# Statistical haemoglobin thresholds to define anaemia across the lifecycle

**DOI:** 10.1101/2023.05.22.23290129

**Authors:** Sabine Braat, Katherine Fielding, Jiru Han, Victoria E. Jackson, Sophie Zaloumis, Jessica Xu Hui Xu, Gemma Moir-Meyer, Sophia M. Blaauwendraad, Vincent W.V. Jaddoe, Romy Gaillard, Patricia C. Parkin, Cornelia M. Borkhoff, Charles D.G. Keown-Stoneman, Catherine S. Birken, Jonathon L. Maguire, Genes & Health Research Team, Melanie Bahlo, Eliza Davidson, Sant-Rayn Pasricha

## Abstract

Detection of anaemia is critical for clinical medicine and public health. Current WHO values that define anaemia are statistical thresholds (5^th^ centile) set over 50 years ago, and are presently <110g/L in children 6-59 months, <115g/L in children 5-11 years, <110g/L in pregnant women, <120g/L in children 12-14 years of age, <120g/L in non-pregnant women, and <130g/L in men. Haemoglobin is sensitive to iron and other nutrient deficiencies, medical illness and inflammation, and is impacted by genetic conditions; thus, careful exclusion of these conditions is crucial to obtain a healthy reference population. We identified data sources from which sufficient clinical and laboratory information was available to determine an apparently healthy reference sample. Individuals were excluded if they had any clinical or biochemical evidence of a condition that may diminish haemoglobin concentration. Discrete 5^th^ centiles were estimated along with two-sided 90% confidence intervals and estimates combined using a fixed-effect approach. Estimates for the 5^th^ centile of the healthy reference population in children were similar between sexes. Thresholds in children 6-23 months were 104.4g/L [90% CI 103.5, 105.3]; in children 24-59 months were 110.2g/L [109.5, 110.9]; and in children 5-11 years were 114.1g/L [113.2, 115.0]. Thresholds diverged by sex in adolescents and adults. In females and males 12-17 years, thresholds were 122.2g/L [121.3, 123.1] and 128.2 [126.4, 130.0], respectively. In adults 18-65 years, thresholds were 119.7g/L [119.1, 120.3] in non-pregnant females and 134.9g/L [134.2, 135.6] in males. Limited analyses indicated 5^th^ centiles in first-trimester pregnancy of 110.3g/L [109.5, 111.0] and 105.9g/L [104.0, 107.7] in the second trimester. All thresholds were robust to variations in definitions and analysis models. Using multiple datasets comprising Asian, African, and European ancestries, we did not identify novel high prevalence genetic variants that influence haemoglobin concentration, other than variants in genes known to cause important clinical disease, suggesting non-clinical genetic factors do not influence the 5^th^ centile between ancestries. Our results directly inform WHO guideline development and provide a platform for global harmonisation of laboratory, clinical and public health haemoglobin thresholds.

## Introduction

Anaemia exists when the red cell mass is insufficient to meet physiologic oxygen-carrying needs, and is usually operationally identified when the haemoglobin concentration falls below a defined threshold for age and sex.^1^ Accurate case definition of anaemia is critical for clinical diagnosis and treatment, and for understanding the magnitude and distribution of this condition as a public health problem. The World Health Organization (WHO) recommends that anaemia be defined when haemoglobin concentration is <110g/L in children 6-59 months, <115g/L in children 5-11 years, <110g/L in pregnant women, <120g/L in children 12-14 years of age, <120g/L in non-pregnant women, and <130g/L in men.^2^ These thresholds reflect the lower 5^th^ centile of the haemoglobin distribution of a reference population of healthy individuals. WHO thresholds were initially proposed in 1958,^3^ updated in 1968,^4^ and have remained essentially unchanged since that time.^5^ WHO thresholds were based on studies with limited measurement of biomarkers of iron and other haematinic deficiency and inflammation. There remains limited consensus on definitions of anaemia, leading to heterogeneous definitions across different sources, expert groups and public health bodies,^6^ translating to inconsistent clinical definitions.^7^

Establishing a valid diagnosis of anaemia is critical for treating patients and detecting the range of diseases that may underlie this condition.^8^ Diagnosis of anaemia lies at the centre of many clinical pathways (for example, investigation of gastrointestinal bleeding,^9^ preoperative optimisation,^10^ antenatal screening for haemoglobinopathy^11^, and eligibility for blood donation^12, 13^). Anaemia is an adverse prognostic factor for multiple conditions, including heart failure,^14^ major surgery,^15^ cancer^16^ and HIV.^17^ Most patients hospitalised with critical illness are anaemic in hospital, and about half remain anaemic six^18^ or even 12 months after hospitalisation.^19^

Valid definitions of anaemia are also critical to guiding population interventions along with tracking progress towards global targets. Based on current definitions, anaemia affects 40% of all preschool children and 30% of women, including 46% of pregnant women globally,^20^ and is a leading cause of Years Lived with Disability.^21^ Reducing anaemia prevalence in women by 50% is a WHO Global Nutrition Target^22^ and a Sustainable Development Goal sub-indicator.^23^ Interventions that directly (for example, iron supplementation^24^ or fortification^25^) or indirectly (for example, malaria control) reduce anaemia are recommended for many low- and middle-income countries, with implementation decisions^26^ and monitoring based on anaemia prevalence.

Haemoglobin thresholds to define anaemia usually vary between men and women, although this distinction has been challenged.^27, 28^ Thresholds may also vary in children and during pregnancy. Haemoglobin levels may be lower in African^29, 30^ or Asian populations,^31^ but it is unclear if this reflects clinically significant genetic conditions affecting the red cell, undefined clinical illnesses, or a different underlying baseline driven by sub-clinical genetic variation, nutritional or environmental factors. It is unknown if individuals with African or Asian heritage still have lower haemoglobin concentrations once carriage of clinically significant genetic polymorphisms that may cause severe genetic syndromes is excluded.

WHO has recognised the need for further evidence for haemoglobin thresholds to define anaemia, including the influence of genetics,^32^ in order to update global guidelines. Here, we present an analysis utilising multiple large-scale datasets in ethnically diverse populations that included sufficiently detailed clinical and laboratory parameters to enable *post-hoc* assembly of reference populations of healthy individuals without discernible risk factors for anaemia, in which 5^th^ centile of haemoglobin distribution can be estimated across the lifecycle. In addition, we estimate ancestry-specific allele frequency and effect on haemoglobin concentrations of genetic polymorphisms and structural variants. We provide a rationale for a single set of thresholds that can be applied globally for clinical and population use.

## Methods

### Estimation of the 5^th^ centile in a healthy population

We searched for datasets comprising appropriate clinical and laboratory information to enable post-hoc derivation of an apparently healthy reference sample (Supplemental Material Section 1). We identified 8 data sources comprising 18 individual datasets that could be included in the analysis. Each included study had received ethics approval and obtained informed consent from participants (Supplemental Material Section 2); use of these datasets for these analyses was deemed by WEHI Governance, Risk and Compliance to meet criteria for exemption from ethics review (Supplemental Material Section 2). Included studies are summarised in Table 1.

**Table 1:**
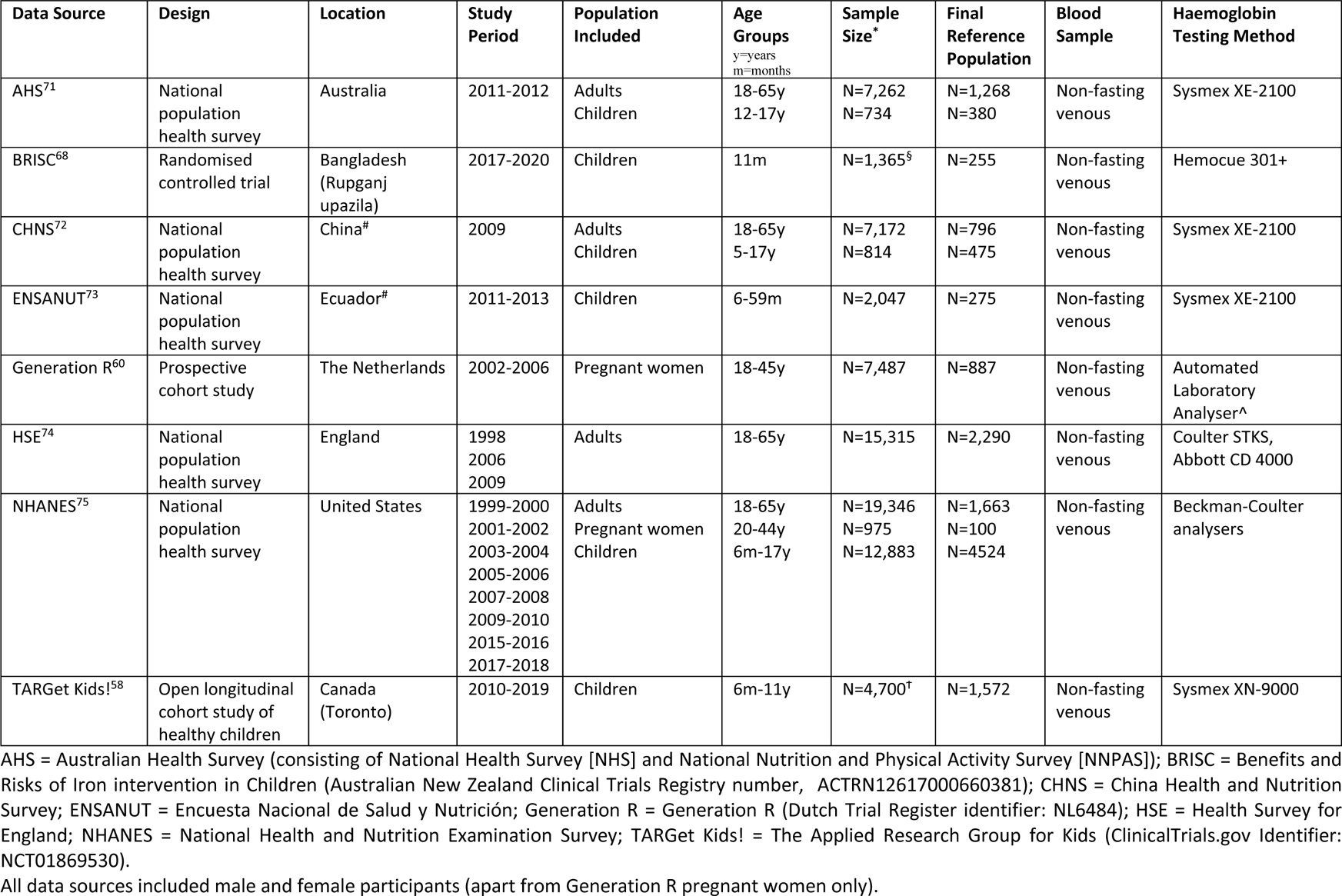
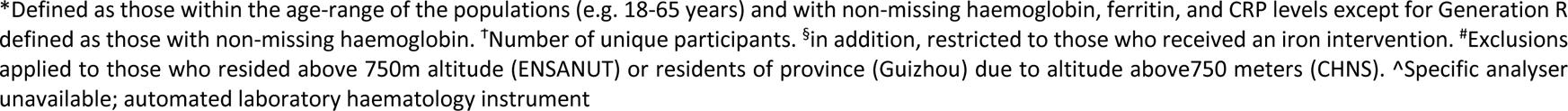
Summary of data used to estimate 5^th^ centiles of haemoglobin concentration

| Data Source | Design | Location | Study Period | Population Included | Age Groups<br>y=years<br>m=months | Sample Size* | Final Reference Population | Blood Sample | Haemoglobin Testing Method |
| --- | --- | --- | --- | --- | --- | --- | --- | --- | --- |
| AHS<sup>71</sup> | National population health survey | Australia | 2011-2012 | Adults<br>Children | 18-65y<br>12-17y | N=7,262<br>N=734 | N=1,268<br>N=380 | Non-fasting venous | Sysmex XE-2100 |
| BRISC<sup>68</sup> | Randomised controlled trial | Bangladesh (Rupganj upazila) | 2017-2020 | Children | 11m | N=1,365<sup>§</sup> | N=255 | Non-fasting venous | Hemocue 301+ |
| CHNS<sup>72</sup> | National population health survey | China<sup>#</sup> | 2009 | Adults<br>Children | 18-65y<br>5-17y | N=7,172<br>N=814 | N=796<br>N=475 | Non-fasting venous | Sysmex XE-2100 |
| ENSANUT<sup>73</sup> | National population health survey | Ecuador<sup>#</sup> | 2011-2013 | Children | 6-59m | N=2,047 | N=275 | Non-fasting venous | Sysmex XE-2100 |
| Generation R<sup>60</sup> | Prospective cohort study | The Netherlands | 2002-2006 | Pregnant women | 18-45y | N=7,487 | N=887 | Non-fasting venous | Automated Laboratory Analyser<sup>^</sup> |
| HSE<sup>74</sup> | National population health survey | England | 1998<br>2006<br>2009 | Adults | 18-65y | N=15,315 | N=2,290 | Non-fasting venous | Coulter STKS, Abbott CD 4000 |
| NHANES<sup>75</sup> | National population health survey | United States | 1999-2000<br>2001-2002<br>2003-2004<br>2005-2006<br>2007-2008<br>2009-2010<br>2015-2016<br>2017-2018 | Adults<br>Pregnant women<br>Children | 18-65y<br>20-44y<br>6m-17y | N=19,346<br>N=975<br>N=12,883 | N=1,663<br>N=100<br>N=4524 | Non-fasting venous | Beckman-Coulter analysers |
| TARGet Kids!<sup>58</sup> | Open longitudinal cohort study of healthy children | Canada (Toronto) | 2010-2019 | Children | 6m-11y | N=4,700<sup>†</sup> | N=1,572 | Non-fasting venous | Sysmex XN-9000 |

AHS = Australian Health Survey (consisting of National Health Survey [NHS] and National Nutrition and Physical Activity Survey [NNPAS]); BRISC = Benefits and Risks of Iron intervention in Children (Australian New Zealand Clinical Trials Registry number, ACTRN12617000660381); CHNS = China Health and Nutrition Survey; ENSANUT = Encuesta Nacional de Salud y Nutrición; Generation R = Generation R (Dutch Trial Register identifier: NL6484); HSE = Health Survey for England; NHANES = National Health and Nutrition Examination Survey; TARGet Kids! = The Applied Research Group for Kids (ClinicalTrials.gov Identifier: NCT01869530).
All data sources included male and female participants (apart from Generation R pregnant women only).
\*Defined as those within the age-range of the populations (e.g. 18-65 years) and with non-missing haemoglobin, ferritin, and CRP levels except for Generation R defined as those with non-missing haemoglobin. <sup>†</sup>Number of unique participants. <sup>§</sup>in addition, restricted to those who received an iron intervention. <sup>#</sup>Exclusions applied to those who resided above 750m altitude (ENSANUT) or residents of province (Guizhou) due to altitude above 750 meters (CHNS). <sup>^</sup>Specific analyser unavailable; automated laboratory haematology instrument

We aimed to define the lower 5^th^ centile of the distribution among healthy individuals (i.e. a healthy reference population). Central to the design of the analysis was the need to establish a reference population from existing datasets (‘*posteriori* approach’) through exclusion of individuals with evidence of conditions or circumstances that may influence haemoglobin concentration.^33^ Haemoglobin levels are reduced by acute, recurrent, chronic medical or surgical illness. Inflammation (for example due to autoimmune disease, infection, solid or haematologic cancer, heart failure and even obesity) suppresses erythropoiesis due to hepcidin-mediated functional iron deficiency and may also reduce red cell survival.^8^ Haemoglobin concentrations are reduced in renal impairment due to reduced erythropoietin production and functional iron deficiency. Many medications may reduce haemoglobin idiosyncratically or in a dose-dependent manner via reduced erythropoiesis, reduced red cell survival or blood loss. Anaemia can persist weeks or months beyond an acute illness^19^ and even beyond normalisation of acute inflammatory markers.^34^ Conversely, hypoxia (for example, smoking, respiratory or cardiac disease, obesity or sleep apnoea, or elevated altitude of residence)^35^ may increase haemoglobin concentrations.

Criteria for exclusion included report of chronic systemic medical illness; any recent illness; recent hospitalisation; use of medications; current smoking or excess alcohol consumption; obesity or low weight. Biomarker evidence of iron status and inflammation were required for all participants; biochemistry for other haematinic deficiencies and renal or hepatic impairment were exclusion criteria where possible. Where data were available, we excluded individuals living at elevations above 750m. Although exclusion criteria were standardised, available data and its coding varied between studies. We sought to ensure ethnic diversity but recognised that population field surveys in impoverished settings may contain a high burden of unreported, undetected or recently resolved inflammation even if acute biomarkers of inflammation had normalised, preventing exclusion of individuals with recent illness that may have lowered haemoglobin concentration.^36^ Haemoglobin measurements were on venous blood using an automated analyser or high-quality point-of-care device (see Table 1). Detailed inclusion and exclusion of individuals from each dataset to obtain the reference sample are shown in Supplemental Material Sections 2.1-2.8.

### Statistical methods

Lower 2.5^th^ and 5^th^ centiles were estimated using the methodology described below and detailed in Supplemental Material Section 2.9. The Clinical & Laboratory Standards Institute recommends two-sided 90% confidence intervals be calculated for each reference limit.^37^ Outliers were identified using Tukey’s method.^38^ All analyses were performed with and without outliers, and presented without outliers. We estimated both discrete thresholds (based on conventionally applied age categories) and continuous thresholds (based on the participant’s age [and sex if applicable] within age categories).

### Discrete thresholds

The parametric theoretical centile of a Gaussian distribution, with corresponding 90% confidence interval, was used to estimate the centile.^39, 40^ For the national population health surveys National Health and Nutrition Examination Survey (NHANES), Health Survey for England (HSE), China Health and Nutrition Survey (CHNS), and Encuesta Nacional de Salud y Nutrición (ENSANUT) both survey-weighted and unweighted (sensitivity) centile estimates were calculated. Survey-weighted centile estimates were calculated using survey-weighted quantile regression. The implementation of survey-weighted quantile regression proceeded as described in “Continuous thresholds” (below), except the survey-weighted quantile regression model only included a single intercept term. Standard errors for the intercept term were derived from samples generated using Canty and Davidson’s bootstrap^41, 42^ and two-sided 90% confidence intervals derived assuming a normal approximation.^43^ Unweighted centile estimates were calculated using the parametric theoretical centile described earlier. Centile estimates were pooled across all data sources using fixed effect and random effects (sensitivity) meta-analyses and presented in forest plots.

### Continuous Thresholds

We used the Hoq^44^ method to estimate age-specific (and by sex, if applicable) continuous haemoglobin thresholds within each age-category by combining all relevant data sets, except for National Health Survey (NHS) and National Nutrition and Physical Activity Survey (NNPAS) due to Australian Bureau of Statistics requirements. This involved identifying the best fitting multivariable fractional polynomial (MFP) model for the mean values of haemoglobin using Royston’s method^45^ followed by a likelihood ratio test for an interaction between sex and the fractional polynomial representation of age. An age-by-sex interaction term was included in the model if they were statistically significant at the nominal significance level of p<0.05. Unweighted quantile regression was then used. Age-dependent haemoglobin predictions were obtained from the quantile regression model and two-sided 90% bootstrap percentile confidence intervals generated, based on 1000 bootstrap samples. Results were presented in plots to indicate how predicted percentiles change with age (and by sex, if applicable), superimposed onto a scatter plot of haemoglobin versus age within each age-category. No continuous thresholds were obtained for pregnant women.

### Genetic analyses

We accessed three large-scale, multi-ancestry GWAS summary statistics associated with haemoglobin concentration trait (Chen 2020^46^; Wheeler 2022^47^; Genes & Health (G&H) Study^48^). Chen and G&H analysed single nucleotide polymorphisms (SNPs), whilst Wheeler considered structural variations (SVs). In each study, individuals were classified as being genetically similar to one of the five “super-populations” defined as part of the 1000 Genomes study: European (EUR), East Asian (EAS), African (AFR), Hispanic/Latino, and South Asian (SAS), allowing for assessment of the effects of haemoglobin-associated variants on haemoglobin concentrations across ancestry groups. Details are given in Supplemental Table 6.1.

Population-specific effect size and allele frequencies were extracted for all variants significantly associated with haemoglobin, in one or more ancestral population. To account for linkage disequilibrium (LD), we sought to select one representative SNP per genetic region for our summaries.

Chen reported independent variants associated with haemoglobin concentrations defined using iterative conditional analyses approach, so we extracted their predefined lists of independent associated SNPs. For the Genes & Health study results, we performed LD clumping with an r^2^<0.1 and a clumping window size of 500 kb to identify independent signals. We examined the overlap of independent signals from the G&H study and Chen analyses, with signals defined as overlapping where the identified representative SNPs from each study were in linkage disequilibrium, with r^2^>0.1. Where signals were overlapping, we selected the top SNP defined in Chen as the representative SNP for that signal.

Both utilised GWAS applied rank-based inverse rank normal transformation (IRNT) to the haemoglobin concentration measurements, prior to association with genetic variants. Effect sizes stated in the text are as reported in terms of the IRNT trait. To provide a better clinical interpretation of the reported effect size estimates, on the plots we also include a scale giving approximate effects in terms of units of haemoglobin measurement (in g/L). We used the UK Biobank (UKBB) population-based cohort (Application Number: 36610; Data-Field 30020) for these approximations by multiplying the IRNT effect size by the standard deviation of haemoglobin concentration in the UKBB cohort.

We also considered structural variants identified in Wheeler.^47^ We extracted ancestry specific frequencies for the identified SVs, but effect estimates were only available from a combined ancestry analysis. This study performed LD and conditional analyses for trait-associated SVs to previous reported GWAS variants to determine whether these SVs were being tagged by the GWAS SNPs.

We accessed gene-based summary statistics from the AstraZeneca PheWAS Portal.^49^ Gene-phenotype associations were tested with multiple collapsing models.^49^ We extracted gene associations of haemoglobin concentrations using the collapsing model Ptv5pcnt (defined as protein-truncating variants; PTVs, with MAF ≤ 5% both within the UKBB cohort and gnomAD). A gene with a p-value less than a Bonferroni corrected threshold of 0.05 divided by the 18,762 genes^49^ tested (P < 2.665 × 10^−6^) was considered significant. For significant genes, we further restricted to those identified as clinically relevant causes of rare anaemia from the Genomics England PanelApp ‘Green’ gene list (Version 3.1)^50^ and investigated the frequency distributions of predicted loss-of-function (pLoF) variants in different populations of gnomAD.^51, 52^ For each gene, we also estimated a cumulative frequency of pLoF variants, within each population, as the sum of alternate allele count for each variant, divided by the mean number of genotypes available across all variants in the gene.

### Software

Reference samples were derived using Stata version 16.1^53^, R version 4.1.1.^54^ (Generation R), or Stata version 17.1.^55^ (NHS and NNPAS). ^54^ Estimation of haemoglobin thresholds was performed using R version 4.2.3,^54^ R version 4.1.1.^54^ (Generation R), or Stata version 17.1.^55^ (NHS and NNPAS) with R packages detailed in Supplemental Material Section 2.9.6. Genetic analyses was performed in R version 4.1.3.^54^ using R packages tidyverse (version 1.3.2), data.table (version 1.14.2), ieugwasr (version 0.1.5), ggplot2 (version 3.4.1), ggrepel (version 0.9.1), UpSetR (version 1.4.0),^56^ and ComplexHeatmap (version 2.13.1).^57^

## Results

We separately analysed thresholds for adult men, adult women, children aged 6 months to 59 months (further sub-categorised as 6-23 months and 24-59 months), 5-11 years, adolescent males and females (12-17 years), and pregnant women (by trimester). For each survey demographic characteristics (including age, self-reported ancestry, and haemoglobin, iron status, inflammation) for the overall (non-missing haemoglobin, ferritin, and C-Reactive Protein [CRP] value) and reference (healthy) sample are shown in Supplemental Tables 3.1.1-3.8.3. Reasons for exclusion from the reference sample are provided in each of Figures 2-5, and details are provided in Supplemental Tables 4.1.1-4.6.3. Pooled analyses for the 5^th^ centile for haemoglobin concentration in healthy individuals is presented in Figures 1-4, while results for the 2.5^th^ centile are available in Supplemental Tables 5.1-5.3.

### Continuous thresholds across the life course

Continuous thresholds in males and females based on all available datasets are summarised in Extended Figure 1, indicating periods of change and stability in haemoglobin across the life course.

**Figure 1:**
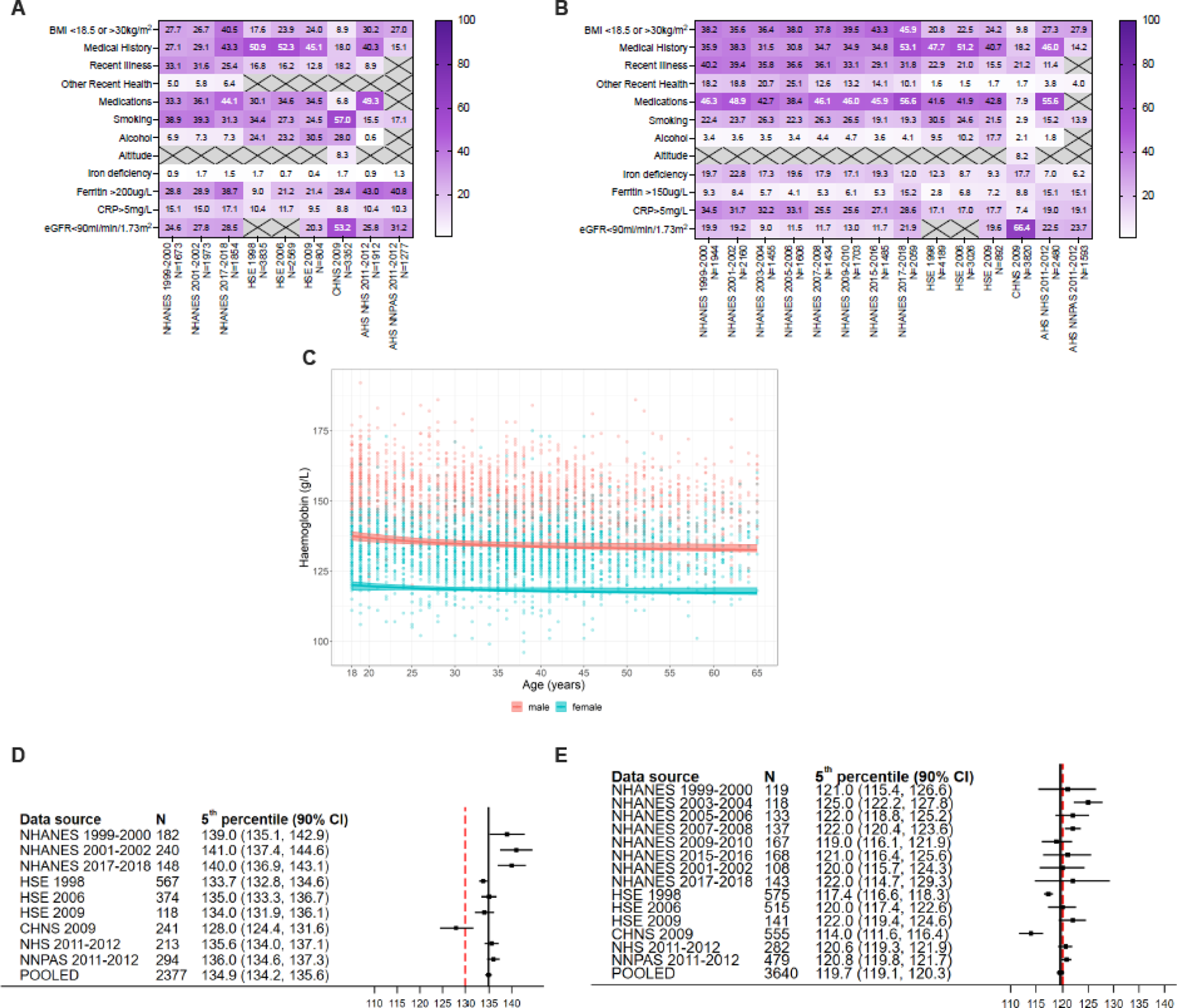
Derivation of haemoglobin thresholds in 18–65-year-old males and females. A) Exclusions from overall dataset in males. B) Exclusions from overall dataset in non-pregnant females. C) Continuous thresholds in 18–65-year-old adult males (red) and non-pregnant females (blue). D) Individual study and pooled (fixed effect) estimates of the 5^th^ centile of the haemoglobin distribution in the healthy reference population in males. E) Individual study and pooled (fixed effect) estimates of the 5^th^ centile of the haemoglobin distribution in the healthy reference population in non-pregnant females.

### Adult women and men

Figure 1 presents estimates of haemoglobin threshold to define anaemia in adult men and women. The data was largely derived from multi-ethnic populations (comprising individuals who self-identified as White, Black, and Asian) across the United States (US), England, and Australia and from China (Supplemental Tables 3.2.1-3.2.14). Reasons for exclusion of individuals from each dataset are summarised in Figures 2A and 2B and detailed in Supplemental Tables 4.1.1-4.1.14. Continuous analyses of the data indicates that in adults between 18 and 65 years of age, the 5^th^ centile haemoglobin concentration of the reference population is higher in males than non-pregnant females (Figure 2C). Pooled analyses indicates that the 5^th^ centile for haemoglobin concentrations in healthy adult men is 134.9g/L [90% Confidence Interval 134.2, 135.6] (Figure 2D). In adult non-pregnant women, the 5^th^ centile is 119.7g/L [119.1, 120.3] (Figure 2E). Sensitivity analyses were performed where the ferritin threshold to indicate iron deficiency is raised from 15ug/L to 45ug/L and 100ug/L; this indicates the haemoglobin 5^th^ centile of ∼120g/L in women is robust and remains lower than in men (Supplementary Figures 5.4-5.5).

**Figure 2:**
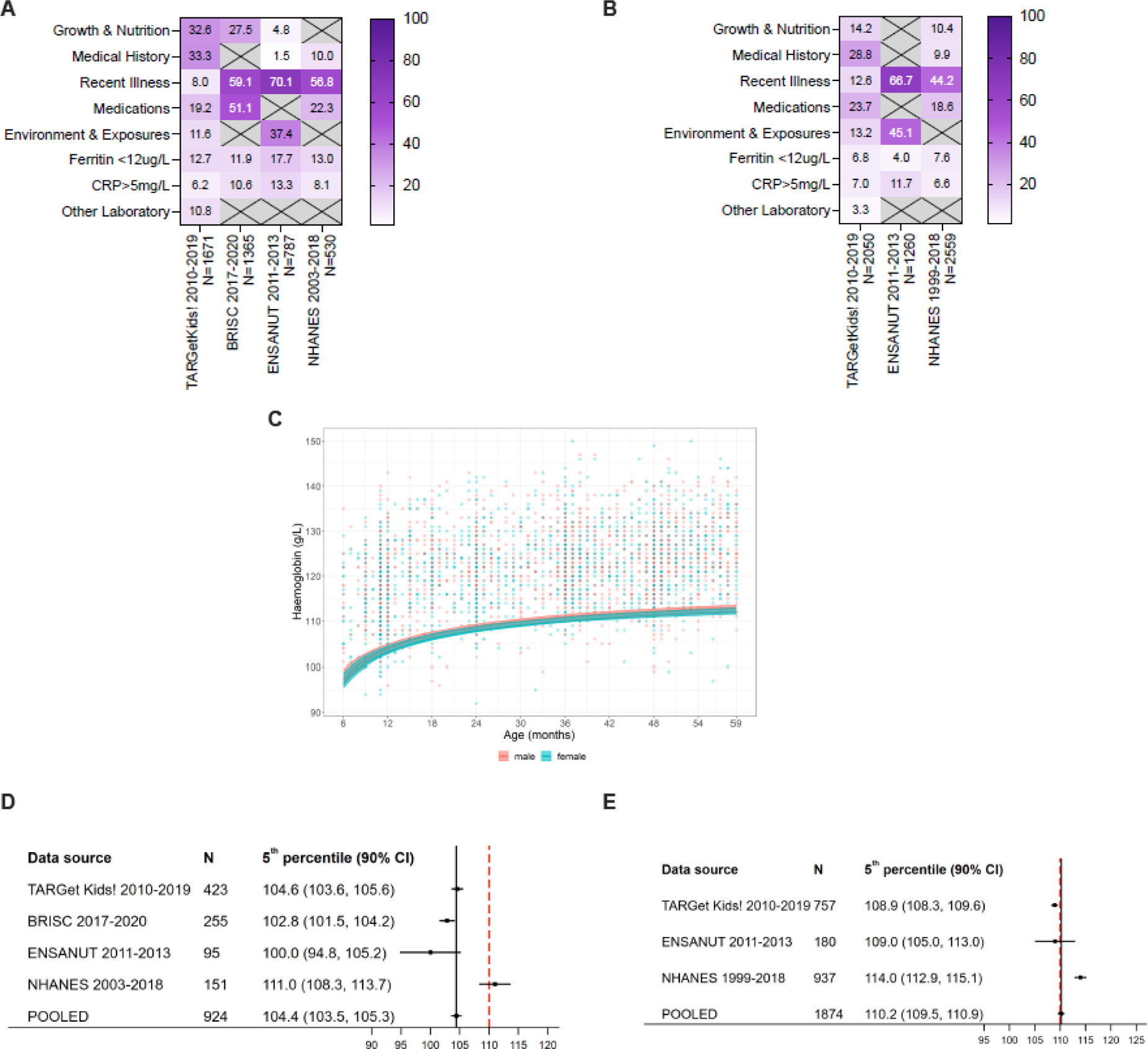
Derivation of haemoglobin thresholds in 6–59-month-old children. Exclusions from overall dataset in children 6-59 months, sub-categorised into children A) 6-23 months and children B) 24-59 months. C) Continuous thresholds in 6–59-month-old male (red) and female (blue) children. D) Individual study and pooled (fixed effect) estimates of the 5^th^ centile of the haemoglobin distribution in the healthy reference population in children 6-23 months. E) Individual study and pooled (fixed effect) estimates of the 5^th^ centile of the haemoglobin distribution in the healthy reference population in children 24-59 months.

### Children

Figure 2 summarises estimates of haemoglobin thresholds in children 6 to 59 months of age. Data were derived from multi-ethnic populations across Canada,^58^ the US, Ecuador^59^, and Bangladesh (Supplemental Tables 3.3.1-3.4.3). Reasons for exclusion from each dataset are summarised in Figure 2A and detailed in Supplemental Tables 4.2.1-4.3.3. Continuous analyses of the data indicates that thresholds are similar in males and females and hence that sex-specific thresholds are not necessary in this age group. Continuous analyses also indicate an increase in thresholds over the first 5 years of life but especially over the first two years (Figure 2B). Discrete thresholds were therefore determined in children 6-23 months of age and 24-59 months of age. Pooled analysis indicates the 5^th^ centile for haemoglobin concentrations in children 6-23 months is 104.4g/L [103.5, 105.3] (Figure 2C), and in children 24-59 months is 110.2g/L [109.5, 110.9] (Figure 2D). Sensitivity analyses were performed where the ferritin threshold to indicate iron deficiency was raised, the CRP threshold to indicate inflammation was lowered, and where only children with a mean cell volume (MCV) above 73 fL (6-23 months) or above 75 fL (24-59 months) were included. These sensitivity analyses indicate the 5^th^ centile estimate is robust to these parameters, as well as to changes in the analytic model (Supplementary Figures 5.6-5.11).

Figure 3 summarises estimates of haemoglobin thresholds in children 5-11 years of age. Data were derived from multi-ethnic populations across Canada, the US, and China (supplementary Table 3.5.1-3.5.3). Reasons for exclusion of individuals from each dataset are summarised in Figure 3A and detailed in Supplemental Table 4.4.1-4.4.3. Continuous analyses of the data indicates that thresholds are similar in males and females and hence that sex-specific thresholds are not necessary in this age group (Figure 3B). Pooled analyses indicated the 5^th^ centile for haemoglobin concentrations in this group is about 114.1g/L [113.2, 115.0] (Figure 3C). Sensitivity analyses indicated these thresholds were robust to changes in assumptions and the analytic model (Supplementary Figures 5.12-5.14).

**Figure 3:**
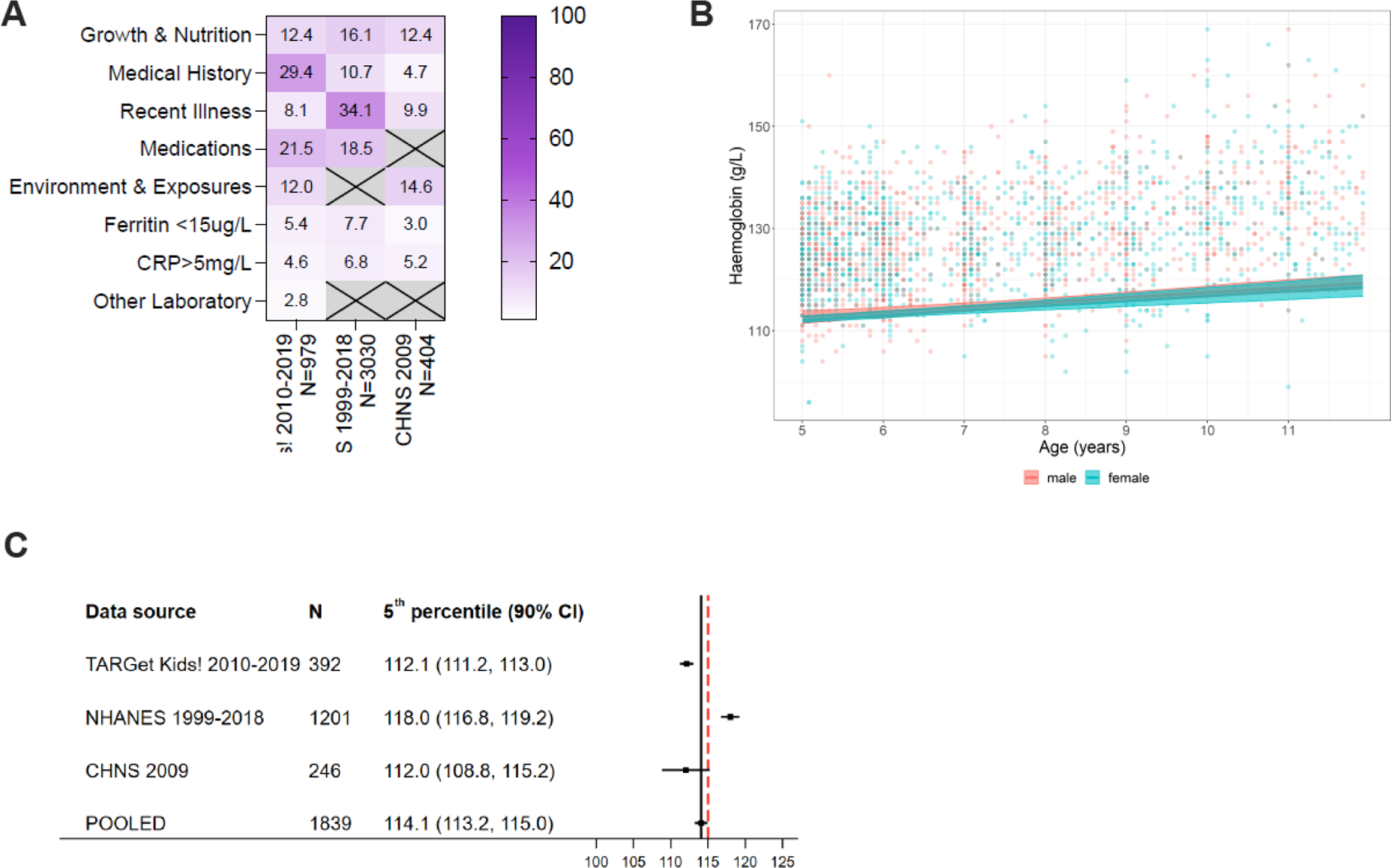
Derivation of haemoglobin thresholds in 5–11-year-old children. A) Exclusions from overall dataset in children 5-11 years. B) Continuous thresholds in 5–11-year-old male (red) and female (blue) children. C) Individual study and pooled (fixed effect) estimates of the 5^th^ centile of the haemoglobin distribution in the healthy reference population in children 5-11 years of age.

Figure 4 summarises estimates of haemoglobin thresholds in children 12-17 years of age. Data were derived from multi-ethnic populations across Canada, the US, Australia, and China (Supplemental Tables 3.7.1-3.7.9). Reasons for exclusion of individuals from each dataset are summarised in Figure 4A and detailed in Supplemental Table 4.5.1-4.5.11. Continuous analyses show that thresholds diverge between males and females over this period (Figure 4B), indicating that sex-specific thresholds are necessary in this age group. Pooled analyses indicated the 5^th^ centile for haemoglobin concentrations in this group is 128.2 g/L [126.4, 130.0] in adolescent males (Figure 4C) and 122.2g/L [121.3, 123.1] in adolescent females (Figure 4D). Sensitivity analyses indicated these thresholds were robust to changes in assumptions and the analytic model (Supplementary Figure 5.15-5.20).

**Figure 4:**
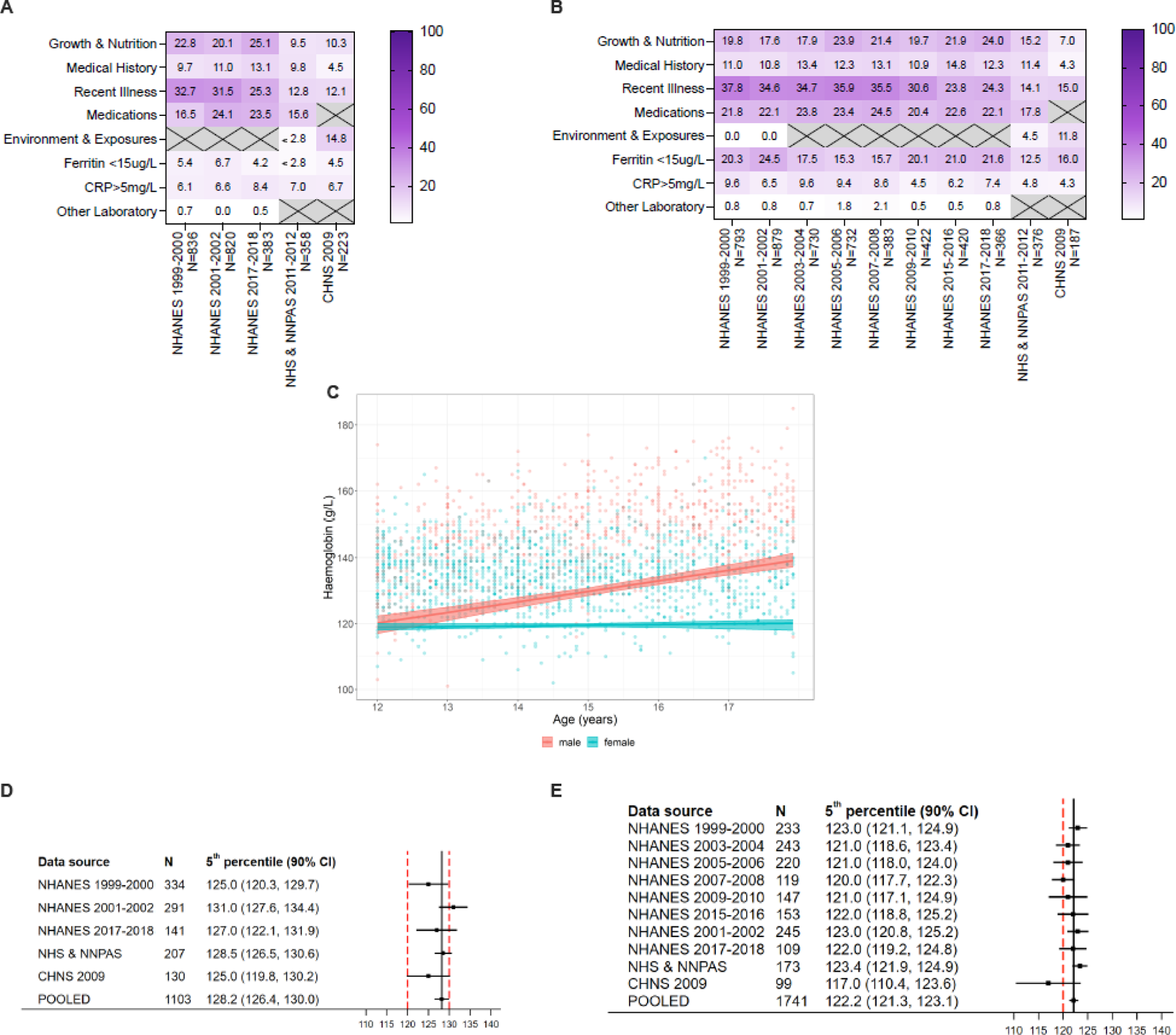
Derivation of haemoglobin thresholds in 12–17-year-old males and females. Exclusions from overall dataset in children 12-17 years, sub-categorised into A) females and B) males. C) Continuous thresholds in 12–17-year-old male (red) and female (blue) children. D) Individual study and pooled (fixed effect) estimates of the 5^th^ centile of the haemoglobin distribution in the healthy reference population in 12–17-year-old males. E) Individual study and pooled (fixed effect) estimates of the 5^th^ centile of the haemoglobin distribution in the healthy reference population in 12–17-year-old females.

### Pregnancy

We were able to access data from only two studies where sufficient clinical information, iron biochemistry and inflammatory biomarkers and haemoglobin concentrations had been measured during pregnancy (NHANES, Generation R^60^). Iron deficiency, inflammation and medical complications were common in these cohorts. Insufficient data was available from NHANES datasets to proceed with analyses. Limited data from Generation R indicated that the 5^th^ centile for haemoglobin thresholds in healthy women was 110.3g/L [109.5, 111.0] in the first trimester, and 105.9g/L [104.0, 107.7] in the second trimester, with insufficient data available for analysis in the third trimester. Participant characteristics are summarised in (Supplementary Table 3.8.1-3.8.3). Exclusions from this dataset are shown in Supplementary Table 4.6.1-4.6.3. The thresholds were robust to changes in assumptions around definition of iron deficiency and inflammation (Supplementary Figure 5.21).

### Ancestry-specific genetic associations with variation in haemoglobin concentration

We investigated whether common genetic variation may explain different haemoglobin concentrations among different ancestry groups using summary statistics from three large-scale, multi-ancestry GWAS of haemoglobin concentrations. We summarised the genetic variants (SNPs and SVs) affecting haemoglobin concentrations and the distribution of effect sizes across different ancestries. We sought to identify whether there are any genetic variants (SNPs and SVs) with large effects that contribute substantially to overall variance in haemoglobin concentration in particular ancestral populations.

We collated the SNP association results from Chen and Genes & Health. We found 402, 21, 2, 2, 3 independent genome-wide significant (P<5×10^−9^) SNPs in the EUR, EAS, AFR, SAS (Chen et al), and SAS Genes & Health groups, respectively (Supplemental Figure 6.1). The larger sample size of EUR and multi-ancestry analyses leads to a higher proportion of loci significant only in EUR and the multi-ancestry analyses. Investigation of the distribution of minor allele frequency (MAF, frequency of the less common allele in the given population) and effect sizes demonstrates an inverse relationship between MAF and effect size (Figure 5), as is commonly seen in complex genetic traits. No ancestries appear to have any common SNPs (MAF > 0.05) with large effect size, except for rs13331259 (a *FAM234A* intronic variant) which shows a larger effect size (P=3.32 × 10^−4^^1^, IRNT effect size = −0.2495, approximately −3.10 g/L per allele) and has a MAF of 0.11 in the AFR population. This variant is in LD (r^2^=0.48 in AFR) with rs76462751 (a *HBA1* downstream variant). This variant was very rare in the EUR and multi-ancestry analyses, with MAFs of 0.0001 and 0.0038, respectively.

**Figure 5:**
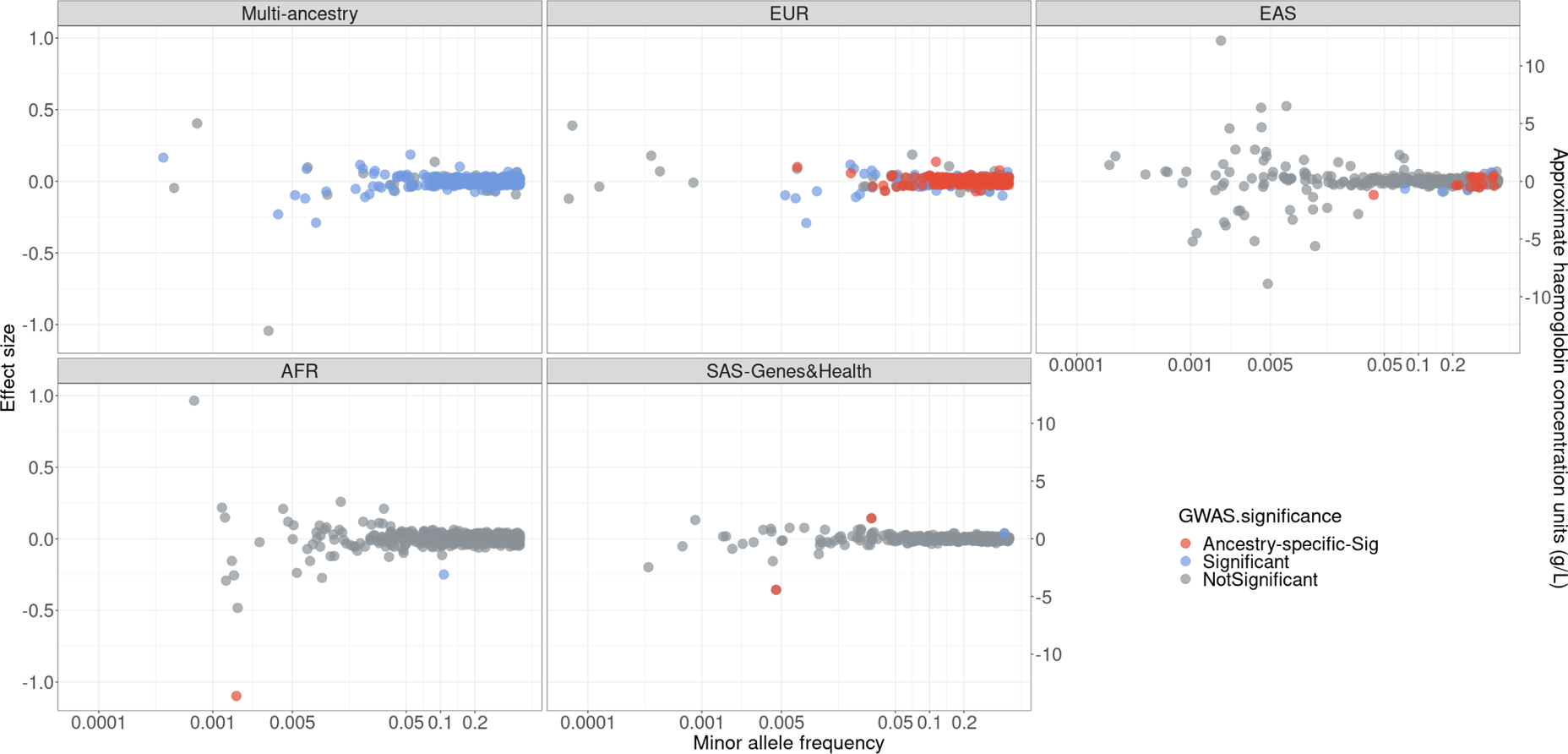
Scatter plot of minor allele frequency (MAF) and effect size. MAF (x-axis on a log10 scale) and standardised effect size (y-axis) of the minor alleles of the 607 variants (both ancestry-specific and cross-ancestry loci) in the GWAS summary statistics. Included in this figure are the 607 haemoglobin variants with a P < 5 × 10^−9^ from a single or multi-ancestry analysis. Genome-wide significant variants for the stated ancestry are indicated in red. Variants that were genome-wide significant in the stated ancestry, and in at least one other ancestry are indicated in blue. Variants that did not reach genome-wide significance in the stated ancestry are indicated in grey. To convert the standardised effect size to an approximate haemoglobin unit (g/L), we regarded standard deviation per minor allele as 12.41 units (g/L), which was estimated from the UKBB population-based cohort.

Across the two analyses of SAS individuals, three SNPs representing independent signals were identified; of these, two were specific to that ancestry and notably did not reach genome-wide significance in the non-SAS GWAS, despite larger sample sizes. These were rs529302116 (an *OR52E3P* and *OR52J1P* intergenic variant on Chr11) and rs563555492 (a *PIEZO1* missense variant) (Supplemental Figure 6.2). rs529302116 is in strong LD (r^2^=0.83) with rs33915217 (a *HBB* intronic variant) in the SAS population. This variant occurs more commonly in populations of South Asian ancestry and is one of the more prominent variants observed in patients with β-thalassaemia^61^.

In addition to the association results of SNPs, we also summarised the SVs association results of Wheeler et al.^62^ (Supplemental Figure 6.3). There were two SVs chr16:172001-177200_DEL (*HBA1, HBA2*) and chr22:37067818-37067888_DUP (*TMPRSS6*) at genome-wide significance (P< 5×10^−8^). Of these, chr22:37067818-37067888_DUP (*TMPRSS6*) is in LD (r^2^=0.77) with rs5756504 (a *TMPRSS6* intronic variant), that has previously been identified through GWAS. The second SV, chr16:172001-177200_DEL (*HBA1, HBA2*) occurred at a high frequency in the AFR ancestry cohort (MAF=0.176) and is located ∼80kb away from the SNP GWAS signal rs13331259 that also showed higher MAF in the AFR ancestry. Conditional analyses found the SNP and SV signals to be independent however, suggesting there may be multiple variants in this region influencing haemoglobin concentrations that occur at higher frequencies in African populations, potentially due to positive selection.

Single-variant-based GWAS is typically focused on the identification of common variants (MAF>0.05) but is underpowered for low-frequency and rare variants due to small sample sizes and low minor allele frequencies. However, these less-common variants could explain additional disease risk or trait variability. To address these limitations, gene-based collapsing analysis of multiple variants in a defined genetic region can identify genes in which low-frequency and rare variants are, in aggregate, associated with a phenotype. To evaluate this, we accessed the AstraZeneca PheWAS Portal^49^ which contains gene–phenotype associations calculated from exome-sequenced UK Biobank participants of European ancestry. From the gene-based results, we identified 13 genes associated with haemoglobin concentrations, with P<2.665×10^−6^, of which five (*HBB, KLF1, PIEZO1, SLC4A1, TMPRSS6*) were considered clinically relevant causes of rare anaemia via the Genomics England PanelApp ‘Green’ gene list^50^ (Supplemental Figure 6.4). Most significant genes show relatively small effect sizes, except for *HBB* which was strongly associated with lower haemoglobin concentrations (P = 2.31 × 10^−9^^2^, IRNT effect size=-1.8941, approximately −23.51 g/L per allele). Across these five clinically relevant rare anaemia genes, we extracted allele frequencies for all pLoF variants from gnomAD and estimated cumulative frequencies of pLoF variants for each gene. The pLoF variants in these genes occurred at varying frequencies in different populations (Supplemental Figure 6.5). For example, *HBB* has a higher prevalence in individuals of EAS, SAS, and Middle Eastern ancestries; however, the cumulative pLoF variants frequency is lower than 0.006, and therefore these variants, even collectively, may be considered rare and unlikely to influence the 5^th^ centile of haemoglobin concentration. Rare pLoF variants in *TMPRSS6* have the highest prevalence in African/African American populations, with a cumulative pLoF variant frequency of around 0.03. The gene-based association with *TMPRSS6* (in Europeans), showed a more modest effect (P=2.41 × 10^-7^, IRNT effect size=-0.1419, approximately −1.76 g/L per allele).

## Discussion

We have estimated the 5^th^ centiles of haemoglobin concentration in healthy young and older children, adolescents, and adults, which can be used to define anaemia across the lifecycle. These analyses largely support continued use of existing WHO haemoglobin thresholds to define anaemia with the exception of a reduction in threshold from 110g/L to 105g/L in children 6-23 months of age, reduction in the second trimester of pregnancy to 105g/L, and perhaps an increase in men from 130g/L to 135g/L. Analyses of genetic associations with haemoglobin concentration across different ancestries shows no evidence of the presence of any non-clinically significant high-frequency variants, indicating that single global definitions for haemoglobin thresholds are appropriate.

Our analyses indicate that statistical anaemia definitions differ between adult women and men, with the lower threshold in women robust to sensitivity analyses utilising higher ferritin thresholds (e.g. <45ug/L^9^) to indicate iron deficiency. Thresholds between sexes diverge in adolescence, when higher thresholds in males may relate to testosterone-induced erythropoiesis.^63^ Our data do not support proposals that statistical thresholds to define anaemia should be similar between adult men and women,^27^ although it is important to recognise that in some studies, similarly reduced haemoglobin concentrations in both sexes have been associated with adverse clinical outcomes.^28^

Our analyses highlight the increase in haemoglobin concentration and hence threshold for anaemia that occurs across childhood, and especially children 6-23 months old. Thresholds based on arbitrary age bands are possibly crude and necessitate jarring changes in the definition of anaemia as children grow and develop. Our data indicate a single 6–59-month categorisation is too broad, and we propose at least dividing this population into two smaller subcategories (i.e. 6-23 months and 24-59 months). Thresholds in male and female children can be combined until adolescence.

Analyses of population surveys in India^64^ and field surveys of biochemically iron-replete, biochemically non-inflamed individuals across several low and middle-income countries have reported substantial heterogeneity of the 5^th^ centile of haemoglobin thresholds between countries in women and preschool children.^36^ Explanations for these heterogenous thresholds are uncertain, but may relate to undetected current or recent inflammatory disease. For example, asymptomatic submicroscopic *Plasmodium* parasitaemia is highly prevalent in sub-Saharan African children,^65, 66^ while in India the incidence of acute respiratory infections in children under 3 years is 2766 per 1000 child years,^67^ and about 22% of Bangladeshi infants receive some form of health care due to illness (usually fever, vomiting or diarrhoea) over a three month period.^68^

We found evidence for heterogeneity of both common and rare genetic effects on haemoglobin across different ancestries, including two variants proximal to the alpha-thalassaemia genes (*HBA1/ HBA2*) which show larger effect sizes than might be expected given their frequency within individuals of African ancestry. These variants are of clinical importance and should be detectable by a diagnosis of anaemia. We also examined rare variants in genes that are recognised to be clinically relevant causes of inherited anaemia and found these to vary in frequency across populations. Although there was some evidence there may be a higher burden of rare variants with large effects in some ancestral populations, clinically relevant rare variants cumulatively occur at low frequencies, and thus are not likely to result in a significantly different distribution of haemoglobin in the overall population. Thus our study does not find it would be clinically safe or appropriate to adopt lower haemoglobin thresholds in populations or individuals of different ancestries. A further limitation of ancestry-specific thresholds is that clinical classification of individuals into ancestral groups is imperfect, as there is substantial genetic diversity within the “super populations”, and further, such approaches could not account for admixed individuals.

Our findings were robust to variations in criteria for iron deficiency and inflammation, to analyses models, and to secular health trends. Our study has several limitations. We estimated statistical thresholds based on the 5^th^ centile of the distribution of haemoglobin values for a ‘healthy’ reference population, rather than ‘functional’ thresholds that indicate risk of symptoms or underlying disease. Ideally, a prospective multicentre study could clinically exclude individuals at risk of reduced haemoglobin concentration to define the reference group.^69, 70^ We sought to overcome this limitation by only utilising datasets where clinical data were available and applying conservative exclusion criteria. Unfortunately, this approach precluded use of cross-sectional datasets from low-income settings where clinical information was limited and undetected subclinical or recurrent clinical illnesses causing inflammation may have suppressed haemoglobin levels, which could lead to under-estimation of thresholds. We were able to utilise datasets from North and South America, Europe, Australia, South and East Asia, but not sub-Saharan Africa. However, the reference samples generally comprised multi-ethnic populations (including African ancestries). Likewise, as is a common constraint with genetic analyses, the largest datasets were representative of European ancestry populations, although we specifically included studies recruiting substantial numbers from South and East Asian and African ancestries. A further limitation of the genetic summaries we have used is that they do not capture the full extent of genetic variation, for example, variants on non-autosomal chromosomes (X, Y, and mitochondria). Analysis of thresholds in those aged less than 6 months, above 65 years and pregnancy was limited, due to insufficient sample size or datasets which simultaneously recorded haemoglobin, iron biochemistry and clinical parameters.

This set of updated haemoglobin thresholds to define anaemia that can be applied by WHO, clinicians, diagnostic laboratories, and public health practitioners.

## Data Availability

Study data used for this analysis are either publicly available or were accessed with agreement from primary data collection sources, and will not be available from the study authors.

https://beta.ukdataservice.ac.uk/datacatalogue/series/series?id=2000021

https://www.cdc.gov/nchs/nhanes/

https://www.abs.gov.au/ausstats/abs@.nsf/mf/4363.0.55.001

https://www.cpc.unc.edu/projects/china

https://www.salud.gob.ec/encuesta-nacional-de-salud-y-nutricion-ensanut/

https://www.targetkids.ca/

https://generationr.nl/

https://www.wehi.edu.au/people/sant-rayn-pasricha/trials/brisc

## Acknowledgements

We thank the many people who have participated in or worked on the studies contributing data to this analysis. We thank Ms Vanessa Pac Soo and Ms Joanna Ling, Methods and Implementation Support for Clinical and Health Research Hub, Faculty of Medicine, Dentistry and Health Sciences, The University of Melbourne, Melbourne, Australia for their assistance with data management. We thank Ms Naomi Von Dinklage, Walter and Eliza Hall Institute of Medical Research, Melbourne, Australia for administrative support.

We thank all members from the Genes & Health Research Team (full list in the Supplementary Information). We thank Social Action for Health, Centre of The Cell, members of our Community Advisory Group, and staff who have recruited and collected data from volunteers. We thank the NIHR National Biosample Centre (UK Biocentre), the Social Genetic & Developmental Psychiatry Centre (King’s College London), Wellcome Sanger Institute, and Broad Institute for sample processing, genotyping, sequencing and variant annotation. We thank: Barts Health NHS Trust, NHS Clinical Commissioning Groups (City and Hackney, Waltham Forest, Tower Hamlets, Newham, Redbridge, Havering, Barking and Dagenham), East London NHS Foundation Trust, Bradford Teaching Hospitals NHS Foundation Trust, Public Health England (especially David Wyllie), Discovery Data Service/Endeavour Health Charitable Trust (especially David Stables), Voror Health Technologies Ltd (especially Sophie Don), NHS England (for what was NHS Digital) - for GDPR-compliant data sharing backed by individual written informed consent. We thank all of the volunteers participating in Genes & Health.

Genes & Health Research Team (in alphabetical order by surname): Shaheen Akhtar, Mohammad Anwar, Elena Arciero, Omar Asgar, Samina Ashraf, Saeed Bidi, Gerome Breen, Raymond Chung, David Collier, Charles J Curtis, Shabana Chaudhary, Megan Clinch, Grainne Colligan, Panos Deloukas, Ceri Durham, Faiza Durrani, Fabiola Eto, Sarah Finer, Joseph Gafton, Ana Angel Garcia, Chris Griffiths, Joanne Harvey, Teng Heng, Sam Hodgson, Qin Qin Huang, Matt Hurles, Karen A Hunt, Shapna Hussain, Kamrul Islam, Vivek Iyer, Ben Jacobs, Ahsan Khan, Cath Lavery, Sang Hyuck Lee, Robin Lerner, Daniel MacArthur, Daniel Malawsky, Hilary Martin, Dan Mason, Rohini Mathur, Mohammed Bodrul Mazid, John McDermott, Caroline Morton, Bill Newman, Elizabeth Owor, Asma Qureshi, Samiha Rahman, Shwetha Ramachandrappa, Mehru Reza, Jessry Russell, Nishat Safa, Miriam Samuel, Michael Simpson, John Solly, Marie Spreckley. Daniel Stow, Michael Taylor, Richard C Trembath, Karen Tricker, Nasir Uddin, David A van Heel, Klaudia Walter, Caroline Winckley, Suzanne Wood, John Wright, Julia Zollner.

We thank the National Institute for Nutrition and Health, China Center for Disease Control and Prevention, Beijing Municipal Center for Disease Control and Prevention, and the Chinese National Human Genome Center at Shanghai. We thank all participating children and families for their time and involvement in TARGet Kids!. We thank the TARGet Kids! Collaboration for supporting this study (details may be found on the website www.targetkids.ca). The TARGet Kids! Collaboration is a primary care practice–based research network and includes practice site physicians, research staff, collaborating investigators, trainees, methodologists, biostatisticians, data management personnel, laboratory management personnel, and advisory committee members.

---

**Extended Figure 1:**
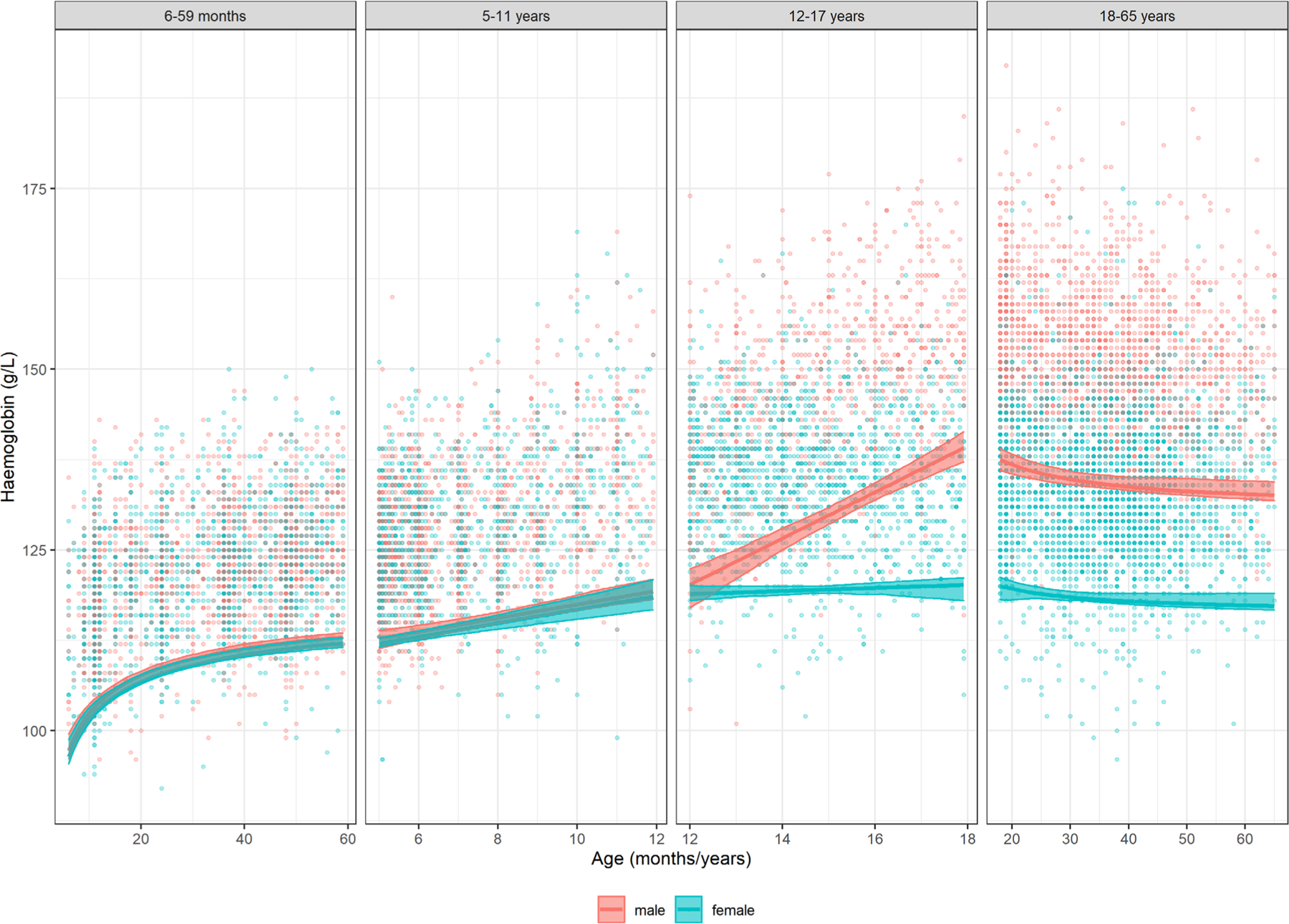
Continuous 5^th^ percentile thresholds across the life course. summary of continuous 5th percentile in males (red) and females (blue) across the life course (6 months to 65 years).

## References

1. Weatherall, D.J. & Hatton, C. Anaemia: pathophysiology, classification, and clinical features. in Oxford Textbook of Medicine (eds. Warrell, D.A., Cox, T.M. & Firth, J.D.) (Oxford University Press, Oxford, 2013).

2. World Health Organisation. Haemoglobin concentrations for the diagnosis of anaemia and assessment of severity. in Vitamin and Mineral Nutrition Information System (World Health Organization, Geneva, 2011).

3. World Health Organisation. Iron Deficiency Anaemia - Report of a study group, (World Health Organization, Geneva, 1959).

4. World Health Organisation. Nutritional Anaemias: Report of a WHO Scientific Group. In Technical Report Series, Vol. 405 (Geneva, 1968).

5. World Health Organization. Nutritional Anaemias: Tools for Effective Prevention and Control, (World Health Organization, Geneva, 2017).

6. Pasricha, S.-R., Colman, K., Centeno-Tablante, E., Garcia-Casal, M.-N. & Peña-Rosas, J.-P. Revisiting WHO haemoglobin thresholds used to define anaemia in clinical medicine and public health. Lancet Haematology Accepted 19/12/2017(2018).

7. Colman, K.S., Wood, E.M., De La Salle, B., Stanworth, S.J. & Pasricha, S.R. Heterogeneous hemoglobin lower thresholds in clinical laboratories. Am J Hematol 93, E142–E144 (2018).

8. Pasricha, S.R., Tye-Din, J., Muckenthaler, M.U. & Swinkels, D.W. Iron deficiency. Lancet (2020).

9. Ko, C.W., et al. AGA Clinical Practice Guidelines on the Gastrointestinal Evaluation of Iron Deficiency Anemia. Gastroenterology 159, 1085–1094 (2020).

10. Munoz, M., et al. International consensus statement on the peri-operative management of anaemia and iron deficiency. Anaesthesia 72, 233–247 (2017).

11. Tan, Y.L. & Kidson-Gerber, G. Antenatal haemoglobinopathy screening in Australia. Med J Aust 204, 226–230 (2016).

12. Goldman, M., Yi, Q.L., Steed, T. & O’Brien, S.F. Changes in minimum hemoglobin and interdonation interval: impact on donor hemoglobin and donation frequency. Transfusion 59, 1734–1741 (2019).

13. Pasricha, S.R., McQuilten, Z.K., Keller, A.J. & Wood, E.M. Hemoglobin and iron indices in nonanemic premenopausal blood donors predict future deferral from whole blood donation. Transfusion 51, 2709–2713 (2011).

14. Anand, I.S. & Gupta, P. Anemia and Iron Deficiency in Heart Failure: Current Concepts and Emerging Therapies. Circulation 138, 80–98 (2018).

15. Musallam, K.M., et al. Preoperative anaemia and postoperative outcomes in non-cardiac surgery: a retrospective cohort study. Lancet 378, 1396–1407 (2011).

16. Caro, J.J., Salas, M., Ward, A. & Goss, G. Anemia as an independent prognostic factor for survival in patients with cancer: a systemic, quantitative review. Cancer 91, 2214–2221 (2001).

17. Sullivan, P.S., Hanson, D.L., Chu, S.Y., Jones, J.L. & Ward, J.W. Epidemiology of anemia in human immunodeficiency virus (HIV)-infected persons: results from the multistate adult and adolescent spectrum of HIV disease surveillance project. Blood 91, 301–308 (1998).

18. Roubinian, N.H., et al. Long-Term Outcomes Among Patients Discharged From the Hospital With Moderate Anemia: A Retrospective Cohort Study. Ann Intern Med 170, 81–89 (2019).

19. Warner, M.A., et al. Prevalence of and Recovery From Anemia Following Hospitalization for Critical Illness Among Adults. JAMA Netw Open 3, e2017843 (2020).

20. Stevens, G.A., et al. National, regional, and global estimates of anaemia by severity in women and children for 2000-19: a pooled analysis of population-representative data. Lancet Glob Health 10, e627–e639 (2022).

21. Kassebaum, N.J., et al. A systematic analysis of global anemia burden from 1990 to 2010. Blood (2013).

22. World Health Organisation. Global nutrition targets 2025: policy brief series (WHO/NMH/NHD/14.2). (World Health Organization, Geneva, 2014).

23. United Nations. SDG Indicators - Metadata repository. Vol. 2023 (United Nations, New York, 2023).

24. WHO. Daily iron supplementation in children 6-23 months of age. in e-Library of Evidence for Nutrition Actions (eLENA), Vol. 2016 (World Health Organization, Geneva, 2016).

25. World Health Organization. WHO guideline: Use of multiple micronutrient powders for point-of-use fortication of foods consumed by infants and young children aged 6–23 months and children aged 2–12 years. (World Health Organization, Geneva, 2016).

26. Pasricha, S.R., et al. Net benefit and cost-effectiveness of universal iron-containing multiple micronutrient powders for young children in 78 countries: a microsimulation study. Lancet Glob Health 8, e1071–e1080 (2020).

27. Weyand, A.C., McGann, P.T. & Sholzberg, M. Sex specific definitions of anaemia contribute to health inequity and sociomedical injustice. Lancet Haematol 9, e6–e8 (2022).

28. Butcher, A., Richards, T., Stanworth, S.J. & Klein, A.A. Diagnostic criteria for pre-operative anaemia-time to end sex discrimination. Anaesthesia 72, 811–814 (2017).

29. Beutler, E. & Waalen, J. The definition of anemia: what is the lower limit of normal of the blood hemoglobin concentration? Blood 107, 1747–1750 (2006).

30. Beutler, E. & West, C. Hematologic differences between African-Americans and whites: the roles of iron deficiency and alpha-thalassemia on hemoglobin levels and mean corpuscular volume. Blood 106, 740–745 (2005).

31. Pasupula, D.K. & Reddy, P.S. When is a South Indian Really Anemic? Indian J Clin Biochem 29, 479–484 (2014).

32. Garcia-Casal, M.N., Pasricha, S.R., Sharma, A.J. & Pena-Rosas, J.P. Use and interpretation of hemoglobin concentrations for assessing anemia status in individuals and populations: results from a WHO technical meeting. Ann N Y Acad Sci 1450, 5–14 (2019).

33. PetitClerc, C. & Wilding, P. International Federation of Clinical Chemistry (IFCC), Scientific Committee, Clinical Section. The theory of reference values. Part 2. Selection of individuals for the production of reference values. J Clin Chem Clin Biochem 22, 203–208 (1984).

34. Shah, A., et al. Intravenous iron to treat anaemia following critical care: a multicentre feasibility randomised trial. Br J Anaesth 128, 272–282 (2022).

35. Sharma, A.J., Addo, O.Y., Mei, Z. & Suchdev, P.S. Reexamination of hemoglobin adjustments to define anemia: altitude and smoking. Ann N Y Acad Sci 1450, 190–203 (2019).

36. Addo, O.Y., et al. Evaluation of Hemoglobin Cutoff Levels to Define Anemia Among Healthy Individuals. JAMA Netw Open 4, e2119123 (2021).

37. (CLSI), C.a.L.S.I. Defining, establishing, and verifying reference intervals in the clinical laboratory; approved guideline. (Clinical and Laboratory Standards Institute (CLSI), Wayne, PA, 2008).

38. Tukey JW. Exploratory data analysis., (Addison-Wesley, 1977).

39. Daly, C.H., Higgins, V., Adeli, K., Grey, V.L. & Hamid, J.S. Reference interval estimation: Methodological comparison using extensive simulations and empirical data. Clin Biochem 50, 1145–1158 (2017).

40. Box GEP & Cox DR. An Analysis of Transformations. Journal of the Royal Statistical Society. Series B (Methodological). 26, 211–252 (1964).

41. Chernick MR. An introduction to bootstrap methods with applications to R, (Wiley, 2011).

42. Davison AC & D, H. Bootstrap Methods and their Application, (Cambridge University Press, Cambridge, 1997).

43. Canty AJ & Davison AC. Resampling-based Variance Estimation for Labour Force Surveys. Journal of the Royal Statistical Society: Series D (The Statistician) 48, 379–391 (1999).

44. Hoq, M., et al. Reference Values for 30 Common Biochemistry Analytes Across 5 Different Analyzers in Neonates and Children 30 Days to 18 Years of Age. Clin Chem 65, 1317–1326 (2019).

45. Royston P & EM, W. A method for estimating age-specific reference intervals (‘normal ranges’) based on fractional polynomials and exponential transformation. Journal of the Royal Statistical Society: Series A (Statistics in Society) 161, 79–101 (1998).

46. Chen, M.-H., et al. Trans-ethnic and Ancestry-Specific Blood-Cell Genetics in 746,667 Individuals from 5 Global Populations. Cell 182, 1198–1213.e1114 (2020).

47. Wheeler, M.M., et al. Whole genome sequencing identifies structural variants contributing to hematologic traits in the NHLBI TOPMed program. Nat Commun 13, 7592 (2022).

48. Finer, S., et al. Cohort Profile: East London Genes &amp; Health (ELGH), a community-based population genomics and health study in British Bangladeshi and British Pakistani people. International Journal of Epidemiology 49, 20–21i (2020).

49. Wang, Q., et al. Rare variant contribution to human disease in 281,104 UK Biobank exomes. Nature 597, 527–532 (2021).

50. Rare anaemia (Version 3.1). in PanelApp, Vol. 2023 (Genomics England 2023).

51. Karczewski, K.J., et al. The mutational constraint spectrum quantified from variation in 141,456 humans. Nature 581, 434–443 (2020).

52. Collins, R.L., et al. A structural variation reference for medical and population genetics. Nature 581, 444–451 (2020).

53. StataCorp. Stata Statistical Software: Release 16. (StataCorp LLC, College Station, RX, 2019).

54. R Core Team. R: A language and environment for statistical computing. (R Foundation for Statistical Computing, Vienna, Austria, 2021).

55. StataCorp. Stata Statistical Software: Release 17. (StataCorp LLC, College Station, RX, 2021).

56. Conway, J.R., Lex, A. & Gehlenborg, N. UpSetR: an R package for the visualization of intersecting sets and their properties. Bioinformatics 33, 2938–2940 (2017).

57. Gu, Z., Eils, R. & Schlesner, M. Complex heatmaps reveal patterns and correlations in multidimensional genomic data. Bioinformatics 32, 2847–2849 (2016).

58. Carsley, S., et al. Cohort Profile: The Applied Research Group for Kids (TARGet Kids!). Int J Epidemiol 44, 776–788 (2015).

59. Ramírez-Luzuriaga MJ., et al. Tomo I: Encuesta Nacional de Salud y Nutrición de la población ecuatoriana de cero a 59 años. ENSANUT-ECU 2012. (Ministerio de Salud Pública/Instituto Nacional de Estadísticas y Censos, Quito-Ecuador, 2014).

60. Jaddoe, V.W., et al. The Generation R Study: Design and cohort profile. Eur J Epidemiol 21, 475–484 (2006).

61. Hu, Y., et al. Whole-genome sequencing association analysis of quantitative red blood cell phenotypes: The NHLBI TOPMed program. Am J Hum Genet 108, 874–893 (2021).

62. Wheeler, M.M., et al. Whole genome sequencing identifies structural variants contributing to hematologic traits in the NHLBI TOPMed program. Nat Commun 13, 7592 (2022).

63. Mancera-Soto, E., et al. Quantification of testosterone-dependent erythropoiesis during male puberty. Exp Physiol 106, 1470–1481 (2021).

64. Sachdev, H.S., et al. Haemoglobin thresholds to define anaemia in a national sample of healthy children and adolescents aged 1-19 years in India: a population-based study. Lancet Glob Health 9, e822–e831 (2021).

65. Vareta, J., et al. Submicroscopic malaria infection is not associated with fever in cross-sectional studies in Malawi. Malar J 19, 233 (2020).

66. Ochwedo, K.O., et al. Hyper-prevalence of submicroscopic Plasmodium falciparum infections in a rural area of western Kenya with declining malaria cases. Malar J 20, 472 (2021).

67. Broor, S., et al. A prospective three-year cohort study of the epidemiology and virology of acute respiratory infections of children in rural India. PLoS One 2, e491 (2007).

68. Pasricha, S.R., et al. Benefits and Risks of Iron Interventions in Infants in Rural Bangladesh. N Engl J Med 385, 982–995 (2021).

69. de Onis, M., et al. The WHO Multicentre Growth Reference Study: planning, study design, and methodology. Food Nutr Bull 25, S15–26 (2004).

70. Papageorghiou, A.T., et al. International standards for fetal growth based on serial ultrasound measurements: the Fetal Growth Longitudinal Study of the INTERGROWTH-21st Project. Lancet 384, 869–879 (2014).

71. Australian Bureau of Statistics. Australian Health Survey: Users’ Guide, 2011-13 Vol. 2022 (Australian Bureau of Statistics, 2017).

72. UNC Chapel Hill. China Health and Nutrition Survey. Vol. 2022 (North Carolina).

73. Instituto Nacional de Estadística y Censos. Encuesta Nacional de Salud, Salud Reproductiva y Nutrición (ENSANUT)-2012. Vol. 2022 (Instituto Nacional de Estadística y Censos, Quito, 2023).

74. NHS Digital. Health Survey for England. Vol. 2022 (NHS Digital, United Kingdom, 2022).

75. National Center for Health Statistics. National Health and Nutrition Examination Survey. Vol. 2022 (Centers for Disease Control and Prevention,, 2023).

