## Supplemental Material for "Statistical haemoglobin thresholds to define anaemia across the lifecycle"

### 1. Database Search Strategy

We focused our initial search on National health or nutrition databases from countries that are part of The Organisation for Economic Co-operation and Development (OECD). Our search terms included “country” AND “national health database”. Databases were required to have venous haemoglobin, ferritin, and inflammation biomarker C-Reactive Protein (CRP), as well as clinical information to ensure that an apparently healthy reference sample could be selected. National Health Surveys of England (Health Survey for England [HSE]), United States of America (National Health and Nutrition Examination Survey [NHANES]), and Australia (Australian Health Survey [AHS]) met these criteria and were included in our study. A National Health Survey from New Zealand was investigated but excluded as it did not contain ferritin, while data from the German Health Interview and Examination Survey for Adults (DEGS) could not be included due to limitations in quality of the haemoglobin point-of-care measurement. Additionally, we were unable to make contact to obtain data from National Health Surveys in Portugal, and were unable to access data from the Canadian Health Measures Survey, due to regulatory requirements for data access. We were unable to identify databases that met our requirements from National surveys in Austria, Chile, Costa Rica, the Czech Republic, Denmark, Estonia, Finland, France, Greece, Hungary, Iceland, Ireland, Israel, Italy, Japan, Latvia, Lithuania, Luxembourg, Mexico, Norway, Poland Slovak Republic, and Sweden.

We also searched for prospective cohort or clinical trial studies from countries with a low background inflammation prevalence, that measured venous haemoglobin, ferritin, and CRP, and collected detailed health outcomes which would enable identification of a healthy reference sample, particularly in children and in pregnancy. This identified studies from Canada (The Applied Research Group for Kids [TARGet Kids!]) and The Netherlands (Generation R), which were included in our analyses. We obtained data from an Australian cohort study (HAPPI Kids) but excluded it upon learning that either ferritin or haemoglobin were measured in individuals, but not both. We expanded our search to non-OECD countries, and identified National Health Surveys from China (China Health and Nutrition Survey [CHNS]) and Ecuador (Encuesta Nacional de Salud y Nutrición [ENSANUT]), which met our criteria. Finally, we identified and included data from a clinical trial of iron supplemented infants in Bangladesh (Benefits and Risks of Iron interventions in Children [BRISC]) in our analyses.

### 2. Methods

#### 2.1 Approach to exclusion criteria to define healthy reference population

For each data source, the healthy reference sample comprised individuals who were not excluded from the study sample based on pre-defined criteria for an apparently healthy individual, defined separately for children (aged 6 months to 18 years), adults aged 18-65 years (non-pregnant) and pregnant women aged 18-45 years.

We applied the criteria to population studies, some of which has been repeated over multiple waves, in order to define a post-hoc healthy reference population. Surveys were ineligible if basic health data along with laboratory measurements (haemoglobin, ferritin, CRP at a minimum) were not available. Surveys were conducted in different countries (e.g. England, Australia, United States [US], Ecuador, China), and in some cases, the same survey platform (e.g. NHANES) was applied over multiple years. Each survey platform, as well as different years of the same platform, collected different information on health outcomes and health status, and some had a specific emphasis (e.g. on "cardiovascular health"). If more detailed criteria were available within an individual survey, these were generally applied (especially regarding health and medication use) even when this caused inconsistencies in criteria between surveys.

The general principles underpinning these inclusion and exclusion criteria are outlined below:

#### 2.2 Demographics

Where available and applicable, data was collected from males and females. Across all surveys, ages 6 months to 65 years were analysed, however the age range in each survey varied, and in many cases varied between years within the same survey platform. A separate analysis of pregnant women was performed, and pregnant women were excluded from the non-pregnant analysis where applicable (i.e. females in the age-groups of 12-17 years and 18-65 years).

Race/ethnicity/ancestry information was collected for each survey, although inconsistent terminology and reported categories were reported between survey platforms. In some surveys (children and adult groups) participants were able to select more than one ancestry group – in these cases, participants were considered as belonging to both ancestry groups. These data were used to describe the demographic of the groups included, but not used as exclusion criteria.

Data regarding other relevant household demographic information (including remoteness, household income/deciles, or poverty ratio) were also collected where available. None of these criteria were used as exclusion criteria for any of the analyses.

#### 2.3 Environmental and Nutritional

The surveys selected for this analysis were performed in countries with low rates of endemic infection. We excluded individuals living above 750m where individual altitude of residence was provided (Ecuador) or in provinces which were known to be elevated (China). Only one study (TARGet Kids!) evaluated exposure to lead (these children were excluded); no surveys asked about other toxins.

Body Mass Index (BMI) was used to exclude those who are underweight or at risk of malnutrition ($BMI<18.5kg/m^2$) and those categorised as obese ($BMI>30kg/m^2$).

Participants who identified themselves as a current smoker were excluded. This exclusion criteria applied to any method of smoking including cigarettes, pipes, and cigars (questioned specifically in most surveys).

Participants were excluded if they reported excessive alcohol use defined using US Centers for Disease Control and Prevention (CDC) recommendations. The US CDC guidelines were chosen to use across all surveys as they aligned with much of Europe, and were the highest recommended limits in any survey country. The US CDC criteria were applied to all surveys as follows (noting that a standard alcoholic drink contains 14g alcohol as defined in the US):

- No more than 1 drink per day for women, or 2 drinks per day for men, on a regular basis AND
- No more than 4 drinks on any one day for women, and no more than 8 drinks on any one day for men

Questions on illicit drug use were not asked, or responses were unavailable, for most surveys.

#### 2.4 Recent Health

All adult surveys comprised population community surveys. Childrens' surveys included children selected as healthy by their routine care provider.

Participants who reported any recent illness in the time period preceding the survey were excluded. All general surveys included questions regarding recent acute illness, although the preceeding time period varied between surveys, with some queried illness within 2 weeks, and some within 1 month. Some surveys asked about 'any' illness, where others specified coughs, colds, pneumonia, gastrointestinal illness.

Any participant who had been admitted to hospital in the last 12 months, or those who reported traumatic injury, major bleeding, pregnancy or childbirth within the preceding 12 months, was excluded where this information was available. Where data were available, participants who had recently donated blood were also excluded.

#### 2.5 General Health

All surveys included questions regarding participants medical history and comorbidities, and these questions were used to exclude participants with significant health conditions. Where possible, participants weren't excluded for resolved health conditions (e.g. historical thyroid disease) or minor issues not expected to contribute to anaemia (e.g. astigmatism). However where specific details were not available, participants who identified themselves as having any health condition were excluded. Outside of questions about general health or "diseases of blood", no survey asked specifically about history of haemoglobinopathy.

In general, all participants requiring any regular prescription medication were excluded. In two surveys, it was possible to exclude all except people on hormonal contraception. Where possible, participants taking supplements or vitamins were included, apart from those specifically on supplementation of haematinics (iron, B12, folate).

In children, we excluded all reporting any use of medications (including over-the-counter medications).

#### 2.6 Laboratory Criteria

For this analysis, surveys were only included where haemoglobin, ferritin, and CRP results were available. In some surveys, specific years targeted specific populations (children, women, pregnant or non pregnant adults) for some testing. If an individual had missing data for any of these parameters, they were excluded from this analysis.

Ferritin limits were set according to the "WHO guidelines on use of ferritin concentrations to assess iron status in individuals and populations",[1] Table 1, for "apparently healthy individuals":

- Age less than 5 years old: ferritin <12ug/L defines iron deficiency
- Age greater than 5 years old, including pregnant women in first trimester: ferritin <15ug/L defines iron deficiency; and ferritin >150ug/L in females, or >200ug/L in males, defines risk of iron overload

A CRP cut off of <5mg/L was used to exclude those with current inflammation.

No criteria were applied to haemoglobin, or any other full blood examination parameters. Mean Corpuscular Volume (MCV) was collected where available, but was not available for most surveys, and no criteria were based on this measurement except for sensitivity analyses in children as described.

Where available, abnormal renal function was an exclusion criteria (eGFR<90ml/min/1.73m$^2$).

Estimated Glomerular Filtration Rate (eGFR) was used to determine if non-pregnant adult participants and pregnant women had kidney disease in the AHS, HSE (relevant data available in year 2009 only), NHANES and CHNS. An eGFR less than 90 ml/min/1.73 m$^2$ was considered an indicator of abnormal kidney function in adults and pregnant women.

The eGFR (ml/min/1.73 m$^2$) was calculated by Chronic Kidney Disease Epidemiology (CKD-EPI)[2] equation using serum creatinine (mg/dL), age (years), and sex (male/female) without race adjustment.[2,3] This formula was used for adults and pregnant women, noting that in pregnancy there is no accepted eGFR calculation or range.

$$eGFR = 142 \times min\left(\frac{creatinine}{K}, 1\right)^{\alpha} \times max\left(\frac{creatinine}{K}, 1\right)^{-1.2} \times 0.9938^{Age} \times SexFactor$$

Where creatinine = serum creatinine (mg/dL), K=0.7 for females or 0.9 for males, α = -0.241 for females or -0.302 for males, min = minimum, max = maximum, age = age (years), and SexFactor

= 1.012 for females and 1 for males. The formula was implemented and calculations were validated by randomly checking eGFR derivations with an online calculator https://www.kidney.org/professionals/kdoqi/gfr_calculator (accessed March 2022) based on the CKD-EPI equation. For some NHANES survey years, serum creatinine as provided required a conversion in units (from µmol/L to mg/dL by multiplying by 1/88.4) and/or calibration due to use of non-standardised laboratory methods before deriving eGFR.

For children, we did not implement an exclusion criterion for eGFR because the rates of undiagnosed chronic kidney disease in children with no other comorbidities are likely be very low, and creatinine was not consistently available across surveys.

Where available, abnormal liver function tests were also used to exclude participants.

#### 2.7 Pregnancy – Additional criteria

Only two surveys included adequate numbers of pregnant participants for analysis – NHANES, and Generation R. Data from pregnant participants were excluded based on all available general health parameters as above where available; with an additional set of criteria to exclude those with pregnancy related illness or complications.

In NHANES, where pregnant participants were a subset of the general survey, the same criteria for health were applied as for other adults. An altered BMI range was used, and all available questions regarding pregnancy status (month of pregnancy) and pregnancy complications (gestational diabetes, and hypertension during pregnancy) were used as exclusion criteria.

Generation R was a targeted study of pregnant women. Fewer questions regarding general health were available, with more data available about pregnancy associated health problems.

Where possible, only those with singleton pregnancies were included. Those with gestational diabetes, pregnancy induced hypertension, pre-eclampsia, or HELLP syndrome were excluded. Pregnancy related cardiomyopathy or other serious complications were excluded.

#### 2.8 Children – Additional criteria

In children, many of the same parameters were sought to identify the reference population as in adults. Additionally, in those under 2 years old, any child noted to have had birth complications was excluded (such as premature birth or low birth weight, NICU admission).

Any child with active health problems was excluded. This included children with potentially minor conditions (e.g. eczema or allergies), however sufficient information on severity was not generally available and thus a restrictive approach as adopted to ensure exclusion of any condition that may affect general health, nutrition, and growth, or that may cause inflammation.

#### 2.9 Statistical Analyses

Individual demographic, clinical, laboratory and haematology data were accessible to the authors for all data sources. Individual data of national population health surveys HSE (web URL: beta.ukdataservice.ac.uk/datacatalogue/series/series?id=2000021), NHANES (web URL: cdc.gov/nchs/nhanes), CHNS (web URL: cpc.unc.edu/projects/china), and ENSANUT (web URL: salud.gob.ec/encuesta-nacional-de-salud-y-nutricion-ensanut/) were downloaded from the relevant websites. Individual data of the randomised controlled BRISC trial (web URL: wehi.edu.au/people/sant-rayn-pasricha/trials/brisc) and the prospective population-based longitudinal cohort study TARGet Kids! (web URL: targetkids.ca) were available to the authors. Individual data of AHS (web URL: abs.gov.au/ausstats/abs@.nsf/mf/4363.0.55.001), consisting of National Health Survey [NHS] and National Nutrition and Physical Activity Survey [NNPAS], was only accessible by the joint first authors through the Australian Bureau of Statistics (ABS) DataLab environment. Analysis output (e.g., centiles) could be retrieved outside of the secure DataLab for public use only after clearance for release from the ABS. Finally, individual data of the prospective population-based longitudinal cohort Generation R study (web URL: generationr.nl) was accessible to the Generation R investigators while analysis output (e.g., centiles) was available to the authors. Further citation details on HSE, NHANES, and AHS are provided in the table below.

| Data Source | Citation |
|---|---|
| HSE | – *Survey 1998*: National Centre for Social Research, University College London. Department of Epidemiology and Public Health. (2010). Health Survey for England, 1998. [data collection]. 5th Release. UK Data Service. SN: 4150, DOI: http://doi.org/10.5255/UKDA-SN-4150-1<br>– *Survey 2006*: National Centre for Social Research, University College London, Department of Epidemiology and Public Health. (2011). Health Survey for England, 2006. [data collection]. 4th Edition. UK Data Service. SN: 5809, DOI: http://doi.org/10.5255/UKDA-SN-5809-1<br>– *Survey 2009*: University College London, Department of Epidemiology and Public Health, National Centre for Social Research. (2015). Health Survey for England, 2009. [data collection]. 3rd Edition. UK Data Service. SN: 6732, DOI: http://doi.org/10.5255/UKDA-SN-6732-2 |
| NHANES | Centers for Disease Control and Prevention (CDC). National Center for Health Statistics (NCHS). National Health and Nutrition Examination Survey Data. Hyattsville, MD: U.S. Department of Health and Human Services, Centers for Disease Control and Prevention, [appropriate year] [web URL] as per below:<br>– [1999-2000] [wwwn.cdc.gov/nchs/nhanes/continuousnhanes/default.aspx?BeginYear=1999]<br>– [2001-2002] [wwwn.cdc.gov/nchs/nhanes/continuousnhanes/default.aspx?BeginYear=2001]<br>– [2003-2004] [wwwn.cdc.gov/nchs/nhanes/continuousnhanes/default.aspx?BeginYear=2003]<br>– [2005-2006] [wwwn.cdc.gov/nchs/nhanes/continuousnhanes/default.aspx?BeginYear=2005]<br>– [2007-2008] [wwwn.cdc.gov/nchs/nhanes/continuousnhanes/default.aspx?BeginYear=2007]<br>– [2009-2010] [wwwn.cdc.gov/nchs/nhanes/continuousnhanes/default.aspx?BeginYear=2009]<br>– [2015-2016] [wwwn.cdc.gov/nchs/nhanes/continuousnhanes/default.aspx?BeginYear=2015]<br>– [2017-2018] [wwwn.cdc.gov/nchs/nhanes/continuousnhanes/default.aspx?BeginYear=2017] |
| AHS | – Australian Bureau of Statistics (2011-12) National Health Survey, Australia [DataLab] (accessed April 2022-23)<br>– Australian Bureau of Statistics (2012-13) National Nutrition and Physical Activity Survey, Australia [DataLab] (accessed April 2022-23) |

HSE = Health Survey for England; NHANES = National Health and Nutrition Examination Survey; AHS = Australian Health Survey (consisting of National Health Survey and National Nutrition and Physical Activity Survey).

##### 2.9.1 General Principles

All analyses used haemoglobin values as provided by the data source and were reported in g/L. The estimates of the 2.5th and 5th centiles along with confidence intervals were obtained for the healthy reference sample for each data sources. The Clinical & Laboratory Standards Institute recommends two-sided 90% confidence intervals be calculated for each reference limit.[4]

With the exception of TARGet Kids!, a single haemoglobin value was available by age-group and sex or trimester of pregnancy. The TARGet Kids! study was a longitudinal cohort study with participants followed between birth to adolescence. Children under 6 years of age were eligible to be included in the study if reported healthy by their parents (including children with asthma). Given that health was central to the estimation of the centiles, only the haemoglobin measurement from the first study visit per participant was used to estimate the centile.

All analyses were run with and without outliers. Outliers were identified using Tukey's method.[5,6] For discrete threshold estimation, outlying values were haemoglobin values < 25th haemoglobin centile – 3 × interquartile range (IQR) and haemoglobin values > 75th haemoglobin centile + 3 × IQR, whereby IQR is the difference between the 75th and 25th centiles. For continuous threshold estimation, haemoglobin in the above description was replaced by the residuals from multivariable fractional polynomial regression of haemoglobin on age after adjusting for sex. Results for the discrete and continuous thresholds are presented excluding outlying values only.

##### 2.9.2 Sample Size

For each data source, the sample size of the healthy reference sample was determined by the number of participants who fulfilled the criteria of an apparently healthy individual. For children in the age-groups of 6-23 months, 24-59 months, and 5-11 years, the available data or all NHANES survey cycles were pooled due to the small sample size by survey cycle. For the Australian NHS and NNPAS, no centiles could be extracted if the sample size was less than 200 (2.5th centile) or 100 (5th centile) as per ABS requirements for output clearance. For children 12-17 years, NHS and NNPAS were pooled to meet this sample size requirement for output clearance. For all other data sources, centiles were not obtained if the sample size was less than 20. No hypothesis testing was undertaken.

##### 2.9.3 Survey Weighting and Design

In national population health surveys (NHANES, HSE, CHNS, ENSANUT), we aimed to account for the sampling weights and sampling design during the analysis. Accounting for the survey design (i.e. cluster and/or stratum to which the individual belongs) is required to correctly estimate standard errors and correct inference. The weighting adjusts the results from a sample survey to infer results for the total target population. To do this, a 'weight' is allocated to each sample unit (person or household), indicating how many population units are represented by the sample unit. In general, the weights account for selection and non-response bias. In the table below, the design, cluster (primary sampling unit) and strata, and weight variables used in centile estimation described later are provided.

Design and weight variables used in centile estimation for each national population health survey

| **Variable Type[1]** | **Variable Name[3]** | **Description** |
|---|---|---|
| *NHANES (Survey 1999-2000, 2001-02, 2003-04, 2005-06, 2007-08, 2009-10, 2015-16, 2017-18)* | | |
| Cluster/PSU[2] | sdmvpsu | County |
| Strata | sdmvstra | Geography (e.g., census region), metropolitan statistical area status, and various population demographics. |
| Weights | wtmec2yr | Generated for individuals who had physical examinations and laboratory tests in mobile examination centers (MEC). |
| *HSE (Survey 2006 and 2009; no design and weight variables applicable for 1998)* | | |
| Cluster/PSU | psu | Postcode sectors |
| Strata | cluster | Local authority and socio-economic group |
| Weights | wt_blood | Generated for individuals aged 16 and over who had a nurse visit and were eligible for a blood sample. |
| *CHNS (Survey 2009)* | | |
| Cluster/PSU | commid | Community/county |
| Strata | N/A[4] | Information unavailable/not released to the public. |
| Weights | N/A | Information unavailable/not released to the public. |
| *ENSANUT (Survey 2011-13)* | | |
| Cluster/PSU | idsector | Census tracts |
| Strata | area | Rural and urban areas |
| Weights | pw | Household expansion factor/weight |

[1]Survey – National population health survey; [2]PSU – Primary sampling unit; [3]Variable Name – Name of variable in dataset; [4]N/A – Not available.

Summary statistics of participant characteristics (Supplemental Material Section 3) and violations of exclusion criteria (Supplemental Material Section 4) were presented unweighted for the national population health surveys.

##### 2.9.4 Discrete Thresholds

The estimates of the 2.5$^{th}$ and 5$^{th}$ discrete centiles along with confidence intervals were obtained for each data source by age-group (6-23 months, 24-59 months, 5-11 years), age-group and sex (12-17 years, 18-65 years), or trimester of pregnancy (pregnant women [18-45 years]).

Simple Random Samples

The parametric theoretical centile of a Gaussian distribution, with corresponding 90% confidence interval, was used to estimate the centile.[7] In addition, we applied a Box-Cox transformation to the reference distribution prior to calculating the theoretical centile and corresponding confidence interval.[8] The simulation study conducted in Daly et al.,[7] found that the parametric approach provided the least biased and most precise estimates for normally distributed reference distributions compared to the other methods examined (non-parametric and robust). Since the reference distribution appeared reasonably normal, the parametric method without Box-Cox transformation was utilised as the preferred method instead of non-parametric and robust methods.

Complex Surveys

For the national population health surveys NHANES, HSE, CHNS, and ENSANUT both unweighted and survey-weighted centile estimates were calculated. Unweighted centile estimates were calculated using the parametric theoretical centile. Survey-weighted centile estimates were calculated using survey-weighted quantile regression. The implementation of survey-weighted quantile regression proceeded the same way as described in Section 2.9.5, except the survey-weighted quantile regression model only included a single intercept term. Standard errors for the intercept term were derived from samples generated using Canty and Davidson's bootstrap and two-sided 90% confidence intervals derived assuming a normal approximation.[9]

Centile estimation from complex survey using the survey-weighted approach implemented by R's svyquantile function was also explored. The 90% confidence intervals for the survey weighted centile estimates were calculated using Woodruff's method,[10] and involves calculating the quantile from the empirical cumulative distribution function (ECDF), estimating a confidence interval for the proportion of values below the quantile, and then transforming that interval using the ECDF. For small centiles (e.g., 2.5$^{th}$, and occasionally 5$^{th}$) and small to moderate sample sizes, the lower limit of the 90% confidence interval could not be calculated. Since this problem was not observed for quantile regression, it was considered the preferred method to obtain the survey-weighted centile estimate while the unweighted centile estimate was obtained to explore the sensitivity of the results.

Pooling

Centile estimates (discrete thresholds) pooled across all data sources using the combined healthy reference sample were calculated using fixed effect and random effects meta-analyses. The fixed effect approach assumes that the variation in the centile estimates is due to chance alone, and the pooled centile estimate is a weighted average of the centile estimates with weights equal to the square of the inverse of the standard error of the estimate. The random effects approach assumes that the variation in the centile estimates is due to more than chance alone, and that between study differences are also contributing. Like the fixed effects approach, the pooled centile estimate from the random effects approach is a weighted average of the centile estimates, but with weights equal to inverse of the total variation (between study variation + within study/residual variation). The fixed and random effects meta-analyses were performed in R using the metafor package.[11]

##### 2.9.5 Continuous Thresholds

The estimates of the 2.5$^{th}$ and 5$^{th}$ continuous centiles along with confidence intervals were obtained for all data sources combined (excluding NHS and NNPAS) by age-group (6-23 months, 24-59 months, 5-11 years, 12-17 years, 18-65 years) (and sex if applicable). NHS and NNPAS data could not be included because individual data was only accessible via the ABS secure DataLab. No continuous thresholds were obtained for pregnant women.

Simple Random Samples

The method proposed in Hoq et al. 2019[12] was used to estimate age-specific continuous haemoglobin thresholds. In brief, Hoq's method implements the following steps:

1. Identify the best fitting multivariable fractional polynomial (MFP) model for the mean values of haemoglobin using Royston's method (8).[13]
2. Likelihood ratio test for an interaction between sex and the fractional polynomial representation of age in the model identified in step 1. Include the age-by-sex interaction term(s) in the model if they are statistically significant at the nominal level of significance of 0.05.
3. Quantile regression was used. The terms included in the quantile regression model were constrained to be the same as those included in the MFP model of the mean (steps 1 and 2). Age-dependent haemoglobin predictions were obtained from the quantile regression model and two-sided 90% bootstrap centile confidence intervals.[14,15] 1000 bootstrap samples were generated.

The residuals from the MFP were examined to assess the fit of the MPF model for the mean haemoglobin values and to identify outliers using Tukey's method.[6] The results from quantile regression modelling were presented in plots showing how the predicted centiles change with age (and by sex if applicable), superimposed onto a scatter plot of haemoglobin versus age (in years or months).

Complex Surveys

To estimate continuous haemoglobin thresholds from national population health surveys NHANES, HSE, CHNS, and ENSANUT, the above steps for simple random samples were performed with the following modifications:

– In step 3 above, survey-weighted quantile regression was performed.

– Canty and Davidson's bootstrap[9] was used to generate bootstrap samples from which two-sided 90% confidence intervals for the predicted centiles were derived using the centile bootstrap method.[14,15]

Pooling

Centile estimates (continuous thresholds) were obtained using the method described under "Simple Random Samples" using the combined healthy reference sample across national population health surveys (NHANES, HSE, CHNS, ENSANUT) and other studies (BRISC, TARGet Kids!).

##### 2.9.6 Software

Reference samples were derived using Stata version 16.1,[16] R version 4.1.1 (Generation R),[17] or Stata version 17.1 (NHS and NNPAS).[18] Estimation of the haemoglobin thresholds was performed using R version 4.2.3,[17] R version 4.1.1 (Generation R),[17] or Stata version 17.1 (NHS and NNPAS).[18] An overview of the R packages used as part of the discrete and continuous threshold estimation as well as the pooling are listed in the table below. In Stata, the command *centile* with option normal was used to obtain the discrete thresholds.

Overview of R packages used as part of the threshold estimation.

| **Package** | **Citation** |
|---|---|
| tidyverse | Wickham et al., (2019). Welcome to the tidyverse. Journal of Open Source Software, 4(43), 1686. https://doi.org/10.21105/joss.01686 |
| magrittr | Stefan Milton Bache and Hadley Wickham (2022). magrittr: A Forward-Pipe Operator for R. R package version 2.0.3. https://CRAN.R-project.org/package=magrittr |
| haven | Hadley Wickham, Evan Miller and Danny Smith (2022). haven: Import and Export 'SPSS', 'Stata' and 'SAS' Files. R package version 2.5.0. https://CRAN.R-project.org/package=haven |
| ggpubr | Alboukadel Kassambara (2020). ggpubr: 'ggplot2' Based Publication Ready Plots. R package version 0.4.0. https://CRAN.R-project.org/package=ggpubr |
| cowplot | Claus O. Wilke (2020). cowplot: Streamlined Plot Theme and Plot Annotations for 'ggplot2'. R package version 1.1.1. https://CRAN.R-project.org/package=cowplot |
| MKinfer | Kohl M (2020). _MKinfer: Inferential Statistics_. R package version 0.6 |

| | |
|---|---|
| | http://www.stamats.de |
| quantreg | Roger Koenker (2022). quantreg: Quantile Regression. R package version 5.88. https://CRAN.R-project.org/package=quantreg |
| survey | T. Lumley (2004) Analysis of complex survey samples. Journal of Statistical Software 9(1): 1-19 T. Lumley (2010) Complex Surveys: A Guide to Analysis Using R. John Wiley and Sons. |
| jtools | Long JA (2020). _jtools: Analysis and Presentation of Social Scientific Data_. R package version 2.1.0. https://cran.r-project.org/package=jtools |
| moments | Lukasz Komsta and Frederick Novomestky (2015). Moments: Moments, cumulants, skewness, kurtosis and related tests. R package version 0.14. https://CRAN.R-project.org/package=moments |
| kableExtra | Hao Zhu (2021). kableExtra: Construct Complex Table with 'kable' and Pipe Syntax. R package version 1.3.4. https://CRAN.R-project.org/package=kableExtra |
| mfp | Gareth Ambler and Axel Benner (2022). mfp: Multivariable Fractional Polynomials. R package version 1.5.2.2. https://CRAN.R-project.org/package=mfp |
| lmtest | Achim Zeileis, Torsten Hothorn (2002). Diagnostic Checking in Regression Relationships. R News 2(3), 7-10. https://CRAN.R-project.org/doc/Rnews/ |
| referenceIntervals | Daniel Finnegan (2020). referenceIntervals: Reference Intervals. R package version 1.2.0. https://CRAN.R-project.org/package=referenceIntervals |
| car | John Fox and Sanford Weisberg (2019). An R Companion to Applied Regression, Third Edition. Thousand Oaks CA: Sage. https://socialsciences.mcmaster.ca/jfox/Books/Companion/ |
| Ecfun | Spencer Graves (2021). Ecfun: Functions for 'Ecdat'. R package version 0.2-5. https://CRAN.R-project.org/package=Ecfun |
| moments | Lukasz Komsta and Frederick Novomestky (2015). moments: Moments, cumulants, skewness, kurtosis and related tests. R package version 0.14. https://CRAN.R-project.org/package=moments |
| nortest | Juergen Gross and Uwe Ligges (2015). nortest: Tests for Normality. R package version 1.0-4. https://CRAN.R-project.org/package=nortest |
| metafor | Viechtbauer, W. (2010). Conducting meta-analyses in R with the metafor package. Journal of Statistical Software, 36(3), 1-48. https://doi.org/10.18637/jss.v036.i03 |
| readxl | Wickham H, Bryan J (2022). _readxl: Read Excel Files_. R package version 1.4.0. https://CRAN.R-project.org/package=readxl |

#### 2.10 Ethics Approvals

Each individual data source obtained ethics approval before conduct of the study/survey, as outlined in the table below.

| Data Source | Ethics |
|---|---|
| TARGet Kids! | The study was approved by the Research Ethics Board of The Hospital for Sick Children, Toronto, Ontario, Canada. |
| Generation R | The general design, all research aims and the specific measurements in the Generation R Study have been approved by the Medical Ethical Committee of Erasmus MC, University Medical Center Rotterdam, The Netherlands. |
| AHS | The AHS biomedical measures component forms the National Health Measures Survey (NHMS). Ethics approval for the NHMS was granted by the Australian Government Department of Health and Ageing Departmental Ethics Committee in February 2011. |
| NHANES | The NHANES protocol was developed and reviewed to be in compliance with the Department of Health and Human Services (HHS) Policy for Protection of Human Research Subjects (45 CFR part 46), guidance/45cfr46.html. It was approved by the National Center for Health Statistics (NCHS), United States, Research Ethics Review Board (ERB) and underwent annual review. |
| HSE | Ethical approval was obtained from the London Multi-centre Research Ethics Committee, England. |
| BRISC | The trial was approved by the Melbourne Health Human Research Ethics Committee, Melbourne, Victoria, Australia (2016.269); the Ethical Review Committee of icddr,b (PR-16063); and the Directorate General of Drug Administration, Ministry of Health and Family Welfare, Dhaka, Bangladesh. |
| ENSANUT | This survey was run with oversight from the National Institute of Statistics and Censuses, Ecuador. |
| CHNS | This is an international collaborative project between the Carolina Population Center at the University of North Carolina at Chapel Hill (United States); and the Chinese Center for Disease Control and Prevention (China). |

TARGet Kids! = The Applied Research Group for Kids; AHS = Australian Health Survey (consisting of National Health Survey and National Nutrition and Physical Activity Survey); NHANES = National Health and Nutrition Examination Survey; HSE = Health Survey for England; BRISC = Benefits and Risks of Iron intervention in Children; ENSANUT = Encuesta Nacional de Salud y Nutrición; CHNS = China Health and Nutrition Survey.

### 3. Demographics

#### 3.1 Demographics Adults (18-65 years) males

##### Table 3.1.1 US NHANES 1999-2000: Participant characteristics (Males) (18-65 years)

| | **Overall sample N=1,673** | **Excluded sample N=1,491** | **Reference sample[1] N=182** |
|---|---|---|---|
| Age (years) | 38.9 (15.0) | 40.2 (14.8) | 28.2 (12.1) |
| Sex | | | |
| Male | 1,673/1,673 (100.0%) | 1,491/1,491 (100.0%) | 182/182 (100.0%) |
| BMI (kg/m$^2$) | 27.4 (5.6) | 27.8 (5.7) | 24.1 (3.1) |
| Poverty | 287/1,475 (19.5%) | 253/1,322 (19.1%) | 34/153 (22.2%) |
| Race and Hispanic Origin | | | |
| Mexican American | 521/1,673 (31.1%) | 434/1,491 (29.1%) | 87/182 (47.8%) |
| Other Hispanic | 95/1,673 (5.7%) | 84/1,491 (5.6%) | 11/182 (6.0%) |
| Non-Hispanic White | 662/1,673 (39.6%) | 610/1,491 (40.9%) | 52/182 (28.6%) |
| Non-Hispanic Black | 332/1,673 (19.8%) | 305/1,491 (20.5%) | 27/182 (14.8%) |
| Other Race - Including Multi-Racial | 63/1,673 (3.8%) | 58/1,491 (3.9%) | 5/182 (2.7%) |
| Haemoglobin (g/L) | 154.2 (11.0) | 154.0 (11.2) | 155.7 (9.2) |
| Anaemia (Haemoglobin<130g/L) | | | |
| No | 1,637/1,673 (97.8%) | 1,456/1,491 (97.7%) | 181/182 (99.5%) |
| Yes | 36/1,673 (2.2%) | 35/1,491 (2.3%) | 1/182 (0.5%) |
| Iron deficiency (Ferritin<15ug/L) | | | |
| No | 1,658/1,673 (99.1%) | 1,476/1,491 (99.0%) | 182/182 (100.0%) |
| Yes | 15/1,673 (0.9%) | 15/1,491 (1.0%) | 0/182 (0.0%) |
| Inflammation (C-Reactive Protein>5mg/L) | | | |
| No | 1,421/1,673 (84.9%) | 1,239/1,491 (83.1%) | 182/182 (100.0%) |
| Yes | 252/1,673 (15.1%) | 252/1,491 (16.9%) | 0/182 (0.0%) |

US = United States, NHANES = National Health and Nutrition Examination Survey, BMI = Body Mass Index, SD = Standard Deviation.
Data are presented as unweighted mean (SD) for continuous measures, and unweighted n/total (%) for categorical measures based on participants with non-missing haemoglobin, ferritin, and C-reactive protein.
[1] Ferritin≥15 and ≤200ug/L and C-reactive protein≤5mg/L.

#### Table 3.1.2 US NHANES 2001-2002: Participant characteristics (Males) (18-65 years)

| | Overall sample N=1,973 | Excluded sample N=1,733 | Reference sample[1] N=240 |
|---|---|---|---|
| Age (years) | 38.6 (14.5) | 40.0 (14.3) | 29.2 (11.8) |
| Sex | | | |
| Male | 1,973/1,973 (100.0%) | 1,733/1,733 (100.0%) | 240/240 (100.0%) |
| BMI (kg/m$^2$) | 27.5 (5.8) | 27.9 (5.9) | 24.3 (2.9) |
| Poverty | 315/1,856 (17.0%) | 265/1,630 (16.3%) | 50/226 (22.1%) |
| Race and Hispanic Origin | | | |
| Mexican American | 497/1,973 (25.2%) | 397/1,733 (22.9%) | 100/240 (41.7%) |
| Other Hispanic | 89/1,973 (4.5%) | 75/1,733 (4.3%) | 14/240 (5.8%) |
| Non-Hispanic White | 898/1,973 (45.5%) | 817/1,733 (47.1%) | 81/240 (33.8%) |
| Non-Hispanic Black | 415/1,973 (21.0%) | 378/1,733 (21.8%) | 37/240 (15.4%) |
| Other Race - Including Multi-Racial | 74/1,973 (3.8%) | 66/1,733 (3.8%) | 8/240 (3.3%) |
| Haemoglobin (g/L) | 154.1 (11.4) | 153.9 (11.7) | 155.6 (9.1) |
| Anaemia (Haemoglobin<130g/L) | | | |
| No | 1,919/1,973 (97.3%) | 1,679/1,733 (96.9%) | 240/240 (100.0%) |
| Yes | 54/1,973 (2.7%) | 54/1,733 (3.1%) | 0/240 (0.0%) |
| Iron deficiency (Ferritin<15ug/L) | | | |
| No | 1,939/1,973 (98.3%) | 1,699/1,733 (98.0%) | 240/240 (100.0%) |
| Yes | 34/1,973 (1.7%) | 34/1,733 (2.0%) | 0/240 (0.0%) |
| Inflammation (C-Reactive Protein>5mg/L) | | | |
| No | 1,678/1,973 (85.0%) | 1,438/1,733 (83.0%) | 240/240 (100.0%) |
| Yes | 295/1,973 (15.0%) | 295/1,733 (17.0%) | 0/240 (0.0%) |

US = United States, NHANES = National Health and Nutrition Examination Survey, BMI = Body Mass Index, SD = Standard Deviation.
Data are presented as unweighted mean (SD) for continuous measures, and unweighted n/total (%) for categorical measures based on participants with non-missing haemoglobin, ferritin, and C-reactive protein.
[1] Ferritin≥15 and ≤200ug/L and C-reactive protein≤5mg/L.

#### Table 3.1.3 US NHANES 2017-2018: Participant characteristics (Males) (18-65 years)

| | Overall sample N=1,854 | Excluded sample N=1,705 | Reference sample[1] N=149 |
|---|---|---|---|
| Age (years) | 42.7 (14.7) | 43.7 (14.5) | 31.4 (12.0) |
| Sex | | | |
| Male | 1,854/1,854 (100.0%) | 1,705/1,705 (100.0%) | 149/149 (100.0%) |
| BMI (kg/m$^2$) | 29.4 (6.7) | 29.8 (6.8) | 24.9 (3.1) |
| Poverty | 318/1,620 (19.6%) | 289/1,493 (19.4%) | 29/127 (22.8%) |
| Race and Hispanic Origin | | | |
| Mexican American | 302/1,854 (16.3%) | 271/1,705 (15.9%) | 31/149 (20.8%) |
| Other Hispanic | 177/1,854 (9.5%) | 156/1,705 (9.1%) | 21/149 (14.1%) |
| Non-Hispanic White | 567/1,854 (30.6%) | 542/1,705 (31.8%) | 25/149 (16.8%) |
| Non-Hispanic Black | 408/1,854 (22.0%) | 379/1,705 (22.2%) | 29/149 (19.5%) |
| Non-Hispanic Asian | 287/1,854 (15.5%) | 246/1,705 (14.4%) | 41/149 (27.5%) |
| Other Race - Including Multi-Racial | 113/1,854 (6.1%) | 111/1,705 (6.5%) | 2/149 (1.3%) |
| Haemoglobin (g/L) | 150.3 (12.2) | 150.2 (12.3) | 152.0 (10.4) |
| Anaemia (Haemoglobin<130g/L) | | | |
| No | 1,777/1,854 (95.8%) | 1,632/1,705 (95.7%) | 145/149 (97.3%) |
| Yes | 77/1,854 (4.2%) | 73/1,705 (4.3%) | 4/149 (2.7%) |
| Iron deficiency (Ferritin<15ug/L) | | | |
| No | 1,826/1,854 (98.5%) | 1,677/1,705 (98.4%) | 149/149 (100.0%) |
| Yes | 28/1,854 (1.5%) | 28/1,705 (1.6%) | 0/149 (0.0%) |
| Inflammation (C-Reactive Protein>5mg/L) | | | |
| No | 1,537/1,854 (82.9%) | 1,388/1,705 (81.4%) | 149/149 (100.0%) |
| Yes | 317/1,854 (17.1%) | 317/1,705 (18.6%) | 0/149 (0.0%) |

US = United States, NHANES = National Health and Nutrition Examination Survey, BMI = Body Mass Index, SD = Standard Deviation.
Data are presented as unweighted mean (SD) for continuous measures, and unweighted n/total (%) for categorical measures based on participants with non-missing haemoglobin, ferritin, and C-reactive protein.
[1] Ferritin≥15 and ≤200ug/L and C-reactive protein≤5mg/L.

#### Table 3.1.4 England HSE 1998: Participant characteristics (Males) (18-65 years)

| | Overall sample N=3,835 | Excluded sample N=3,268 | Reference sample[1] N=567 |
|---|---|---|---|
| Age (years) | 42.3 (12.5) | 42.8 (12.6) | 39.7 (11.5) |
| Sex | | | |
| Male | 3,835/3,835 (100.0%) | 3,268/3,268 (100.0%) | 567/567 (100.0%) |
| BMI (kg/m$^2$) | 26.6 (4.0) | 26.8 (4.1) | 25.4 (2.5) |
| Income | | | |
| bottom quintile (≤£7,186) | 414/3,408 (12.1%) | 363/2,900 (12.5%) | 51/508 (10.0%) |
| 2nd quintile (>£7,186 ≤ £10,834) | 406/3,408 (11.9%) | 349/2,900 (12.0%) | 57/508 (11.2%) |
| 3rd quintile (>£10,834 ≤ £17,890) | 809/3,408 (23.7%) | 684/2,900 (23.6%) | 125/508 (24.6%) |
| 4th quintile (>£17,890 ≤ £27,705) | 858/3,408 (25.2%) | 739/2,900 (25.5%) | 119/508 (23.4%) |
| highest quintile (>£27,705) | 921/3,408 (27.0%) | 765/2,900 (26.4%) | 156/508 (30.7%) |
| Ethnic group | | | |
| White | 3,608/3,833 (94.1%) | 3,094/3,266 (94.7%) | 514/567 (90.7%) |
| Black - caribbean | 33/3,833 (0.9%) | 24/3,266 (0.7%) | 9/567 (1.6%) |
| Black - african | 24/3,833 (0.6%) | 20/3,266 (0.6%) | 4/567 (0.7%) |
| Black - other black groups | 8/3,833 (0.2%) | 8/3,266 (0.2%) | 0/567 (0.0%) |
| Indian | 63/3,833 (1.6%) | 44/3,266 (1.3%) | 19/567 (3.4%) |
| Pakistani | 33/3,833 (0.9%) | 26/3,266 (0.8%) | 7/567 (1.2%) |
| Bangladeshi | 15/3,833 (0.4%) | 11/3,266 (0.3%) | 4/567 (0.7%) |
| Chinese | 9/3,833 (0.2%) | 9/3,266 (0.3%) | 0/567 (0.0%) |
| None of these | 40/3,833 (1.0%) | 30/3,266 (0.9%) | 10/567 (1.8%) |
| Haemoglobin (g/L) | 148.3 (10.1) | 148.4 (10.4) | 147.6 (8.4) |
| Anaemia (Haemoglobin<130g/L) | | | |
| No | 3,725/3,835 (97.1%) | 3,163/3,268 (96.8%) | 562/567 (99.1%) |
| Yes | 110/3,835 (2.9%) | 105/3,268 (3.2%) | 5/567 (0.9%) |
| Iron deficiency (Ferritin<15ug/L) | | | |
| No | 3,769/3,835 (98.3%) | 3,202/3,268 (98.0%) | 567/567 (100.0%) |
| Yes | 66/3,835 (1.7%) | 66/3,268 (2.0%) | 0/567 (0.0%) |
| Inflammation (C-Reactive Protein>5mg/L) | | | |
| No | 3,437/3,835 (89.6%) | 2,870/3,268 (87.8%) | 567/567 (100.0%) |
| Yes | 398/3,835 (10.4%) | 398/3,268 (12.2%) | 0/567 (0.0%) |

HSE = Health Survey for England, BMI = Body Mass Index, SD = Standard Deviation.
Data are presented as unweighted mean (SD) for continuous measures, and unweighted n/total (%) for categorical measures based on participants with non-missing haemoglobin, ferritin, and C-reactive protein.
[1] Ferritin≥15 and ≤200ug/L and C-reactive protein≤5mg/L.

#### Table 3.1.5 England HSE 2006: Participant characteristics (Males) (18-65 years)

| | Overall sample N=2,569 | Excluded sample N=2,195 | Reference sample[1] N=374 |
|---|---|---|---|
| Age (years) | 44.6 (12.6) | 45.3 (12.6) | 40.8 (12.2) |
| Sex | | | |
| Male | 2,569/2,569 (100.0%) | 2,195/2,195 (100.0%) | 374/374 (100.0%) |
| BMI (kg/m$^2$) | 27.5 (4.4) | 27.9 (4.5) | 25.5 (2.6) |
| Income | | | |
| bottom quintile (<£10,598) | 259/2,196 (11.8%) | 238/1,888 (12.6%) | 21/308 (6.8%) |
| 2nd quintile (≥£10,598 < £16,852) | 261/2,196 (11.9%) | 232/1,888 (12.3%) | 29/308 (9.4%) |
| 3rd quintile (≥£16,852< £25,114) | 432/2,196 (19.7%) | 386/1,888 (20.4%) | 46/308 (14.9%) |
| 4th quintile (≥£25,114< £40,373) | 604/2,196 (27.5%) | 501/1,888 (26.5%) | 103/308 (33.4%) |
| highest quintile (≥£40,373) | 640/2,196 (29.1%) | 531/1,888 (28.1%) | 109/308 (35.4%) |
| Ethnic group | | | |
| White | 2,378/2,568 (92.6%) | 2,059/2,194 (93.8%) | 319/374 (85.3%) |
| Mixed | 21/2,568 (0.8%) | 14/2,194 (0.6%) | 7/374 (1.9%) |
| Asian or Asian British | 109/2,568 (4.2%) | 76/2,194 (3.5%) | 33/374 (8.8%) |
| Black or Black British | 43/2,568 (1.7%) | 32/2,194 (1.5%) | 11/374 (2.9%) |
| Chinese or other ethnic group | 17/2,568 (0.7%) | 13/2,194 (0.6%) | 4/374 (1.1%) |
| Haemoglobin (g/L) | 150.5 (10.0) | 150.7 (10.2) | 149.8 (8.4) |
| Anaemia (Haemoglobin<130g/L) | | | |
| No | 2,522/2,569 (98.2%) | 2,153/2,195 (98.1%) | 369/374 (98.7%) |
| Yes | 47/2,569 (1.8%) | 42/2,195 (1.9%) | 5/374 (1.3%) |
| Iron deficiency (Ferritin<15ug/L) | | | |
| No | 2,552/2,569 (99.3%) | 2,178/2,195 (99.2%) | 374/374 (100.0%) |
| Yes | 17/2,569 (0.7%) | 17/2,195 (0.8%) | 0/374 (0.0%) |
| Inflammation (C-Reactive Protein>5mg/L) | | | |
| No | 2,269/2,569 (88.3%) | 1,895/2,195 (86.3%) | 374/374 (100.0%) |
| Yes | 300/2,569 (11.7%) | 300/2,195 (13.7%) | 0/374 (0.0%) |

HSE = Health Survey for England, BMI = Body Mass Index, SD = Standard Deviation.
Data are presented as unweighted mean (SD) for continuous measures, and unweighted n/total (%) for categorical measures based on participants with non-missing haemoglobin, ferritin, and C-reactive protein.
[1] Ferritin≥15 and ≤200ug/L and C-reactive protein≤5mg/L.

#### Table 3.1.6 England HSE 2009: Participant characteristics (Males) (18-65 years)

| | Overall sample N=804 | Excluded sample N=686 | Reference sample[1] N=118 |
|---|---|---|---|
| Age (years) | 44.5 (13.0) | 45.6 (12.8) | 38.1 (12.6) |
| Sex | | | |
| Male | 804/804 (100.0%) | 686/686 (100.0%) | 118/118 (100.0%) |
| BMI (kg/m$^2$) | 27.5 (4.4) | 27.9 (4.5) | 24.7 (2.9) |
| Income | | | |
| bottom quintile (≤£10,655.74) | 84/693 (12.1%) | 79/584 (13.5%) | 5/109 (4.6%) |
| 2nd quintile (>£10,655.74 ≤£16,900.00) | 89/693 (12.8%) | 75/584 (12.8%) | 14/109 (12.8%) |
| 3rd quintile (>£16,900.00 ≤£26,787.88) | 115/693 (16.6%) | 92/584 (15.8%) | 23/109 (21.1%) |
| 4th quintile (>£26,787.88 ≤£41,864.41) | 197/693 (28.4%) | 167/584 (28.6%) | 30/109 (27.5%) |
| highest quintile (>£41,864.41) | 208/693 (30.0%) | 171/584 (29.3%) | 37/109 (33.9%) |
| Ethnic group | | | |
| White - British | 699/804 (86.9%) | 599/686 (87.3%) | 100/118 (84.7%) |
| White - Irish | 5/804 (0.6%) | 5/686 (0.7%) | 0/118 (0.0%) |
| Any other white background | 22/804 (2.7%) | 20/686 (2.9%) | 2/118 (1.7%) |
| Mixed - White and Black Caribbean | 4/804 (0.5%) | 3/686 (0.4%) | 1/118 (0.8%) |
| Mixed - White and Black African | 2/804 (0.2%) | 1/686 (0.1%) | 1/118 (0.8%) |
| Mixed - White and Asian | 1/804 (0.1%) | 1/686 (0.1%) | 0/118 (0.0%) |
| Any other mixed background | 8/804 (1.0%) | 6/686 (0.9%) | 2/118 (1.7%) |
| Asian or Asian British - Indian | 11/804 (1.4%) | 11/686 (1.6%) | 0/118 (0.0%) |
| Asian or Asian British - Pakistani | 11/804 (1.4%) | 8/686 (1.2%) | 3/118 (2.5%) |
| Asian or Asian British - Bangladeshi | 4/804 (0.5%) | 3/686 (0.4%) | 1/118 (0.8%) |
| Any other Asian/Asian British background | 9/804 (1.1%) | 7/686 (1.0%) | 2/118 (1.7%) |
| Black or Black British - Caribbean | 7/804 (0.9%) | 6/686 (0.9%) | 1/118 (0.8%) |
| Black or Black British - African | 16/804 (2.0%) | 12/686 (1.7%) | 4/118 (3.4%) |
| Any other Black/Black British background | 1/804 (0.1%) | 0/686 (0.0%) | 1/118 (0.8%) |
| Chinese | 1/804 (0.1%) | 1/686 (0.1%) | 0/118 (0.0%) |
| Any other | 3/804 (0.4%) | 3/686 (0.4%) | 0/118 (0.0%) |
| Haemoglobin (g/L) | 149.9 (9.6) | 150.2 (9.7) | 148.3 (9.2) |
| Anaemia (Haemoglobin<130g/L) | | | |
| No | 791/804 (98.4%) | 674/686 (98.3%) | 117/118 (99.2%) |
| Yes | 13/804 (1.6%) | 12/686 (1.7%) | 1/118 (0.8%) |
| Iron deficiency (Ferritin<15ug/L) | | | |
| No | 801/804 (99.6%) | 683/686 (99.6%) | 118/118 (100.0%) |
| Yes | 3/804 (0.4%) | 3/686 (0.4%) | 0/118 (0.0%) |
| Inflammation (C-Reactive Protein>5mg/L) | | | |
| No | 728/804 (90.5%) | 610/686 (88.9%) | 118/118 (100.0%) |
| Yes | 76/804 (9.5%) | 76/686 (11.1%) | 0/118 (0.0%) |

HSE = Health Survey for England, BMI = Body Mass Index, SD = Standard Deviation.
Data are presented as unweighted mean (SD) for continuous measures, and unweighted n/total (%) for categorical measures based on participants with non-missing haemoglobin, ferritin, and C-reactive protein.
[1] Ferritin≥15 and ≤200ug/L and C-reactive protein≤5mg/L.

#### Table 3.1.7 CHN CHNS 2009: Participant characteristics (Males) (18-65 years)

| | Overall sample N=3,352 | Excluded sample N=3,107 | Reference sample[1] N=245 |
|---|---|---|---|
| Age (years) | 45.7 (12.1) | 46.3 (11.8) | 37.2 (12.9) |
| Sex | | | |
| Male | 3,352/3,352 (100.0%) | 3,107/3,107 (100.0%) | 245/245 (100.0%) |
| BMI (kg/m$^2$) | 23.5 (3.4) | 23.5 (3.4) | 22.9 (2.7) |
| Nationality | | | |
| Han | 2,978/3,340 (89.2%) | 2,748/3,095 (88.8%) | 230/245 (93.9%) |
| Mongolian | 4/3,340 (0.1%) | 4/3,095 (0.1%) | 0/245 (0.0%) |
| Hui | 10/3,340 (0.3%) | 9/3,095 (0.3%) | 1/245 (0.4%) |
| Miao | 91/3,340 (2.7%) | 90/3,095 (2.9%) | 1/245 (0.4%) |
| Zhuang | 22/3,340 (0.7%) | 20/3,095 (0.6%) | 2/245 (0.8%) |
| Buyi | 69/3,340 (2.1%) | 69/3,095 (2.2%) | 0/245 (0.0%) |
| Korean | 3/3,340 (0.1%) | 3/3,095 (0.1%) | 0/245 (0.0%) |
| Man | 88/3,340 (2.6%) | 79/3,095 (2.6%) | 9/245 (3.7%) |
| Dong | 1/3,340 (0.0%) | 1/3,095 (0.0%) | 0/245 (0.0%) |
| Tujia | 44/3,340 (1.3%) | 42/3,095 (1.4%) | 2/245 (0.8%) |
| Other | 30/3,340 (0.9%) | 30/3,095 (1.0%) | 0/245 (0.0%) |
| Haemoglobin (g/L) | 153.2 (17.3) | 153.1 (17.5) | 153.4 (15.6) |
| Anaemia (Haemoglobin<130g/L) | | | |
| No | 3,252/3,352 (97.0%) | 3,008/3,107 (96.8%) | 244/245 (99.6%) |
| Yes | 100/3,352 (3.0%) | 99/3,107 (3.2%) | 1/245 (0.4%) |
| Iron deficiency (Ferritin<15ug/L) | | | |
| No | 3,294/3,352 (98.3%) | 3,049/3,107 (98.1%) | 245/245 (100.0%) |
| Yes | 58/3,352 (1.7%) | 58/3,107 (1.9%) | 0/245 (0.0%) |
| Inflammation (C-Reactive Protein>5mg/L) | | | |
| No | 3,058/3,352 (91.2%) | 2,813/3,107 (90.5%) | 245/245 (100.0%) |
| Yes | 294/3,352 (8.8%) | 294/3,107 (9.5%) | 0/245 (0.0%) |

CHN = China, CHNS = China Health and Nutrition Survey, BMI = Body Mass Index, SD = Standard Deviation.
Data are presented as unweighted mean (SD) for continuous measures, and unweighted n/total (%) for categorical measures based on participants with non-missing haemoglobin, ferritin, and C-reactive protein.
[1] Ferritin≥15 and ≤200ug/L and C-reactive protein≤5mg/L.

Table 3.1.8 AUS NHS 2011-2012: Participant characteristics (Males) (18-65 years)

| | **Overall sample N=1,912** | **Excluded sample N=1,699** | **Reference sample[1] N=213** |
|---|---|---|---|
| Age (years) | 46.2 (12.6) | 47.5 (12.2) | 36.6 (11.6) |
| Sex | | | |
| Male | 1,912/1,912 (100.0%) | 1,699/1,699 (100.0%) | 213/213 (100.0%) |
| BMI (kg/m$^2$)* | 28.2 (5.0) | 28.6 (5.0) | 24.7 (2.6) |
| Equivalised income of household | | | |
| bottom quintile | 196/1,735 (11.3%) | 181/1,546 (11.7%) | 15/189 (7.9%) |
| 2nd quintile | 209/1,735 (12.0%) | 188/1,546 (12.2%) | 21/189 (11.1%) |
| 3rd quintile | 367/1,735 (21.2%) | 320/1,546 (20.7%) | 47/189 (24.9%) |
| 4th quintile | 447/1,735 (25.8%) | 400/1,546 (25.9%) | 47/189 (24.9%) |
| highest quintile | 516/1,735 (29.7%) | 457/1,546 (29.6%) | 59/189 (31.2%) |
| Ancestry (multiple response) | | | |
| Oceanian and Antarctican | 455/1,912 (23.8%) | 411/1,699 (24.2%) | 44/213 (20.7%) |
| North-West European | 1,025/1,912 (53.6%) | 930/1,699 (54.7%) | 95/213 (44.6%) |
| Southern and Eastern European | 174/1,912 (9.1%) | 159/1,699 (9.4%) | 15/213 (7.0%) |
| South-East Asian | 42/1,912 (2.2%) | 32/1,699 (1.9%) | 10/213 (4.7%) |
| North-East Asian | 60/1,912 (3.1%) | 44/1,699 (2.6%) | 16/213 (7.5%) |
| Southern and Central Asian | 78/1,912 (4.1%) | 56/1,699 (3.3%) | 22/213 (10.3%) |
| Other[†] | 78/1,912 (4.1%) | 67/1,699 (3.9%) | 11/213 (5.2%) |
| Haemoglobin (g/L) | 152.6 (10.5) | 152.9 (10.6) | 150.3 (8.9) |
| Anaemia (Haemoglobin<130g/L) | | | |
| No | 1,884/1,912 (98.5%) | Not displayed[§] | Not displayed[§] |
| Yes | 28/1,912 (1.5%) | Not displayed[§] | <10/213 (<4.7%) |
| Iron deficiency (Ferritin<15ug/L) | | | |
| No | 1,895/1,912 (99.1%) | Not displayed[§] | Not displayed[§] |
| Yes | 17/1,912 (0.9%) | Not displayed[§] | <10/213 (<4.7%) |
| Inflammation (C-Reactive Protein>5mg/L) | | | |
| No | 1,713/1,912 (89.6%) | Not displayed[§] | Not displayed[§] |
| Yes | 199/1,912 (10.4%) | Not displayed[§] | <10/213 (<4.7%) |

AUS = Australia, NHS = National Health Survey, BMI = Body Mass Index, SD = Standard Deviation.
Data are presented as unweighted mean (SD) for continuous measures, and unweighted n/total (%) for categorical measures based on participants with non-missing haemoglobin, ferritin, and C-reactive protein.
*BMI is missing for 53 participants.
[†] Other includes North African and Middle Eastern, American, Sub-Saharan African, Not stated, inadequately described.
[§]Not displayed due to Australian Bureau of Statistics requirements for output clearance.
[1] Ferritin≥15 and ≤200ug/L and C-reactive protein≤5mg/L.

#### Table 3.1.9 AUS NNPAS 2011-2012: Participant characteristics (Males) (18-65 years)

| | **Overall sample N=1,277** | **Excluded sample N=983** | **Reference sample[1] N=294** |
|---|---|---|---|
| Age (years) | 45.4 (12.8) | 47.6 (11.9) | 37.7 (12.8) |
| Sex | | | |
| Male | 1,277/1,277 (100.0%) | 983/983 (100.0%) | 294/294 (100.0%) |
| BMI (kg/m$^2$)* | 27.9 (4.7) | 28.6 (4.8) | 25.2 (2.7) |
| Equivalised income of household | | | |
| bottom quintile | 152/1,218 (12.5%) | 124/947 (13.1%) | 28/271 (10.3%) |
| 2nd quintile | 160/1,218 (13.1%) | 129/947 (13.6%) | 31/271 (11.4%) |
| 3rd quintile | 245/1,218 (20.1%) | 184/947 (19.4%) | 61/271 (22.5%) |
| 4th quintile | 326/1,218 (26.8%) | 243/947 (25.7%) | 83/271 (30.6%) |
| highest quintile | 335/1,218 (27.5%) | 267/947 (28.2%) | 68/271 (25.1%) |
| Country of birth | | | |
| Australia | 891/1,277 (69.8%) | 699/983 (71.1%) | 192/294 (65.3%) |
| United Kingdom | 94/1,277 (7.4%) | 71/983 (7.2%) | 23/294 (7.8%) |
| Other[†] | 292/1,277 (22.9%) | 213/983 (21.7%) | 79/294 (26.9%) |
| Haemoglobin (g/L) | 152.3 (10.2) | 152.5 (10.5) | 151.3 (9.3) |
| Anaemia (Haemoglobin<120g/L) | | | |
| No | 1,252/1,277 (98.0%) | Not displayed[§] | Not displayed[§] |
| Yes | 25/1,277 (2.0%) | Not displayed[§] | <10/294 (<3.4%) |
| Iron deficiency (Ferritin<15ug/L) | | | |
| No | 1,261/1,277 (98.7%) | Not displayed[§] | Not displayed[§] |
| Yes | 16/1,277 (1.3%) | Not displayed[§] | <10/294 (<3.4%) |
| Inflammation (C-Reactive Protein>5mg/L) | | | |
| No | 1,145/1,277 (89.7%) | Not displayed[§] | Not displayed[§] |
| Yes | 132/1,277 (10.3%) | Not displayed[§] | <10/294 (<3.4%) |

AUS = Australia, NNPAS = National Nutrition and Physical Activity Survey, BMI = Body Mass Index, SD = Standard Deviation.

Data are presented as unweighted mean (SD) for continuous measures, and unweighted n/total (%) for categorical measures based on participants with non-missing haemoglobin, ferritin, and C-reactive protein.

*BMI is missing for 43 participants.

[†] Other: New Zealand, Italy, Viet Nam, China (excl. SARs and Taiwan Province), Greece, Germany, Philippines, India, Oceania and Antarctica (excl. Australia and New Zealand), North-West Europe (excl. United Kingdom and Germany), Southern and Eastern Europe (excl. Italy and Greece), North Africa and the Middle East, South-East Asia (excl. Viet Nam and Philippines), North-East Asia (excl. China), Southern & Central Asia (excl. India), Americas, Sub-Saharan Africa.

[§] Not displayed due to Australian Bureau of Statistics requirements for output clearance.

[1] Ferritin≥15 and ≤200ug/L and C-reactive protein≤5mg/L.

#### 3.2 Demographics Adults (18-65 years) females

##### Table 3.2.1 US NHANES 1999-2000: Participant characteristics (Females) (18-65 years)

| | **Overall sample N=1,944** | **Excluded sample N=1,825** | **Reference sample[1] N=119** |
|---|---|---|---|
| Age (years) | 37.8 (14.6) | 38.3 (14.6) | 29.4 (10.8) |
| Sex | | | |
| Female | 1,944/1,944 (100.0%) | 1,825/1,825 (100.0%) | 119/119 (100.0%) |
| BMI (kg/m$^2$) | 28.7 (7.2) | 29.1 (7.2) | 23.7 (2.9) |
| Poverty | 416/1,674 (24.9%) | 388/1,570 (24.7%) | 28/104 (26.9%) |
| Race and Hispanic Origin | | | |
| Mexican American | 622/1,944 (32.0%) | 568/1,825 (31.1%) | 54/119 (45.4%) |
| Other Hispanic | 148/1,944 (7.6%) | 136/1,825 (7.5%) | 12/119 (10.1%) |
| Non-Hispanic White | 724/1,944 (37.2%) | 691/1,825 (37.9%) | 33/119 (27.7%) |
| Non-Hispanic Black | 382/1,944 (19.7%) | 368/1,825 (20.2%) | 14/119 (11.8%) |
| Other Race - Including Multi-Racial | 68/1,944 (3.5%) | 62/1,825 (3.4%) | 6/119 (5.0%) |
| Haemoglobin (g/L) | 133.1 (12.0) | 132.9 (12.2) | 136.9 (8.6) |
| Anaemia (Haemoglobin<120g/L) | | | |
| No | 1,783/1,944 (91.7%) | 1,669/1,825 (91.5%) | 114/119 (95.8%) |
| Yes | 161/1,944 (8.3%) | 156/1,825 (8.5%) | 5/119 (4.2%) |
| Iron deficiency (Ferritin<15ug/L) | | | |
| No | 1,561/1,944 (80.3%) | 1,442/1,825 (79.0%) | 119/119 (100.0%) |
| Yes | 383/1,944 (19.7%) | 383/1,825 (21.0%) | 0/119 (0.0%) |
| Inflammation (C-Reactive Protein>5mg/L) | | | |
| No | 1,273/1,944 (65.5%) | 1,154/1,825 (63.2%) | 119/119 (100.0%) |
| Yes | 671/1,944 (34.5%) | 671/1,825 (36.8%) | 0/119 (0.0%) |

US = United States, NHANES = National Health and Nutrition Examination Survey, BMI = Body Mass Index, SD = Standard Deviation.
Data are presented as unweighted mean (SD) for continuous measures, and unweighted n/total (%) for categorical measures based on participants with non-missing haemoglobin, ferritin, and C-reactive protein.
[1] Ferritin≥15 and ≤150ug/L and C-reactive protein≤5mg/L.

#### Table 3.2.2 US NHANES 2001-2002: Participant characteristics (Females) (18-65 years)

| | **Overall sample N=2,160** | **Excluded sample N=2,052** | **Reference sample[1] N=108** |
|---|---|---|---|
| Age (years) | 37.5 (14.1) | 37.9 (14.2) | 30.0 (10.8) |
| Sex | | | |
| Female | 2,160/2,160 (100.0%) | 2,052/2,052 (100.0%) | 108/108 (100.0%) |
| BMI (kg/m$^2$) | 28.3 (7.0) | 28.6 (7.1) | 24.2 (2.8) |
| Poverty | 430/2,026 (21.2%) | 401/1,925 (20.8%) | 29/101 (28.7%) |
| Race and Hispanic Origin | | | |
| Mexican American | 526/2,160 (24.4%) | 489/2,052 (23.8%) | 37/108 (34.3%) |
| Other Hispanic | 97/2,160 (4.5%) | 86/2,052 (4.2%) | 11/108 (10.2%) |
| Non-Hispanic White | 1,001/2,160 (46.3%) | 966/2,052 (47.1%) | 35/108 (32.4%) |
| Non-Hispanic Black | 445/2,160 (20.6%) | 428/2,052 (20.9%) | 17/108 (15.7%) |
| Other Race - Including Multi-Racial | 91/2,160 (4.2%) | 83/2,052 (4.0%) | 8/108 (7.4%) |
| Haemoglobin (g/L) | 132.7 (12.9) | 132.5 (13.0) | 136.5 (9.9) |
| Anaemia (Haemoglobin<120g/L) | | | |
| No | 1,946/2,160 (90.1%) | 1,843/2,052 (89.8%) | 103/108 (95.4%) |
| Yes | 214/2,160 (9.9%) | 209/2,052 (10.2%) | 5/108 (4.6%) |
| Iron deficiency (Ferritin<15ug/L) | | | |
| No | 1,668/2,160 (77.2%) | 1,560/2,052 (76.0%) | 108/108 (100.0%) |
| Yes | 492/2,160 (22.8%) | 492/2,052 (24.0%) | 0/108 (0.0%) |
| Inflammation (C-Reactive Protein>5mg/L) | | | |
| No | 1,475/2,160 (68.3%) | 1,367/2,052 (66.6%) | 108/108 (100.0%) |
| Yes | 685/2,160 (31.7%) | 685/2,052 (33.4%) | 0/108 (0.0%) |

US = United States, NHANES = National Health and Nutrition Examination Survey, BMI = Body Mass Index, SD = Standard Deviation.
Data are presented as unweighted mean (SD) for continuous measures, and unweighted n/total (%) for categorical measures based on participants with non-missing haemoglobin, ferritin, and C-reactive protein.
[1] Ferritin≥15 and ≤150ug/L and C-reactive protein≤5mg/L.

#### Table 3.2.3 US NHANES 2003-2004: Participant characteristics (Females) (18-65 years)

| | **Overall sample N=1,455** | **Excluded sample N=1,337** | **Reference sample[1] N=118** |
|---|---|---|---|
| Age (years) | 30.9 (9.8) | 31.2 (9.8) | 27.5 (9.2) |
| Sex | | | |
| Female | 1,455/1,455 (100.0%) | 1,337/1,337 (100.0%) | 118/118 (100.0%) |
| BMI (kg/m$^2$) | 28.3 (7.4) | 28.7 (7.5) | 23.6 (3.0) |
| Poverty | 403/1,381 (29.2%) | 371/1,272 (29.2%) | 32/109 (29.4%) |
| Race and Hispanic Origin | | | |
| Mexican American | 336/1,455 (23.1%) | 295/1,337 (22.1%) | 41/118 (34.7%) |
| Other Hispanic | 54/1,455 (3.7%) | 46/1,337 (3.4%) | 8/118 (6.8%) |
| Non-Hispanic White | 648/1,455 (44.5%) | 615/1,337 (46.0%) | 33/118 (28.0%) |
| Non-Hispanic Black | 360/1,455 (24.7%) | 334/1,337 (25.0%) | 26/118 (22.0%) |
| Other Race - Including Multi-Racial | 57/1,455 (3.9%) | 47/1,337 (3.5%) | 10/118 (8.5%) |
| Haemoglobin (g/L) | 133.2 (12.4) | 132.9 (12.6) | 136.4 (8.9) |
| Anaemia (Haemoglobin<120g/L) | | | |
| No | 1,334/1,455 (91.7%) | 1,218/1,337 (91.1%) | 116/118 (98.3%) |
| Yes | 121/1,455 (8.3%) | 119/1,337 (8.9%) | 2/118 (1.7%) |
| Iron deficiency (Ferritin<15ug/L) | | | |
| No | 1,204/1,455 (82.7%) | 1,086/1,337 (81.2%) | 118/118 (100.0%) |
| Yes | 251/1,455 (17.3%) | 251/1,337 (18.8%) | 0/118 (0.0%) |
| Inflammation (C-Reactive Protein>5mg/L) | | | |
| No | 986/1,455 (67.8%) | 868/1,337 (64.9%) | 118/118 (100.0%) |
| Yes | 469/1,455 (32.2%) | 469/1,337 (35.1%) | 0/118 (0.0%) |

US = United States, NHANES = National Health and Nutrition Examination Survey, BMI = Body Mass Index, SD = Standard Deviation.
Data are presented as unweighted mean (SD) for continuous measures, and unweighted n/total (%) for categorical measures based on participants with non-missing haemoglobin, ferritin, and C-reactive protein. No ferritin was measured in participants more than 49 years.
[1] Ferritin≥15 and ≤150ug/L and C-reactive protein≤5mg/L.

#### Table 3.2.4 US NHANES 2005-2006: Participant characteristics (Females) (18-65 years)

| | Overall sample N=1,606 | Excluded sample N=1,473 | Reference sample[1] N=133 |
|---|---|---|---|
| Age (years) | 30.7 (9.5) | 31.0 (9.5) | 28.1 (9.2) |
| Sex | | | |
| Female | 1,606/1,606 (100.0%) | 1,473/1,473 (100.0%) | 133/133 (100.0%) |
| BMI (kg/m$^2$) | 28.6 (7.4) | 29.0 (7.5) | 23.9 (3.2) |
| Poverty | 392/1,539 (25.5%) | 357/1,414 (25.2%) | 35/125 (28.0%) |
| Race and Hispanic Origin | | | |
| Mexican American | 416/1,606 (25.9%) | 366/1,473 (24.8%) | 50/133 (37.6%) |
| Other Hispanic | 64/1,606 (4.0%) | 58/1,473 (3.9%) | 6/133 (4.5%) |
| Non-Hispanic White | 638/1,606 (39.7%) | 606/1,473 (41.1%) | 32/133 (24.1%) |
| Non-Hispanic Black | 395/1,606 (24.6%) | 364/1,473 (24.7%) | 31/133 (23.3%) |
| Other Race - Including Multi-Racial | 93/1,606 (5.8%) | 79/1,473 (5.4%) | 14/133 (10.5%) |
| Haemoglobin (g/L) | 131.3 (12.5) | 130.9 (12.8) | 135.5 (8.5) |
| Anaemia (Haemoglobin<120g/L) | | | |
| No | 1,451/1,606 (90.3%) | 1,320/1,473 (89.6%) | 131/133 (98.5%) |
| Yes | 155/1,606 (9.7%) | 153/1,473 (10.4%) | 2/133 (1.5%) |
| Iron deficiency (Ferritin<15ug/L) | | | |
| No | 1,291/1,606 (80.4%) | 1,158/1,473 (78.6%) | 133/133 (100.0%) |
| Yes | 315/1,606 (19.6%) | 315/1,473 (21.4%) | 0/133 (0.0%) |
| Inflammation (C-Reactive Protein>5mg/L) | | | |
| No | 1,074/1,606 (66.9%) | 941/1,473 (63.9%) | 133/133 (100.0%) |
| Yes | 532/1,606 (33.1%) | 532/1,473 (36.1%) | 0/133 (0.0%) |

US = United States, NHANES = National Health and Nutrition Examination Survey, BMI = Body Mass Index, SD = Standard Deviation.

Data are presented as unweighted mean (SD) for continuous measures, and unweighted n/total (%) for categorical measures based on participants with non-missing haemoglobin, ferritin, and C-reactive protein. No ferritin was measured in participants more than 49 years.

[1] Ferritin≥15 and ≤150ug/L and C-reactive protein≤5mg/L.

#### Table 3.2.5 US NHANES 2007-2008: Participant characteristics (Females) (18-65 years)

| | **Overall sample N=1,434** | **Excluded sample N=1,297** | **Reference sample[1] N=137** |
|---|---|---|---|
| Age (years) | 33.8 (9.3) | 34.1 (9.3) | 30.8 (9.2) |
| Sex | | | |
| Female | 1,434/1,434 (100.0%) | 1,297/1,297 (100.0%) | 137/137 (100.0%) |
| BMI (kg/m$^2$) | 28.6 (7.6) | 29.0 (7.7) | 24.1 (2.9) |
| Poverty | 364/1,328 (27.4%) | 338/1,207 (28.0%) | 26/121 (21.5%) |
| Race and Hispanic Origin | | | |
| Mexican American | 304/1,434 (21.2%) | 259/1,297 (20.0%) | 45/137 (32.8%) |
| Other Hispanic | 197/1,434 (13.7%) | 167/1,297 (12.9%) | 30/137 (21.9%) |
| Non-Hispanic White | 582/1,434 (40.6%) | 553/1,297 (42.6%) | 29/137 (21.2%) |
| Non-Hispanic Black | 289/1,434 (20.2%) | 267/1,297 (20.6%) | 22/137 (16.1%) |
| Other Race - Including Multi-Racial | 62/1,434 (4.3%) | 51/1,297 (3.9%) | 11/137 (8.0%) |
| Haemoglobin (g/L) | 132.3 (12.6) | 132.2 (13.0) | 133.2 (8.5) |
| Anaemia (Haemoglobin<120g/L) | | | |
| No | 1,273/1,434 (88.8%) | 1,140/1,297 (87.9%) | 133/137 (97.1%) |
| Yes | 161/1,434 (11.2%) | 157/1,297 (12.1%) | 4/137 (2.9%) |
| Iron deficiency (Ferritin<15ug/L) | | | |
| No | 1,177/1,434 (82.1%) | 1,040/1,297 (80.2%) | 137/137 (100.0%) |
| Yes | 257/1,434 (17.9%) | 257/1,297 (19.8%) | 0/137 (0.0%) |
| Inflammation (C-Reactive Protein>5mg/L) | | | |
| No | 1,068/1,434 (74.5%) | 931/1,297 (71.8%) | 137/137 (100.0%) |
| Yes | 366/1,434 (25.5%) | 366/1,297 (28.2%) | 0/137 (0.0%) |

US = United States, NHANES = National Health and Nutrition Examination Survey, BMI = Body Mass Index, SD = Standard Deviation.
Data are presented as unweighted mean (SD) for continuous measures, and unweighted n/total (%) for categorical measures based on participants with non-missing haemoglobin, ferritin, and C-reactive protein. No ferritin was measured in participants more than 49 years.
[1] Ferritin≥15 and ≤150ug/L and C-reactive protein≤5mg/L.

#### Table 3.2.6 US NHANES 2009-2010: Participant characteristics (Females) (18-65 years)

| | **Overall sample N=1,703** | **Excluded sample N=1,536** | **Reference sample[1] N=167** |
|---|---|---|---|
| Age (years) | 33.5 (9.5) | 33.8 (9.4) | 30.7 (9.2) |
| Sex | | | |
| Female | 1,703/1,703 (100.0%) | 1,536/1,536 (100.0%) | 167/167 (100.0%) |
| BMI (kg/m$^2$) | 28.8 (8.0) | 29.4 (8.2) | 24.0 (3.0) |
| Poverty | 475/1,557 (30.5%) | 438/1,416 (30.9%) | 37/141 (26.2%) |
| Race and Hispanic Origin | | | |
| Mexican American | 346/1,703 (20.3%) | 299/1,536 (19.5%) | 47/167 (28.1%) |
| Other Hispanic | 201/1,703 (11.8%) | 175/1,536 (11.4%) | 26/167 (15.6%) |
| Non-Hispanic White | 738/1,703 (43.3%) | 685/1,536 (44.6%) | 53/167 (31.7%) |
| Non-Hispanic Black | 303/1,703 (17.8%) | 285/1,536 (18.6%) | 18/167 (10.8%) |
| Other Race - Including Multi-Racial | 115/1,703 (6.8%) | 92/1,536 (6.0%) | 23/167 (13.8%) |
| Haemoglobin (g/L) | 131.9 (12.0) | 131.7 (12.3) | 133.8 (8.6) |
| Anaemia (Haemoglobin<120g/L) | | | |
| No | 1,504/1,703 (88.3%) | 1,346/1,536 (87.6%) | 158/167 (94.6%) |
| Yes | 199/1,703 (11.7%) | 190/1,536 (12.4%) | 9/167 (5.4%) |
| Iron deficiency (Ferritin<15ug/L) | | | |
| No | 1,411/1,703 (82.9%) | 1,244/1,536 (81.0%) | 167/167 (100.0%) |
| Yes | 292/1,703 (17.1%) | 292/1,536 (19.0%) | 0/167 (0.0%) |
| Inflammation (C-Reactive Protein>5mg/L) | | | |
| No | 1,267/1,703 (74.4%) | 1,100/1,536 (71.6%) | 167/167 (100.0%) |
| Yes | 436/1,703 (25.6%) | 436/1,536 (28.4%) | 0/167 (0.0%) |

US = United States, NHANES = National Health and Nutrition Examination Survey, BMI = Body Mass Index, SD = Standard Deviation.
Data are presented as unweighted mean (SD) for continuous measures, and unweighted n/total (%) for categorical measures based on participants with non-missing haemoglobin, ferritin, and C-reactive protein. No ferritin was measured in participants more than 49 years.
[1] Ferritin≥15 and ≤150ug/L and C-reactive protein≤5mg/L.

#### Table 3.2.7 US NHANES 2015-2016: Participant characteristics (Females) (18-65 years)

| | **Overall sample N=1,485** | **Excluded sample N=1,317** | **Reference sample[1] N=168** |
|---|---|---|---|
| Age (years) | 33.5 (9.2) | 33.7 (9.3) | 31.7 (8.6) |
| Sex | | | |
| Female | 1,485/1,485 (100.0%) | 1,317/1,317 (100.0%) | 168/168 (100.0%) |
| BMI (kg/m$^2$) | 29.7 (8.1) | 30.4 (8.3) | 24.1 (3.2) |
| Poverty | 331/1,360 (24.3%) | 297/1,212 (24.5%) | 34/148 (23.0%) |
| Race and Hispanic Origin | | | |
| Mexican American | 289/1,485 (19.5%) | 246/1,317 (18.7%) | 43/168 (25.6%) |
| Other Hispanic | 194/1,485 (13.1%) | 169/1,317 (12.8%) | 25/168 (14.9%) |
| Non-Hispanic White | 407/1,485 (27.4%) | 378/1,317 (28.7%) | 29/168 (17.3%) |
| Non-Hispanic Black | 339/1,485 (22.8%) | 323/1,317 (24.5%) | 16/168 (9.5%) |
| Non-Hispanic Asian | 194/1,485 (13.1%) | 142/1,317 (10.8%) | 52/168 (31.0%) |
| Other Race - Including Multi-Racial | 62/1,485 (4.2%) | 59/1,317 (4.5%) | 3/168 (1.8%) |
| Haemoglobin (g/L) | 128.8 (13.0) | 128.3 (13.3) | 132.8 (8.4) |
| Anaemia (Haemoglobin<120g/L) | | | |
| No | 1,227/1,485 (82.6%) | 1,067/1,317 (81.0%) | 160/168 (95.2%) |
| Yes | 258/1,485 (17.4%) | 250/1,317 (19.0%) | 8/168 (4.8%) |
| Iron deficiency (Ferritin<15ug/L) | | | |
| No | 1,198/1,485 (80.7%) | 1,030/1,317 (78.2%) | 168/168 (100.0%) |
| Yes | 287/1,485 (19.3%) | 287/1,317 (21.8%) | 0/168 (0.0%) |
| Inflammation (C-Reactive Protein>5mg/L) | | | |
| No | 1,083/1,485 (72.9%) | 915/1,317 (69.5%) | 168/168 (100.0%) |
| Yes | 402/1,485 (27.1%) | 402/1,317 (30.5%) | 0/168 (0.0%) |

US = United States, NHANES = National Health and Nutrition Examination Survey, BMI = Body Mass Index, SD = Standard Deviation.
Data are presented as unweighted mean (SD) for continuous measures, and unweighted n/total (%) for categorical measures based on participants with non-missing haemoglobin, ferritin, and C-reactive protein. No ferritin was measured in participants more than 49 years.
[1] Ferritin≥15 and ≤150ug/L and C-reactive protein≤5mg/L.

#### Table 3.2.8 US NHANES 2017-2018: Participant characteristics (Females) (18-65 years)

| | Overall sample N=2,059 | Excluded sample N=1,916 | Reference sample[1] N=143 |
|---|---|---|---|
| Age (years) | 42.4 (14.3) | 43.0 (14.3) | 34.6 (12.0) |
| Sex | | | |
| Female | 2,059/2,059 (100.0%) | 1,916/1,916 (100.0%) | 143/143 (100.0%) |
| BMI (kg/m$^2$) | 30.3 (8.4) | 30.8 (8.5) | 24.0 (3.0) |
| Poverty | 412/1,790 (23.0%) | 383/1,672 (22.9%) | 29/118 (24.6%) |
| Race and Hispanic Origin | | | |
| Mexican American | 318/2,059 (15.4%) | 295/1,916 (15.4%) | 23/143 (16.1%) |
| Other Hispanic | 223/2,059 (10.8%) | 201/1,916 (10.5%) | 22/143 (15.4%) |
| Non-Hispanic White | 616/2,059 (29.9%) | 591/1,916 (30.8%) | 25/143 (17.5%) |
| Non-Hispanic Black | 486/2,059 (23.6%) | 469/1,916 (24.5%) | 17/143 (11.9%) |
| Non-Hispanic Asian | 307/2,059 (14.9%) | 254/1,916 (13.3%) | 53/143 (37.1%) |
| Other Race - Including Multi-Racial | 109/2,059 (5.3%) | 106/1,916 (5.5%) | 3/143 (2.1%) |
| Haemoglobin (g/L) | 132.0 (13.0) | 131.8 (13.3) | 134.6 (7.9) |
| Anaemia (Haemoglobin<120g/L) | | | |
| No | 1,803/2,059 (87.6%) | 1,665/1,916 (86.9%) | 138/143 (96.5%) |
| Yes | 256/2,059 (12.4%) | 251/1,916 (13.1%) | 5/143 (3.5%) |
| Iron deficiency (Ferritin<15ug/L) | | | |
| No | 1,812/2,059 (88.0%) | 1,669/1,916 (87.1%) | 143/143 (100.0%) |
| Yes | 247/2,059 (12.0%) | 247/1,916 (12.9%) | 0/143 (0.0%) |
| Inflammation (C-Reactive Protein>5mg/L) | | | |
| No | 1,471/2,059 (71.4%) | 1,328/1,916 (69.3%) | 143/143 (100.0%) |
| Yes | 588/2,059 (28.6%) | 588/1,916 (30.7%) | 0/143 (0.0%) |

US = United States, NHANES = National Health and Nutrition Examination Survey, BMI = Body Mass Index, SD = Standard Deviation.
Data are presented as unweighted mean (SD) for continuous measures, and unweighted n/total (%) for categorical measures based on participants with non-missing haemoglobin, ferritin, and C-reactive protein.
[1] Ferritin≥15 and ≤150ug/L and C-reactive protein≤5mg/L.

#### Table 3.2.9 England HSE 1998: Participant characteristics (Females) (18-65 years)

| | **Overall sample N=4,189** | **Excluded sample N=3,613** | **Reference sample[1] N=576** |
|---|---|---|---|
| Age (years) | 42.5 (12.5) | 42.8 (12.6) | 40.6 (11.8) |
| Sex | | | |
| Female | 4,189/4,189 (100.0%) | 3,613/3,613 (100.0%) | 576/576 (100.0%) |
| BMI (kg/m$^2$) | 26.3 (5.2) | 26.6 (5.4) | 24.2 (2.7) |
| Income | | | |
| bottom quintile (≤£7,186) | 604/3,699 (16.3%) | 553/3,194 (17.3%) | 51/505 (10.1%) |
| 2nd quintile (>£7,186 ≤ £10,834) | 492/3,699 (13.3%) | 449/3,194 (14.1%) | 43/505 (8.5%) |
| 3rd quintile (>£10,834 ≤ £17,890) | 905/3,699 (24.5%) | 778/3,194 (24.4%) | 127/505 (25.1%) |
| 4th quintile (>£17,890 ≤ £27,705) | 865/3,699 (23.4%) | 733/3,194 (22.9%) | 132/505 (26.1%) |
| highest quintile (>£27,705) | 833/3,699 (22.5%) | 681/3,194 (21.3%) | 152/505 (30.1%) |
| Ethnic group | | | |
| White | 3,957/4,184 (94.6%) | 3,421/3,608 (94.8%) | 536/576 (93.1%) |
| Black - Caribbean | 24/4,184 (0.6%) | 20/3,608 (0.6%) | 4/576 (0.7%) |
| Black - African | 35/4,184 (0.8%) | 29/3,608 (0.8%) | 6/576 (1.0%) |
| Black - other black groups | 10/4,184 (0.2%) | 10/3,608 (0.3%) | 0/576 (0.0%) |
| Indian | 67/4,184 (1.6%) | 57/3,608 (1.6%) | 10/576 (1.7%) |
| Pakistani | 28/4,184 (0.7%) | 24/3,608 (0.7%) | 4/576 (0.7%) |
| Bangladeshi | 11/4,184 (0.3%) | 9/3,608 (0.2%) | 2/576 (0.3%) |
| Chinese | 8/4,184 (0.2%) | 7/3,608 (0.2%) | 1/576 (0.2%) |
| None of these | 44/4,184 (1.1%) | 31/3,608 (0.9%) | 13/576 (2.3%) |
| Haemoglobin (g/L) | 131.7 (10.4) | 131.8 (10.6) | 131.1 (8.4) |
| Anaemia (Haemoglobin<120g/L) | | | |
| No | 3,782/4,189 (90.3%) | 3,248/3,613 (89.9%) | 534/576 (92.7%) |
| Yes | 407/4,189 (9.7%) | 365/3,613 (10.1%) | 42/576 (7.3%) |
| Iron deficiency (Ferritin<15ug/L) | | | |
| No | 3,673/4,189 (87.7%) | 3,097/3,613 (85.7%) | 576/576 (100.0%) |
| Yes | 516/4,189 (12.3%) | 516/3,613 (14.3%) | 0/576 (0.0%) |
| Inflammation (C-Reactive Protein>5mg/L) | | | |
| No | 3,472/4,189 (82.9%) | 2,896/3,613 (80.2%) | 576/576 (100.0%) |
| Yes | 717/4,189 (17.1%) | 717/3,613 (19.8%) | 0/576 (0.0%) |

HSE = Health Survey for England, BMI = Body Mass Index, SD = Standard Deviation.
Data are presented as unweighted mean (SD) for continuous measures, and unweighted n/total (%) for categorical measures based on participants with non-missing haemoglobin, ferritin, and C-reactive protein.
[1] Ferritin≥15 and ≤150ug/L and C-reactive protein≤5mg/L.

#### Table 3.2.10 England HSE 2006: Participant characteristics (Females) (18-65 years)

| | Overall sample N=3,026 | Excluded sample N=2,511 | Reference sample[1] N=515 |
|---|---|---|---|
| Age (years) | 44.5 (12.4) | 45.0 (12.4) | 42.1 (11.8) |
| Sex | | | |
| Female | 3,026/3,026 (100.0%) | 2,511/2,511 (100.0%) | 515/515 (100.0%) |
| BMI (kg/m$^2$) | 26.7 (5.4) | 27.2 (5.7) | 24.2 (2.8) |
| Income | | | |
| bottom quintile (<£10,598) | 380/2,591 (14.7%) | 342/2,149 (15.9%) | 38/442 (8.6%) |
| 2nd quintile (≥£10,598< £16,852) | 404/2,591 (15.6%) | 351/2,149 (16.3%) | 53/442 (12.0%) |
| 3rd quintile (≥£16,852< £25,114) | 537/2,591 (20.7%) | 456/2,149 (21.2%) | 81/442 (18.3%) |
| 4th quintile (≥£25,114< £40,373) | 638/2,591 (24.6%) | 524/2,149 (24.4%) | 114/442 (25.8%) |
| highest quintile (≥£40,373) | 632/2,591 (24.4%) | 476/2,149 (22.1%) | 156/442 (35.3%) |
| Ethnic group | | | |
| White | 2,792/3,025 (92.3%) | 2,336/2,510 (93.1%) | 456/515 (88.5%) |
| Mixed | 31/3,025 (1.0%) | 27/2,510 (1.1%) | 4/515 (0.8%) |
| Asian or Asian British | 123/3,025 (4.1%) | 92/2,510 (3.7%) | 31/515 (6.0%) |
| Black or Black British | 52/3,025 (1.7%) | 40/2,510 (1.6%) | 12/515 (2.3%) |
| Chinese or other ethnic group | 27/3,025 (0.9%) | 15/2,510 (0.6%) | 12/515 (2.3%) |
| Haemoglobin (g/L) | 134.0 (10.2) | 134.2 (10.6) | 133.1 (8.3) |
| Anaemia (Haemoglobin<120g/L) | | | |
| No | 2,819/3,026 (93.2%) | 2,328/2,511 (92.7%) | 491/515 (95.3%) |
| Yes | 207/3,026 (6.8%) | 183/2,511 (7.3%) | 24/515 (4.7%) |
| Iron deficiency (Ferritin<15ug/L) | | | |
| No | 2,762/3,026 (91.3%) | 2,247/2,511 (89.5%) | 515/515 (100.0%) |
| Yes | 264/3,026 (8.7%) | 264/2,511 (10.5%) | 0/515 (0.0%) |
| Inflammation (C-Reactive Protein>5mg/L) | | | |
| No | 2,513/3,026 (83.0%) | 1,998/2,511 (79.6%) | 515/515 (100.0%) |
| Yes | 513/3,026 (17.0%) | 513/2,511 (20.4%) | 0/515 (0.0%) |

HSE = Health Survey for England, BMI = Body Mass Index, SD = Standard Deviation.
Data are presented as unweighted mean (SD) for continuous measures, and unweighted n/total (%) for categorical measures based on participants with non-missing haemoglobin, ferritin, and C-reactive protein.
[1] Ferritin≥15 and ≤150ug/L and C-reactive protein≤5mg/L.

#### Table 3.2.11 England HSE 2009: Participant characteristics (Females) (18-65 years)

| | Overall sample N=892 | Excluded sample N=750 | Reference sample[1] N=142 |
|---|---|---|---|
| Age (years) | 44.9 (12.5) | 45.5 (12.5) | 41.9 (11.8) |
| Sex | | | |
| Female | 892/892 (100.0%) | 750/750 (100.0%) | 142/142 (100.0%) |
| BMI (kg/m$^2$) | 27.3 (5.7) | 27.8 (6.0) | 24.4 (2.9) |
| Income | | | |
| bottom quintile (≤£10,655.74) | 110/768 (14.3%) | 101/646 (15.6%) | 9/122 (7.4%) |
| 2nd quintile (>£10,655.74 ≤£16,900.00) | 135/768 (17.6%) | 119/646 (18.4%) | 16/122 (13.1%) |
| 3rd quintile (>£16,900.00 ≤£26,787.88) | 140/768 (18.2%) | 123/646 (19.0%) | 17/122 (13.9%) |
| 4th quintile (>£26,787.88 ≤£41,864.41) | 192/768 (25.0%) | 155/646 (24.0%) | 37/122 (30.3%) |
| highest quintile (>£41,864.41) | 191/768 (24.9%) | 148/646 (22.9%) | 43/122 (35.2%) |
| Ethnic group | | | |
| White - British | 781/891 (87.7%) | 672/750 (89.6%) | 109/141 (77.3%) |
| White - Irish | 8/891 (0.9%) | 5/750 (0.7%) | 3/141 (2.1%) |
| Any other white background | 36/891 (4.0%) | 27/750 (3.6%) | 9/141 (6.4%) |
| Mixed - White and Black Caribbean | 1/891 (0.1%) | 1/750 (0.1%) | 0/141 (0.0%) |
| Mixed - White and Black African | 2/891 (0.2%) | 1/750 (0.1%) | 1/141 (0.7%) |
| Mixed - White and Asian | 2/891 (0.2%) | 1/750 (0.1%) | 1/141 (0.7%) |
| Any other mixed background | 4/891 (0.4%) | 4/750 (0.5%) | 0/141 (0.0%) |
| Asian or Asian British - Indian | 9/891 (1.0%) | 8/750 (1.1%) | 1/141 (0.7%) |
| Asian or Asian British - Pakistani | 11/891 (1.2%) | 10/750 (1.3%) | 1/141 (0.7%) |
| Asian or Asian British - Bangladeshi | 5/891 (0.6%) | 4/750 (0.5%) | 1/141 (0.7%) |
| Any other Asian/Asian British background | 7/891 (0.8%) | 2/750 (0.3%) | 5/141 (3.5%) |
| Black or Black British - Caribbean | 5/891 (0.6%) | 3/750 (0.4%) | 2/141 (1.4%) |
| Black or Black British - African | 14/891 (1.6%) | 8/750 (1.1%) | 6/141 (4.3%) |
| Any other Black/Black British background | 1/891 (0.1%) | 1/750 (0.1%) | 0/141 (0.0%) |
| Chinese | 1/891 (0.1%) | 1/750 (0.1%) | 0/141 (0.0%) |
| Any other | 4/891 (0.4%) | 2/750 (0.3%) | 2/141 (1.4%) |
| Haemoglobin (g/L) | 132.9 (10.3) | 132.8 (10.4) | 133.7 (9.6) |
| Anaemia (Haemoglobin<120g/L) | | | |
| No | 823/892 (92.3%) | 683/750 (91.1%) | 140/142 (98.6%) |
| Yes | 69/892 (7.7%) | 67/750 (8.9%) | 2/142 (1.4%) |
| Iron deficiency (Ferritin<15ug/L) | | | |
| No | 809/892 (90.7%) | 667/750 (88.9%) | 142/142 (100.0%) |
| Yes | 83/892 (9.3%) | 83/750 (11.1%) | 0/142 (0.0%) |
| Inflammation (C-Reactive Protein>5mg/L) | | | |
| No | 734/892 (82.3%) | 592/750 (78.9%) | 142/142 (100.0%) |
| Yes | 158/892 (17.7%) | 158/750 (21.1%) | 0/142 (0.0%) |

HSE = Health Survey for England, BMI = Body Mass Index, SD = Standard Deviation.
Data are presented as unweighted mean (SD) for continuous measures, and unweighted n/total (%) for categorical measures based on participants with non-missing haemoglobin, ferritin, and C-reactive protein.
[1] Ferritin≥15 and ≤150ug/L and C-reactive protein≤5mg/L.

#### Table 3.2.12 CHN CHNS 2009: Participant characteristics (Females) (18-65 years)

| | **Overall sample N=3,820** | **Excluded sample N=3,254** | **Reference sample[1] N=566** |
|---|---|---|---|
| Age (years) | 45.6 (11.8) | 47.1 (11.4) | 37.2 (10.7) |
| Sex | | | |
| Female | 3,820/3,820 (100.0%) | 3,254/3,254 (100.0%) | 566/566 (100.0%) |
| BMI (kg/m$^2$) | 23.4 (3.5) | 23.5 (3.6) | 22.9 (2.6) |
| Nationality | | | |
| Han | 3,374/3,808 (88.6%) | 2,856/3,246 (88.0%) | 518/562 (92.2%) |
| Mongolian | 4/3,808 (0.1%) | 4/3,246 (0.1%) | 0/562 (0.0%) |
| Hui | 7/3,808 (0.2%) | 4/3,246 (0.1%) | 3/562 (0.5%) |
| Miao | 121/3,808 (3.2%) | 112/3,246 (3.5%) | 9/562 (1.6%) |
| Zhuang | 28/3,808 (0.7%) | 26/3,246 (0.8%) | 2/562 (0.4%) |
| Buyi | 68/3,808 (1.8%) | 68/3,246 (2.1%) | 0/562 (0.0%) |
| Korean | 3/3,808 (0.1%) | 2/3,246 (0.1%) | 1/562 (0.2%) |
| Man | 117/3,808 (3.1%) | 97/3,246 (3.0%) | 20/562 (3.6%) |
| Tujia | 43/3,808 (1.1%) | 41/3,246 (1.3%) | 2/562 (0.4%) |
| Other | 43/3,808 (1.1%) | 36/3,246 (1.1%) | 7/562 (1.2%) |
| Haemoglobin (g/L) | 132.4 (17.9) | 132.1 (18.2) | 134.3 (16.0) |
| Anaemia (Haemoglobin<120g/L) | | | |
| No | 2,213/3,820 (57.9%) | 1,859/3,254 (57.1%) | 354/566 (62.5%) |
| Yes | 1,607/3,820 (42.1%) | 1,395/3,254 (42.9%) | 212/566 (37.5%) |
| Iron deficiency (Ferritin<15ug/L) | | | |
| No | 3,145/3,820 (82.3%) | 2,579/3,254 (79.3%) | 566/566 (100.0%) |
| Yes | 675/3,820 (17.7%) | 675/3,254 (20.7%) | 0/566 (0.0%) |
| Inflammation (C-Reactive Protein>5mg/L) | | | |
| No | 3,538/3,820 (92.6%) | 2,972/3,254 (91.3%) | 566/566 (100.0%) |
| Yes | 282/3,820 (7.4%) | 282/3,254 (8.7%) | 0/566 (0.0%) |

CHN = China, CHNS = China Health and Nutrition Survey, BMI = Body Mass Index, SD = Standard Deviation.
Data are presented as unweighted mean (SD) for continuous measures, and unweighted n/total (%) for categorical measures based on participants with non-missing haemoglobin, ferritin, and C-reactive protein.
[1] Ferritin≥15 and ≤150ug/L and C-reactive protein≤5mg/L.

#### Table 3.2.13 AUS NHS 2011-2012: Participant characteristics (Females) (18-65 years)

| | Overall sample N=2,480 | Excluded sample N=2,198 | Reference sample N=282 |
|---|---|---|---|
| Age (years) | 45.3 (12.6) | 46.2 (12.5) | 38.3 (11.0) |
| Sex | | | |
| Female | 2,480/2,480 (100.0%) | 2,198/2,198 (100.0%) | 282/282 (100.0%) |
| BMI (kg/m$^2$)* | 27.5 (6.3) | 28.0 (6.5) | 23.7 (2.7) |
| Equivalised income of household | | | |
| bottom quintile | 270/2,121 (12.7%) | 251/1,883 (13.3%) | 19/238 (8.0%) |
| 2nd quintile | 352/2,121 (16.6%) | 317/1,883 (16.8%) | 35/238 (14.7%) |
| 3rd quintile | 487/2,121 (23.0%) | 430/1,883 (22.8%) | 57/238 (23.9%) |
| 4th quintile | 521/2,121 (24.6%) | 450/1,883 (23.9%) | 71/238 (29.8%) |
| highest quintile | 491/2,121 (23.1%) | 435/1,883 (23.1%) | 56/238 (23.5%) |
| Ancestry (multiple response) | | | |
| Oceanian and Antarctican | 621/2,480 (25.0%) | 565/2,198 (25.7%) | 56/282 (19.9%) |
| North-West European | 1,326/2,480 (53.5%) | 1,206/2,198 (54.9%) | 120/282 (42.6%) |
| Southern and Eastern European | 217/2,480 (8.8%) | 185/2,198 (8.4%) | 32/282 (11.3%) |
| South-East Asian | 73/2,480 (2.9%) | 57/2,198 (2.6%) | 16/282 (5.7%) |
| North-East Asian | 63/2,480 (2.5%) | 43/2,198 (2.0%) | 20/282 (7.1%) |
| Southern and Central Asian | 74/2,480 (3.0%) | 58/2,198 (2.6%) | 16/282 (5.7%) |
| Other[†] | 106/2,480 (4.3%) | 84/2,198 (3.8%) | 22/282 (7.8%) |
| Haemoglobin (g/L) | 135.5 (9.9) | 135.7 (10.1) | 134.7 (8.6) |
| Anaemia (Haemoglobin<120g/L) | | | |
| No | 2,354/2,480 (94.9%) | Not displayed[§] | Not displayed[§] |
| Yes | 126/2,480 (5.1%) | Not displayed[§] | <10/282 (<3.5%) |
| Iron deficiency (Ferritin<15ug/L) | | | |
| No | 2,306/2,480 (93.0%) | Not displayed[§] | Not displayed[§] |
| Yes | 174/2,480 (7.0%) | Not displayed[§] | <10/282 (<3.5%) |
| Inflammation (C-Reactive Protein>5mg/L) | | | |
| No | 2,010/2,480 (81.0%) | Not displayed[§] | Not displayed[§] |
| Yes | 470/2,480 (19.0%) | Not displayed[§] | <10/282 (<3.5%) |

AUS = Australia, NHS= National Health Survey, BMI = Body Mass Index, SD = Standard Deviation.
Data are presented as unweighted mean (SD) for continuous measures, and unweighted n/total (%) for categorical measures based on participants with non-missing haemoglobin, ferritin, and C-reactive protein.
*BMI is missing for 176 participants.
[†] Other includes North African and Middle Eastern, American, Sub-Saharan African, Not stated, inadequately described.
[§] Not displayed due to Australian Bureau of Statistics requirements for output clearance.
[1] Ferritin≥15 and ≤150ug/L and C-reactive protein≤5mg/L.

#### Table 3.2.14 AUS NNPAS 2011-2012: Participant characteristics (Females) (18-65 years)

| | Overall sample N=1,593 | Excluded sample N=1,114 | Reference sample N=479 |
|---|---|---|---|
| Age (years) | 44.6 (12.7) | 46.2 (12.7) | 41.0 (11.9) |
| Sex | | | |
| Female | 1,593/1,593 (100.0%) | 1,114/1,114 (100.0%) | 479/479 (100.0%) |
| BMI (kg/m$^2$)* | 27.3 (6.2) | 28.9 (6.6) | 23.8 (2.9) |
| Equivalised income of household | | | |
| bottom quintile | 245/1,504 (16.3%) | 189/1,057 (17.9%) | 56/447 (12.5%) |
| 2nd quintile | 252/1,504 (16.8%) | 192/1,057 (18.2%) | 60/447 (13.4%) |
| 3rd quintile | 307/1,504 (20.4%) | 211/1,057 (20.0%) | 96/447 (21.5%) |
| 4th quintile | 379/1,504 (25.2%) | 247/1,057 (23.4%) | 132/447 (29.5%) |
| highest quintile | 321/1,504 (21.3%) | 218/1,057 (20.6%) | 103/447 (23.0%) |
| Country of birth | | | |
| Australia | 1,140/1,593 (71.6%) | 835/1,114 (75.0%) | 305/479 (63.7%) |
| United Kingdom | 109/1,593 (6.8%) | 66/1,114 (5.9%) | 43/479 (9.0%) |
| Other | 344/1,593 (21.6%) | 213/1,114 (19.1%) | 131/479 (27.3%) |
| Other[†] | 135.2 (9.6) | 135.4 (10.0) | 134.6 (8.4) |
| Haemoglobin (g/L) | | | |
| Anaemia (Haemoglobin<130g/L) | 1,512/1,593 (94.9%) | 1,052/1,114 (94.4%) | 460/479 (96.0%) |
| No | 81/1,593 (5.1%) | 62/1,114 (5.6%) | 19/479 (4.0%) |
| Yes | | | |
| Iron deficiency (Ferritin<15ug/L) | 1,495/1,593 (93.8%) | Not displayed[§] | Not displayed[§] |
| No | 98/1,593 (6.2%) | Not displayed[§] | <10/479 (<2.1%) |
| Yes | | | |
| Inflammation (C-Reactive Protein>5mg/L) | 1,288/1,593 (80.9%) | Not displayed[§] | Not displayed[§] |
| No | 305/1,593 (19.1%) | Not displayed[§] | <10/479 (<2.1%) |

AUS = Australia, NNPAS= National Nutrition and Physical Activity Survey, BMI = Body Mass Index, SD = Standard Deviation.

Data are presented as unweighted mean (SD) for continuous measures, and unweighted n/total (%) for categorical measures based on participants with non-missing haemoglobin, ferritin, and C-reactive protein.

*BMI is missing for 95 participants.

[†] Other: New Zealand, Italy, Viet Nam, China (excl. SARs and Taiwan Province), Greece, Germany, Philippines, India, Oceania and Antarctica (excl. Australia and New Zealand), North-West Europe (excl. United Kingdom and Germany), Southern and Eastern Europe (excl. Italy and Greece), North Africa and the Middle East, South-East Asia (excl. Viet Nam and Philippines), North-East Asia (excl. China), Southern & Central Asia (excl. India), Americas, Sub-Saharan Africa.

[§] Not displayed due to Australian Bureau of Statistics requirements for output clearance.

[1] Ferritin≥15 and ≤150ug/L and C-reactive protein≤5mg/L.

#### 3.3 Demographics Children (6-23 months)

##### Table 3.3.1 CAN TARGet Kids! 2010-2019: Participant characteristics (6-23 months)

| | Overall sample N=1,671 | Excluded sample N=1,248 | Reference sample[1] N=423 |
|---|---|---|---|
| Age (months) | 13.5 (4.2) | 13.8 (4.3) | 12.9 (3.9) |
| Sex | | | |
| Male | 906/1,671 (54.2%) | 699/1,248 (56.0%) | 207/423 (48.9%) |
| Female | 765/1,671 (45.8%) | 549/1,248 (44.0%) | 216/423 (51.1%) |
| Maternal ethnicity | | | |
| European | 934/1,458 (64.1%) | 694/1,093 (63.5%) | 240/365 (65.8%) |
| East Asian | 83/1,458 (5.7%) | 62/1,093 (5.7%) | 21/365 (5.8%) |
| South Asian | 119/1,458 (8.2%) | 93/1,093 (8.5%) | 26/365 (7.1%) |
| Southeast Asian | 45/1,458 (3.1%) | 37/1,093 (3.4%) | 8/365 (2.2%) |
| Arab | 30/1,458 (2.1%) | 20/1,093 (1.8%) | 10/365 (2.7%) |
| African | 94/1,458 (6.4%) | 75/1,093 (6.9%) | 19/365 (5.2%) |
| Latin American | 45/1,458 (3.1%) | 30/1,093 (2.7%) | 15/365 (4.1%) |
| Mixed Ethnicity | 102/1,458 (7.0%) | 76/1,093 (7.0%) | 26/365 (7.1%) |
| Other | 6/1,458 (0.4%) | 6/1,093 (0.5%) | 0/365 (0.0%) |
| Paternal ethnicity | | | |
| European | 932/1,429 (65.2%) | 691/1,070 (64.6%) | 241/359 (67.1%) |
| East Asian | 55/1,429 (3.8%) | 43/1,070 (4.0%) | 12/359 (3.3%) |
| South Asian | 119/1,429 (8.3%) | 92/1,070 (8.6%) | 27/359 (7.5%) |
| Southeast Asian | 35/1,429 (2.4%) | 27/1,070 (2.5%) | 8/359 (2.2%) |
| Arab | 34/1,429 (2.4%) | 28/1,070 (2.6%) | 6/359 (1.7%) |
| African | 117/1,429 (8.2%) | 95/1,070 (8.9%) | 22/359 (6.1%) |
| Latin American | 44/1,429 (3.1%) | 29/1,070 (2.7%) | 15/359 (4.2%) |
| Mixed Ethnicity | 86/1,429 (6.0%) | 58/1,070 (5.4%) | 28/359 (7.8%) |
| Other | 7/1,429 (0.5%) | 7/1,070 (0.7%) | 0/359 (0.0%) |
| Haemoglobin (g/L) | 117.2 (9.6) | 116.9 (10.0) | 117.9 (8.1) |
| Anaemia (Haemoglobin<110g/L) | | | |
| No | 1,348/1,671 (80.7%) | 991/1,248 (79.4%) | 357/423 (84.4%) |
| Yes | 323/1,671 (19.3%) | 257/1,248 (20.6%) | 66/423 (15.6%) |
| Iron deficiency (Ferritin<12ug/L) | | | |
| No | 1,459/1,671 (87.3%) | 1,036/1,248 (83.0%) | 423/423 (100.0%) |
| Yes | 212/1,671 (12.7%) | 212/1,248 (17.0%) | 0/423 (0.0%) |
| Inflammation (C-Reactive Protein>5mg/L) | | | |
| No | 1,568/1,671 (93.8%) | 1,145/1,248 (91.7%) | 423/423 (100.0%) |
| Yes | 103/1,671 (6.2%) | 103/1,248 (8.3%) | 0/423 (0.0%) |

CAN = Canada, TARGet Kids! = The Applied Research Group for Kids, BMI = Body Mass Index, SD = Standard Deviation.
Data are presented as mean (SD) for continuous measures, and n/total (%) for categorical measures based on participants with non-missing haemoglobin, ferritin, and C-reactive protein. Body mass index not presented, z-scores were used as exclusion criteria.
[1] Ferritin≥12ug/L and C-reactive protein≤5mg/L.

#### Table 3.3.2 BAN BRISC 2017-2020: Participant characteristics (11 months)

| | Overall sample N=1,365 | Excluded sample N=1,110 | Reference sample[1] N=255 |
|---|---|---|---|
| Age (months) | 11.1 (0.3) | 11.1 (0.3) | 11.0 (0.3) |
| Sex | | | |
| Female | 689/1,365 (50.5%) | 536/1,110 (48.3%) | 153/255 (60.0%) |
| Male | 676/1,365 (49.5%) | 574/1,110 (51.7%) | 102/255 (40.0%) |
| Wealth index | | | |
| Quintile 1 (relative poorest) | 298/1,364 (21.8%) | 244/1,109 (22.0%) | 54/255 (21.2%) |
| Quintile 2 | 267/1,364 (19.6%) | 227/1,109 (20.5%) | 40/255 (15.7%) |
| Quintile 3 (relative middle) | 264/1,364 (19.4%) | 209/1,109 (18.8%) | 55/255 (21.6%) |
| Quintile 4 | 274/1,364 (20.1%) | 225/1,109 (20.3%) | 49/255 (19.2%) |
| Quintile 5 (relative wealthiest) | 261/1,364 (19.1%) | 204/1,109 (18.4%) | 57/255 (22.4%) |
| Country | | | |
| Bangladesh | 1,365/1,365 (100.0%) | 1,110/1,110 (100.0%) | 255/255 (100.0%) |
| Haemoglobin (g/L) | 115.1 (9.3) | 114.8 (9.5) | 116.5 (8.3) |
| Anaemia (Haemoglobin<110g/L) | | | |
| No | 1,031/1,365 (75.5%) | 826/1,110 (74.4%) | 205/255 (80.4%) |
| Yes | 334/1,365 (24.5%) | 284/1,110 (25.6%) | 50/255 (19.6%) |
| Iron deficiency (Ferritin<12ug/L) | | | |
| No | 1,203/1,365 (88.1%) | 948/1,110 (85.4%) | 255/255 (100.0%) |
| Yes | 162/1,365 (11.9%) | 162/1,110 (14.6%) | 0/255 (0.0%) |
| Inflammation (C-Reactive Protein>5mg/L) | | | |
| No | 1,220/1,365 (89.4%) | 965/1,110 (86.9%) | 255/255 (100.0%) |
| Yes | 145/1,365 (10.6%) | 145/1,110 (13.1%) | 0/255 (0.0%) |

BAN = Bangladesh, BRISC = Benefits and Risks of Iron intervention in Children, SD = Standard Deviation.
Data are presented as mean (SD) for continuous measures, and n/total (%) for categorical measures based on participants with non-missing haemoglobin, ferritin, and C-reactive protein. Body mass index not presented, z-scores were used as exclusion criteria.
[1] Ferritin≥12ug/L and C-reactive protein≤5mg/L.

#### Table 3.3.3 ECU ENSANUT 2011-2013: Participant characteristics (6-23 months)

| | Overall sample N=787 | Excluded sample N=692 | Reference sample[1] N=95 |
|---|---|---|---|
| Age (months) | 14.3 (5.1) | 14.3 (5.1) | 14.1 (5.3) |
| Sex | | | |
| Male | 395/787 (50.2%) | 347/692 (50.1%) | 48/95 (50.5%) |
| Female | 392/787 (49.8%) | 345/692 (49.9%) | 47/95 (49.5%) |
| Economic quintiles | | | |
| Quintile 1 (relative poorest) | 239/787 (30.4%) | 215/692 (31.1%) | 24/95 (25.3%) |
| Quintile 2 | 176/787 (22.4%) | 148/692 (21.4%) | 28/95 (29.5%) |
| Quintile 3 (relative middle) | 157/787 (19.9%) | 133/692 (19.2%) | 24/95 (25.3%) |
| Quintile 4 | 126/787 (16.0%) | 117/692 (16.9%) | 9/95 (9.5%) |
| Quintile 5 (relative wealthiest) | 89/787 (11.3%) | 79/692 (11.4%) | 10/95 (10.5%) |
| Ethnicity (self-reported) | | | |
| Afro-Ecuadorian | 32/787 (4.1%) | 28/692 (4.0%) | 4/95 (4.2%) |
| Indigenous | 78/787 (9.9%) | 70/692 (10.1%) | 8/95 (8.4%) |
| Montubio | 34/787 (4.3%) | 31/692 (4.5%) | 3/95 (3.2%) |
| Rest of the Population | 643/787 (81.7%) | 563/692 (81.4%) | 80/95 (84.2%) |
| Haemoglobin (g/L) | 114.3 (12.7) | 114.4 (13.1) | 113.7 (9.1) |
| Anaemia (Haemoglobin<110g/L) | | | |
| No | 521/787 (66.2%) | 461/692 (66.6%) | 60/95 (63.2%) |
| Yes | 266/787 (33.8%) | 231/692 (33.4%) | 35/95 (36.8%) |
| Iron deficiency (Ferritin<12ug/L) | | | |
| No | 648/787 (82.3%) | 553/692 (79.9%) | 95/95 (100.0%) |
| Yes | 139/787 (17.7%) | 139/692 (20.1%) | 0/95 (0.0%) |
| Inflammation (C-Reactive Protein>5mg/L) | | | |
| No | 682/787 (86.7%) | 587/692 (84.8%) | 95/95 (100.0%) |
| Yes | 105/787 (13.3%) | 105/692 (15.2%) | 0/95 (0.0%) |

ECU = Ecuador, ENSANUT = Encuesta National de Salud y Nutricion, SD = Standard Deviation.
Data are presented as unweighted mean (SD) for continuous measures, and unweighted n/total (%) for categorical measures based on participants with non-missing haemoglobin, ferritin, and C-reactive protein. No information on body mass index was available.
[1] Ferritin≥12ug/L and C-reactive protein≤5mg/L.

#### Table 3.3.4 US NHANES 2003-2018: Participant characteristics (12-23 months)

| | Overall sample N=530 | Excluded sample N=379 | Reference sample[1] N=151 |
|---|---|---|---|
| Age (months) | 18.2 (3.2) | 18.1 (3.2) | 18.5 (3.3) |
| Sex | | | |
| Male | 279/530 (52.6%) | 213/379 (56.2%) | 66/151 (43.7%) |
| Female | 251/530 (47.4%) | 166/379 (43.8%) | 85/151 (56.3%) |
| Poverty | 198/496 (39.9%) | 147/358 (41.1%) | 51/138 (37.0%) |
| Race and Hispanic Origin | | | |
| Mexican American | 139/530 (26.2%) | 100/379 (26.4%) | 39/151 (25.8%) |
| Other Hispanic | 38/530 (7.2%) | 28/379 (7.4%) | 10/151 (6.6%) |
| Non-Hispanic White | 171/530 (32.3%) | 126/379 (33.2%) | 45/151 (29.8%) |
| Non-Hispanic Black | 136/530 (25.7%) | 93/379 (24.5%) | 43/151 (28.5%) |
| Other Race - Including Multi-Racial | 46/530 (8.7%) | 32/379 (8.4%) | 14/151 (9.3%) |
| Haemoglobin (g/L) | 122.7 (8.2) | 122.4 (8.4) | 123.6 (7.5) |
| Anaemia (Haemoglobin<110g/L) | | | |
| No | 500/530 (94.3%) | 354/379 (93.4%) | 146/151 (96.7%) |
| Yes | 30/530 (5.7%) | 25/379 (6.6%) | 5/151 (3.3%) |
| Iron deficiency (Ferritin<12ug/L) | | | |
| No | 461/530 (87.0%) | 310/379 (81.8%) | 151/151 (100.0%) |
| Yes | 69/530 (13.0%) | 69/379 (18.2%) | 0/151 (0.0%) |
| Inflammation (C-Reactive Protein>5mg/L) | | | |
| No | 487/530 (91.9%) | 336/379 (88.7%) | 151/151 (100.0%) |
| Yes | 43/530 (8.1%) | 43/379 (11.3%) | 0/151 (0.0%) |

US = United States, NHANES = National Health and Nutrition Examination Survey, SD = Standard Deviation.
Data are presented as unweighted mean (SD) for continuous measures, and unweighted n/total (%) for categorical measures based on participants with non-missing haemoglobin, ferritin, and C-reactive protein. Data was combined from 1999-2018. No ferritin was measured in participants in survey years 1999-2000, 2001-2002, 2007-2008, 2009-2010, 2011-2012, 2013-2014. No information on body mass index was available.
[1] Ferritin≥12ug/L and C-reactive protein≤5mg/L.

#### 3.4 Demographics Children (24-59 months)

##### Table 3.4.1 CAN TARGet Kids! 2010-2019: Participant characteristics (24-59 months)

| | **Overall sample N=2,050** | **Excluded sample N=1,293** | **Reference sample[1] N=757** |
|---|---|---|---|
| Age (months) | 38.0 (10.6) | 37.5 (10.9) | 38.8 (10.0) |
| Sex | | | |
| Male | 1,075/2,050 (52.4%) | 688/1,293 (53.2%) | 387/757 (51.1%) |
| Female | 975/2,050 (47.6%) | 605/1,293 (46.8%) | 370/757 (48.9%) |
| BMI (kg/m$^2$) | 16.0 (1.5) | 16.1 (1.7) | 15.9 (1.2) |
| Maternal ethnicity | | | |
| European | 1,110/1,802 (61.6%) | 663/1,155 (57.4%) | 447/647 (69.1%) |
| East Asian | 112/1,802 (6.2%) | 77/1,155 (6.7%) | 35/647 (5.4%) |
| South Asian | 197/1,802 (10.9%) | 150/1,155 (13.0%) | 47/647 (7.3%) |
| Southeast Asian | 79/1,802 (4.4%) | 52/1,155 (4.5%) | 27/647 (4.2%) |
| Arab | 35/1,802 (1.9%) | 23/1,155 (2.0%) | 12/647 (1.9%) |
| African | 106/1,802 (5.9%) | 77/1,155 (6.7%) | 29/647 (4.5%) |
| Latin American | 56/1,802 (3.1%) | 40/1,155 (3.5%) | 16/647 (2.5%) |
| Mixed Ethnicity | 100/1,802 (5.5%) | 69/1,155 (6.0%) | 31/647 (4.8%) |
| Other | 7/1,802 (0.4%) | 4/1,155 (0.3%) | 3/647 (0.5%) |
| Paternal ethnicity | | | |
| European | 1,132/1,782 (63.5%) | 678/1,142 (59.4%) | 454/640 (70.9%) |
| East Asian | 79/1,782 (4.4%) | 53/1,142 (4.6%) | 26/640 (4.1%) |
| South Asian | 201/1,782 (11.3%) | 153/1,142 (13.4%) | 48/640 (7.5%) |
| Southeast Asian | 57/1,782 (3.2%) | 37/1,142 (3.2%) | 20/640 (3.1%) |
| Arab | 44/1,782 (2.5%) | 30/1,142 (2.6%) | 14/640 (2.2%) |
| African | 131/1,782 (7.4%) | 91/1,142 (8.0%) | 40/640 (6.3%) |
| Latin American | 46/1,782 (2.6%) | 32/1,142 (2.8%) | 14/640 (2.2%) |
| Mixed Ethnicity | 85/1,782 (4.8%) | 62/1,142 (5.4%) | 23/640 (3.6%) |
| Other | 7/1,782 (0.4%) | 6/1,142 (0.5%) | 1/640 (0.2%) |
| Haemoglobin (g/L) | 120.2 (8.1) | 119.7 (8.4) | 121.0 (7.3) |
| Anaemia (Haemoglobin<110g/L) | | | |
| No | 1,889/2,050 (92.1%) | 1,172/1,293 (90.6%) | 717/757 (94.7%) |
| Yes | 161/2,050 (7.9%) | 121/1,293 (9.4%) | 40/757 (5.3%) |
| Iron deficiency (Ferritin<12ug/L) | | | |
| No | 1,911/2,050 (93.2%) | 1,154/1,293 (89.2%) | 757/757 (100.0%) |
| Yes | 139/2,050 (6.8%) | 139/1,293 (10.8%) | 0/757 (0.0%) |
| Inflammation (C-Reactive Protein>5mg/L) | | | |
| No | 1,906/2,050 (93.0%) | 1,149/1,293 (88.9%) | 757/757 (100.0%) |
| Yes | 144/2,050 (7.0%) | 144/1,293 (11.1%) | 0/757 (0.0%) |

CAN = Canada, TARGet Kids! = The Applied Research Group for Kids, BMI = Body Mass Index, SD = Standard Deviation.
Data are presented as mean (SD) for continuous measures, and n/total (%) for categorical measures based on participants with non-missing haemoglobin, ferritin, and C-reactive protein.
[1] Ferritin≥12ug/L and C-reactive protein≤5mg/L.

#### Table 3.4.2 ECU ENSANUT 2011-2013: Participant characteristics (24-59 months)

| | Overall sample N=1,260 | Excluded sample N=1,080 | Reference sample[1] N=180 |
|---|---|---|---|
| Age (months) | 39.6 (10.4) | 39.6 (10.6) | 39.6 (9.7) |
| Sex | | | |
| Male | 644/1,260 (51.1%) | 556/1,080 (51.5%) | 88/180 (48.9%) |
| Female | 616/1,260 (48.9%) | 524/1,080 (48.5%) | 92/180 (51.1%) |
| Economic quintiles | | | |
| Quintile 1 (relative poorest) | 362/1,260 (28.7%) | 319/1,080 (29.5%) | 43/180 (23.9%) |
| Quintile 2 | 309/1,260 (24.5%) | 270/1,080 (25.0%) | 39/180 (21.7%) |
| Quintile 3 (relative middle) | 263/1,260 (20.9%) | 220/1,080 (20.4%) | 43/180 (23.9%) |
| Quintile 4 | 192/1,260 (15.2%) | 159/1,080 (14.7%) | 33/180 (18.3%) |
| Quintile 5 (relative wealthiest) | 134/1,260 (10.6%) | 112/1,080 (10.4%) | 22/180 (12.2%) |
| Ethnicity (self-reported) | | | |
| Afro-Ecuadorian | 41/1,260 (3.3%) | 32/1,080 (3.0%) | 9/180 (5.0%) |
| Indigenous | 124/1,260 (9.8%) | 112/1,080 (10.4%) | 12/180 (6.7%) |
| Montubio | 64/1,260 (5.1%) | 56/1,080 (5.2%) | 8/180 (4.4%) |
| Rest of the Population | 1,031/1,260 (81.8%) | 880/1,080 (81.5%) | 151/180 (83.9%) |
| Haemoglobin (g/L) | 125.0 (11.0) | 125.5 (11.3) | 121.7 (8.3) |
| Anaemia (Haemoglobin<110g/L) | | | |
| No | 1,189/1,260 (94.4%) | 1,020/1,080 (94.4%) | 169/180 (93.9%) |
| Yes | 71/1,260 (5.6%) | 60/1,080 (5.6%) | 11/180 (6.1%) |
| Iron deficiency (Ferritin<12ug/L) | | | |
| No | 1,210/1,260 (96.0%) | 1,030/1,080 (95.4%) | 180/180 (100.0%) |
| Yes | 50/1,260 (4.0%) | 50/1,080 (4.6%) | 0/180 (0.0%) |
| Inflammation (C-Reactive Protein>5mg/L) | | | |
| No | 1,112/1,260 (88.3%) | 932/1,080 (86.3%) | 180/180 (100.0%) |
| Yes | 148/1,260 (11.7%) | 148/1,080 (13.7%) | 0/180 (0.0%) |

ECU = Ecuador, ENSANUT = Encuesta National de Salud y Nutricion, SD = Standard Deviation.
Data are presented as unweighted mean (SD) for continuous measures, and unweighted n/total (%) for categorical measures based on participants with non-missing haemoglobin, ferritin, and C-reactive protein.
[1] Ferritin≥12ug/L and C-reactive protein≤5mg/L.

#### Table 3.4.3 US NHANES 1999-2018: Participant characteristics (24-59 months)

| | Overall sample N=2,559 | Excluded sample N=1,622 | Reference sample[1] N=937 |
|---|---|---|---|
| Age (months) | 44.5 (9.6) | 44.1 (9.7) | 45.1 (9.5) |
| Sex | | | |
| Male | 1,334/2,559 (52.1%) | 869/1,622 (53.6%) | 465/937 (49.6%) |
| Female | 1,225/2,559 (47.9%) | 753/1,622 (46.4%) | 472/937 (50.4%) |
| BMI (kg/m$^2$) | 16.4 (1.8) | 16.7 (2.1) | 16.0 (1.1) |
| Poverty | 902/2,370 (38.1%) | 585/1,500 (39.0%) | 317/870 (36.4%) |
| Race and Hispanic Origin | | | |
| Mexican American | 700/2,559 (27.4%) | 476/1,622 (29.3%) | 224/937 (23.9%) |
| Other Hispanic | 210/2,559 (8.2%) | 142/1,622 (8.8%) | 68/937 (7.3%) |
| Non-Hispanic White | 784/2,559 (30.6%) | 488/1,622 (30.1%) | 296/937 (31.6%) |
| Non-Hispanic Black | 645/2,559 (25.2%) | 389/1,622 (24.0%) | 256/937 (27.3%) |
| Other Race - Including Multi-Racial | 220/2,559 (8.6%) | 127/1,622 (7.8%) | 93/937 (9.9%) |
| Haemoglobin (g/L) | 125.3 (8.2) | 124.9 (8.5) | 125.8 (7.8) |
| Anaemia (Haemoglobin<110g/L) | | | |
| No | 2,492/2,559 (97.4%) | 1,570/1,622 (96.8%) | 922/937 (98.4%) |
| Yes | 67/2,559 (2.6%) | 52/1,622 (3.2%) | 15/937 (1.6%) |
| Iron deficiency (Ferritin<12ug/L) | | | |
| No | 2,365/2,559 (92.4%) | 1,428/1,622 (88.0%) | 937/937 (100.0%) |
| Yes | 194/2,559 (7.6%) | 194/1,622 (12.0%) | 0/937 (0.0%) |
| Inflammation (C-Reactive Protein>5mg/L) | | | |
| No | 2,390/2,559 (93.4%) | 1,453/1,622 (89.6%) | 937/937 (100.0%) |
| Yes | 169/2,559 (6.6%) | 169/1,622 (10.4%) | 0/937 (0.0%) |

US = United States, NHANES = National Health and Nutrition Examination Survey, BMI = Body Mass Index, SD = Standard Deviation.
Data are presented as unweighted mean (SD) for continuous measures, and unweighted n/total (%) for categorical measures based on participants with non-missing haemoglobin, ferritin, and C-reactive protein. Data was combined from 1999-2018. No ferritin was measured in participants less than 3 years in survey years 1999-2000, 2001-2002, 2007-2008 and 2009-2010. No ferritin was measured in participants in survey years 2011-2012 and 2013-2014.
[1] Ferritin≥12ug/L and C-reactive protein≤5mg/L.

#### 3.5 Demographics Children (5-11 years)

##### Table 3.5.1 CAN TARGet Kids! 2010-2019: Participant characteristics (5-11 years)

| | Overall sample N=979 | Excluded sample N=587 | Reference sample[1] N=392 |
|---|---|---|---|
| Age (months) | 6.2 (1.4) | 6.3 (1.5) | 6.2 (1.3) |
| Sex | | | |
| Male | 528/979 (53.9%) | 329/587 (56.0%) | 199/392 (50.8%) |
| Female | 451/979 (46.1%) | 258/587 (44.0%) | 193/392 (49.2%) |
| BMI (kg/m$^2$) | 15.9 (2.1) | 16.2 (2.4) | 15.5 (1.4) |
| Maternal ethnicity | | | |
| European | 587/923 (63.6%) | 343/555 (61.8%) | 244/368 (66.3%) |
| East Asian | 81/923 (8.8%) | 46/555 (8.3%) | 35/368 (9.5%) |
| South Asian | 70/923 (7.6%) | 50/555 (9.0%) | 20/368 (5.4%) |
| Southeast Asian | 33/923 (3.6%) | 19/555 (3.4%) | 14/368 (3.8%) |
| Arab | 16/923 (1.7%) | 10/555 (1.8%) | 6/368 (1.6%) |
| African | 46/923 (5.0%) | 34/555 (6.1%) | 12/368 (3.3%) |
| Latin American | 35/923 (3.8%) | 24/555 (4.3%) | 11/368 (3.0%) |
| Mixed Ethnicity | 53/923 (5.7%) | 28/555 (5.0%) | 25/368 (6.8%) |
| Other | 2/923 (0.2%) | 1/555 (0.2%) | 1/368 (0.3%) |
| Paternal ethnicity | | | |
| European | 582/888 (65.5%) | 342/535 (63.9%) | 240/353 (68.0%) |
| East Asian | 42/888 (4.7%) | 25/535 (4.7%) | 17/353 (4.8%) |
| South Asian | 81/888 (9.1%) | 52/535 (9.7%) | 29/353 (8.2%) |
| Southeast Asian | 20/888 (2.3%) | 9/535 (1.7%) | 11/353 (3.1%) |
| Arab | 15/888 (1.7%) | 9/535 (1.7%) | 6/353 (1.7%) |
| African | 61/888 (6.9%) | 43/535 (8.0%) | 18/353 (5.1%) |
| Latin American | 30/888 (3.4%) | 19/535 (3.6%) | 11/353 (3.1%) |
| Mixed Ethnicity | 55/888 (6.2%) | 35/535 (6.5%) | 20/353 (5.7%) |
| Other | 2/888 (0.2%) | 1/535 (0.2%) | 1/353 (0.3%) |
| Haemoglobin (g/L) | 124.1 (7.9) | 124.2 (8.4) | 123.9 (7.2) |
| Anaemia (Haemoglobin<115g/L) | | | |
| No | 874/979 (89.3%) | 514/587 (87.6%) | 360/392 (91.8%) |
| Yes | 105/979 (10.7%) | 73/587 (12.4%) | 32/392 (8.2%) |
| Iron deficiency (Ferritin<15ug/L) | | | |
| No | 926/979 (94.6%) | 534/587 (91.0%) | 392/392 (100.0%) |
| Yes | 53/979 (5.4%) | 53/587 (9.0%) | 0/392 (0.0%) |
| Inflammation (C-Reactive Protein>5mg/L) | | | |
| No | 934/979 (95.4%) | 542/587 (92.3%) | 392/392 (100.0%) |
| Yes | 45/979 (4.6%) | 45/587 (7.7%) | 0/392 (0.0%) |

CAN = Canada, TARGet Kids! = The Applied Research Group for Kids, BMI = Body Mass Index, SD = Standard Deviation.
Data are presented as mean (SD) for continuous measures, and n/total (%) for categorical measures based on participants with non-missing haemoglobin, ferritin, and C-reactive protein.
[1] Ferritin≥15ug/L and C-reactive protein≤5mg/L.

#### Table 3.5.2 US NHANES 1999-2018: Participant characteristics (5-11 years)

| | Overall sample N=3,030 | Excluded sample N=1,829 | Reference sample[1] N=1,201 |
|---|---|---|---|
| Age (years) | 7.7 (2.2) | 7.7 (2.2) | 7.7 (2.2) |
| Sex | | | |
| Male | 1,531/3,030 (50.5%) | 947/1,829 (51.8%) | 584/1,201 (48.6%) |
| Female | 1,499/3,030 (49.5%) | 882/1,829 (48.2%) | 617/1,201 (51.4%) |
| BMI (kg/m$^2$) | 17.8 (3.7) | 18.5 (4.3) | 16.6 (2.1) |
| Poverty | 1,024/2,778 (36.9%) | 594/1,680 (35.4%) | 430/1,098 (39.2%) |
| Race and Hispanic Origin | | | |
| Mexican American | 973/3,030 (32.1%) | 581/1,829 (31.8%) | 392/1,201 (32.6%) |
| Other Hispanic | 166/3,030 (5.5%) | 111/1,829 (6.1%) | 55/1,201 (4.6%) |
| Non-Hispanic White | 818/3,030 (27.0%) | 500/1,829 (27.3%) | 318/1,201 (26.5%) |
| Non-Hispanic Black | 910/3,030 (30.0%) | 530/1,829 (29.0%) | 380/1,201 (31.6%) |
| Other Race - Including Multi-Racial | 163/3,030 (5.4%) | 107/1,829 (5.9%) | 56/1,201 (4.7%) |
| Haemoglobin (g/L) | 130.6 (9.0) | 130.4 (9.0) | 130.8 (9.1) |
| Anaemia (Haemoglobin<115g/L) | | | |
| No | 2,944/3,030 (97.2%) | 1,778/1,829 (97.2%) | 1,166/1,201 (97.1%) |
| Yes | 86/3,030 (2.8%) | 51/1,829 (2.8%) | 35/1,201 (2.9%) |
| Iron deficiency (Ferritin<15ug/L) | | | |
| No | 2,798/3,030 (92.3%) | 1,597/1,829 (87.3%) | 1,201/1,201 (100.0%) |
| Yes | 232/3,030 (7.7%) | 232/1,829 (12.7%) | 0/1,201 (0.0%) |
| Inflammation (C-Reactive Protein>5mg/L) | | | |
| No | 2,824/3,030 (93.2%) | 1,623/1,829 (88.7%) | 1,201/1,201 (100.0%) |
| Yes | 206/3,030 (6.8%) | 206/1,829 (11.3%) | 0/1,201 (0.0%) |

US = United States, NHANES = National Health and Nutrition Examination Survey, BMI = Body Mass Index, SD = Standard Deviation.
Data are presented as unweighted mean (SD) for continuous measures, and unweighted n/total (%) for categorical measures based on participants with non-missing haemoglobin, ferritin, and C-reactive protein. Data was combined from 1999-2018. No ferritin was measured in participants 6 to 11 years in survey years 2003-2004, 2005-2006, 2007-2008, 2009-2010, 2015-2016, 2017-2018. No ferritin was measured in participants in survey years 2011-2012 and 2013-2014.
[1] Ferritin≥15ug/L and C-reactive protein≤5mg/L.

#### Table 3.5.3 CHN CHNS 2009: Participant characteristics (5-11 years)

| | Overall sample N=404 | Excluded sample N=155 | Reference sample[1] N=249 |
|---|---|---|---|
| Age (years) | 9.0 (1.5) | 9.0 (1.6) | 9.0 (1.5) |
| Sex | | | |
| Male | 227/404 (56.2%) | 84/155 (54.2%) | 143/249 (57.4%) |
| Female | 177/404 (43.8%) | 71/155 (45.8%) | 106/249 (42.6%) |
| BMI (kg/m$^2$) | 16.3 (2.9) | 16.7 (3.9) | 16.1 (2.0) |
| Nationality | | | |
| Han | 338/403 (83.9%) | 112/155 (72.3%) | 226/248 (91.1%) |
| Hui | 1/403 (0.2%) | 0/155 (0.0%) | 1/248 (0.4%) |
| Miao | 16/403 (4.0%) | 8/155 (5.2%) | 8/248 (3.2%) |
| Zhuang | 5/403 (1.2%) | 1/155 (0.6%) | 4/248 (1.6%) |
| Buyi | 11/403 (2.7%) | 11/155 (7.1%) | 0/248 (0.0%) |
| Korean | 2/403 (0.5%) | 1/155 (0.6%) | 1/248 (0.4%) |
| Man | 7/403 (1.7%) | 2/155 (1.3%) | 5/248 (2.0%) |
| Tujia | 19/403 (4.7%) | 19/155 (12.3%) | 0/248 (0.0%) |
| Other | 4/403 (1.0%) | 1/155 (0.6%) | 3/248 (1.2%) |
| Haemoglobin (g/L) | 133.9 (15.2) | 133.1 (13.8) | 134.3 (16.1) |
| Anaemia (Haemoglobin<120g/L) | | | |
| No | 380/404 (94.1%) | 150/155 (96.8%) | 230/249 (92.4%) |
| Yes | 24/404 (5.9%) | 5/155 (3.2%) | 19/249 (7.6%) |
| Iron deficiency (Ferritin<15ug/L) | | | |
| No | 392/404 (97.0%) | 143/155 (92.3%) | 249/249 (100.0%) |
| Yes | 12/404 (3.0%) | 12/155 (7.7%) | 0/249 (0.0%) |
| Inflammation (C-Reactive Protein>5mg/L) | | | |
| No | 383/404 (94.8%) | 134/155 (86.5%) | 249/249 (100.0%) |
| Yes | 21/404 (5.2%) | 21/155 (13.5%) | 0/249 (0.0%) |

CHN = China, CHNS = China Health and Nutrition Survey, BMI = Body Mass Index, SD = Standard Deviation.
Data are presented as unweighted mean (SD) for continuous measures, and unweighted n/total (%) for categorical measures based on participants with non-missing haemoglobin, ferritin, and C-reactive protein.
[1] Ferritin≥15ug/L and C-reactive protein≤5mg/L.

#### 3.6 Demographics Children (12-17 years) males

##### Table 3.6.1 US NHANES 1999-2000: Participant characteristics (Males) (12-17 years)

| | **Overall sample N=836** | **Excluded sample N=502** | **Reference sample[1] N=334** |
|---|---|---|---|
| Age (years) | 15.0 (1.8) | 14.9 (1.7) | 15.1 (1.9) |
| Sex | | | |
| Male | 836/836 (100.0%) | 502/502 (100.0%) | 334/334 (100.0%) |
| BMI (kg/m$^2$) | 23.2 (5.8) | 24.7 (6.7) | 21.0 (3.1) |
| Poverty | 269/730 (36.8%) | 148/433 (34.2%) | 121/297 (40.7%) |
| Race and Hispanic Origin | | | |
| Mexican American | 371/836 (44.4%) | 225/502 (44.8%) | 146/334 (43.7%) |
| Other Hispanic | 38/836 (4.5%) | 22/502 (4.4%) | 16/334 (4.8%) |
| Non-Hispanic White | 159/836 (19.0%) | 102/502 (20.3%) | 57/334 (17.1%) |
| Non-Hispanic Black | 243/836 (29.1%) | 140/502 (27.9%) | 103/334 (30.8%) |
| Other Race - Including Multi-Racial | 25/836 (3.0%) | 13/502 (2.6%) | 12/334 (3.6%) |
| Haemoglobin (g/L) | 146.5 (11.7) | 145.8 (11.5) | 147.6 (12.0) |
| Anaemia (Haemoglobin<120g/L [<15 years] or 130g/L [>=15 years]) | | | |
| No | 818/836 (97.8%) | 490/502 (97.6%) | 328/334 (98.2%) |
| Yes | 18/836 (2.2%) | 12/502 (2.4%) | 6/334 (1.8%) |
| Iron deficiency (Ferritin<15ug/L) | | | |
| No | 791/836 (94.6%) | 457/502 (91.0%) | 334/334 (100.0%) |
| Yes | 45/836 (5.4%) | 45/502 (9.0%) | 0/334 (0.0%) |
| Inflammation (C-Reactive Protein>5mg/L) | | | |
| No | 785/836 (93.9%) | 451/502 (89.8%) | 334/334 (100.0%) |
| Yes | 51/836 (6.1%) | 51/502 (10.2%) | 0/334 (0.0%) |

US = United States, NHANES = National Health and Nutrition Examination Survey, BMI = Body Mass Index, SD = Standard Deviation.
Data are presented as unweighted mean (SD) for continuous measures, and unweighted n/total (%) for categorical measures based on participants with non-missing haemoglobin, ferritin, and C-reactive protein.
[1] Ferritin≥15ug/L and C-reactive protein≤5mg/L.

#### Table 3.6.2 US NHANES 2001-2002: Participant characteristics (Males) (12-17 years)

| | Overall sample N=820 | Excluded sample N=529 | Reference sample[1] N=291 |
|---|---|---|---|
| Age (years) | 15.1 (1.8) | 15.2 (1.8) | 15.1 (1.7) |
| Sex | | | |
| Male | 820/820 (100.0%) | 529/529 (100.0%) | 291/291 (100.0%) |
| BMI (kg/m$^2$) | 23.0 (5.5) | 24.2 (6.1) | 20.9 (3.2) |
| Poverty | 206/760 (27.1%) | 132/491 (26.9%) | 74/269 (27.5%) |
| Race and Hispanic Origin | | | |
| Mexican American | 246/820 (30.0%) | 155/529 (29.3%) | 91/291 (31.3%) |
| Other Hispanic | 37/820 (4.5%) | 22/529 (4.2%) | 15/291 (5.2%) |
| Non-Hispanic White | 243/820 (29.6%) | 167/529 (31.6%) | 76/291 (26.1%) |
| Non-Hispanic Black | 258/820 (31.5%) | 163/529 (30.8%) | 95/291 (32.6%) |
| Other Race - Including Multi-Racial | 36/820 (4.4%) | 22/529 (4.2%) | 14/291 (4.8%) |
| Haemoglobin (g/L) | 147.1 (12.3) | 146.9 (12.6) | 147.7 (11.8) |
| Anaemia (Haemoglobin<120g/L [<15 years] or 130g/L [>=15 years]) | | | |
| No | 804/820 (98.0%) | 515/529 (97.4%) | 289/291 (99.3%) |
| Yes | 16/820 (2.0%) | 14/529 (2.6%) | 2/291 (0.7%) |
| Iron deficiency (Ferritin<15ug/L) | | | |
| No | 765/820 (93.3%) | 474/529 (89.6%) | 291/291 (100.0%) |
| Yes | 55/820 (6.7%) | 55/529 (10.4%) | 0/291 (0.0%) |
| Inflammation (C-Reactive Protein>5mg/L) | | | |
| No | 766/820 (93.4%) | 475/529 (89.8%) | 291/291 (100.0%) |
| Yes | 54/820 (6.6%) | 54/529 (10.2%) | 0/291 (0.0%) |

US = United States, NHANES = National Health and Nutrition Examination Survey, BMI = Body Mass Index, SD = Standard Deviation.

Data are presented as unweighted mean (SD) for continuous measures, and unweighted n/total (%) for categorical measures based on participants with non-missing haemoglobin, ferritin, and C-reactive protein.

[1] Ferritin≥15ug/L and C-reactive protein≤5mg/L.

#### Table 3.6.3 US NHANES 2017-2018: Participant characteristics (Males) (12-17 years)

| | Overall sample N=383 | Excluded sample N=242 | Reference sample[1] N=141 |
|---|---|---|---|
| Age (years) | 15.0 (1.8) | 15.1 (1.8) | 14.9 (1.8) |
| Sex | | | |
| Male | 383/383 (100.0%) | 242/242 (100.0%) | 141/141 (100.0%) |
| BMI (kg/m$^2$) | 23.5 (6.1) | 25.3 (6.7) | 20.5 (3.0) |
| Poverty | 80/346 (23.1%) | 48/219 (21.9%) | 32/127 (25.2%) |
| Race and Hispanic Origin | | | |
| Mexican American | 59/383 (15.4%) | 42/242 (17.4%) | 17/141 (12.1%) |
| Other Hispanic | 29/383 (7.6%) | 15/242 (6.2%) | 14/141 (9.9%) |
| Non-Hispanic White | 124/383 (32.4%) | 82/242 (33.9%) | 42/141 (29.8%) |
| Non-Hispanic Black | 80/383 (20.9%) | 46/242 (19.0%) | 34/141 (24.1%) |
| Non-Hispanic Asian | 51/383 (13.3%) | 30/242 (12.4%) | 21/141 (14.9%) |
| Other Race - Including Multi-Racial | 40/383 (10.4%) | 27/242 (11.2%) | 13/141 (9.2%) |
| Haemoglobin (g/L) | 145.2 (12.2) | 145.4 (13.3) | 144.8 (10.3) |
| Anaemia (Haemoglobin<120g/L [<15 years] or 130g/L [>=15 years]) | | | |
| No | 376/383 (98.2%) | 236/242 (97.5%) | 140/141 (99.3%) |
| Yes | 7/383 (1.8%) | 6/242 (2.5%) | 1/141 (0.7%) |
| Iron deficiency (Ferritin<15ug/L) | | | |
| No | 367/383 (95.8%) | 226/242 (93.4%) | 141/141 (100.0%) |
| Yes | 16/383 (4.2%) | 16/242 (6.6%) | 0/141 (0.0%) |
| Inflammation (C-Reactive Protein>5mg/L) | | | |
| No | 351/383 (91.6%) | 210/242 (86.8%) | 141/141 (100.0%) |
| Yes | 32/383 (8.4%) | 32/242 (13.2%) | 0/141 (0.0%) |

US = United States, NHANES = National Health and Nutrition Examination Survey, BMI = Body Mass Index, SD = Standard Deviation.
Data are presented as unweighted mean (SD) for continuous measures, and unweighted n/total (%) for categorical measures based on participants with non-missing haemoglobin, ferritin, and C-reactive protein.
[1] Ferritin≥15ug/L and C-reactive protein≤5mg/L.

#### Table 3.6.4 AUS NHS and NNPAS 2011-2012: Participant characteristics (Males) (12-17 years)

| | **Overall sample N=358** | **Excluded sample N=151** | **Reference sample N=207** |
|---|---|---|---|
| Age (years) | 14.6 (1.7) | 14.6 (1.6) | 14.6 (1.7) |
| Sex | | | |
| Male | 358/358 (100.0%) | 151/151 (100.0%) | 207/207 (100.0%) |
| BMI (kg/m$^2$)* | 21.5 (3.7) | 22.3 (4.6) | 20.9 (2.9) |
| Equivalised income of household | | | |
| bottom quintile | 36/312 (11.5%) | 11/133 (8.3%) | 25/179 (14.0%) |
| 2nd quintile | 63/312 (20.2%) | 28/133 (21.1%) | 35/179 (19.6%) |
| 3rd quintile | 84/312 (26.9%) | 40/133 (30.1%) | 44/179 (24.6%) |
| 4th quintile | 86/312 (27.6%) | 36/133 (27.1%) | 50/179 (27.9%) |
| highest quintile | 43/312 (13.8%) | 18/133 (13.5%) | 25/179 (14.0%) |
| Haemoglobin (g/L) | 147.7 (11.1) | 147.5 (10.2) | 147.8 (11.7) |
| Anaemia (Haemoglobin<120g/L [<15 years] or 130g/L [≥15 years]) | | | |
| No | Not displayed[†] | Not displayed[†] | Not displayed[†] |
| Yes | <10/358 (<2.8%) | Not displayed[†] | Not displayed[†] |
| Iron deficiency (Ferritin<15ug/L) | | | |
| No | Not displayed[†] | Not displayed[†] | Not displayed[†] |
| Yes | <10/358 (<2.8%) | Not displayed[†] | Not displayed[†] |
| Inflammation (C-Reactive Protein>5mg/L) | | | |
| No | 333/358 (93.0%) | Not displayed[†] | Not displayed[†] |
| Yes | 25/358 (7.0%) | Not displayed[†] | <10/207 (<4.8%) |

AUS = Australia, NHS = National Health Survey, NNPAS = National Nutrition and Physical Activity Survey, BMI = Body Mass Index, SD = Standard Deviation.
Data are presented as unweighted mean (SD) for continuous measures, and unweighted n/total (%) for categorical measures based on participants with non-missing haemoglobin, ferritin, and C-reactive protein. Data was combined from NHS and NNPAS 2011-2012.
*BMI is missing for 21 participants.
[†] Not presented due to Australian Bureau of Statistics requirements for output clearance.
[1] Ferritin≥15ug/L and C-reactive protein≤5mg/L.

#### Table 3.6.5 CHN CHNS 2009: Participant characteristics (Males) (12-17 years)

| | **Overall sample N=223** | **Excluded sample N=93** | **Reference sample[1] N=130** |
|---|---|---|---|
| Age (years) | 14.1 (1.7) | 14.1 (1.6) | 14.1 (1.7) |
| Sex | | | |
| Male | 223/223 (100.0%) | 93/93 (100.0%) | 130/130 (100.0%) |
| BMI (kg/m$^2$) | 19.0 (3.5) | 19.0 (4.5) | 19.1 (2.6) |
| Nationality | | | |
| Han | 192/223 (86.1%) | 70/93 (75.3%) | 122/130 (93.8%) |
| Vaguer | 1/223 (0.4%) | 1/93 (1.1%) | 0/130 (0.0%) |
| Miao | 4/223 (1.8%) | 4/93 (4.3%) | 0/130 (0.0%) |
| Zhuang | 3/223 (1.3%) | 1/93 (1.1%) | 2/130 (1.5%) |
| Buyi | 8/223 (3.6%) | 8/93 (8.6%) | 0/130 (0.0%) |
| Man | 4/223 (1.8%) | 1/93 (1.1%) | 3/130 (2.3%) |
| Tujia | 8/223 (3.6%) | 7/93 (7.5%) | 1/130 (0.8%) |
| Other | 3/223 (1.3%) | 1/93 (1.1%) | 2/130 (1.5%) |
| Haemoglobin (g/L) | 146.3 (15.0) | 145.5 (16.8) | 146.8 (13.7) |
| Anaemia (Haemoglobin<120g/L [<15 years] or 130g/L [>=15 years]) | | | |
| No | 211/223 (94.6%) | 86/93 (92.5%) | 125/130 (96.2%) |
| Yes | 12/223 (5.4%) | 7/93 (7.5%) | 5/130 (3.8%) |
| Iron deficiency (Ferritin<15ug/L) | | | |
| No | 213/223 (95.5%) | 83/93 (89.2%) | 130/130 (100.0%) |
| Yes | 10/223 (4.5%) | 10/93 (10.8%) | 0/130 (0.0%) |
| Inflammation (C-Reactive Protein>5mg/L) | | | |
| No | 208/223 (93.3%) | 78/93 (83.9%) | 130/130 (100.0%) |
| Yes | 15/223 (6.7%) | 15/93 (16.1%) | 0/130 (0.0%) |

CHN = China, CHNS = China Health and Nutrition Survey, BMI = Body Mass Index, SD = Standard Deviation.
Data are presented as unweighted mean (SD) for continuous measures, and unweighted n/total (%) for categorical measures based on participants with non-missing haemoglobin, ferritin, and C-reactive protein.
[1] Ferritin≥15ug/L and C-reactive protein≤5mg/L.

#### 3.7 Demographics Children (12-17 years) females

##### Table 3.7.1 US NHANES 1999-2000: Participant characteristics (Females) (12-17 years)

| | **Overall sample N=793** | **Excluded sample N=560** | **Reference sample[1] N=233** |
|---|---|---|---|
| Age (years) | 14.9 (1.7) | 15.0 (1.8) | 14.6 (1.7) |
| Sex | | | |
| Female | 793/793 (100.0%) | 560/560 (100.0%) | 233/233 (100.0%) |
| BMI (kg/m$^2$) | 23.9 (5.7) | 24.9 (6.2) | 21.4 (2.9) |
| Poverty | 255/701 (36.4%) | 181/497 (36.4%) | 74/204 (36.3%) |
| Race and Hispanic Origin | | | |
| Mexican American | 332/793 (41.9%) | 241/560 (43.0%) | 91/233 (39.1%) |
| Other Hispanic | 36/793 (4.5%) | 27/560 (4.8%) | 9/233 (3.9%) |
| Non-Hispanic White | 160/793 (20.2%) | 112/560 (20.0%) | 48/233 (20.6%) |
| Non-Hispanic Black | 231/793 (29.1%) | 157/560 (28.0%) | 74/233 (31.8%) |
| Other Race - Including Multi-Racial | 34/793 (4.3%) | 23/560 (4.1%) | 11/233 (4.7%) |
| Haemoglobin (g/L) | 133.3 (9.5) | 132.9 (10.0) | 134.4 (8.3) |
| Anaemia (Haemoglobin<120g/L) | | | |
| No | 736/793 (92.8%) | 518/560 (92.5%) | 218/233 (93.6%) |
| Yes | 57/793 (7.2%) | 42/560 (7.5%) | 15/233 (6.4%) |
| Iron deficiency (Ferritin<15ug/L) | | | |
| No | 632/793 (79.7%) | 399/560 (71.3%) | 233/233 (100.0%) |
| Yes | 161/793 (20.3%) | 161/560 (28.7%) | 0/233 (0.0%) |
| Inflammation (C-Reactive Protein>5mg/L) | | | |
| No | 717/793 (90.4%) | 484/560 (86.4%) | 233/233 (100.0%) |
| Yes | 76/793 (9.6%) | 76/560 (13.6%) | 0/233 (0.0%) |

US = United States, NHANES = National Health and Nutrition Examination Survey, BMI = Body Mass Index, SD = Standard Deviation.
Data are presented as unweighted mean (SD) for continuous measures, and unweighted n/total (%) for categorical measures based on participants with non-missing haemoglobin, ferritin, and C-reactive protein.
[1] Ferritin≥15ug/L and C-reactive protein≤5mg/L.

#### Table 3.7.2 US NHANES 2001-2002: Participant characteristics (Females) (12-17 years)

| | **Overall sample N=879** | **Excluded sample N=634** | **Reference sample[1] N=245** |
|---|---|---|---|
| Age (years) | 15.0 (1.8) | 15.1 (1.8) | 14.6 (1.7) |
| Sex | | | |
| Female | 879/879 (100.0%) | 634/634 (100.0%) | 245/245 (100.0%) |
| BMI (kg/m$^2$) | 23.3 (5.6) | 24.2 (6.1) | 21.0 (3.0) |
| Poverty | 227/841 (27.0%) | 166/609 (27.3%) | 61/232 (26.3%) |
| Race and Hispanic Origin | | | |
| Mexican American | 287/879 (32.7%) | 206/634 (32.5%) | 81/245 (33.1%) |
| Other Hispanic | 44/879 (5.0%) | 29/634 (4.6%) | 15/245 (6.1%) |
| Non-Hispanic White | 262/879 (29.8%) | 185/634 (29.2%) | 77/245 (31.4%) |
| Non-Hispanic Black | 260/879 (29.6%) | 197/634 (31.1%) | 63/245 (25.7%) |
| Other Race - Including Multi-Racial | 26/879 (3.0%) | 17/634 (2.7%) | 9/245 (3.7%) |
| Haemoglobin (g/L) | 133.1 (10.8) | 132.3 (11.1) | 135.3 (9.7) |
| Anaemia (Haemoglobin<120g/L) | | | |
| No | 801/879 (91.1%) | 567/634 (89.4%) | 234/245 (95.5%) |
| Yes | 78/879 (8.9%) | 67/634 (10.6%) | 11/245 (4.5%) |
| Iron deficiency (Ferritin<15ug/L) | | | |
| No | 664/879 (75.5%) | 419/634 (66.1%) | 245/245 (100.0%) |
| Yes | 215/879 (24.5%) | 215/634 (33.9%) | 0/245 (0.0%) |
| Inflammation (C-Reactive Protein>5mg/L) | | | |
| No | 822/879 (93.5%) | 577/634 (91.0%) | 245/245 (100.0%) |
| Yes | 57/879 (6.5%) | 57/634 (9.0%) | 0/245 (0.0%) |

US = United States, NHANES = National Health and Nutrition Examination Survey, BMI = Body Mass Index, SD = Standard Deviation.
Data are presented as unweighted mean (SD) for continuous measures, and unweighted n/total (%) for categorical measures based on participants with non-missing haemoglobin, ferritin, and C-reactive protein.
[1] Ferritin≥15ug/L and C-reactive protein≤5mg/L.

#### Table 3.7.3 US NHANES 2003-2004: Participant characteristics (Females) (12-17 years)

| | **Overall sample N=730** | **Excluded sample N=487** | **Reference sample[1] N=243** |
|---|---|---|---|
| Age (years) | 15.0 (1.8) | 15.2 (1.8) | 14.6 (1.7) |
| Sex | | | |
| Female | 730/730 (100.0%) | 487/487 (100.0%) | 243/243 (100.0%) |
| BMI (kg/m$^2$) | 23.6 (5.9) | 24.7 (6.6) | 21.5 (2.9) |
| Poverty | 229/706 (32.4%) | 153/470 (32.6%) | 76/236 (32.2%) |
| Race and Hispanic Origin | | | |
| Mexican American | 227/730 (31.1%) | 141/487 (29.0%) | 86/243 (35.4%) |
| Other Hispanic | 21/730 (2.9%) | 18/487 (3.7%) | 3/243 (1.2%) |
| Non-Hispanic White | 191/730 (26.2%) | 138/487 (28.3%) | 53/243 (21.8%) |
| Non-Hispanic Black | 258/730 (35.3%) | 170/487 (34.9%) | 88/243 (36.2%) |
| Other Race - Including Multi-Racial | 33/730 (4.5%) | 20/487 (4.1%) | 13/243 (5.3%) |
| Haemoglobin (g/L) | 133.7 (10.3) | 132.9 (10.6) | 135.3 (9.5) |
| Anaemia (Haemoglobin<120g/L) | | | |
| No | 669/730 (91.6%) | 439/487 (90.1%) | 230/243 (94.7%) |
| Yes | 61/730 (8.4%) | 48/487 (9.9%) | 13/243 (5.3%) |
| Iron deficiency (Ferritin<15ug/L) | | | |
| No | 602/730 (82.5%) | 359/487 (73.7%) | 243/243 (100.0%) |
| Yes | 128/730 (17.5%) | 128/487 (26.3%) | 0/243 (0.0%) |
| Inflammation (C-Reactive Protein>5mg/L) | | | |
| No | 660/730 (90.4%) | 417/487 (85.6%) | 243/243 (100.0%) |
| Yes | 70/730 (9.6%) | 70/487 (14.4%) | 0/243 (0.0%) |

US = United States, NHANES = National Health and Nutrition Examination Survey, BMI = Body Mass Index, SD = Standard Deviation.
Data are presented as unweighted mean (SD) for continuous measures, and unweighted n/total (%) for categorical measures based on participants with non-missing haemoglobin, ferritin, and C-reactive protein.
[1] Ferritin≥15ug/L and C-reactive protein≤5mg/L.

#### Table 3.7.4 US NHANES 2005-2006: Participant characteristics (Females) (12-17 years)

| | Overall sample N=732 | Excluded sample N=512 | Reference sample[1] N=220 |
|---|---|---|---|
| Age (years) | 15.1 (1.7) | 15.2 (1.7) | 14.7 (1.7) |
| Sex | | | |
| Female | 732/732 (100.0%) | 512/512 (100.0%) | 220/220 (100.0%) |
| BMI (kg/m$^2$) | 24.2 (6.2) | 25.4 (6.9) | 21.5 (2.9) |
| Poverty | 199/707 (28.1%) | 145/496 (29.2%) | 54/211 (25.6%) |
| Race and Hispanic Origin | | | |
| Mexican American | 255/732 (34.8%) | 176/512 (34.4%) | 79/220 (35.9%) |
| Other Hispanic | 19/732 (2.6%) | 14/512 (2.7%) | 5/220 (2.3%) |
| Non-Hispanic White | 192/732 (26.2%) | 143/512 (27.9%) | 49/220 (22.3%) |
| Non-Hispanic Black | 233/732 (31.8%) | 157/512 (30.7%) | 76/220 (34.5%) |
| Other Race - Including Multi-Racial | 33/732 (4.5%) | 22/512 (4.3%) | 11/220 (5.0%) |
| Haemoglobin (g/L) | 133.5 (10.2) | 132.9 (10.5) | 134.7 (9.3) |
| Anaemia (Haemoglobin<120g/L) | | | |
| No | 675/732 (92.2%) | 466/512 (91.0%) | 209/220 (95.0%) |
| Yes | 57/732 (7.8%) | 46/512 (9.0%) | 11/220 (5.0%) |
| Iron deficiency (Ferritin<15ug/L) | | | |
| No | 620/732 (84.7%) | 400/512 (78.1%) | 220/220 (100.0%) |
| Yes | 112/732 (15.3%) | 112/512 (21.9%) | 0/220 (0.0%) |
| Inflammation (C-Reactive Protein>5mg/L) | | | |
| No | 663/732 (90.6%) | 443/512 (86.5%) | 220/220 (100.0%) |
| Yes | 69/732 (9.4%) | 69/512 (13.5%) | 0/220 (0.0%) |

US = United States, NHANES = National Health and Nutrition Examination Survey, BMI = Body Mass Index, SD = Standard Deviation.
Data are presented as unweighted mean (SD) for continuous measures, and unweighted n/total (%) for categorical measures based on participants with non-missing haemoglobin, ferritin, and C-reactive protein.
[1] Ferritin≥15ug/L and C-reactive protein≤5mg/L.

#### Table 3.7.5 US NHANES 2007-2008: Participant characteristics (Females) (12-17 years)

| | Overall sample N=383 | Excluded sample N=264 | Reference sample[1] N=119 |
|---|---|---|---|
| Age (years) | 15.0 (1.7) | 15.2 (1.7) | 14.7 (1.6) |
| Sex | | | |
| Female | 383/383 (100.0%) | 264/264 (100.0%) | 119/119 (100.0%) |
| BMI (kg/m$^2$) | 24.1 (5.9) | 25.3 (6.4) | 21.4 (3.1) |
| Poverty | 107/359 (29.8%) | 72/248 (29.0%) | 35/111 (31.5%) |
| Race and Hispanic Origin | | | |
| Mexican American | 92/383 (24.0%) | 67/264 (25.4%) | 25/119 (21.0%) |
| Other Hispanic | 52/383 (13.6%) | 38/264 (14.4%) | 14/119 (11.8%) |
| Non-Hispanic White | 122/383 (31.9%) | 86/264 (32.6%) | 36/119 (30.3%) |
| Non-Hispanic Black | 103/383 (26.9%) | 64/264 (24.2%) | 39/119 (32.8%) |
| Other Race - Including Multi-Racial | 14/383 (3.7%) | 9/264 (3.4%) | 5/119 (4.2%) |
| Haemoglobin (g/L) | 131.8 (11.3) | 131.0 (11.4) | 133.4 (10.9) |
| Anaemia (Haemoglobin<120g/L) | | | |
| No | 346/383 (90.3%) | 232/264 (87.9%) | 114/119 (95.8%) |
| Yes | 37/383 (9.7%) | 32/264 (12.1%) | 5/119 (4.2%) |
| Iron deficiency (Ferritin<15ug/L) | | | |
| No | 323/383 (84.3%) | 204/264 (77.3%) | 119/119 (100.0%) |
| Yes | 60/383 (15.7%) | 60/264 (22.7%) | 0/119 (0.0%) |
| Inflammation (C-Reactive Protein>5mg/L) | | | |
| No | 350/383 (91.4%) | 231/264 (87.5%) | 119/119 (100.0%) |
| Yes | 33/383 (8.6%) | 33/264 (12.5%) | 0/119 (0.0%) |

US = United States, NHANES = National Health and Nutrition Examination Survey, BMI = Body Mass Index, SD = Standard Deviation.
Data are presented as unweighted mean (SD) for continuous measures, and unweighted n/total (%) for categorical measures based on participants with non-missing haemoglobin, ferritin, and C-reactive protein.
[1] Ferritin≥15ug/L and C-reactive protein≤5mg/L.

#### Table 3.7.6 US NHANES 2015-2016: Participant characteristics (Females) (12-17 years)

| | Overall sample N=420 | Excluded sample N=266 | Reference sample[1] N=154 |
|---|---|---|---|
| Age (years) | 15.1 (1.7) | 15.3 (1.8) | 14.8 (1.7) |
| Sex | | | |
| Female | 420/420 (100.0%) | 266/266 (100.0%) | 154/154 (100.0%) |
| BMI (kg/m$^2$) | 24.6 (5.9) | 26.0 (6.6) | 22.1 (3.3) |
| Poverty | 102/387 (26.4%) | 64/244 (26.2%) | 38/143 (26.6%) |
| Race and Hispanic Origin | | | |
| Mexican American | 105/420 (25.0%) | 68/266 (25.6%) | 37/154 (24.0%) |
| Other Hispanic | 53/420 (12.6%) | 34/266 (12.8%) | 19/154 (12.3%) |
| Non-Hispanic White | 113/420 (26.9%) | 71/266 (26.7%) | 42/154 (27.3%) |
| Non-Hispanic Black | 85/420 (20.2%) | 58/266 (21.8%) | 27/154 (17.5%) |
| Non-Hispanic Asian | 39/420 (9.3%) | 19/266 (7.1%) | 20/154 (13.0%) |
| Other Race - Including Multi-Racial | 25/420 (6.0%) | 16/266 (6.0%) | 9/154 (5.8%) |
| Haemoglobin (g/L) | 130.4 (11.1) | 128.0 (11.7) | 134.7 (8.5) |
| Anaemia (Haemoglobin<120g/L) | | | |
| No | 368/420 (87.6%) | 220/266 (82.7%) | 148/154 (96.1%) |
| Yes | 52/420 (12.4%) | 46/266 (17.3%) | 6/154 (3.9%) |
| Iron deficiency (Ferritin<15ug/L) | | | |
| No | 332/420 (79.0%) | 178/266 (66.9%) | 154/154 (100.0%) |
| Yes | 88/420 (21.0%) | 88/266 (33.1%) | 0/154 (0.0%) |
| Inflammation (C-Reactive Protein>5mg/L) | | | |
| No | 394/420 (93.8%) | 240/266 (90.2%) | 154/154 (100.0%) |
| Yes | 26/420 (6.2%) | 26/266 (9.8%) | 0/154 (0.0%) |

US = United States, NHANES = National Health and Nutrition Examination Survey, BMI = Body Mass Index, SD = Standard Deviation.
Data are presented as unweighted mean (SD) for continuous measures, and unweighted n/total (%) for categorical measures based on participants with non-missing haemoglobin, ferritin, and C-reactive protein.
[1] Ferritin≥15ug/L and C-reactive protein≤5mg/L.

#### Table 3.7.7 US NHANES 2017-2018: Participant characteristics (Females) (12-17 years)

| | Overall sample N=366 | Excluded sample N=257 | Reference sample[1] N=109 |
|---|---|---|---|
| Age (years) | 15.1 (1.8) | 15.2 (1.7) | 14.8 (1.9) |
| Sex | | | |
| Female | 366/366 (100.0%) | 257/257 (100.0%) | 109/109 (100.0%) |
| BMI (kg/m$^2$) | 24.8 (6.4) | 26.1 (6.9) | 21.7 (3.4) |
| Poverty | 81/324 (25.0%) | 64/229 (27.9%) | 17/95 (17.9%) |
| Race and Hispanic Origin | | | |
| Mexican American | 80/366 (21.9%) | 52/257 (20.2%) | 28/109 (25.7%) |
| Other Hispanic | 26/366 (7.1%) | 17/257 (6.6%) | 9/109 (8.3%) |
| Non-Hispanic White | 104/366 (28.4%) | 78/257 (30.4%) | 26/109 (23.9%) |
| Non-Hispanic Black | 86/366 (23.5%) | 63/257 (24.5%) | 23/109 (21.1%) |
| Non-Hispanic Asian | 43/366 (11.7%) | 27/257 (10.5%) | 16/109 (14.7%) |
| Other Race - Including Multi-Racial | 27/366 (7.4%) | 20/257 (7.8%) | 7/109 (6.4%) |
| Haemoglobin (g/L) | 130.2 (11.2) | 128.5 (12.1) | 134.2 (7.4) |
| Anaemia (Haemoglobin<120g/L) | | | |
| No | 315/366 (86.1%) | 209/257 (81.3%) | 106/109 (97.2%) |
| Yes | 51/366 (13.9%) | 48/257 (18.7%) | 3/109 (2.8%) |
| Iron deficiency (Ferritin<15ug/L) | | | |
| No | 287/366 (78.4%) | 178/257 (69.3%) | 109/109 (100.0%) |
| Yes | 79/366 (21.6%) | 79/257 (30.7%) | 0/109 (0.0%) |
| Inflammation (C-Reactive Protein>5mg/L) | | | |
| No | 339/366 (92.6%) | 230/257 (89.5%) | 109/109 (100.0%) |
| Yes | 27/366 (7.4%) | 27/257 (10.5%) | 0/109 (0.0%) |

US = United States, NHANES = National Health and Nutrition Examination Survey, BMI = Body Mass Index, SD = Standard Deviation.
Data are presented as unweighted mean (SD) for continuous measures, and unweighted n/total (%) for categorical measures based on participants with non-missing haemoglobin, ferritin, and C-reactive protein.
[1] Ferritin≥15ug/L and C-reactive protein≤5mg/L.

#### Table 3.7.8 AUS NHS and NNPAS 2011-2012: Participant characteristics (Females) (12-17 years)

| | Overall sample N=376 | Excluded sample N=203 | Reference sample[1] N=173 |
|---|---|---|---|
| Age (years) | 14.7 (1.7) | 15.0 (1.7) | 14.4 (1.6) |
| Sex | | | |
| Female | 376/376 (100.0%) | 203/203 (100.0%) | 173/173 (100.0%) |
| BMI (kg/m$^2$)* | 22.3 (4.4) | 22.9 (5.2) | 21.6 (3.1) |
| Equivalised income of household | | | |
| bottom quintile | 43/320 (13.4%) | 21/169 (12.4%) | 22/151 (14.6%) |
| 2nd quintile | 67/320 (20.9%) | 38/169 (22.5%) | 29/151 (19.2%) |
| 3rd quintile | 82/320 (25.6%) | 47/169 (27.8%) | 35/151 (23.2%) |
| 4th quintile | 81/320 (25.3%) | 39/169 (23.1%) | 42/151 (27.8%) |
| highest quintile | 47/320 (14.7%) | 24/169 (14.2%) | 23/151 (15.2%) |
| Haemoglobin (g/L) | 134.9 (8.7) | 133.7 (9.2) | 136.3 (7.9) |
| Anaemia (Haemoglobin<120g/L) | | | |
| No | 360/376 (95.7%) | Not displayed[†] | Not displayed[†] |
| Yes | 16/376 (4.3%) | Not displayed[†] | <10/173 (<5.8%) |
| Iron deficiency (Ferritin<15ug/L) | | | |
| No | 329/376 (87.5%) | Not displayed[†] | Not displayed[†] |
| Yes | 47/376 (12.5%) | Not displayed[†] | <10/173 (<5.8%) |
| Inflammation (C-Reactive Protein>5mg/L) | | | |
| No | 358/376 (95.2%) | Not displayed[†] | Not displayed[†] |
| Yes | 18/376 (4.8%) | Not displayed[†] | <10/173 (<5.8%) |

AUS = Australia, NHS = National Health Survey, NNPAS = National Nutrition and Physical Activity Survey, BMI = Body Mass Index, SD = Standard Deviation.
Data are presented as unweighted mean (SD) for continuous measures, and unweighted n/total (%) for categorical measures based on participants with non-missing haemoglobin, ferritin, and C-reactive protein. Data was combined from NHS and NNPAS 2011-2012.
* BMI is missing for 25 participants.
[†] Not presented due to Australian Bureau of Statistics requirements for output clearance.
[1] Ferritin≥15ug/L and C-reactive protein≤5mg/L.

##### Table 3.7.9 CHN CHNS 2009: Participant characteristics (Females) (12-17 years)

| | Overall sample N=187 | Excluded sample N=85 | Reference sample[1] N=102 |
|---|---|---|---|
| Age (years) | 13.9 (1.5) | 13.8 (1.4) | 14.0 (1.6) |
| Sex | | | |
| Female | 187/187 (100.0%) | 85/85 (100.0%) | 102/102 (100.0%) |
| BMI (kg/m$^2$) | 19.0 (2.8) | 18.7 (2.9) | 19.1 (2.8) |
| Nationality | | | |
| Han | 154/186 (82.8%) | 62/85 (72.9%) | 92/101 (91.1%) |
| Hui | 1/186 (0.5%) | 0/85 (0.0%) | 1/101 (1.0%) |
| Miao | 4/186 (2.2%) | 3/85 (3.5%) | 1/101 (1.0%) |
| Zhuang | 4/186 (2.2%) | 1/85 (1.2%) | 3/101 (3.0%) |
| Buyi | 12/186 (6.5%) | 12/85 (14.1%) | 0/101 (0.0%) |
| Man | 6/186 (3.2%) | 2/85 (2.4%) | 4/101 (4.0%) |
| Tujia | 4/186 (2.2%) | 4/85 (4.7%) | 0/101 (0.0%) |
| Other | 1/186 (0.5%) | 1/85 (1.2%) | 0/101 (0.0%) |
| Haemoglobin (g/L) | 134.7 (15.9) | 132.4 (16.3) | 136.7 (15.3) |
| Anaemia (Haemoglobin<120g/L) | | | |
| No | 172/187 (92.0%) | 76/85 (89.4%) | 96/102 (94.1%) |
| Yes | 15/187 (8.0%) | 9/85 (10.6%) | 6/102 (5.9%) |
| Iron deficiency (Ferritin<15ug/L) | | | |
| No | 157/187 (84.0%) | 55/85 (64.7%) | 102/102 (100.0%) |
| Yes | 30/187 (16.0%) | 30/85 (35.3%) | 0/102 (0.0%) |
| Inflammation (C-Reactive Protein>5mg/L) | | | |
| No | 179/187 (95.7%) | 77/85 (90.6%) | 102/102 (100.0%) |
| Yes | 8/187 (4.3%) | 8/85 (9.4%) | 0/102 (0.0%) |

CHN = China, CHNS = China Health and Nutrition Survey, BMI = Body Mass Index, SD = Standard Deviation.
Data are presented as unweighted mean (SD) for continuous measures, and unweighted n/total (%) for categorical measures based on participants with non-missing haemoglobin, ferritin, and C-reactive protein.
[1] Ferritin≥15ug/L and C-reactive protein≤5mg/L.

#### 3.8 Demographics Pregnant women

##### Table 3.8.1 NL Generation R: Participant characteristics (Trimester 1) (18-45 years)

| | **Overall sample N=5,054** | **Excluded sample N=4,279** | **Reference sample [1] N=775** |
|---|---|---|---|
| Age (years) | 29.8 (4.9) | 29.6 (5.0) | 30.8 (4.5) |
| BMI* (kg/m$^2$) | 24.5 (4.4) | 24.8 (4.6) | 22.8 (2.4) |
| Nationality | | | |
| Dutch | 2,469/4,844 (51.0%) | 1,992/4,070 (48.9%) | 477/774 (61.6%) |
| Other† | 2,375/4,844 (49.0%) | 2,078/4,070 (51.1%) | 297/774 (38.4%) |
| Haemoglobin (g/L) | 123.4 (9.2) | 123.4 (9.3) | 123.8 (8.7) |
| Anaemia (Haemoglobin<110g/L) | 372/5,054 (7.4%) | 327/4,279 (7.6%) | 45/775 (5.8%) |
| Iron deficiency (Ferritin<15ug/L)* | 366/4,785 (7.6%) | 366/4,010 (9.1%) | 0/775 (0%) |
| Inflammation (C-Reactive Protein>5mg/L)* | 2,136/4,737 (45.1%) | 2,136/3,962 (53.9%) | 0/775 (0%) |

NL = The Netherlands, BMI = Body Mass Index, SD = Standard Deviation.
Data are presented as mean (SD) for continuous measures, and n/total (%) for categorical measures based on participants with non-missing haemoglobin in first trimester of pregnancy.
* BMI, iron deficiency, and inflammation in the first trimester of pregnancy.
† Other includes Indonesian, Cape Verde, Moroccan, Dutch Antilles, Surinamese, Turkish, African, American, western, American, non-western, Asian, western, Asian, non-western, European, Oceania.
[1] Ferritin≥15 and ≤150ug/L and C-reactive protein≤5mg/L (both first trimester of pregnancy). If a measurement of one or more of the healthy criteria is missing, then the participant is excluded from the reference sample.

Table 3.8.2 NL Generation R: Participant characteristics (Trimester 2) (18-45 years)

| | **Overall sample N=2,194** | **Excluded sample N=2,083** | **Reference sample [1] N=111** |
|---|---|---|---|
| Age (years) | 29.9 (5.6) | 29.8 (5.6) | 31.5 (4.6) |
| BMI* (kg/m$^2$) | 24.7 (4.5) | 25.0 (4.7) | 22.7 (2.4) |
| Nationality | | | |
| Dutch | 886/2,029 (43.7%) | 813/1,918 (42.4%) | 73/111 (65.8%) |
| Other[†] | 1,143/2,029 (56.3%) | 1,105/1,918 (57.6%) | 38/111 (34.5%) |
| Haemoglobin (g/L) | 116.3 (9.7) | 116.2 (9.7) | 118.4 (7.6) |
| Anaemia (Haemoglobin<110g/L) | 549/2,149 (25.0%) | 535/2,083 (25.7%) | 14/111 (12.6%) |
| Iron deficiency (Ferritin<15ug/L)* | 45/658 (6.8%) | 45/547 (8.2%) | 0/111 (0%) |
| Inflammation (C-Reactive Protein>5mg/L)* | 276/649 (42.5%) | 276/538 (51.3%) | 0/111 (0%) |

NL = The Netherlands, BMI = Body Mass Index, SD = Standard Deviation.
Data are presented as mean (SD) for continuous measures, and n/total (%) for categorical measures based on participants with non-missing haemoglobin in second trimester of pregnancy.
* BMI, iron deficiency, and inflammation in the first trimester of pregnancy. Approx. 75% of the participants in their second trimester of pregnancy did not have a first trimester blood sample.
[†] Other includes Indonesian, Cape Verde, Moroccan, Dutch Antilles, Surinamese, Turkish, African, American-western, American-non-western, Asian-western, Asian- non-western, European, Oceania.
[1] Ferritin≥15 and ≤150ug/L and C-reactive protein≤5mg/L (both first trimester for pregnancy). If a measurement of one or more of the healthy criteria is missing, then the participant is excluded from the reference sample.

#### Table 3.8.3 NL Generation R: Participant characteristics (Trimester 3) (18-45 years)

No participant characteristics presented because of the reference sample size (N=1).

### 4. Detailed Individual Exclusions

#### 4.1 Detailed Individual Exclusions Adults (18-65 years)

##### Table 4.1.1 US NHANES 1999-2000: Participant characteristics of violations (18-65 years)

| | Overall sample | |
|---|---|---|
| | **Male N=1,673** | **Female N=1,944** |
| Reference sample[1] | | |
| No | 1,491/1,673 (89.1%) | 1,825/1,944 (93.9%) |
| Yes | 182/1,673 (10.9%) | 119/1,944 (6.1%) |
| VIOLATION: BMI<18.5 or >30 kg/m$^2$ | | |
| No | 1,197/1,673 (71.5%) | 1,195/1,944 (61.5%) |
| Yes | 463/1,673 (27.7%) | 742/1,944 (38.2%) |
| No data | 13/1,673 (0.8%) | 7/1,944 (0.4%) |
| VIOLATION: Doctor told you have diabetes | | |
| No | 1,563/1,673 (93.4%) | 1,840/1,944 (94.7%) |
| Yes | 108/1,673 (6.5%) | 104/1,944 (5.3%) |
| No data | 2/1,673 (0.1%) | 0/1,944 (0.0%) |
| VIOLATION: Ever told you had weak/failing kidneys? | | |
| No | 1,390/1,673 (83.1%) | 1,642/1,944 (84.5%) |
| Yes | 33/1,673 (2.0%) | 45/1,944 (2.3%) |
| No data | 5/1,673 (0.3%) | 0/1,944 (0.0%) |
| Missing | 245/1,673 (14.6%) | 257/1,944 (13.2%) |
| VIOLATION: Still have asthma | | |
| No | 12/1,673 (0.7%) | 15/1,944 (0.8%) |
| Yes | 22/1,673 (1.3%) | 24/1,944 (1.2%) |
| Missing | 1,639/1,673 (98.0%) | 1,905/1,944 (98.0%) |
| VIOLATION: Taking treatment for anemia/past 3 months | | |
| No | 1,662/1,673 (99.3%) | 1,829/1,944 (94.1%) |
| Yes | 11/1,673 (0.7%) | 114/1,944 (5.9%) |
| No data | 0/1,673 (0.0%) | 1/1,944 (0.1%) |
| VIOLATION: Doctor ever said you had arthritis | | |
| No | 1,230/1,673 (73.5%) | 1,375/1,944 (70.7%) |
| Yes | 197/1,673 (11.8%) | 311/1,944 (16.0%) |
| No data | 1/1,673 (0.1%) | 1/1,944 (0.1%) |
| Missing | 245/1,673 (14.6%) | 257/1,944 (13.2%) |
| VIOLATION: Ever told had congestive heart failure | | |
| No | 1,401/1,673 (83.7%) | 1,665/1,944 (85.6%) |
| Yes | 25/1,673 (1.5%) | 20/1,944 (1.0%) |
| No data | 2/1,673 (0.1%) | 2/1,944 (0.1%) |
| Missing | 245/1,673 (14.6%) | 257/1,944 (13.2%) |
| VIOLATION: Ever told you had coronary heart disease | | |
| No | 1,384/1,673 (82.7%) | 1,662/1,944 (85.5%) |
| Yes | 44/1,673 (2.6%) | 20/1,944 (1.0%) |
| No data | 0/1,673 (0.0%) | 5/1,944 (0.3%) |
| Missing | 245/1,673 (14.6%) | 257/1,944 (13.2%) |
| VIOLATION: Ever told you had angina/angina pectoris | | |
| No | 1,395/1,673 (83.4%) | 1,652/1,944 (85.0%) |
| Yes | 31/1,673 (1.9%) | 26/1,944 (1.3%) |
| No data | 2/1,673 (0.1%) | 9/1,944 (0.5%) |

| | Overall sample | |
|---|---|---|
| | Male N=1,673 | Female N=1,944 |
| Missing | 245/1,673 (14.6%) | 257/1,944 (13.2%) |
| VIOLATION: Ever told you had heart attack | | |
| No | 1,370/1,673 (81.9%) | 1,664/1,944 (85.6%) |
| Yes | 57/1,673 (3.4%) | 22/1,944 (1.1%) |
| No data | 1/1,673 (0.1%) | 1/1,944 (0.1%) |
| Missing | 245/1,673 (14.6%) | 257/1,944 (13.2%) |
| VIOLATION: Ever told you had a stroke | | |
| No | 1,401/1,673 (83.7%) | 1,666/1,944 (85.7%) |
| Yes | 27/1,673 (1.6%) | 21/1,944 (1.1%) |
| Missing | 245/1,673 (14.6%) | 257/1,944 (13.2%) |
| VIOLATION: Do you still have thyroid problem | | |
| No | 4/1,673 (0.2%) | 48/1,944 (2.5%) |
| Yes | 18/1,673 (1.1%) | 72/1,944 (3.7%) |
| No data | 0/1,673 (0.0%) | 7/1,944 (0.4%) |
| Missing | 1,651/1,673 (98.7%) | 1,817/1,944 (93.5%) |
| VIOLATION: Ever told you had emphysema | | |
| No | 1,416/1,673 (84.6%) | 1,676/1,944 (86.2%) |
| Yes | 10/1,673 (0.6%) | 11/1,944 (0.6%) |
| No data | 2/1,673 (0.1%) | 0/1,944 (0.0%) |
| Missing | 245/1,673 (14.6%) | 257/1,944 (13.2%) |
| VIOLATION: Ever told you had chronic bronchitis | | |
| No | 1,378/1,673 (82.4%) | 1,552/1,944 (79.8%) |
| Yes | 48/1,673 (2.9%) | 135/1,944 (6.9%) |
| No data | 2/1,673 (0.1%) | 0/1,944 (0.0%) |
| Missing | 245/1,673 (14.6%) | 257/1,944 (13.2%) |
| VIOLATION: Ever told you had any liver condition | | |
| No | 1,368/1,673 (81.8%) | 1,642/1,944 (84.5%) |
| Yes | 58/1,673 (3.5%) | 41/1,944 (2.1%) |
| No data | 2/1,673 (0.1%) | 4/1,944 (0.2%) |
| Missing | 245/1,673 (14.6%) | 257/1,944 (13.2%) |
| VIOLATION: Ever told you had cancer or malignancy | | |
| No | 1,387/1,673 (82.9%) | 1,611/1,944 (82.9%) |
| Yes | 40/1,673 (2.4%) | 75/1,944 (3.9%) |
| No data | 1/1,673 (0.1%) | 1/1,944 (0.1%) |
| Missing | 245/1,673 (14.6%) | 257/1,944 (13.2%) |
| VIOLATION: Overnight hospital patient in last year | | |
| No | 1,542/1,673 (92.2%) | 1,685/1,944 (86.7%) |
| Yes | 128/1,673 (7.7%) | 256/1,944 (13.2%) |
| No data | 3/1,673 (0.2%) | 3/1,944 (0.2%) |
| VIOLATION: SP have head cold or chest cold? | | |
| No | 1,264/1,673 (75.6%) | 1,398/1,944 (71.9%) |
| Yes | 333/1,673 (19.9%) | 438/1,944 (22.5%) |
| No data | 76/1,673 (4.5%) | 108/1,944 (5.6%) |
| VIOLATION: SP have stomach or intestinal illness? | | |
| No | 1,462/1,673 (87.4%) | 1,643/1,944 (84.5%) |
| Yes | 133/1,673 (7.9%) | 191/1,944 (9.8%) |
| No data | 78/1,673 (4.7%) | 110/1,944 (5.7%) |
| VIOLATION: SP have flu, pneumonia, ear infection? | | |
| No | 1,497/1,673 (89.5%) | 1,730/1,944 (89.0%) |

| | Overall sample | |
|---|---|---|
| | **Male N=1,673** | **Female N=1,944** |
| Yes | 97/1,673 (5.8%) | 103/1,944 (5.3%) |
| No data | 79/1,673 (4.7%) | 111/1,944 (5.7%) |
| VIOLATION: SP donated blood in past 12 months | | |
| No | 1,507/1,673 (90.1%) | 1,726/1,944 (88.8%) |
| Yes | 84/1,673 (5.0%) | 106/1,944 (5.5%) |
| No data | 82/1,673 (4.9%) | 112/1,944 (5.8%) |
| VIOLATION: Are you pregnant now? | | |
| No | 0/1,673 (0.0%) | 489/1,944 (25.2%) |
| Yes | 0/1,673 (0.0%) | 227/1,944 (11.7%) |
| No data | 0/1,673 (0.0%) | 610/1,944 (31.4%) |
| Missing | 1,673/1,673 (100.0%) | 618/1,944 (31.8%) |
| VIOLATION: Pregnancy status at exam | | |
| No | 0/1,673 (0.0%) | 1,379/1,944 (70.9%) |
| Yes | 0/1,673 (0.0%) | 265/1,944 (13.6%) |
| No data | 0/1,673 (0.0%) | 78/1,944 (4.0%) |
| Missing | 1,673/1,673 (100.0%) | 222/1,944 (11.4%) |
| VIOLATION: Urine Pregnancy Result | | |
| No | 0/1,673 (0.0%) | 1,441/1,944 (74.1%) |
| Yes | 0/1,673 (0.0%) | 263/1,944 (13.5%) |
| No data | 0/1,673 (0.0%) | 18/1,944 (0.9%) |
| Missing | 1,673/1,673 (100.0%) | 222/1,944 (11.4%) |
| VIOLATION: Taken prescription medicine, past month | | |
| No | 1,113/1,673 (66.5%) | 1,041/1,944 (53.5%) |
| Yes | 557/1,673 (33.3%) | 901/1,944 (46.3%) |
| No data | 3/1,673 (0.2%) | 2/1,944 (0.1%) |
| VIOLATION: Do you now smoke cigarettes? | | |
| No | 390/1,673 (23.3%) | 308/1,944 (15.8%) |
| Yes | 420/1,673 (25.1%) | 325/1,944 (16.7%) |
| Missing | 863/1,673 (51.6%) | 1,311/1,944 (67.4%) |
| VIOLATION: Smoked tobacco last 5 days? | | |
| No | 986/1,673 (58.9%) | 1,420/1,944 (73.0%) |
| Yes | 603/1,673 (36.0%) | 409/1,944 (21.0%) |
| No data | 84/1,673 (5.0%) | 115/1,944 (5.9%) |
| VIOLATION: Do you now smoke a pipe? | | |
| No | 120/1,673 (7.2%) | 3/1,944 (0.2%) |
| Yes | 4/1,673 (0.2%) | 0/1,944 (0.0%) |
| Missing | 1,549/1,673 (92.6%) | 1,941/1,944 (99.8%) |
| VIOLATION: Do you now smoke a cigar? | | |
| No | 82/1,673 (4.9%) | 4/1,944 (0.2%) |
| Yes | 64/1,673 (3.8%) | 6/1,944 (0.3%) |
| No data | 1/1,673 (0.1%) | 0/1,944 (0.0%) |
| Missing | 1,526/1,673 (91.2%) | 1,934/1,944 (99.5%) |
| VIOLATION: Do you now use snuff? | | |
| No | 48/1,673 (2.9%) | 3/1,944 (0.2%) |
| Yes | 44/1,673 (2.6%) | 3/1,944 (0.2%) |
| Missing | 1,581/1,673 (94.5%) | 1,938/1,944 (99.7%) |
| VIOLATION: Do you now chew tobacco? | | |
| No | 63/1,673 (3.8%) | 2/1,944 (0.1%) |
| Yes | 14/1,673 (0.8%) | 1/1,944 (0.1%) |

| | Overall sample | |
|---|---|---|
| | Male<br>N=1,673 | Female<br>N=1,944 |
| Missing | 1,596/1,673 (95.4%) | 1,941/1,944 (99.8%) |
| VIOLATION: Excessive alcohol intake | | |
| No | 307/1,673 (18.4%) | 590/1,944 (30.3%) |
| Yes | 116/1,673 (6.9%) | 67/1,944 (3.4%) |
| No data | 64/1,673 (3.8%) | 105/1,944 (5.4%) |
| Missing | 1,186/1,673 (70.9%) | 1,182/1,944 (60.8%) |
| VIOLATION: Ferritin<15ug/L or Ferritin>150/200ug/L (F/M) | | |
| No | 1,176/1,673 (70.3%) | 1,380/1,944 (71.0%) |
| Yes | 497/1,673 (29.7%) | 564/1,944 (29.0%) |
| VIOLATION: C-Reactive Protein>5mg/L | | |
| No | 1,421/1,673 (84.9%) | 1,273/1,944 (65.5%) |
| Yes | 252/1,673 (15.1%) | 671/1,944 (34.5%) |
| VIOLATION: eGFR<90 ml/min/1.73m$^2$ | | |
| No | 1,252/1,673 (74.8%) | 1,549/1,944 (79.7%) |
| Yes | 411/1,673 (24.6%) | 387/1,944 (19.9%) |
| No data | 10/1,673 (0.6%) | 8/1,944 (0.4%) |

US = United States, NHANES = National Health and Nutrition Examination Survey, BMI = Body Mass Index, SP = Survey Participant, F = Female, M = Male.
Data are presented as unweighted n/total (%) based on participants with non-missing haemoglobin, ferritin, and C-reactive protein.
No data: No answer/refused, Don't know; Missing: Not applicable, Not asked.
[1] Males: ferritin≥15 and ≤200ug/L and C-reactive protein≤5mg/L; females: ferritin≥15 and ≤150ug/L and C-reactive protein≤5mg/L.

#### Table 4.1.2 US NHANES 2001-2002: Participant characteristics of violations (18-65 years)

| | Overall sample | |
|---|---|---|
| | **Male N=1,973** | **Female N=2,160** |
| Reference sample[1] | | |
| No | 1,733/1,973 (87.8%) | 2,052/2,160 (95.0%) |
| Yes | 240/1,973 (12.2%) | 108/2,160 (5.0%) |
| VIOLATION: BMI<18.5 or >30 kg/m² | | |
| No | 1,390/1,973 (70.5%) | 1,327/2,160 (61.4%) |
| Yes | 526/1,973 (26.7%) | 768/2,160 (35.6%) |
| No data | 57/1,973 (2.9%) | 65/2,160 (3.0%) |
| VIOLATION: Doctor told you have diabetes | | |
| No | 1,840/1,973 (93.3%) | 2,031/2,160 (94.0%) |
| Yes | 132/1,973 (6.7%) | 129/2,160 (6.0%) |
| No data | 1/1,973 (0.1%) | 0/2,160 (0.0%) |
| VIOLATION: Ever told you had weak/failing kidneys? | | |
| No | 1,659/1,973 (84.1%) | 1,881/2,160 (87.1%) |
| Yes | 33/1,973 (1.7%) | 31/2,160 (1.4%) |
| No data | 2/1,973 (0.1%) | 4/2,160 (0.2%) |
| Missing | 279/1,973 (14.1%) | 244/2,160 (11.3%) |
| VIOLATION: Still have asthma | | |
| No | 83/1,973 (4.2%) | 97/2,160 (4.5%) |
| Yes | 104/1,973 (5.3%) | 169/2,160 (7.8%) |
| No data | 4/1,973 (0.2%) | 4/2,160 (0.2%) |
| Missing | 1,782/1,973 (90.3%) | 1,890/2,160 (87.5%) |
| VIOLATION: Taking treatment for anemia/past 3 months | | |
| No | 1,958/1,973 (99.2%) | 2,040/2,160 (94.4%) |
| Yes | 15/1,973 (0.8%) | 119/2,160 (5.5%) |
| No data | 0/1,973 (0.0%) | 1/2,160 (0.0%) |
| VIOLATION: Doctor ever said you had arthritis | | |
| No | 1,445/1,973 (73.2%) | 1,570/2,160 (72.7%) |
| Yes | 248/1,973 (12.6%) | 345/2,160 (16.0%) |
| No data | 1/1,973 (0.1%) | 1/2,160 (0.0%) |
| Missing | 279/1,973 (14.1%) | 244/2,160 (11.3%) |
| VIOLATION: Ever told had congestive heart failure | | |
| No | 1,659/1,973 (84.1%) | 1,896/2,160 (87.8%) |
| Yes | 32/1,973 (1.6%) | 19/2,160 (0.9%) |
| No data | 3/1,973 (0.2%) | 1/2,160 (0.0%) |
| Missing | 279/1,973 (14.1%) | 244/2,160 (11.3%) |
| VIOLATION: Ever told you had coronary heart disease | | |
| No | 1,638/1,973 (83.0%) | 1,890/2,160 (87.5%) |
| Yes | 52/1,973 (2.6%) | 26/2,160 (1.2%) |
| No data | 4/1,973 (0.2%) | 0/2,160 (0.0%) |
| Missing | 279/1,973 (14.1%) | 244/2,160 (11.3%) |
| VIOLATION: Ever told you had angina/angina pectoris | | |
| No | 1,657/1,973 (84.0%) | 1,877/2,160 (86.9%) |
| Yes | 34/1,973 (1.7%) | 35/2,160 (1.6%) |
| No data | 3/1,973 (0.2%) | 4/2,160 (0.2%) |
| Missing | 279/1,973 (14.1%) | 244/2,160 (11.3%) |
| VIOLATION: Ever told you had heart attack | | |
| No | 1,645/1,973 (83.4%) | 1,892/2,160 (87.6%) |
| Yes | 48/1,973 (2.4%) | 24/2,160 (1.1%) |

| | Overall sample | |
|---|---|---|
| | Male<br>N=1,973 | Female<br>N=2,160 |
| No data | 1/1,973 (0.1%) | 0/2,160 (0.0%) |
| Missing | 279/1,973 (14.1%) | 244/2,160 (11.3%) |
| VIOLATION: Ever told you had a stroke | | |
| No | 1,672/1,973 (84.7%) | 1,886/2,160 (87.3%) |
| Yes | 21/1,973 (1.1%) | 29/2,160 (1.3%) |
| No data | 1/1,973 (0.1%) | 1/2,160 (0.0%) |
| Missing | 279/1,973 (14.1%) | 244/2,160 (11.3%) |
| VIOLATION: Do you still have thyroid problem | | |
| No | 10/1,973 (0.5%) | 56/2,160 (2.6%) |
| Yes | 21/1,973 (1.1%) | 104/2,160 (4.8%) |
| No data | 2/1,973 (0.1%) | 6/2,160 (0.3%) |
| Missing | 1,940/1,973 (98.3%) | 1,994/2,160 (92.3%) |
| VIOLATION: Ever told you had emphysema | | |
| No | 1,677/1,973 (85.0%) | 1,904/2,160 (88.1%) |
| Yes | 17/1,973 (0.9%) | 12/2,160 (0.6%) |
| Missing | 279/1,973 (14.1%) | 244/2,160 (11.3%) |
| VIOLATION: Ever told you had chronic bronchitis | | |
| No | 1,626/1,973 (82.4%) | 1,788/2,160 (82.8%) |
| Yes | 62/1,973 (3.1%) | 122/2,160 (5.6%) |
| No data | 6/1,973 (0.3%) | 6/2,160 (0.3%) |
| Missing | 279/1,973 (14.1%) | 244/2,160 (11.3%) |
| VIOLATION: Ever told you had any liver condition | | |
| No | 1,621/1,973 (82.2%) | 1,876/2,160 (86.9%) |
| Yes | 70/1,973 (3.5%) | 39/2,160 (1.8%) |
| No data | 3/1,973 (0.2%) | 1/2,160 (0.0%) |
| Missing | 279/1,973 (14.1%) | 244/2,160 (11.3%) |
| VIOLATION: Ever told you had cancer or malignancy | | |
| No | 1,631/1,973 (82.7%) | 1,802/2,160 (83.4%) |
| Yes | 63/1,973 (3.2%) | 110/2,160 (5.1%) |
| No data | 0/1,973 (0.0%) | 4/2,160 (0.2%) |
| Missing | 279/1,973 (14.1%) | 244/2,160 (11.3%) |
| VIOLATION: Overnight hospital patient in last year | | |
| No | 1,839/1,973 (93.2%) | 1,846/2,160 (85.5%) |
| Yes | 134/1,973 (6.8%) | 314/2,160 (14.5%) |
| VIOLATION: SP have head cold or chest cold? | | |
| No | 1,464/1,973 (74.2%) | 1,510/2,160 (69.9%) |
| Yes | 401/1,973 (20.3%) | 466/2,160 (21.6%) |
| No data | 108/1,973 (5.5%) | 184/2,160 (8.5%) |
| VIOLATION: SP have stomach or intestinal illness? | | |
| No | 1,704/1,973 (86.4%) | 1,773/2,160 (82.1%) |
| Yes | 161/1,973 (8.2%) | 203/2,160 (9.4%) |
| No data | 108/1,973 (5.5%) | 184/2,160 (8.5%) |
| VIOLATION: SP have flu, pneumonia, ear infection? | | |
| No | 1,791/1,973 (90.8%) | 1,853/2,160 (85.8%) |
| Yes | 73/1,973 (3.7%) | 121/2,160 (5.6%) |
| No data | 109/1,973 (5.5%) | 186/2,160 (8.6%) |
| VIOLATION: SP donated blood in past 12 months | | |
| No | 1,750/1,973 (88.7%) | 1,865/2,160 (86.3%) |
| Yes | 114/1,973 (5.8%) | 111/2,160 (5.1%) |

| | Overall sample | |
|---|---|---|
| | Male<br>N=1,973 | Female<br>N=2,160 |
| No data | 109/1,973 (5.5%) | 184/2,160 (8.5%) |
| VIOLATION: Are you pregnant now? | | |
| No | 0/1,973 (0.0%) | 625/2,160 (28.9%) |
| Yes | 0/1,973 (0.0%) | 243/2,160 (11.3%) |
| No data | 0/1,973 (0.0%) | 1,118/2,160 (51.8%) |
| Missing | 1,973/1,973 (100.0%) | 174/2,160 (8.1%) |
| VIOLATION: Pregnancy status at exam | | |
| No | 0/1,973 (0.0%) | 1,556/2,160 (72.0%) |
| Yes | 0/1,973 (0.0%) | 305/2,160 (14.1%) |
| No data | 0/1,973 (0.0%) | 84/2,160 (3.9%) |
| Missing | 1,973/1,973 (100.0%) | 215/2,160 (10.0%) |
| VIOLATION: Urine Pregnancy Result | | |
| No | 0/1,973 (0.0%) | 1,621/2,160 (75.0%) |
| Yes | 0/1,973 (0.0%) | 301/2,160 (13.9%) |
| No data | 0/1,973 (0.0%) | 23/2,160 (1.1%) |
| Missing | 1,973/1,973 (100.0%) | 215/2,160 (10.0%) |
| VIOLATION: Taken prescription medicine, past month | | |
| No | 1,260/1,973 (63.9%) | 1,103/2,160 (51.1%) |
| Yes | 713/1,973 (36.1%) | 1,057/2,160 (48.9%) |
| VIOLATION: Do you now smoke cigarettes? | | |
| No | 447/1,973 (22.7%) | 343/2,160 (15.9%) |
| Yes | 539/1,973 (27.3%) | 415/2,160 (19.2%) |
| Missing | 987/1,973 (50.0%) | 1,402/2,160 (64.9%) |
| VIOLATION: Smoked tobacco last 5 days? | | |
| No | 1,160/1,973 (58.8%) | 1,511/2,160 (70.0%) |
| Yes | 703/1,973 (35.6%) | 461/2,160 (21.3%) |
| No data | 110/1,973 (5.6%) | 188/2,160 (8.7%) |
| VIOLATION: Do you now smoke a pipe? | | |
| No | 122/1,973 (6.2%) | 6/2,160 (0.3%) |
| Yes | 16/1,973 (0.8%) | 1/2,160 (0.0%) |
| Missing | 1,835/1,973 (93.0%) | 2,153/2,160 (99.7%) |
| VIOLATION: Do you now smoke cigars? | | |
| No | 87/1,973 (4.4%) | 9/2,160 (0.4%) |
| Yes | 67/1,973 (3.4%) | 4/2,160 (0.2%) |
| Missing | 1,819/1,973 (92.2%) | 2,147/2,160 (99.4%) |
| VIOLATION: Do you now use snuff? | | |
| No | 75/1,973 (3.8%) | 2/2,160 (0.1%) |
| Yes | 43/1,973 (2.2%) | 0/2,160 (0.0%) |
| Missing | 1,855/1,973 (94.0%) | 2,158/2,160 (99.9%) |
| VIOLATION: Do you now chew tobacco? | | |
| No | 78/1,973 (4.0%) | 5/2,160 (0.2%) |
| Yes | 25/1,973 (1.3%) | 2/2,160 (0.1%) |
| Missing | 1,870/1,973 (94.8%) | 2,153/2,160 (99.7%) |
| VIOLATION: Excessive alcohol intake | | |
| No | 368/1,973 (18.7%) | 661/2,160 (30.6%) |
| Yes | 144/1,973 (7.3%) | 78/2,160 (3.6%) |
| No data | 88/1,973 (4.5%) | 172/2,160 (8.0%) |
| Missing | 1,373/1,973 (69.6%) | 1,249/2,160 (57.8%) |
| VIOLATION: Ferritin<15ug/L or Ferritin>150/200ug/L (F/M) | | |

| | Overall sample | |
|---|---|---|
| | Male<br>N=1,973 | Female<br>N=2,160 |
| No | 1,370/1,973 (69.4%) | 1,486/2,160 (68.8%) |
| Yes | 603/1,973 (30.6%) | 674/2,160 (31.2%) |
| VIOLATION: C-Reactive Protein>5mg/L | | |
| No | 1,678/1,973 (85.0%) | 1,475/2,160 (68.3%) |
| Yes | 295/1,973 (15.0%) | 685/2,160 (31.7%) |
| VIOLATION: eGFR<90 ml/min/1.73m[2] | | |
| No | 1,419/1,973 (71.9%) | 1,726/2,160 (79.9%) |
| Yes | 548/1,973 (27.8%) | 414/2,160 (19.2%) |
| No data | 6/1,973 (0.3%) | 20/2,160 (0.9%) |

US = United States, NHANES = National Health and Nutrition Examination Survey, BMI = Body Mass Index, SP = Survey Participant, F = Female, M = Male.
Data are presented as unweighted n/total (%) based on participants with non-missing haemoglobin, ferritin, and C-reactive protein.
No data: No answer/refused, Don't know; Missing: Not applicable, Not asked.
[1] Males: ferritin≥15 and ≤200ug/L and C-reactive protein≤5mg/L; females: ferritin≥15 and ≤150ug/L and C-reactive protein≤5mg/L.

#### Table 4.1.3 US NHANES 2003-2004: Participant characteristics of violations (Females) (18-65 years)

| | Overall sample Female N=1,455 |
|---|---|
| Reference sample[1] | |
| No | 1,337/1,455 (91.9%) |
| Yes | 118/1,455 (8.1%) |
| VIOLATION: BMI<18.5 or >30 kg/m$^2$ | |
| No | 903/1,455 (62.1%) |
| Yes | 530/1,455 (36.4%) |
| No data | 22/1,455 (1.5%) |
| VIOLATION: Doctor told you have diabetes | |
| No | 1,417/1,455 (97.4%) |
| Yes | 37/1,455 (2.5%) |
| No data | 1/1,455 (0.1%) |
| VIOLATION: Ever told you had weak/failing kidneys? | |
| No | 1,166/1,455 (80.1%) |
| Yes | 16/1,455 (1.1%) |
| No data | 5/1,455 (0.3%) |
| Missing | 268/1,455 (18.4%) |
| VIOLATION: Still have asthma | |
| No | 79/1,455 (5.4%) |
| Yes | 143/1,455 (9.8%) |
| No data | 4/1,455 (0.3%) |
| Missing | 1,229/1,455 (84.5%) |
| VIOLATION: Taking treatment for anemia/past 3 months | |
| No | 1,378/1,455 (94.7%) |
| Yes | 76/1,455 (5.2%) |
| No data | 1/1,455 (0.1%) |
| VIOLATION: Doctor ever said you had arthritis | |
| No | 1,048/1,455 (72.0%) |
| Yes | 136/1,455 (9.3%) |
| No data | 3/1,455 (0.2%) |
| Missing | 268/1,455 (18.4%) |
| VIOLATION: Ever told had congestive heart failure | |
| No | 1,180/1,455 (81.1%) |
| Yes | 5/1,455 (0.3%) |
| No data | 2/1,455 (0.1%) |
| Missing | 268/1,455 (18.4%) |
| VIOLATION: Ever told you had coronary heart disease | |
| No | 1,180/1,455 (81.1%) |
| Yes | 5/1,455 (0.3%) |
| No data | 2/1,455 (0.1%) |
| Missing | 268/1,455 (18.4%) |
| VIOLATION: Ever told you had angina/angina pectoris | |
| No | 1,176/1,455 (80.8%) |
| Yes | 9/1,455 (0.6%) |
| No data | 2/1,455 (0.1%) |
| Missing | 268/1,455 (18.4%) |
| VIOLATION: Ever told you had heart attack | |
| No | 1,178/1,455 (81.0%) |

| | Overall sample<br>Female<br>N=1,455 |
|---|---|
| Yes | 8/1,455 (0.5%) |
| No data | 1/1,455 (0.1%) |
| Missing | 268/1,455 (18.4%) |
| VIOLATION: Ever told you had a stroke | |
| No | 1,175/1,455 (80.8%) |
| Yes | 11/1,455 (0.8%) |
| No data | 1/1,455 (0.1%) |
| Missing | 268/1,455 (18.4%) |
| VIOLATION: Do you still have thyroid problem | |
| No | 21/1,455 (1.4%) |
| Yes | 77/1,455 (5.3%) |
| No data | 6/1,455 (0.4%) |
| Missing | 1,351/1,455 (92.9%) |
| VIOLATION: Ever told you had emphysema | |
| No | 1,179/1,455 (81.0%) |
| Yes | 7/1,455 (0.5%) |
| No data | 1/1,455 (0.1%) |
| Missing | 268/1,455 (18.4%) |
| VIOLATION: Ever told you had chronic bronchitis | |
| No | 1,109/1,455 (76.2%) |
| Yes | 76/1,455 (5.2%) |
| No data | 2/1,455 (0.1%) |
| Missing | 268/1,455 (18.4%) |
| VIOLATION: Ever told you had any liver condition | |
| No | 1,163/1,455 (79.9%) |
| Yes | 21/1,455 (1.4%) |
| No data | 3/1,455 (0.2%) |
| Missing | 268/1,455 (18.4%) |
| VIOLATION: Ever told you had cancer or malignancy | |
| No | 1,141/1,455 (78.4%) |
| Yes | 41/1,455 (2.8%) |
| No data | 5/1,455 (0.3%) |
| Missing | 268/1,455 (18.4%) |
| VIOLATION: Overnight hospital patient in last year | |
| No | 1,269/1,455 (87.2%) |
| Yes | 186/1,455 (12.8%) |
| VIOLATION: SP have head cold or chest cold? | |
| No | 1,040/1,455 (71.5%) |
| Yes | 272/1,455 (18.7%) |
| No data | 143/1,455 (9.8%) |
| VIOLATION: SP have stomach or intestinal illness? | |
| No | 1,188/1,455 (81.6%) |
| Yes | 124/1,455 (8.5%) |
| No data | 143/1,455 (9.8%) |
| VIOLATION: SP have flu, pneumonia, ear infection? | |
| No | 1,237/1,455 (85.0%) |
| Yes | 73/1,455 (5.0%) |
| No data | 145/1,455 (10.0%) |
| VIOLATION: SP donated blood in past 12 months | |

| | Overall sample<br>Female<br>N=1,455 |
|---|---|
| No | 1,229/1,455 (84.5%) |
| Yes | 81/1,455 (5.6%) |
| No data | 145/1,455 (10.0%) |
| VIOLATION: Are you pregnant now? | |
| No | 645/1,455 (44.3%) |
| Yes | 198/1,455 (13.6%) |
| No data | 612/1,455 (42.1%) |
| VIOLATION: Pregnancy status at exam | |
| No | 1,188/1,455 (81.6%) |
| Yes | 230/1,455 (15.8%) |
| No data | 37/1,455 (2.5%) |
| VIOLATION: Urine Pregnancy Result | |
| No | 1,217/1,455 (83.6%) |
| Yes | 228/1,455 (15.7%) |
| No data | 10/1,455 (0.7%) |
| VIOLATION: Taken prescription medicine, past month | |
| No | 831/1,455 (57.1%) |
| Yes | 622/1,455 (42.7%) |
| No data | 2/1,455 (0.1%) |
| VIOLATION: Do you now smoke cigarettes? | |
| No | 170/1,455 (11.7%) |
| Yes | 291/1,455 (20.0%) |
| Missing | 994/1,455 (68.3%) |
| VIOLATION: Smoked tobacco last 5 days? | |
| No | 974/1,455 (66.9%) |
| Yes | 335/1,455 (23.0%) |
| No data | 146/1,455 (10.0%) |
| VIOLATION: Do you now smoke a pipe? | |
| No | 3/1,455 (0.2%) |
| Missing | 1,452/1,455 (99.8%) |
| VIOLATION: Do you now smoke cigars? | |
| No | 3/1,455 (0.2%) |
| Yes | 9/1,455 (0.6%) |
| Missing | 1,443/1,455 (99.2%) |
| VIOLATION: Do you now use snuff? | |
| No | 3/1,455 (0.2%) |
| Yes | 1/1,455 (0.1%) |
| Missing | 1,451/1,455 (99.7%) |
| VIOLATION: Do you now chew tobacco? | |
| Missing | 1,455/1,455 (100.0%) |
| VIOLATION: Excessive alcohol intake | |
| No | 354/1,455 (24.3%) |
| Yes | 51/1,455 (3.5%) |
| No data | 131/1,455 (9.0%) |
| Missing | 919/1,455 (63.2%) |
| VIOLATION: Ferritin<15ug/L or Ferritin>150ug/L | |
| No | 1,120/1,455 (77.0%) |
| Yes | 335/1,455 (23.0%) |
| VIOLATION: C-Reactive Protein>5mg/L | |

| | Overall sample Female N=1,455 |
|---|---|
| No | 986/1,455 (67.8%) |
| Yes | 469/1,455 (32.2%) |
| VIOLATION: eGFR<90 ml/min/1.73m$^2$ | |
| No | 1,315/1,455 (90.4%) |
| Yes | 131/1,455 (9.0%) |
| No data | 9/1,455 (0.6%) |

US = United States, NHANES = National Health and Nutrition Examination Survey, BMI = Body Mass Index, SP = Survey Participant.
Data are presented as unweighted n/total (%) based on participants with non-missing haemoglobin, ferritin, and C-reactive protein.  No ferritin was measured in participants more than 49 years. No ferritin was collected in males.
No data: No answer/refused, Don't know; Missing: Not applicable, Not asked.
[1] Ferritin≥15 and ≤150ug/L and C-reactive protein≤5mg/L.

Table 4.1.4 US NHANES 2005-2006: Participant characteristics of violations (Females) (18-65 years)

| | Overall sample Female N=1,606 |
|---|---|
| Reference sample[1] | |
| No | 1,473/1,606 (91.7%) |
| Yes | 133/1,606 (8.3%) |
| VIOLATION: BMI<18.5 or >30 kg/m$^2$ | |
| No | 984/1,606 (61.3%) |
| Yes | 611/1,606 (38.0%) |
| No data | 11/1,606 (0.7%) |
| VIOLATION: Doctor told you have diabetes | |
| No | 1,555/1,606 (96.8%) |
| Yes | 50/1,606 (3.1%) |
| No data | 1/1,606 (0.1%) |
| VIOLATION: Ever told you had weak/failing kidneys? | |
| No | 1,313/1,606 (81.8%) |
| Yes | 29/1,606 (1.8%) |
| No data | 5/1,606 (0.3%) |
| Missing | 259/1,606 (16.1%) |
| VIOLATION: Still have asthma | |
| No | 86/1,606 (5.4%) |
| Yes | 153/1,606 (9.5%) |
| No data | 4/1,606 (0.2%) |
| Missing | 1,363/1,606 (84.9%) |
| VIOLATION: Taking treatment for anemia/past 3 months | |
| No | 1,515/1,606 (94.3%) |
| Yes | 90/1,606 (5.6%) |
| No data | 1/1,606 (0.1%) |
| VIOLATION: Doctor ever said you had arthritis | |
| No | 1,202/1,606 (74.8%) |
| Yes | 143/1,606 (8.9%) |
| No data | 2/1,606 (0.1%) |
| Missing | 259/1,606 (16.1%) |
| VIOLATION: Ever told had congestive heart failure | |
| No | 1,337/1,606 (83.3%) |
| Yes | 8/1,606 (0.5%) |
| No data | 2/1,606 (0.1%) |
| Missing | 259/1,606 (16.1%) |
| VIOLATION: Ever told you had coronary heart disease | |
| No | 1,338/1,606 (83.3%) |
| Yes | 7/1,606 (0.4%) |
| No data | 2/1,606 (0.1%) |
| Missing | 259/1,606 (16.1%) |
| VIOLATION: Ever told you had angina/angina pectoris | |
| No | 1,336/1,606 (83.2%) |
| Yes | 8/1,606 (0.5%) |
| No data | 3/1,606 (0.2%) |
| Missing | 259/1,606 (16.1%) |
| VIOLATION: Ever told you had heart attack | |
| No | 1,339/1,606 (83.4%) |

| | Overall sample<br>Female<br>N=1,606 |
|---|---|
| Yes | 7/1,606 (0.4%) |
| No data | 1/1,606 (0.1%) |
| Missing | 259/1,606 (16.1%) |
| VIOLATION: Ever told you had a stroke | |
| No | 1,331/1,606 (82.9%) |
| Yes | 16/1,606 (1.0%) |
| Missing | 259/1,606 (16.1%) |
| VIOLATION: Do you still have thyroid problem | |
| No | 23/1,606 (1.4%) |
| Yes | 71/1,606 (4.4%) |
| No data | 4/1,606 (0.2%) |
| Missing | 1,508/1,606 (93.9%) |
| VIOLATION: Ever told you had emphysema | |
| No | 1,342/1,606 (83.6%) |
| Yes | 5/1,606 (0.3%) |
| Missing | 259/1,606 (16.1%) |
| VIOLATION: Ever told you had chronic bronchitis | |
| No | 1,264/1,606 (78.7%) |
| Yes | 80/1,606 (5.0%) |
| No data | 3/1,606 (0.2%) |
| Missing | 259/1,606 (16.1%) |
| VIOLATION: Ever told you had any liver condition | |
| No | 1,316/1,606 (81.9%) |
| Yes | 30/1,606 (1.9%) |
| No data | 1/1,606 (0.1%) |
| Missing | 259/1,606 (16.1%) |
| VIOLATION: Ever told you had cancer or malignancy | |
| No | 1,296/1,606 (80.7%) |
| Yes | 51/1,606 (3.2%) |
| Missing | 259/1,606 (16.1%) |
| VIOLATION: Overnight hospital patient in last year | |
| No | 1,401/1,606 (87.2%) |
| Yes | 205/1,606 (12.8%) |
| VIOLATION: SP have head cold or chest cold? | |
| No | 1,170/1,606 (72.9%) |
| Yes | 309/1,606 (19.2%) |
| No data | 127/1,606 (7.9%) |
| VIOLATION: SP have stomach or intestinal illness? | |
| No | 1,321/1,606 (82.3%) |
| Yes | 160/1,606 (10.0%) |
| No data | 125/1,606 (7.8%) |
| VIOLATION: SP have flu, pneumonia, ear infection? | |
| No | 1,405/1,606 (87.5%) |
| Yes | 76/1,606 (4.7%) |
| No data | 125/1,606 (7.8%) |
| VIOLATION: SP donated blood in past 12 months | |
| No | 1,404/1,606 (87.4%) |
| Yes | 77/1,606 (4.8%) |
| No data | 125/1,606 (7.8%) |

| | Overall sample<br>Female<br>N=1,606 |
|---|---|
| VIOLATION: Are you pregnant now? | |
| No | 711/1,606 (44.3%) |
| Yes | 299/1,606 (18.6%) |
| No data | 596/1,606 (37.1%) |
| VIOLATION: Pregnancy status at exam | |
| No | 1,229/1,606 (76.5%) |
| Yes | 334/1,606 (20.8%) |
| No data | 43/1,606 (2.7%) |
| VIOLATION: Urine Pregnancy Result | |
| No | 1,259/1,606 (78.4%) |
| Yes | 331/1,606 (20.6%) |
| No data | 16/1,606 (1.0%) |
| VIOLATION: Taken prescription medicine, past month | |
| No | 989/1,606 (61.6%) |
| Yes | 616/1,606 (38.4%) |
| No data | 1/1,606 (0.1%) |
| VIOLATION: Do you now smoke cigarettes? | |
| No | 214/1,606 (13.3%) |
| Yes | 270/1,606 (16.8%) |
| Missing | 1,122/1,606 (69.9%) |
| VIOLATION: Smoked tobacco last 5 days? | |
| No | 1,152/1,606 (71.7%) |
| Yes | 321/1,606 (20.0%) |
| No data | 133/1,606 (8.3%) |
| VIOLATION: Excessive alcohol intake | |
| No | 423/1,606 (26.3%) |
| Yes | 54/1,606 (3.4%) |
| No data | 116/1,606 (7.2%) |
| Missing | 1,013/1,606 (63.1%) |
| VIOLATION: Ferritin<15ug/L or Ferritin>150ug/L | |
| No | 1,226/1,606 (76.3%) |
| Yes | 380/1,606 (23.7%) |
| VIOLATION: C-Reactive Protein>5mg/L | |
| No | 1,074/1,606 (66.9%) |
| Yes | 532/1,606 (33.1%) |
| VIOLATION: eGFR<90 ml/min/1.73m[2] | |
| No | 1,414/1,606 (88.0%) |
| Yes | 185/1,606 (11.5%) |
| No data | 7/1,606 (0.4%) |

US = United States, NHANES = National Health and Nutrition Examination Survey, BMI = Body Mass Index, SP = Survey Participant.
Data are presented as unweighted n/total (%) based on participants with non-missing haemoglobin, ferritin, and C-reactive protein. No ferritin was measured in participants more than 49 years. No ferritin was collected in males.
No data: No answer/refused, Don't know; Missing: Not applicable, Not asked.
[1] Ferritin≥15 and ≤150ug/L and C-reactive protein≤5mg/L.

#### Table 4.1.5 US NHANES 2007-2008: Participant characteristics of violations (Females) (18-65 years)

| | Overall sample Female N=1,434 |
|---|---|
| Reference sample[1] | |
| No | 1,297/1,434 (90.4%) |
| Yes | 137/1,434 (9.6%) |
| VIOLATION: BMI<18.5 or >30 kg/m$^2$ | |
| No | 871/1,434 (60.7%) |
| Yes | 542/1,434 (37.8%) |
| No data | 21/1,434 (1.5%) |
| VIOLATION: Doctor told you have diabetes | |
| No | 1,376/1,434 (96.0%) |
| Yes | 57/1,434 (4.0%) |
| No data | 1/1,434 (0.1%) |
| VIOLATION: Ever told you had weak/failing kidneys? | |
| No | 1,294/1,434 (90.2%) |
| Yes | 29/1,434 (2.0%) |
| Missing | 111/1,434 (7.7%) |
| VIOLATION: Still have asthma | |
| No | 82/1,434 (5.7%) |
| Yes | 128/1,434 (8.9%) |
| No data | 7/1,434 (0.5%) |
| Missing | 1,217/1,434 (84.9%) |
| VIOLATION: Taking treatment for anemia/past 3 months | |
| No | 1,355/1,434 (94.5%) |
| Yes | 79/1,434 (5.5%) |
| VIOLATION: Doctor ever said you had arthritis | |
| No | 1,139/1,434 (79.4%) |
| Yes | 183/1,434 (12.8%) |
| No data | 1/1,434 (0.1%) |
| Missing | 111/1,434 (7.7%) |
| VIOLATION: Ever told had congestive heart failure | |
| No | 1,313/1,434 (91.6%) |
| Yes | 9/1,434 (0.6%) |
| No data | 1/1,434 (0.1%) |
| Missing | 111/1,434 (7.7%) |
| VIOLATION: Ever told you had coronary heart disease | |
| No | 1,318/1,434 (91.9%) |
| Yes | 3/1,434 (0.2%) |
| No data | 2/1,434 (0.1%) |
| Missing | 111/1,434 (7.7%) |
| VIOLATION: Ever told you had angina/angina pectoris | |
| No | 1,317/1,434 (91.8%) |
| Yes | 4/1,434 (0.3%) |
| No data | 2/1,434 (0.1%) |
| Missing | 111/1,434 (7.7%) |
| VIOLATION: Ever told you had heart attack | |
| No | 1,309/1,434 (91.3%) |
| Yes | 13/1,434 (0.9%) |
| No data | 1/1,434 (0.1%) |

| | Overall sample Female N=1,434 |
|---|---|
| Missing | 111/1,434 (7.7%) |
| VIOLATION: Ever told you had a stroke | |
| No | 1,306/1,434 (91.1%) |
| Yes | 14/1,434 (1.0%) |
| No data | 3/1,434 (0.2%) |
| Missing | 111/1,434 (7.7%) |
| VIOLATION: Do you still have thyroid problem | |
| No | 30/1,434 (2.1%) |
| Yes | 77/1,434 (5.4%) |
| No data | 5/1,434 (0.3%) |
| Missing | 1,322/1,434 (92.2%) |
| VIOLATION: Ever told you had emphysema | |
| No | 1,311/1,434 (91.4%) |
| Yes | 11/1,434 (0.8%) |
| No data | 1/1,434 (0.1%) |
| Missing | 111/1,434 (7.7%) |
| VIOLATION: Ever told you had chronic bronchitis | |
| No | 1,241/1,434 (86.5%) |
| Yes | 81/1,434 (5.6%) |
| No data | 1/1,434 (0.1%) |
| Missing | 111/1,434 (7.7%) |
| VIOLATION: Ever told you had any liver condition | |
| No | 1,277/1,434 (89.1%) |
| Yes | 46/1,434 (3.2%) |
| Missing | 111/1,434 (7.7%) |
| VIOLATION: Ever told you had cancer or malignancy | |
| No | 1,262/1,434 (88.0%) |
| Yes | 59/1,434 (4.1%) |
| No data | 2/1,434 (0.1%) |
| Missing | 111/1,434 (7.7%) |
| VIOLATION: Doctor ever told you that you had gout? | |
| No | 1,311/1,434 (91.4%) |
| Yes | 10/1,434 (0.7%) |
| No data | 2/1,434 (0.1%) |
| Missing | 111/1,434 (7.7%) |
| VIOLATION: Overnight hospital patient in last year | |
| No | 1,247/1,434 (87.0%) |
| Yes | 187/1,434 (13.0%) |
| VIOLATION: SP have head cold or chest cold? | |
| No | 1,051/1,434 (73.3%) |
| Yes | 259/1,434 (18.1%) |
| No data | 124/1,434 (8.6%) |
| VIOLATION: SP have stomach or intestinal illness? | |
| No | 1,142/1,434 (79.6%) |
| Yes | 171/1,434 (11.9%) |
| No data | 121/1,434 (8.4%) |
| VIOLATION: SP have flu, pneumonia, ear infection? | |
| No | 1,238/1,434 (86.3%) |
| Yes | 74/1,434 (5.2%) |

| | Overall sample<br>Female<br>N=1,434 |
|---|---|
| No data | 122/1,434 (8.5%) |
| VIOLATION: SP donated blood in past 12 months | |
| No | 1,251/1,434 (87.2%) |
| Yes | 62/1,434 (4.3%) |
| No data | 121/1,434 (8.4%) |
| VIOLATION: Are you pregnant now? | |
| No | 680/1,434 (47.4%) |
| Yes | 40/1,434 (2.8%) |
| No data | 369/1,434 (25.7%) |
| Missing | 345/1,434 (24.1%) |
| VIOLATION: Pregnancy status at exam | |
| No | 1,020/1,434 (71.1%) |
| Yes | 49/1,434 (3.4%) |
| No data | 20/1,434 (1.4%) |
| Missing | 345/1,434 (24.1%) |
| VIOLATION: Urine Pregnancy Result | |
| No | 1,028/1,434 (71.7%) |
| Yes | 49/1,434 (3.4%) |
| No data | 12/1,434 (0.8%) |
| Missing | 345/1,434 (24.1%) |
| VIOLATION: How many months ago have baby? | |
| No | 44/1,434 (3.1%) |
| Yes | 78/1,434 (5.4%) |
| No data | 3/1,434 (0.2%) |
| Missing | 1,309/1,434 (91.3%) |
| VIOLATION: Taken prescription medicine, past month | |
| No | 798/1,434 (55.6%) |
| Yes | 636/1,434 (44.4%) |
| VIOLATION: For anemia, such as low iron | |
| Yes | 51/1,434 (3.6%) |
| No data | 1,383/1,434 (96.4%) |
| VIOLATION: Do you now smoke cigarettes? | |
| No | 166/1,434 (11.6%) |
| Yes | 317/1,434 (22.1%) |
| Missing | 951/1,434 (66.3%) |
| VIOLATION: Smoked tobacco last 5 days? | |
| No | 974/1,434 (67.9%) |
| Yes | 339/1,434 (23.6%) |
| No data | 121/1,434 (8.4%) |
| VIOLATION: Excessive alcohol intake | |
| No | 364/1,434 (25.4%) |
| Yes | 63/1,434 (4.4%) |
| No data | 110/1,434 (7.7%) |
| Missing | 897/1,434 (62.6%) |
| VIOLATION: Ferritin<15ug/L or Ferritin>150ug/L | |
| No | 1,102/1,434 (76.8%) |
| Yes | 332/1,434 (23.2%) |
| VIOLATION: C-Reactive Protein>5mg/L | |
| No | 1,068/1,434 (74.5%) |

| | Overall sample<br>Female<br>N=1,434 |
|---|---|
| Yes | 366/1,434 (25.5%) |
| VIOLATION: eGFR<90 ml/min/1.73m$^2$ | |
| No | 1,261/1,434 (87.9%) |
| Yes | 168/1,434 (11.7%) |
| No data | 5/1,434 (0.3%) |

US = United States, NHANES = National Health and Nutrition Examination Survey, BMI = Body Mass Index, SP = Survey Participant.
Data are presented as unweighted n/total (%) based on participants with non-missing haemoglobin, ferritin, and C-reactive protein. No ferritin was measured in participants more than 49 years. No ferritin was collected in males.
No data: No answer/refused, Don't know; Missing: Not applicable, Not asked.
[1] Ferritin≥15 and ≤150ug/L and C-reactive protein≤5mg/L.

Table 4.1.6 US NHANES 2009-2010: Participant characteristics of violations (Females) (18-65 years)

| | Overall sample Female N=1,703 |
|---|---|
| Reference sample[1] | |
| No | 1,536/1,703 (90.2%) |
| Yes | 167/1,703 (9.8%) |
| VIOLATION: BMI<18.5 or >30 kg/m$^2$ | |
| No | 1,020/1,703 (59.9%) |
| Yes | 672/1,703 (39.5%) |
| No data | 11/1,703 (0.6%) |
| VIOLATION: Doctor told you have diabetes | |
| No | 1,647/1,703 (96.7%) |
| Yes | 54/1,703 (3.2%) |
| No data | 2/1,703 (0.1%) |
| VIOLATION: Ever told you had weak/failing kidneys? | |
| No | 1,553/1,703 (91.2%) |
| Yes | 22/1,703 (1.3%) |
| No data | 2/1,703 (0.1%) |
| Missing | 126/1,703 (7.4%) |
| VIOLATION: Still have asthma | |
| No | 114/1,703 (6.7%) |
| Yes | 169/1,703 (9.9%) |
| No data | 6/1,703 (0.4%) |
| Missing | 1,414/1,703 (83.0%) |
| VIOLATION: Taking treatment for anemia/past 3 months | |
| No | 1,606/1,703 (94.3%) |
| Yes | 97/1,703 (5.7%) |
| VIOLATION: Doctor ever said you had arthritis | |
| No | 1,390/1,703 (81.6%) |
| Yes | 185/1,703 (10.9%) |
| No data | 2/1,703 (0.1%) |
| Missing | 126/1,703 (7.4%) |
| VIOLATION: Ever told had congestive heart failure | |
| No | 1,567/1,703 (92.0%) |
| Yes | 7/1,703 (0.4%) |
| No data | 3/1,703 (0.2%) |
| Missing | 126/1,703 (7.4%) |
| VIOLATION: Ever told you had coronary heart disease | |
| No | 1,570/1,703 (92.2%) |
| Yes | 4/1,703 (0.2%) |
| No data | 3/1,703 (0.2%) |
| Missing | 126/1,703 (7.4%) |
| VIOLATION: Ever told you had angina/angina pectoris | |
| No | 1,569/1,703 (92.1%) |
| Yes | 6/1,703 (0.4%) |
| No data | 2/1,703 (0.1%) |
| Missing | 126/1,703 (7.4%) |
| VIOLATION: Ever told you had heart attack | |
| No | 1,559/1,703 (91.5%) |
| Yes | 13/1,703 (0.8%) |

| | Overall sample Female N=1,703 |
|---|---|
| No data | 5/1,703 (0.3%) |
| Missing | 126/1,703 (7.4%) |
| VIOLATION: Ever told you had a stroke | |
| No | 1,557/1,703 (91.4%) |
| Yes | 20/1,703 (1.2%) |
| Missing | 126/1,703 (7.4%) |
| VIOLATION: Do you still have thyroid problem | |
| No | 23/1,703 (1.4%) |
| Yes | 97/1,703 (5.7%) |
| No data | 7/1,703 (0.4%) |
| Missing | 1,576/1,703 (92.5%) |
| VIOLATION: Ever told you had emphysema | |
| No | 1,563/1,703 (91.8%) |
| Yes | 14/1,703 (0.8%) |
| Missing | 126/1,703 (7.4%) |
| VIOLATION: Ever told you had chronic bronchitis | |
| No | 1,500/1,703 (88.1%) |
| Yes | 76/1,703 (4.5%) |
| No data | 1/1,703 (0.1%) |
| Missing | 126/1,703 (7.4%) |
| VIOLATION: Ever told you had any liver condition | |
| No | 1,542/1,703 (90.5%) |
| Yes | 34/1,703 (2.0%) |
| No data | 1/1,703 (0.1%) |
| Missing | 126/1,703 (7.4%) |
| VIOLATION: Ever told you had cancer or malignancy | |
| No | 1,503/1,703 (88.3%) |
| Yes | 74/1,703 (4.3%) |
| Missing | 126/1,703 (7.4%) |
| VIOLATION: Doctor ever told you that you had gout? | |
| No | 1,570/1,703 (92.2%) |
| Yes | 7/1,703 (0.4%) |
| Missing | 126/1,703 (7.4%) |
| VIOLATION: Overnight hospital patient in last year | |
| No | 1,460/1,703 (85.7%) |
| Yes | 243/1,703 (14.3%) |
| VIOLATION: SP have head cold or chest cold? | |
| No | 1,175/1,703 (69.0%) |
| Yes | 280/1,703 (16.4%) |
| No data | 248/1,703 (14.6%) |
| VIOLATION: SP have stomach or intestinal illness? | |
| No | 1,348/1,703 (79.2%) |
| Yes | 108/1,703 (6.3%) |
| No data | 247/1,703 (14.5%) |
| VIOLATION: SP have flu, pneumonia, ear infection? | |
| No | 1,384/1,703 (81.3%) |
| Yes | 67/1,703 (3.9%) |
| No data | 252/1,703 (14.8%) |
| VIOLATION: SP donated blood in past 12 months | |

| | Overall sample Female N=1,703 |
|---|---|
| No | 1,368/1,703 (80.3%) |
| Yes | 88/1,703 (5.2%) |
| No data | 247/1,703 (14.5%) |
| VIOLATION: Are you pregnant now? | |
| No | 713/1,703 (41.9%) |
| Yes | 44/1,703 (2.6%) |
| No data | 549/1,703 (32.2%) |
| Missing | 397/1,703 (23.3%) |
| VIOLATION: Pregnancy status at exam | |
| No | 1,212/1,703 (71.2%) |
| Yes | 64/1,703 (3.8%) |
| No data | 30/1,703 (1.8%) |
| Missing | 397/1,703 (23.3%) |
| VIOLATION: Urine Pregnancy Result | |
| No | 1,232/1,703 (72.3%) |
| Yes | 64/1,703 (3.8%) |
| No data | 10/1,703 (0.6%) |
| Missing | 397/1,703 (23.3%) |
| VIOLATION: How many months ago have baby? | |
| No | 58/1,703 (3.4%) |
| Yes | 80/1,703 (4.7%) |
| Missing | 1,565/1,703 (91.9%) |
| VIOLATION: Taken prescription medicine, past month | |
| No | 942/1,703 (55.3%) |
| Yes | 761/1,703 (44.7%) |
| VIOLATION: For anemia, such as low iron | |
| Yes | 53/1,703 (3.1%) |
| No data | 1,650/1,703 (96.9%) |
| VIOLATION: Do you now smoke cigarettes? | |
| No | 187/1,703 (11.0%) |
| Yes | 397/1,703 (23.3%) |
| Missing | 1,119/1,703 (65.7%) |
| VIOLATION: Smoked tobacco last 5 days? | |
| No | 1,072/1,703 (62.9%) |
| Yes | 382/1,703 (22.4%) |
| No data | 249/1,703 (14.6%) |
| VIOLATION: Excessive alcohol intake | |
| No | 370/1,703 (21.7%) |
| Yes | 80/1,703 (4.7%) |
| No data | 234/1,703 (13.7%) |
| Missing | 1,019/1,703 (59.8%) |
| VIOLATION: Ferritin<15ug/L or Ferritin>150ug/L (F/M) | |
| No | 1,308/1,703 (76.8%) |
| Yes | 395/1,703 (23.2%) |
| VIOLATION: C-Reactive Protein>5mg/L | |
| No | 1,267/1,703 (74.4%) |
| Yes | 436/1,703 (25.6%) |
| VIOLATION: eGFR<90 ml/min/1.73m$^2$ | |
| No | 1,465/1,703 (86.0%) |

| | Overall sample<br>Female<br>N=1,703 |
|---|---|
| Yes | 222/1,703 (13.0%) |
| No data | 16/1,703 (0.9%) |

US = United States, NHANES = National Health and Nutrition Examination Survey, BMI = Body Mass Index, SP = Survey Participant.
Data are presented as unweighted n/total (%) based on participants with non-missing haemoglobin, ferritin, and C-reactive protein. No ferritin was measured in participants more than 49 years. No ferritin was collected in males.
No data: No answer/refused, Don't know; Missing: Not applicable, Not asked.
[1] Ferritin≥15 and ≤150ug/L and C-reactive protein≤5mg/L.

#### Table 4.1.7 US NHANES 2015-2016: Participant characteristics of violations (Females) (18-65 years)

| | Overall sample Female N=1,485 |
|---|---|
| Reference sample[1] | |
| No | 1,317/1,485 (88.7%) |
| Yes | 168/1,485 (11.3%) |
| VIOLATION: BMI<18.5 or >30 kg/m$^2$ | |
| No | 830/1,485 (55.9%) |
| Yes | 643/1,485 (43.3%) |
| No data | 12/1,485 (0.8%) |
| VIOLATION: Doctor told you have diabetes | |
| No | 1,403/1,485 (94.5%) |
| Yes | 81/1,485 (5.5%) |
| No data | 1/1,485 (0.1%) |
| VIOLATION: Ever told you had weak/failing kidneys? | |
| No | 1,352/1,485 (91.0%) |
| Yes | 23/1,485 (1.5%) |
| No data | 1/1,485 (0.1%) |
| Missing | 109/1,485 (7.3%) |
| VIOLATION: Still have asthma | |
| No | 91/1,485 (6.1%) |
| Yes | 157/1,485 (10.6%) |
| No data | 4/1,485 (0.3%) |
| Missing | 1,233/1,485 (83.0%) |
| VIOLATION: Taking treatment for anemia/past 3 months | |
| No | 1,384/1,485 (93.2%) |
| Yes | 99/1,485 (6.7%) |
| No data | 2/1,485 (0.1%) |
| VIOLATION: Doctor ever said you had arthritis | |
| No | 1,209/1,485 (81.4%) |
| Yes | 165/1,485 (11.1%) |
| No data | 2/1,485 (0.1%) |
| Missing | 109/1,485 (7.3%) |
| VIOLATION: Ever told had congestive heart failure | |
| No | 1,369/1,485 (92.2%) |
| Yes | 7/1,485 (0.5%) |
| Missing | 109/1,485 (7.3%) |
| VIOLATION: Ever told you had coronary heart disease | |
| No | 1,369/1,485 (92.2%) |
| Yes | 6/1,485 (0.4%) |
| No data | 1/1,485 (0.1%) |
| Missing | 109/1,485 (7.3%) |
| VIOLATION: Ever told you had angina/angina pectoris | |
| No | 1,366/1,485 (92.0%) |
| Yes | 9/1,485 (0.6%) |
| No data | 1/1,485 (0.1%) |
| Missing | 109/1,485 (7.3%) |
| VIOLATION: Ever told you had heart attack | |
| No | 1,365/1,485 (91.9%) |
| Yes | 11/1,485 (0.7%) |

| | Overall sample Female N=1,485 |
|---|---|
| Missing | 109/1,485 (7.3%) |
| VIOLATION: Ever told you had a stroke | |
| No | 1,364/1,485 (91.9%) |
| Yes | 12/1,485 (0.8%) |
| Missing | 109/1,485 (7.3%) |
| VIOLATION: Do you still have thyroid problem | |
| No | 29/1,485 (2.0%) |
| Yes | 81/1,485 (5.5%) |
| No data | 13/1,485 (0.9%) |
| Missing | 1,362/1,485 (91.7%) |
| VIOLATION: Ever told you had emphysema | |
| No | 1,370/1,485 (92.3%) |
| Yes | 6/1,485 (0.4%) |
| Missing | 109/1,485 (7.3%) |
| VIOLATION: Ever told you had chronic bronchitis | |
| No | 1,305/1,485 (87.9%) |
| Yes | 67/1,485 (4.5%) |
| No data | 4/1,485 (0.3%) |
| Missing | 109/1,485 (7.3%) |
| VIOLATION: Ever told you had any liver condition | |
| No | 1,340/1,485 (90.2%) |
| Yes | 32/1,485 (2.2%) |
| No data | 4/1,485 (0.3%) |
| Missing | 109/1,485 (7.3%) |
| VIOLATION: Ever told you had COPD? | |
| No | 1,363/1,485 (91.8%) |
| Yes | 13/1,485 (0.9%) |
| Missing | 109/1,485 (7.3%) |
| VIOLATION: Ever been told you have jaundice? | |
| No | 1,452/1,485 (97.8%) |
| Yes | 31/1,485 (2.1%) |
| No data | 2/1,485 (0.1%) |
| VIOLATION: Ever told you had cancer or malignancy | |
| No | 1,336/1,485 (90.0%) |
| Yes | 40/1,485 (2.7%) |
| Missing | 109/1,485 (7.3%) |
| VIOLATION: Ever been told you have jaundice? | |
| No | 1,452/1,485 (97.8%) |
| Yes | 31/1,485 (2.1%) |
| No data | 2/1,485 (0.1%) |
| VIOLATION: Overnight hospital patient in last year | |
| No | 1,318/1,485 (88.8%) |
| Yes | 167/1,485 (11.2%) |
| VIOLATION: SP have head cold or chest cold? | |
| No | 1,062/1,485 (71.5%) |
| Yes | 230/1,485 (15.5%) |
| No data | 193/1,485 (13.0%) |
| VIOLATION: SP have stomach or intestinal illness? | |
| No | 1,177/1,485 (79.3%) |

| | Overall sample<br>Female<br>N=1,485 |
|---|---|
| Yes | 114/1,485 (7.7%) |
| No data | 194/1,485 (13.1%) |
| VIOLATION: SP have flu, pneumonia, ear infection? | |
| No | 1,248/1,485 (84.0%) |
| Yes | 43/1,485 (2.9%) |
| No data | 194/1,485 (13.1%) |
| VIOLATION: SP donated blood in past 12 months | |
| No | 1,216/1,485 (81.9%) |
| Yes | 76/1,485 (5.1%) |
| No data | 193/1,485 (13.0%) |
| VIOLATION: Are you pregnant now? | |
| No | 611/1,485 (41.1%) |
| Yes | 46/1,485 (3.1%) |
| No data | 59/1,485 (4.0%) |
| Missing | 769/1,485 (51.8%) |
| VIOLATION: Pregnancy status at exam | |
| No | 1,060/1,485 (71.4%) |
| Yes | 60/1,485 (4.0%) |
| No data | 28/1,485 (1.9%) |
| Missing | 337/1,485 (22.7%) |
| VIOLATION: Urine Pregnancy Result | |
| No | 1,081/1,485 (72.8%) |
| Yes | 59/1,485 (4.0%) |
| No data | 8/1,485 (0.5%) |
| Missing | 337/1,485 (22.7%) |
| VIOLATION: How many months ago have baby? | |
| No | 82/1,485 (5.5%) |
| Yes | 84/1,485 (5.7%) |
| No data | 982/1,485 (66.1%) |
| Missing | 337/1,485 (22.7%) |
| VIOLATION: Taken prescription medicine, past month | |
| No | 840/1,485 (56.6%) |
| Yes | 644/1,485 (43.4%) |
| No data | 1/1,485 (0.1%) |
| VIOLATION: For anemia, such as low iron | |
| Yes | 71/1,485 (4.8%) |
| No data | 1,414/1,485 (95.2%) |
| VIOLATION: Do you now smoke cigarettes? | |
| No | 149/1,485 (10.0%) |
| Yes | 220/1,485 (14.8%) |
| Missing | 1,116/1,485 (75.2%) |
| VIOLATION: Smoked tobacco last 5 days? | |
| No | 1,049/1,485 (70.6%) |
| Yes | 243/1,485 (16.4%) |
| No data | 193/1,485 (13.0%) |
| VIOLATION: Do you now smoke cigarettes? | |
| No | 401/1,485 (27.0%) |
| Yes | 17/1,485 (1.1%) |
| Missing | 1,067/1,485 (71.9%) |

| | Overall sample Female N=1,485 |
|---|---|
| VIOLATION: Excessive alcohol intake | |
| No | 419/1,485 (28.2%) |
| Yes | 54/1,485 (3.6%) |
| No data | 194/1,485 (13.1%) |
| Missing | 818/1,485 (55.1%) |
| VIOLATION: Ferritin<15ug/L or Ferritin>150ug/L (F/M) | |
| No | 1,120/1,485 (75.4%) |
| Yes | 365/1,485 (24.6%) |
| VIOLATION: C-Reactive Protein>5mg/L | |
| No | 1,083/1,485 (72.9%) |
| Yes | 402/1,485 (27.1%) |
| VIOLATION: eGFR<90 ml/min/1.73m$^2$ | |
| No | 1,309/1,485 (88.1%) |
| Yes | 174/1,485 (11.7%) |
| No data | 2/1,485 (0.1%) |

US = United States, NHANES = National Health and Nutrition Examination Survey, BMI = Body Mass Index, SP = Survey Participant.
Data are presented as unweighted n/total (%) based on participants with non-missing haemoglobin, ferritin, and C-reactive protein. No ferritin was measured in participants more than 49 years. No ferritin was collected in males.
No data: No answer/refused, Don't know; Missing: Not applicable, Not asked.
[1] Ferritin≥15 and ≤150ug/L and C-reactive protein≤5mg/L.

#### Table 4.1.8 US NHANES 2017-2018: Participant characteristics of violations (18-65 years)

| | Overall sample | |
|---|---|---|
| | Male<br>N=1,854 | Female<br>N=2,059 |
| Reference sample[1] | | |
| No | 1,705/1,854 (92.0%) | 1,916/2,059 (93.1%) |
| Yes | 149/1,854 (8.0%) | 143/2,059 (6.9%) |
| VIOLATION: BMI<18.5 or >30 kg/m[2] | | |
| No | 1,080/1,854 (58.3%) | 1,094/2,059 (53.1%) |
| Yes | 750/1,854 (40.5%) | 946/2,059 (45.9%) |
| No data | 24/1,854 (1.3%) | 19/2,059 (0.9%) |
| VIOLATION: Doctor told you have diabetes | | |
| No | 1,626/1,854 (87.7%) | 1,856/2,059 (90.1%) |
| Yes | 227/1,854 (12.2%) | 201/2,059 (9.8%) |
| No data | 1/1,854 (0.1%) | 2/2,059 (0.1%) |
| VIOLATION: Ever told you had weak/failing kidneys? | | |
| No | 1,693/1,854 (91.3%) | 1,888/2,059 (91.7%) |
| Yes | 45/1,854 (2.4%) | 48/2,059 (2.3%) |
| No data | 1/1,854 (0.1%) | 1/2,059 (0.0%) |
| Missing | 115/1,854 (6.2%) | 122/2,059 (5.9%) |
| VIOLATION: Still have asthma | | |
| No | 125/1,854 (6.7%) | 103/2,059 (5.0%) |
| Yes | 140/1,854 (7.6%) | 245/2,059 (11.9%) |
| No data | 8/1,854 (0.4%) | 5/2,059 (0.2%) |
| Missing | 1,581/1,854 (85.3%) | 1,706/2,059 (82.9%) |
| VIOLATION: Taking treatment for anemia/past 3 months | | |
| No | 1,835/1,854 (99.0%) | 1,891/2,059 (91.8%) |
| Yes | 17/1,854 (0.9%) | 166/2,059 (8.1%) |
| No data | 2/1,854 (0.1%) | 2/2,059 (0.1%) |
| VIOLATION: Doctor ever said you had arthritis | | |
| No | 1,389/1,854 (74.9%) | 1,449/2,059 (70.4%) |
| Yes | 346/1,854 (18.7%) | 483/2,059 (23.5%) |
| No data | 4/1,854 (0.2%) | 5/2,059 (0.2%) |
| Missing | 115/1,854 (6.2%) | 122/2,059 (5.9%) |
| VIOLATION: Ever told had congestive heart failure | | |
| No | 1,692/1,854 (91.3%) | 1,911/2,059 (92.8%) |
| Yes | 43/1,854 (2.3%) | 23/2,059 (1.1%) |
| No data | 4/1,854 (0.2%) | 3/2,059 (0.1%) |
| Missing | 115/1,854 (6.2%) | 122/2,059 (5.9%) |
| VIOLATION: Ever told you had coronary heart disease | | |
| No | 1,677/1,854 (90.5%) | 1,912/2,059 (92.9%) |
| Yes | 58/1,854 (3.1%) | 24/2,059 (1.2%) |
| No data | 4/1,854 (0.2%) | 1/2,059 (0.0%) |
| Missing | 115/1,854 (6.2%) | 122/2,059 (5.9%) |
| VIOLATION: Ever told you had angina/angina pectoris | | |
| No | 1,698/1,854 (91.6%) | 1,898/2,059 (92.2%) |
| Yes | 34/1,854 (1.8%) | 33/2,059 (1.6%) |
| No data | 7/1,854 (0.4%) | 6/2,059 (0.3%) |
| Missing | 115/1,854 (6.2%) | 122/2,059 (5.9%) |
| VIOLATION: Ever told you had heart attack | | |
| No | 1,665/1,854 (89.8%) | 1,899/2,059 (92.2%) |
| Yes | 71/1,854 (3.8%) | 34/2,059 (1.7%) |

| | Overall sample | |
|---|---|---|
| | Male<br>N=1,854 | Female<br>N=2,059 |
| No data | 3/1,854 (0.2%) | 4/2,059 (0.2%) |
| Missing | 115/1,854 (6.2%) | 122/2,059 (5.9%) |
| VIOLATION: Ever told you had a stroke | | |
| No | 1,683/1,854 (90.8%) | 1,877/2,059 (91.2%) |
| Yes | 54/1,854 (2.9%) | 57/2,059 (2.8%) |
| No data | 2/1,854 (0.1%) | 3/2,059 (0.1%) |
| Missing | 115/1,854 (6.2%) | 122/2,059 (5.9%) |
| VIOLATION: Do you still have thyroid problem | | |
| No | 16/1,854 (0.9%) | 54/2,059 (2.6%) |
| Yes | 53/1,854 (2.9%) | 200/2,059 (9.7%) |
| No data | 3/1,854 (0.2%) | 11/2,059 (0.5%) |
| Missing | 1,782/1,854 (96.1%) | 1,794/2,059 (87.1%) |
| VIOLATION: Ever told you had emphysema | | |
| No | 1,713/1,854 (92.4%) | 1,917/2,059 (93.1%) |
| Yes | 24/1,854 (1.3%) | 17/2,059 (0.8%) |
| No data | 2/1,854 (0.1%) | 3/2,059 (0.1%) |
| Missing | 115/1,854 (6.2%) | 122/2,059 (5.9%) |
| VIOLATION: Ever told you had chronic bronchitis | | |
| No | 1,643/1,854 (88.6%) | 1,809/2,059 (87.9%) |
| Yes | 96/1,854 (5.2%) | 125/2,059 (6.1%) |
| No data | 0/1,854 (0.0%) | 3/2,059 (0.1%) |
| Missing | 115/1,854 (6.2%) | 122/2,059 (5.9%) |
| VIOLATION: Ever told you had any liver condition | | |
| No | 1,633/1,854 (88.1%) | 1,849/2,059 (89.8%) |
| Yes | 103/1,854 (5.6%) | 82/2,059 (4.0%) |
| No data | 3/1,854 (0.2%) | 6/2,059 (0.3%) |
| Missing | 115/1,854 (6.2%) | 122/2,059 (5.9%) |
| VIOLATION: Ever told you had cancer or malignancy | | |
| No | 1,668/1,854 (90.0%) | 1,806/2,059 (87.7%) |
| Yes | 71/1,854 (3.8%) | 129/2,059 (6.3%) |
| No data | 0/1,854 (0.0%) | 2/2,059 (0.1%) |
| Missing | 115/1,854 (6.2%) | 122/2,059 (5.9%) |
| VIOLATION: Ever been told you have jaundice? | | |
| No | 1,798/1,854 (97.0%) | 2,013/2,059 (97.8%) |
| Yes | 53/1,854 (2.9%) | 46/2,059 (2.2%) |
| No data | 3/1,854 (0.2%) | 0/2,059 (0.0%) |
| VIOLATION: Ever told you had COPD? | | |
| No | 1,680/1,854 (90.6%) | 1,869/2,059 (90.8%) |
| Yes | 57/1,854 (3.1%) | 67/2,059 (3.3%) |
| No data | 2/1,854 (0.1%) | 1/2,059 (0.0%) |
| Missing | 115/1,854 (6.2%) | 122/2,059 (5.9%) |
| VIOLATION: Has DR ever said you have gallstones | | |
| No | 1,661/1,854 (89.6%) | 1,666/2,059 (80.9%) |
| Yes | 75/1,854 (4.0%) | 265/2,059 (12.9%) |
| No data | 3/1,854 (0.2%) | 6/2,059 (0.3%) |
| Missing | 115/1,854 (6.2%) | 122/2,059 (5.9%) |
| VIOLATION: Overnight hospital patient in last year | | |
| No | 1,721/1,854 (92.8%) | 1,848/2,059 (89.8%) |
| Yes | 133/1,854 (7.2%) | 211/2,059 (10.2%) |

| | Overall sample | |
|---|---|---|
| | Male<br>N=1,854 | Female<br>N=2,059 |
| VIOLATION: SP have head cold or chest cold? | | |
| No | 1,485/1,854 (80.1%) | 1,541/2,059 (74.8%) |
| Yes | 273/1,854 (14.7%) | 359/2,059 (17.4%) |
| No data | 96/1,854 (5.2%) | 159/2,059 (7.7%) |
| VIOLATION: SP have stomach or intestinal illness? | | |
| No | 1,641/1,854 (88.5%) | 1,746/2,059 (84.8%) |
| Yes | 120/1,854 (6.5%) | 160/2,059 (7.8%) |
| No data | 93/1,854 (5.0%) | 153/2,059 (7.4%) |
| VIOLATION: SP have flu, pneumonia, ear infection? | | |
| No | 1,692/1,854 (91.3%) | 1,814/2,059 (88.1%) |
| Yes | 71/1,854 (3.8%) | 93/2,059 (4.5%) |
| No data | 91/1,854 (4.9%) | 152/2,059 (7.4%) |
| VIOLATION: SP donated blood in past 12 months | | |
| No | 1,644/1,854 (88.7%) | 1,820/2,059 (88.4%) |
| Yes | 118/1,854 (6.4%) | 87/2,059 (4.2%) |
| No data | 92/1,854 (5.0%) | 152/2,059 (7.4%) |
| VIOLATION: Are you pregnant now? | | |
| No | 0/1,854 (0.0%) | 579/2,059 (28.1%) |
| Yes | 0/1,854 (0.0%) | 37/2,059 (1.8%) |
| No data | 0/1,854 (0.0%) | 48/2,059 (2.3%) |
| Missing | 1,854/1,854 (100.0%) | 1,395/2,059 (67.8%) |
| VIOLATION: Pregnancy status at exam | | |
| No | 0/1,854 (0.0%) | 911/2,059 (44.2%) |
| Yes | 0/1,854 (0.0%) | 47/2,059 (2.3%) |
| No data | 0/1,854 (0.0%) | 22/2,059 (1.1%) |
| Missing | 1,854/1,854 (100.0%) | 1,079/2,059 (52.4%) |
| VIOLATION: Urine Pregnancy Result | | |
| No | 0/1,854 (0.0%) | 927/2,059 (45.0%) |
| Yes | 0/1,854 (0.0%) | 47/2,059 (2.3%) |
| No data | 0/1,854 (0.0%) | 6/2,059 (0.3%) |
| Missing | 1,854/1,854 (100.0%) | 1,079/2,059 (52.4%) |
| VIOLATION: How many months ago have baby? | | |
| No | 0/1,854 (0.0%) | 904/2,059 (43.9%) |
| Yes | 0/1,854 (0.0%) | 76/2,059 (3.7%) |
| Missing | 1,854/1,854 (100.0%) | 1,079/2,059 (52.4%) |
| VIOLATION: Taken prescription medicine, past month | | |
| No | 1,039/1,854 (56.0%) | 938/2,059 (45.6%) |
| Yes | 812/1,854 (43.8%) | 1,117/2,059 (54.2%) |
| No data | 3/1,854 (0.2%) | 4/2,059 (0.2%) |
| VIOLATION: For anemia, such as low iron | | |
| No | 1,829/1,854 (98.7%) | 1,930/2,059 (93.7%) |
| Yes | 25/1,854 (1.3%) | 129/2,059 (6.3%) |
| VIOLATION: Do you now smoke cigarettes? | | |
| No | 432/1,854 (23.3%) | 284/2,059 (13.8%) |
| Yes | 443/1,854 (23.9%) | 329/2,059 (16.0%) |
| Missing | 979/1,854 (52.8%) | 1,446/2,059 (70.2%) |
| VIOLATION: Smoked tobacco last 5 days? | | |
| No | 1,230/1,854 (66.3%) | 1,539/2,059 (74.7%) |
| Yes | 533/1,854 (28.7%) | 367/2,059 (17.8%) |

| | Overall sample | |
|---|---|---|
| | Male<br>N=1,854 | Female<br>N=2,059 |
| No data | 91/1,854 (4.9%) | 153/2,059 (7.4%) |
| VIOLATION: Excessive alcohol intake | | |
| No | 414/1,854 (22.3%) | 548/2,059 (26.6%) |
| Yes | 136/1,854 (7.3%) | 84/2,059 (4.1%) |
| No data | 94/1,854 (5.1%) | 156/2,059 (7.6%) |
| Missing | 1,210/1,854 (65.3%) | 1,271/2,059 (61.7%) |
| VIOLATION: Ferritin<15ug/L or Ferritin>150/200ug/L (F/M) | | |
| No | 1,108/1,854 (59.8%) | 1,499/2,059 (72.8%) |
| Yes | 746/1,854 (40.2%) | 560/2,059 (27.2%) |
| VIOLATION: C-Reactive Protein>5mg/L | | |
| No | 1,537/1,854 (82.9%) | 1,471/2,059 (71.4%) |
| Yes | 317/1,854 (17.1%) | 588/2,059 (28.6%) |
| VIOLATION: eGFR<90 ml/min/1.73m$^2$ | | |
| No | 1,322/1,854 (71.3%) | 1,606/2,059 (78.0%) |
| Yes | 529/1,854 (28.5%) | 450/2,059 (21.9%) |
| No data | 3/1,854 (0.2%) | 3/2,059 (0.1%) |

US = United States, NHANES = National Health and Nutrition Examination Survey, BMI = Body Mass Index, SP = Survey Participant, F = Female, M = Male.
Data are presented as unweighted n/total (%) based on participants with non-missing haemoglobin, ferritin, and C-reactive protein.
No data: No answer/refused, Don't know; Missing: Not applicable, Not asked.
[1] Males: ferritin≥15 and ≤200ug/L and C-reactive protein≤5mg/L; females: ferritin≥15 and ≤150ug/L and C-reactive protein≤5mg/L.

#### Table 4.1.9 England HSE 1998: Participant characteristics of violations (18-65 years)

| | Overall sample | |
|---|---|---|
| | Male N=3,835 | Female N=4,189 |
| Reference sample[1] | | |
| No | 3,268/3,835 (85.2%) | 3,613/4,189 (86.2%) |
| Yes | 567/3,835 (14.8%) | 576/4,189 (13.8%) |
| VIOLATION: BMI<18.5 or >30 kg/m[2] | | |
| No | 3,009/3,835 (78.5%) | 3,120/4,189 (74.5%) |
| Yes | 675/3,835 (17.6%) | 871/4,189 (20.8%) |
| No data | 151/3,835 (3.9%) | 198/4,189 (4.7%) |
| VIOLATION: whether has clotting disorder | | |
| No | 3,806/3,835 (99.2%) | 4,169/4,189 (99.5%) |
| Yes | 29/3,835 (0.8%) | 20/4,189 (0.5%) |
| VIOLATION: (d) had cardiovascular condition | | |
| No | 2,946/3,835 (76.8%) | 3,330/4,189 (79.5%) |
| Yes | 885/3,835 (23.1%) | 855/4,189 (20.4%) |
| No data | 4/3,835 (0.1%) | 4/4,189 (0.1%) |
| VIOLATION: (d) had cvd: excludes those with high bp | | |
| No | 3,408/3,835 (88.9%) | 3,791/4,189 (90.5%) |
| Yes | 424/3,835 (11.1%) | 394/4,189 (9.4%) |
| No data | 3/3,835 (0.1%) | 4/4,189 (0.1%) |
| VIOLATION: (d) had ihd (angina or heart attack) | | |
| No | 3,686/3,835 (96.1%) | 4,117/4,189 (98.3%) |
| Yes | 148/3,835 (3.9%) | 71/4,189 (1.7%) |
| No data | 1/3,835 (0.0%) | 1/4,189 (0.0%) |
| VIOLATION: (d) had cvd (angina, heart attack or stroke) | | |
| No | 3,660/3,835 (95.4%) | 4,096/4,189 (97.8%) |
| Yes | 174/3,835 (4.5%) | 92/4,189 (2.2%) |
| No data | 1/3,835 (0.0%) | 1/4,189 (0.0%) |
| VIOLATION: (d) angina or mi (rose angina qure) | | |
| No | 3,474/3,835 (90.6%) | 3,905/4,189 (93.2%) |
| Yes | 354/3,835 (9.2%) | 282/4,189 (6.7%) |
| No data | 7/3,835 (0.2%) | 2/4,189 (0.0%) |
| VIOLATION: (d) angina symptoms (rose angina qure) | | |
| No | 3,762/3,835 (98.1%) | 4,087/4,189 (97.6%) |
| Yes | 66/3,835 (1.7%) | 101/4,189 (2.4%) |
| No data | 7/3,835 (0.2%) | 1/4,189 (0.0%) |
| VIOLATION: (d) possible infarction (rose angina qure) | | |
| No | 3,525/3,835 (91.9%) | 3,987/4,189 (95.2%) |
| Yes | 310/3,835 (8.1%) | 201/4,189 (4.8%) |
| No data | 0/3,835 (0.0%) | 1/4,189 (0.0%) |
| VIOLATION: (d) doctor diagnosed angina | | |
| No | 3,728/3,835 (97.2%) | 4,130/4,189 (98.6%) |
| Yes | 107/3,835 (2.8%) | 59/4,189 (1.4%) |
| VIOLATION: (d) doctor diagnosed high blood pressure (excluding pregnant) | | |
| No | 3,209/3,835 (83.7%) | 3,599/4,189 (85.9%) |
| Yes | 625/3,835 (16.3%) | 590/4,189 (14.1%) |
| No data | 1/3,835 (0.0%) | 0/4,189 (0.0%) |
| VIOLATION: (d) doctor diagnosed heart attack | | |
| No | 3,745/3,835 (97.7%) | 4,163/4,189 (99.4%) |

| | Overall sample | |
|---|---|---|
| | Male<br>N=3,835 | Female<br>N=4,189 |
| Yes | 89/3,835 (2.3%) | 25/4,189 (0.6%) |
| No data | 1/3,835 (0.0%) | 1/4,189 (0.0%) |
| VIOLATION: (d) doctor diagnosed stroke | | |
| No | 3,796/3,835 (99.0%) | 4,162/4,189 (99.4%) |
| Yes | 39/3,835 (1.0%) | 27/4,189 (0.6%) |
| VIOLATION: (d) doctor diagnosed diabetes (excluding pregnant) | | |
| No | 3,742/3,835 (97.6%) | 4,129/4,189 (98.6%) |
| Yes | 93/3,835 (2.4%) | 60/4,189 (1.4%) |
| VIOLATION: (d) doctor diagnosed heart murmur (excluding pregnant) | | |
| No | 3,738/3,835 (97.5%) | 4,052/4,189 (96.7%) |
| Yes | 96/3,835 (2.5%) | 136/4,189 (3.2%) |
| No data | 1/3,835 (0.0%) | 1/4,189 (0.0%) |
| VIOLATION: (d) doctor diagnosed irregular heart rhythm | | |
| No | 3,682/3,835 (96.0%) | 4,029/4,189 (96.2%) |
| Yes | 151/3,835 (3.9%) | 159/4,189 (3.8%) |
| No data | 2/3,835 (0.1%) | 1/4,189 (0.0%) |
| VIOLATION: (d) doctor diagnosed other heart condition | | |
| No | 3,796/3,835 (99.0%) | 4,151/4,189 (99.1%) |
| Yes | 39/3,835 (1.0%) | 37/4,189 (0.9%) |
| No data | 0/3,835 (0.0%) | 1/4,189 (0.0%) |
| VIOLATION: surgery for heart condition | | |
| No | 3,785/3,835 (98.7%) | 4,166/4,189 (99.5%) |
| Yes | 50/3,835 (1.3%) | 23/4,189 (0.5%) |
| VIOLATION: any surgery for heart murmur | | |
| No | 3,829/3,835 (99.8%) | 4,185/4,189 (99.9%) |
| Yes | 6/3,835 (0.2%) | 4/4,189 (0.1%) |
| VIOLATION: (d) i infectious disease | | |
| No | 3,825/3,835 (99.7%) | 4,182/4,189 (99.8%) |
| Yes | 8/3,835 (0.2%) | 6/4,189 (0.1%) |
| No data | 2/3,835 (0.1%) | 1/4,189 (0.0%) |
| VIOLATION: (d) ii neoplasms & benign growths | | |
| No | 3,809/3,835 (99.3%) | 4,129/4,189 (98.6%) |
| Yes | 24/3,835 (0.6%) | 59/4,189 (1.4%) |
| No data | 2/3,835 (0.1%) | 1/4,189 (0.0%) |
| VIOLATION: (d) iii endocrine & metabolic | | |
| No | 3,696/3,835 (96.4%) | 4,006/4,189 (95.6%) |
| Yes | 137/3,835 (3.6%) | 182/4,189 (4.3%) |
| No data | 2/3,835 (0.1%) | 1/4,189 (0.0%) |
| VIOLATION: (d) iv blood and related organs | | |
| No | 3,822/3,835 (99.7%) | 4,168/4,189 (99.5%) |
| Yes | 11/3,835 (0.3%) | 20/4,189 (0.5%) |
| No data | 2/3,835 (0.1%) | 1/4,189 (0.0%) |
| VIOLATION: (d) v mental disorders | | |
| No | 3,757/3,835 (98.0%) | 4,066/4,189 (97.1%) |
| Yes | 76/3,835 (2.0%) | 122/4,189 (2.9%) |
| No data | 2/3,835 (0.1%) | 1/4,189 (0.0%) |
| VIOLATION: (d) vi nervous system | | |
| No | 3,737/3,835 (97.4%) | 4,031/4,189 (96.2%) |

| | Overall sample | |
|---|---|---|
| | Male<br>N=3,835 | Female<br>N=4,189 |
| Yes | 96/3,835 (2.5%) | 157/4,189 (3.7%) |
| No data | 2/3,835 (0.1%) | 1/4,189 (0.0%) |
| VIOLATION: (d) vi eye complaints | | |
| No | 3,778/3,835 (98.5%) | 4,143/4,189 (98.9%) |
| Yes | 55/3,835 (1.4%) | 45/4,189 (1.1%) |
| No data | 2/3,835 (0.1%) | 1/4,189 (0.0%) |
| VIOLATION: (d) vi ear complaints | | |
| No | 3,725/3,835 (97.1%) | 4,126/4,189 (98.5%) |
| Yes | 108/3,835 (2.8%) | 62/4,189 (1.5%) |
| No data | 2/3,835 (0.1%) | 1/4,189 (0.0%) |
| VIOLATION: (d) vii heart and circulatory system | | |
| No | 3,566/3,835 (93.0%) | 3,949/4,189 (94.3%) |
| Yes | 267/3,835 (7.0%) | 239/4,189 (5.7%) |
| No data | 2/3,835 (0.1%) | 1/4,189 (0.0%) |
| VIOLATION: (d) respiratory system | | |
| No | 3,488/3,835 (91.0%) | 3,825/4,189 (91.3%) |
| Yes | 345/3,835 (9.0%) | 363/4,189 (8.7%) |
| No data | 2/3,835 (0.1%) | 1/4,189 (0.0%) |
| VIOLATION: (d) ix digestive system | | |
| No | 3,657/3,835 (95.4%) | 4,005/4,189 (95.6%) |
| Yes | 176/3,835 (4.6%) | 183/4,189 (4.4%) |
| No data | 2/3,835 (0.1%) | 1/4,189 (0.0%) |
| VIOLATION: (d) x genito-urinary system | | |
| No | 3,777/3,835 (98.5%) | 4,089/4,189 (97.6%) |
| Yes | 56/3,835 (1.5%) | 99/4,189 (2.4%) |
| No data | 2/3,835 (0.1%) | 1/4,189 (0.0%) |
| VIOLATION: (d) xii skin complaints | | |
| No | 3,758/3,835 (98.0%) | 4,120/4,189 (98.4%) |
| Yes | 75/3,835 (2.0%) | 68/4,189 (1.6%) |
| No data | 2/3,835 (0.1%) | 1/4,189 (0.0%) |
| VIOLATION: (d) xiii musculoskeletal system | | |
| No | 3,150/3,835 (82.1%) | 3,504/4,189 (83.6%) |
| Yes | 683/3,835 (17.8%) | 684/4,189 (16.3%) |
| No data | 2/3,835 (0.1%) | 1/4,189 (0.0%) |
| VIOLATION: (d) other complaints | | |
| No | 3,830/3,835 (99.9%) | 4,186/4,189 (99.9%) |
| Yes | 3/3,835 (0.1%) | 2/4,189 (0.0%) |
| No data | 2/3,835 (0.1%) | 1/4,189 (0.0%) |
| VIOLATION: (d) long standing illness | | |
| No | 2,306/3,835 (60.1%) | 2,586/4,189 (61.7%) |
| Yes | 1,527/3,835 (39.8%) | 1,602/4,189 (38.2%) |
| No data | 2/3,835 (0.1%) | 1/4,189 (0.0%) |
| VIOLATION: (d) acute sickness last 2 weeks | | |
| No | 3,343/3,835 (87.2%) | 3,483/4,189 (83.1%) |
| Yes | 491/3,835 (12.8%) | 705/4,189 (16.8%) |
| No data | 1/3,835 (0.0%) | 1/4,189 (0.0%) |
| VIOLATION: been an inpatient in hospital in last 12 months | | |
| No | 3,604/3,835 (94.0%) | 3,829/4,189 (91.4%) |
| Yes | 228/3,835 (5.9%) | 359/4,189 (8.6%) |

| | Overall sample | |
|---|---|---|
| | Male<br>N=3,835 | Female<br>N=4,189 |
| No data | 3/3,835 (0.1%) | 1/4,189 (0.0%) |
| VIOLATION: whether currently pregnant 16 plus | | |
| No | 3,835/3,835 (100.0%) | 4,182/4,189 (99.8%) |
| No data | 0/3,835 (0.0%) | 7/4,189 (0.2%) |
| VIOLATION: pregnant in the last 12 months | | |
| No | 3,835/3,835 (100.0%) | 4,129/4,189 (98.6%) |
| Yes | 0/3,835 (0.0%) | 59/4,189 (1.4%) |
| No data | 0/3,835 (0.0%) | 1/4,189 (0.0%) |
| VIOLATION: (d) whether taking medication - excluding contraceptives only | | |
| No | 2,680/3,835 (69.9%) | 2,489/4,189 (59.4%) |
| Yes | 1,155/3,835 (30.1%) | 1,700/4,189 (40.6%) |
| VIOLATION: Do you currently inject insulin for diabetes? | | |
| No | 3,808/3,835 (99.3%) | 4,171/4,189 (99.6%) |
| Yes | 27/3,835 (0.7%) | 18/4,189 (0.4%) |
| VIOLATION: Are you currently taking any medicines, tablets or pills | | |
| No | 3,793/3,835 (98.9%) | 4,157/4,189 (99.2%) |
| Yes | 42/3,835 (1.1%) | 32/4,189 (0.8%) |
| VIOLATION: whether still on hrt | | |
| No | 3,835/3,835 (100.0%) | 3,664/4,189 (87.5%) |
| Yes | 0/3,835 (0.0%) | 520/4,189 (12.4%) |
| No data | 0/3,835 (0.0%) | 5/4,189 (0.1%) |
| VIOLATION: (d) diuretics (blood pressure) | | |
| No | 3,768/3,835 (98.3%) | 4,057/4,189 (96.8%) |
| Yes | 67/3,835 (1.7%) | 132/4,189 (3.2%) |
| VIOLATION: (d) beta blockers (blood pressure fibrinogen) | | |
| No | 3,731/3,835 (97.3%) | 4,074/4,189 (97.3%) |
| Yes | 104/3,835 (2.7%) | 115/4,189 (2.7%) |
| VIOLATION: (d) ace inhibitors (blood pressure) | | |
| No | 3,729/3,835 (97.2%) | 4,127/4,189 (98.5%) |
| Yes | 106/3,835 (2.8%) | 62/4,189 (1.5%) |
| VIOLATION: (d) calcium blockers (blood pressure) | | |
| No | 3,739/3,835 (97.5%) | 4,125/4,189 (98.5%) |
| Yes | 96/3,835 (2.5%) | 64/4,189 (1.5%) |
| VIOLATION: (d) other drugs affecting bp | | |
| No | 3,820/3,835 (99.6%) | 4,174/4,189 (99.6%) |
| Yes | 15/3,835 (0.4%) | 15/4,189 (0.4%) |
| VIOLATION: (d) lipid lowering (cholesterol fibrinogen) | | |
| No | 3,750/3,835 (97.8%) | 4,134/4,189 (98.7%) |
| Yes | 85/3,835 (2.2%) | 55/4,189 (1.3%) |
| VIOLATION: (d) iron deficiency (haemoglobin ferritin) | | |
| No | 3,835/3,835 (100.0%) | 4,189/4,189 (100.0%) |
| VIOLATION: (d) whether taking drugs affecting blood pressure | | |
| No | 3,559/3,835 (92.8%) | 3,897/4,189 (93.0%) |
| Yes | 276/3,835 (7.2%) | 292/4,189 (7.0%) |
| VIOLATION: (d) whether taking drugs prescribed for blood pressure | | |
| No | 3,632/3,835 (94.7%) | 3,983/4,189 (95.1%) |

| | Overall sample | |
|---|---|---|
| | Male<br>N=3,835 | Female<br>N=4,189 |
| Yes | 203/3,835 (5.3%) | 206/4,189 (4.9%) |
| VIOLATION: (d) units drunk on heaviest day in last 7 | | |
| No | 2,911/3,835 (75.9%) | 3,792/4,189 (90.5%) |
| Yes | 888/3,835 (23.2%) | 381/4,189 (9.1%) |
| No data | 36/3,835 (0.9%) | 16/4,189 (0.4%) |
| VIOLATION: (d) cigarette smoking status – never/ ex-regular/ ex-occasional/ current | | |
| No | 2,687/3,835 (70.1%) | 2,956/4,189 (70.6%) |
| Yes | 1,145/3,835 (29.9%) | 1,232/4,189 (29.4%) |
| No data | 3/3,835 (0.1%) | 1/4,189 (0.0%) |
| VIOLATION: (d) number of cigarettes smoke a day - including non-smokers | | |
| No | 2,705/3,835 (70.5%) | 2,978/4,189 (71.1%) |
| Yes | 1,125/3,835 (29.3%) | 1,207/4,189 (28.8%) |
| No data | 5/3,835 (0.1%) | 4/4,189 (0.1%) |
| VIOLATION: currently smokes cigarettes | | |
| No | 2,704/3,835 (70.5%) | 2,964/4,189 (70.8%) |
| Yes | 1,131/3,835 (29.5%) | 1,225/4,189 (29.2%) |
| VIOLATION: currently smokes cigars | | |
| No | 3,700/3,835 (96.5%) | 4,180/4,189 (99.8%) |
| Yes | 135/3,835 (3.5%) | 9/4,189 (0.2%) |
| VIOLATION: currently smokes pipe | | |
| No | 3,797/3,835 (99.0%) | 4,189/4,189 (100.0%) |
| Yes | 38/3,835 (1.0%) | 0/4,189 (0.0%) |
| VIOLATION: Ferritin<15ug/L or Ferritin>150/200ug/L (F/M) | | |
| No | 3,423/3,835 (89.3%) | 3,558/4,189 (84.9%) |
| Yes | 412/3,835 (10.7%) | 631/4,189 (15.1%) |
| VIOLATION: C-Reactive Protein>5mg/L | | |
| No | 3,437/3,835 (89.6%) | 3,472/4,189 (82.9%) |
| Yes | 398/3,835 (10.4%) | 717/4,189 (17.1%) |

HSE = Health Survey for England, BMI = Body Mass Index, F = Female, M = Male.
Data are presented as unweighted n/total (%) based on participants with non-missing haemoglobin, ferritin, and C-reactive protein.
No data: No answer/refused, Don't know, Not applicable, Not obtained.
[1] Males: ferritin≥15 and ≤200ug/L and C-reactive protein≤5mg/L; females: ferritin≥15 and ≤150ug/L and C-reactive protein≤5mg/L.

#### Table 4.1.10 England HSE 2006: Participant characteristics of violations (18-65 years)

| | Overall sample | |
|---|---|---|
| | Male N=2,569 | Female N=3,026 |
| Reference sample[1] | | |
| No | 2,195/2,569 (85.4%) | 2,511/3,026 (83.0%) |
| Yes | 374/2,569 (14.6%) | 515/3,026 (17.0%) |
| VIOLATION: BMI<18.5 or >30 kg/m$^2$ | | |
| No | 1,820/2,569 (70.8%) | 2,162/3,026 (71.4%) |
| Yes | 614/2,569 (23.9%) | 682/3,026 (22.5%) |
| No data | 135/2,569 (5.3%) | 182/3,026 (6.0%) |
| VIOLATION: whether has clotting disorder | | |
| No | 2,569/2,569 (100.0%) | 3,026/3,026 (100.0%) |
| VIOLATION: (d) had cardiovascular condition | | |
| No | 1,843/2,569 (71.7%) | 2,269/3,026 (75.0%) |
| Yes | 725/2,569 (28.2%) | 752/3,026 (24.9%) |
| No data | 1/2,569 (0.0%) | 5/3,026 (0.2%) |
| VIOLATION: (d) had cvd: excludes those with high bp | | |
| No | 2,267/2,569 (88.2%) | 2,706/3,026 (89.4%) |
| Yes | 301/2,569 (11.7%) | 317/3,026 (10.5%) |
| No data | 1/2,569 (0.0%) | 3/3,026 (0.1%) |
| VIOLATION: (d) had ihd (angina or heart attack) | | |
| No | 2,486/2,569 (96.8%) | 3,001/3,026 (99.2%) |
| Yes | 83/2,569 (3.2%) | 25/3,026 (0.8%) |
| VIOLATION: (d) had cvd (angina, heart attack or stroke) | | |
| No | 2,477/2,569 (96.4%) | 2,982/3,026 (98.5%) |
| Yes | 92/2,569 (3.6%) | 44/3,026 (1.5%) |
| VIOLATION: (d) angina or mi (rose angina qure) | | |
| No | 2,330/2,569 (90.7%) | 2,835/3,026 (93.7%) |
| Yes | 237/2,569 (9.2%) | 187/3,026 (6.2%) |
| No data | 2/2,569 (0.1%) | 4/3,026 (0.1%) |
| VIOLATION: (d) angina symptoms (rose angina qure) | | |
| No | 2,532/2,569 (98.6%) | 2,962/3,026 (97.9%) |
| Yes | 36/2,569 (1.4%) | 60/3,026 (2.0%) |
| No data | 1/2,569 (0.0%) | 4/3,026 (0.1%) |
| VIOLATION: (d) possible infarction (rose angina qure) | | |
| No | 2,353/2,569 (91.6%) | 2,884/3,026 (95.3%) |
| Yes | 215/2,569 (8.4%) | 142/3,026 (4.7%) |
| No data | 1/2,569 (0.0%) | 0/3,026 (0.0%) |
| VIOLATION: (d) doctor diagnosed angina | | |
| No | 2,509/2,569 (97.7%) | 3,007/3,026 (99.4%) |
| Yes | 60/2,569 (2.3%) | 19/3,026 (0.6%) |
| VIOLATION: (d) doctor diagnosed high blood pressure (excluding pregnant) | | |
| No | 2,006/2,569 (78.1%) | 2,453/3,026 (81.1%) |
| Yes | 563/2,569 (21.9%) | 571/3,026 (18.9%) |
| No data | 0/2,569 (0.0%) | 2/3,026 (0.1%) |
| VIOLATION: (d) doctor diagnosed heart attack | | |
| No | 2,523/2,569 (98.2%) | 3,012/3,026 (99.5%) |
| Yes | 46/2,569 (1.8%) | 14/3,026 (0.5%) |
| VIOLATION: (d) doctor diagnosed stroke | | |
| No | 2,554/2,569 (99.4%) | 3,003/3,026 (99.2%) |

| | Overall sample | |
|---|---|---|
| | Male<br>N=2,569 | Female<br>N=3,026 |
| Yes | 15/2,569 (0.6%) | 23/3,026 (0.8%) |
| VIOLATION: (d) doctor diagnosed diabetes (excluding pregnant) | | |
| No | 2,471/2,569 (96.2%) | 2,965/3,026 (98.0%) |
| Yes | 98/2,569 (3.8%) | 61/3,026 (2.0%) |
| VIOLATION: (d) doctor diagnosed heart murmur (excluding pregnant) | | |
| No | 2,521/2,569 (98.1%) | 2,929/3,026 (96.8%) |
| Yes | 48/2,569 (1.9%) | 94/3,026 (3.1%) |
| No data | 0/2,569 (0.0%) | 3/3,026 (0.1%) |
| VIOLATION: (d) doctor diagnosed irregular heart rhythm | | |
| No | 2,464/2,569 (95.9%) | 2,891/3,026 (95.5%) |
| Yes | 104/2,569 (4.0%) | 135/3,026 (4.5%) |
| No data | 1/2,569 (0.0%) | 0/3,026 (0.0%) |
| VIOLATION: (d) doctor diagnosed other heart condition | | |
| No | 2,543/2,569 (99.0%) | 2,990/3,026 (98.8%) |
| Yes | 26/2,569 (1.0%) | 36/3,026 (1.2%) |
| VIOLATION: surgery for heart condition | | |
| No | 2,522/2,569 (98.2%) | 3,010/3,026 (99.5%) |
| Yes | 47/2,569 (1.8%) | 16/3,026 (0.5%) |
| VIOLATION: any surgery for heart murmur | | |
| No | 2,567/2,569 (99.9%) | 3,024/3,026 (99.9%) |
| Yes | 2/2,569 (0.1%) | 2/3,026 (0.1%) |
| VIOLATION: (d) i infectious disease | | |
| No | 2,565/2,569 (99.8%) | 3,021/3,026 (99.8%) |
| Yes | 3/2,569 (0.1%) | 4/3,026 (0.1%) |
| No data | 1/2,569 (0.0%) | 1/3,026 (0.0%) |
| VIOLATION: (d) ii neoplasms & benign growths | | |
| No | 2,543/2,569 (99.0%) | 2,985/3,026 (98.6%) |
| Yes | 25/2,569 (1.0%) | 40/3,026 (1.3%) |
| No data | 1/2,569 (0.0%) | 1/3,026 (0.0%) |
| VIOLATION: (d) iii endocrine & metabolic | | |
| No | 2,436/2,569 (94.8%) | 2,853/3,026 (94.3%) |
| Yes | 132/2,569 (5.1%) | 172/3,026 (5.7%) |
| No data | 1/2,569 (0.0%) | 1/3,026 (0.0%) |
| VIOLATION: (d) iv blood and related organs | | |
| No | 2,562/2,569 (99.7%) | 2,999/3,026 (99.1%) |
| Yes | 6/2,569 (0.2%) | 26/3,026 (0.9%) |
| No data | 1/2,569 (0.0%) | 1/3,026 (0.0%) |
| VIOLATION: (d) v mental disorders | | |
| No | 2,486/2,569 (96.8%) | 2,887/3,026 (95.4%) |
| Yes | 82/2,569 (3.2%) | 138/3,026 (4.6%) |
| No data | 1/2,569 (0.0%) | 1/3,026 (0.0%) |
| VIOLATION: (d) vi nervous system | | |
| No | 2,491/2,569 (97.0%) | 2,893/3,026 (95.6%) |
| Yes | 77/2,569 (3.0%) | 132/3,026 (4.4%) |
| No data | 1/2,569 (0.0%) | 1/3,026 (0.0%) |
| VIOLATION: (d) vi eye complaints | | |
| No | 2,530/2,569 (98.5%) | 2,995/3,026 (99.0%) |
| Yes | 38/2,569 (1.5%) | 30/3,026 (1.0%) |

| | Overall sample | |
|---|---|---|
| | Male<br>N=2,569 | Female<br>N=3,026 |
| No data | 1/2,569 (0.0%) | 1/3,026 (0.0%) |
| VIOLATION: (d) vi ear complaints | | |
| No | 2,519/2,569 (98.1%) | 2,977/3,026 (98.4%) |
| Yes | 49/2,569 (1.9%) | 48/3,026 (1.6%) |
| No data | 1/2,569 (0.0%) | 1/3,026 (0.0%) |
| VIOLATION: (d) vii heart and circulatory system | | |
| No | 2,351/2,569 (91.5%) | 2,797/3,026 (92.4%) |
| Yes | 217/2,569 (8.4%) | 228/3,026 (7.5%) |
| No data | 1/2,569 (0.0%) | 1/3,026 (0.0%) |
| VIOLATION: (d) respiratory system | | |
| No | 2,359/2,569 (91.8%) | 2,757/3,026 (91.1%) |
| Yes | 209/2,569 (8.1%) | 268/3,026 (8.9%) |
| No data | 1/2,569 (0.0%) | 1/3,026 (0.0%) |
| VIOLATION: (d) ix digestive system | | |
| No | 2,453/2,569 (95.5%) | 2,886/3,026 (95.4%) |
| Yes | 115/2,569 (4.5%) | 139/3,026 (4.6%) |
| No data | 1/2,569 (0.0%) | 1/3,026 (0.0%) |
| VIOLATION: (d) x genito-urinary system | | |
| No | 2,530/2,569 (98.5%) | 2,945/3,026 (97.3%) |
| Yes | 38/2,569 (1.5%) | 80/3,026 (2.6%) |
| No data | 1/2,569 (0.0%) | 1/3,026 (0.0%) |
| VIOLATION: (d) xii skin complaints | | |
| No | 2,524/2,569 (98.2%) | 2,970/3,026 (98.1%) |
| Yes | 44/2,569 (1.7%) | 55/3,026 (1.8%) |
| No data | 1/2,569 (0.0%) | 1/3,026 (0.0%) |
| VIOLATION: (d) xiii musculoskeletal system | | |
| No | 2,116/2,569 (82.4%) | 2,540/3,026 (83.9%) |
| Yes | 452/2,569 (17.6%) | 485/3,026 (16.0%) |
| No data | 1/2,569 (0.0%) | 1/3,026 (0.0%) |
| VIOLATION: (d) other complaints | | |
| No | 2,564/2,569 (99.8%) | 3,014/3,026 (99.6%) |
| Yes | 4/2,569 (0.2%) | 11/3,026 (0.4%) |
| No data | 1/2,569 (0.0%) | 1/3,026 (0.0%) |
| VIOLATION: (d) long standing illness | | |
| No | 1,550/2,569 (60.3%) | 1,790/3,026 (59.2%) |
| Yes | 1,018/2,569 (39.6%) | 1,235/3,026 (40.8%) |
| No data | 1/2,569 (0.0%) | 1/3,026 (0.0%) |
| VIOLATION: been an inpatient in hospital in last 12 months | | |
| No | 2,498/2,569 (97.2%) | 2,889/3,026 (95.5%) |
| Yes | 71/2,569 (2.8%) | 137/3,026 (4.5%) |
| VIOLATION: (d) acute sickness last 2 weeks | | |
| No | 2,202/2,569 (85.7%) | 2,477/3,026 (81.9%) |
| Yes | 367/2,569 (14.3%) | 548/3,026 (18.1%) |
| No data | 0/2,569 (0.0%) | 1/3,026 (0.0%) |
| VIOLATION: how many times fractured bones in the last 12 months | | |
| No | 2,559/2,569 (99.6%) | 3,019/3,026 (99.8%) |
| Yes | 10/2,569 (0.4%) | 7/3,026 (0.2%) |
| VIOLATION: whether currently pregnant 16 plus | | |

| | Overall sample | |
|---|---|---|
| | Male<br>N=2,569 | Female<br>N=3,026 |
| No | 2,569/2,569 (100.0%) | 3,026/3,026 (100.0%) |
| VIOLATION: pregnant in the last 12months | | |
| No | 2,569/2,569 (100.0%) | 2,982/3,026 (98.5%) |
| Yes | 0/2,569 (0.0%) | 43/3,026 (1.4%) |
| No data | 0/2,569 (0.0%) | 1/3,026 (0.0%) |
| VIOLATION: (d) whether taking medication - excluding contraceptives only | | |
| No | 1,681/2,569 (65.4%) | 1,777/3,026 (58.7%) |
| Yes | 888/2,569 (34.6%) | 1,249/3,026 (41.3%) |
| VIOLATION: Do you currently inject insulin for diabetes? | | |
| No | 2,538/2,569 (98.8%) | 3,005/3,026 (99.3%) |
| Yes | 31/2,569 (1.2%) | 21/3,026 (0.7%) |
| VIOLATION: Are you currently taking any medicines, tablets or pills | | |
| No | 2,504/2,569 (97.5%) | 2,988/3,026 (98.7%) |
| Yes | 65/2,569 (2.5%) | 38/3,026 (1.3%) |
| VIOLATION: whether still on hrt | | |
| No | 2,569/2,569 (100.0%) | 2,860/3,026 (94.5%) |
| Yes | 0/2,569 (0.0%) | 165/3,026 (5.5%) |
| No data | 0/2,569 (0.0%) | 1/3,026 (0.0%) |
| VIOLATION: (d) diuretics (blood pressure) | | |
| No | 2,487/2,569 (96.8%) | 2,900/3,026 (95.8%) |
| Yes | 82/2,569 (3.2%) | 126/3,026 (4.2%) |
| VIOLATION: (d) beta blockers (blood pressure fibrinogen) | | |
| No | 2,460/2,569 (95.8%) | 2,910/3,026 (96.2%) |
| Yes | 109/2,569 (4.2%) | 116/3,026 (3.8%) |
| VIOLATION: (d) ace inhibitors (blood pressure) | | |
| No | 2,385/2,569 (92.8%) | 2,885/3,026 (95.3%) |
| Yes | 184/2,569 (7.2%) | 141/3,026 (4.7%) |
| VIOLATION: (d) calcium blockers (blood pressure) | | |
| No | 2,479/2,569 (96.5%) | 2,949/3,026 (97.5%) |
| Yes | 90/2,569 (3.5%) | 77/3,026 (2.5%) |
| VIOLATION: (d) other drugs affecting bp | | |
| No | 2,541/2,569 (98.9%) | 3,004/3,026 (99.3%) |
| Yes | 28/2,569 (1.1%) | 22/3,026 (0.7%) |
| VIOLATION: (d) lipid lowering (cholesterol fibrinogen) | | |
| No | 2,352/2,569 (91.6%) | 2,887/3,026 (95.4%) |
| Yes | 217/2,569 (8.4%) | 139/3,026 (4.6%) |
| VIOLATION: (d) iron deficiency (haemoglobin ferritin) | | |
| No | 2,569/2,569 (100.0%) | 3,026/3,026 (100.0%) |
| VIOLATION: (d) whether taking drugs affecting blood pressure | | |
| No | 2,268/2,569 (88.3%) | 2,730/3,026 (90.2%) |
| Yes | 301/2,569 (11.7%) | 296/3,026 (9.8%) |
| VIOLATION: (d) whether taking drugs prescribed for blood pressure | | |
| No | 2,342/2,569 (91.2%) | 2,785/3,026 (92.0%) |
| Yes | 227/2,569 (8.8%) | 241/3,026 (8.0%) |
| VIOLATION: are you taking statins (drugs to lower cholesterol) | | |
| No | 2,558/2,569 (99.6%) | 3,013/3,026 (99.6%) |

| | Overall sample | |
|---|---|---|
| | Male<br>N=2,569 | Female<br>N=3,026 |
| Yes | 11/2,569 (0.4%) | 13/3,026 (0.4%) |
| VIOLATION: (d) units drunk on heaviest day in last 7 | | |
| No | 1,973/2,569 (76.8%) | 2,717/3,026 (89.8%) |
| Yes | 593/2,569 (23.1%) | 307/3,026 (10.1%) |
| No data | 3/2,569 (0.1%) | 2/3,026 (0.1%) |
| VIOLATION: (d) cigarette smoking status – never/ ex-regular/ ex-occasional/ current | | |
| No | 1,956/2,569 (76.1%) | 2,315/3,026 (76.5%) |
| Yes | 613/2,569 (23.9%) | 709/3,026 (23.4%) |
| No data | 0/2,569 (0.0%) | 2/3,026 (0.1%) |
| VIOLATION: (d) number of cigarettes smoke a day - including non-smokers | | |
| No | 1,974/2,569 (76.8%) | 2,334/3,026 (77.1%) |
| Yes | 590/2,569 (23.0%) | 689/3,026 (22.8%) |
| No data | 5/2,569 (0.2%) | 3/3,026 (0.1%) |
| VIOLATION: currently smokes cigarettes | | |
| No | 1,959/2,569 (76.3%) | 2,332/3,026 (77.1%) |
| Yes | 610/2,569 (23.7%) | 694/3,026 (22.9%) |
| VIOLATION: currently smokes cigars | | |
| No | 2,522/2,569 (98.2%) | 3,024/3,026 (99.9%) |
| Yes | 47/2,569 (1.8%) | 2/3,026 (0.1%) |
| VIOLATION: currently smokes pipe | | |
| No | 2,558/2,569 (99.6%) | 3,026/3,026 (100.0%) |
| Yes | 11/2,569 (0.4%) | 0/3,026 (0.0%) |
| VIOLATION: Ferritin<15ug/L or Ferritin>150/200ug/L (F/M) | | |
| No | 2,007/2,569 (78.1%) | 2,556/3,026 (84.5%) |
| Yes | 562/2,569 (21.9%) | 470/3,026 (15.5%) |
| VIOLATION: C-Reactive Protein>5mg/L | | |
| No | 2,269/2,569 (88.3%) | 2,513/3,026 (83.0%) |
| Yes | 300/2,569 (11.7%) | 513/3,026 (17.0%) |

HSE = Health Survey for England, BMI = Body Mass Index, F = Female, M = Male.
Data are presented as unweighted n/total (%) based on participants with non-missing haemoglobin, ferritin, and C-reactive protein.
No data: No answer/refused, Don't know, Not applicable, Not obtained.
[1] Males: ferritin≥15 and ≤200ug/L and C-reactive protein≤5mg/L; females: ferritin≥15 and ≤150ug/L and C-reactive protein≤5mg/L.

#### Table 4.1.11 England HSE 2009: Participant characteristics of violations (18-65 years)

| | Overall sample | |
|---|---|---|
| | Male<br>N=804 | Female<br>N=892 |
| Reference sample[1] | | |
| No | 686/804 (85.3%) | 750/892 (84.1%) |
| Yes | 118/804 (14.7%) | 142/892 (15.9%) |
| VIOLATION: BMI<18.5 or >30 kg/m$^2$ | | |
| No | 557/804 (69.3%) | 624/892 (70.0%) |
| Yes | 193/804 (24.0%) | 216/892 (24.2%) |
| No data | 54/804 (6.7%) | 52/892 (5.8%) |
| VIOLATION: whether has clotting disorder | | |
| No | 804/804 (100.0%) | 892/892 (100.0%) |
| VIOLATION: (d) doctor diagnosed high blood pressure (excluding pregnant) | | |
| No | 634/804 (78.9%) | 764/892 (85.7%) |
| Yes | 170/804 (21.1%) | 128/892 (14.3%) |
| VIOLATION: (d) doctor diagnosed diabetes (excluding pregnant) | | |
| No | 776/804 (96.5%) | 871/892 (97.6%) |
| Yes | 28/804 (3.5%) | 21/892 (2.4%) |
| VIOLATION: (d) i infectious disease | | |
| No | 803/804 (99.9%) | 891/892 (99.9%) |
| Yes | 0/804 (0.0%) | 1/892 (0.1%) |
| No data | 1/804 (0.1%) | 0/892 (0.0%) |
| VIOLATION: (d) ii neoplasms & benign growths | | |
| No | 795/804 (98.9%) | 883/892 (99.0%) |
| Yes | 8/804 (1.0%) | 9/892 (1.0%) |
| No data | 1/804 (0.1%) | 0/892 (0.0%) |
| VIOLATION: (d) iii endocrine & metabolic | | |
| No | 760/804 (94.5%) | 828/892 (92.8%) |
| Yes | 43/804 (5.3%) | 64/892 (7.2%) |
| No data | 1/804 (0.1%) | 0/892 (0.0%) |
| VIOLATION: (d) iv blood and related organs | | |
| No | 802/804 (99.8%) | 885/892 (99.2%) |
| Yes | 1/804 (0.1%) | 7/892 (0.8%) |
| No data | 1/804 (0.1%) | 0/892 (0.0%) |
| VIOLATION: (d) v mental disorders | | |
| No | 777/804 (96.6%) | 855/892 (95.9%) |
| Yes | 26/804 (3.2%) | 37/892 (4.1%) |
| No data | 1/804 (0.1%) | 0/892 (0.0%) |
| VIOLATION: (d) vi nervous system | | |
| No | 781/804 (97.1%) | 848/892 (95.1%) |
| Yes | 22/804 (2.7%) | 44/892 (4.9%) |
| No data | 1/804 (0.1%) | 0/892 (0.0%) |
| VIOLATION: (d) vi eye complaints | | |
| No | 794/804 (98.8%) | 885/892 (99.2%) |
| Yes | 9/804 (1.1%) | 7/892 (0.8%) |
| No data | 1/804 (0.1%) | 0/892 (0.0%) |
| VIOLATION: (d) vi ear complaints | | |
| No | 793/804 (98.6%) | 881/892 (98.8%) |
| Yes | 10/804 (1.2%) | 11/892 (1.2%) |
| No data | 1/804 (0.1%) | 0/892 (0.0%) |

| | Overall sample | |
|---|---|---|
| | Male<br>N=804 | Female<br>N=892 |
| VIOLATION: (d) vii heart and circulatory system | | |
| No | 720/804 (89.6%) | 840/892 (94.2%) |
| Yes | 83/804 (10.3%) | 52/892 (5.8%) |
| No data | 1/804 (0.1%) | 0/892 (0.0%) |
| VIOLATION: (d) respiratory system | | |
| No | 755/804 (93.9%) | 828/892 (92.8%) |
| Yes | 48/804 (6.0%) | 64/892 (7.2%) |
| No data | 1/804 (0.1%) | 0/892 (0.0%) |
| VIOLATION: (d) ix digestive system | | |
| No | 772/804 (96.0%) | 849/892 (95.2%) |
| Yes | 31/804 (3.9%) | 43/892 (4.8%) |
| No data | 1/804 (0.1%) | 0/892 (0.0%) |
| VIOLATION: (d) x genito-urinary system | | |
| No | 794/804 (98.8%) | 872/892 (97.8%) |
| Yes | 9/804 (1.1%) | 20/892 (2.2%) |
| No data | 1/804 (0.1%) | 0/892 (0.0%) |
| VIOLATION: (d) xii skin complaints | | |
| No | 790/804 (98.3%) | 880/892 (98.7%) |
| Yes | 13/804 (1.6%) | 12/892 (1.3%) |
| No data | 1/804 (0.1%) | 0/892 (0.0%) |
| VIOLATION: (d) xiii musculoskeletal system | | |
| No | 684/804 (85.1%) | 767/892 (86.0%) |
| Yes | 119/804 (14.8%) | 125/892 (14.0%) |
| No data | 1/804 (0.1%) | 0/892 (0.0%) |
| VIOLATION: (d) other complaints | | |
| No | 800/804 (99.5%) | 891/892 (99.9%) |
| Yes | 3/804 (0.4%) | 1/892 (0.1%) |
| No data | 1/804 (0.1%) | 0/892 (0.0%) |
| VIOLATION: (d) long standing illness | | |
| No | 509/804 (63.3%) | 572/892 (64.1%) |
| Yes | 294/804 (36.6%) | 320/892 (35.9%) |
| No data | 1/804 (0.1%) | 0/892 (0.0%) |
| VIOLATION: (d) acute sickness last 2 weeks | | |
| No | 701/804 (87.2%) | 754/892 (84.5%) |
| Yes | 103/804 (12.8%) | 138/892 (15.5%) |
| VIOLATION: (d) whether taking medication - excluding contraceptives only | | |
| No | 530/804 (65.9%) | 511/892 (57.3%) |
| Yes | 274/804 (34.1%) | 381/892 (42.7%) |
| VIOLATION: Do you currently inject insulin for diabetes? | | |
| No | 796/804 (99.0%) | 885/892 (99.2%) |
| Yes | 8/804 (1.0%) | 7/892 (0.8%) |
| VIOLATION: Are you currently taking any medicines, tablets or pills | | |
| No | 784/804 (97.5%) | 878/892 (98.4%) |
| Yes | 20/804 (2.5%) | 14/892 (1.6%) |
| VIOLATION: (d) diuretics (blood pressure) | | |
| No | 773/804 (96.1%) | 867/892 (97.2%) |
| Yes | 31/804 (3.9%) | 25/892 (2.8%) |
| VIOLATION: (d) beta blockers (blood pressure fibrinogen) | | |

| | Overall sample | |
|---|---|---|
| | Male<br>N=804 | Female<br>N=892 |
| No | 777/804 (96.6%) | 869/892 (97.4%) |
| Yes | 27/804 (3.4%) | 23/892 (2.6%) |
| VIOLATION: (d) ace inhibitors (blood pressure) | | |
| No | 730/804 (90.8%) | 845/892 (94.7%) |
| Yes | 74/804 (9.2%) | 47/892 (5.3%) |
| VIOLATION: (d) calcium blockers (blood pressure) | | |
| No | 762/804 (94.8%) | 881/892 (98.8%) |
| Yes | 42/804 (5.2%) | 11/892 (1.2%) |
| VIOLATION: (d) other drugs affecting bp | | |
| No | 795/804 (98.9%) | 887/892 (99.4%) |
| Yes | 9/804 (1.1%) | 5/892 (0.6%) |
| VIOLATION: (d) lipid lowering (cholesterol fibrinogen) | | |
| No | 729/804 (90.7%) | 847/892 (95.0%) |
| Yes | 75/804 (9.3%) | 45/892 (5.0%) |
| VIOLATION: (d) iron deficiency (haemoglobin ferritin) | | |
| No | 804/804 (100.0%) | 892/892 (100.0%) |
| VIOLATION: (d) whether taking drugs affecting blood pressure | | |
| No | 689/804 (85.7%) | 814/892 (91.3%) |
| Yes | 115/804 (14.3%) | 78/892 (8.7%) |
| VIOLATION: (d) whether taking drugs prescribed for blood pressure | | |
| No | 714/804 (88.8%) | 827/892 (92.7%) |
| Yes | 90/804 (11.2%) | 65/892 (7.3%) |
| VIOLATION: are you taking statins (drugs to lower cholesterol) | | |
| No | 797/804 (99.1%) | 885/892 (99.2%) |
| Yes | 7/804 (0.9%) | 7/892 (0.8%) |
| VIOLATION: (D) NEW Units drunk on heaviest day in last 7 (16yrs+) | | |
| No | 559/804 (69.5%) | 734/892 (82.3%) |
| Yes | 243/804 (30.2%) | 154/892 (17.3%) |
| No data | 2/804 (0.2%) | 4/892 (0.4%) |
| VIOLATION: (d) cigarette smoking status – never/ ex-regular/ ex-occasional/ current | | |
| No | 628/804 (78.1%) | 707/892 (79.3%) |
| Yes | 176/804 (21.9%) | 185/892 (20.7%) |
| VIOLATION: (d) number of cigarettes smoke a day - including non-smokers | | |
| No | 631/804 (78.5%) | 709/892 (79.5%) |
| Yes | 172/804 (21.4%) | 183/892 (20.5%) |
| No data | 1/804 (0.1%) | 0/892 (0.0%) |
| VIOLATION: currently smokes cigarettes | | |
| No | 627/804 (78.0%) | 709/892 (79.5%) |
| Yes | 177/804 (22.0%) | 183/892 (20.5%) |
| VIOLATION: currently smokes cigars | | |
| No | 786/804 (97.8%) | 892/892 (100.0%) |
| Yes | 18/804 (2.2%) | 0/892 (0.0%) |
| VIOLATION: currently smokes pipe | | |
| No | 803/804 (99.9%) | 892/892 (100.0%) |
| Yes | 1/804 (0.1%) | 0/892 (0.0%) |
| VIOLATION: Ferritin<15ug/L or Ferritin>150/200ug/L (F/M) | | |
| No | 629/804 (78.2%) | 745/892 (83.5%) |

| | Overall sample | |
| --- | --- | --- |
| | Male<br>N=804 | Female<br>N=892 |
| Yes | 175/804 (21.8%) | 147/892 (16.5%) |
| VIOLATION: C-Reactive Protein>5mg/L | | |
| No | 728/804 (90.5%) | 734/892 (82.3%) |
| Yes | 76/804 (9.5%) | 158/892 (17.7%) |
| VIOLATION: eGFR<90 ml/min/1.73m$^2$ | | |
| No | 606/804 (75.4%) | 677/892 (75.9%) |
| Yes | 163/804 (20.3%) | 175/892 (19.6%) |
| No data | 35/804 (4.4%) | 40/892 (4.5%) |

HSE = Health Survey for England, BMI = Body Mass Index, F = Female, M = Male.
Data are presented as unweighted n/total (%) based on participants with non-missing haemoglobin, ferritin, and C-reactive protein.
No data: No answer/refused, Don't know, Not applicable, Not obtained.
[1] Males: ferritin≥15 and ≤200ug/L and C-reactive protein≤5mg/L; females: ferritin≥15 and ≤150ug/L and C-reactive protein≤5mg/L.

#### Table 4.1.12 CHN CHNS 2009: Participant characteristics of violations (18-65 years)

| | Overall sample | |
|---|---|---|
| | **Male** | **Female** |
| | **N=3,352** | **N=3,820** |
| Reference sample[1] | | |
| No | 3,107/3,352 (92.7%) | 3,254/3,820 (85.2%) |
| Yes | 245/3,352 (7.3%) | 566/3,820 (14.8%) |
| VIOLATION: BMI<18.5 or >30 kg/m$^2$ | | |
| No | 2,967/3,352 (88.5%) | 3,382/3,820 (88.5%) |
| Yes | 298/3,352 (8.9%) | 376/3,820 (9.8%) |
| Missing | 87/3,352 (2.6%) | 62/3,820 (1.6%) |
| VIOLATION: Illness/Injury - Infectious/parasitic disease | | |
| No | 342/3,352 (10.2%) | 483/3,820 (12.6%) |
| Yes | 2/3,352 (0.1%) | 1/3,820 (0.0%) |
| Missing | 3,008/3,352 (89.7%) | 3,336/3,820 (87.3%) |
| VIOLATION: Illness/Injury - Heart disease | | |
| No | 331/3,352 (9.9%) | 459/3,820 (12.0%) |
| Yes | 13/3,352 (0.4%) | 25/3,820 (0.7%) |
| Missing | 3,008/3,352 (89.7%) | 3,336/3,820 (87.3%) |
| VIOLATION: Illness/Injury - Tumor | | |
| No | 342/3,352 (10.2%) | 482/3,820 (12.6%) |
| Yes | 2/3,352 (0.1%) | 2/3,820 (0.1%) |
| Missing | 3,008/3,352 (89.7%) | 3,336/3,820 (87.3%) |
| VIOLATION: Illness/Injury - Respiratory disease | | |
| No | 244/3,352 (7.3%) | 355/3,820 (9.3%) |
| Yes | 100/3,352 (3.0%) | 129/3,820 (3.4%) |
| Missing | 3,008/3,352 (89.7%) | 3,336/3,820 (87.3%) |
| VIOLATION: Illness/Injury - Injury | | |
| No | 340/3,352 (10.1%) | 477/3,820 (12.5%) |
| Yes | 4/3,352 (0.1%) | 7/3,820 (0.2%) |
| Missing | 3,008/3,352 (89.7%) | 3,336/3,820 (87.3%) |
| VIOLATION: Illness/Injury - Alcohol poisoning | | |
| No | 343/3,352 (10.2%) | 484/3,820 (12.7%) |
| Yes | 1/3,352 (0.0%) | 0/3,820 (0.0%) |
| Missing | 3,008/3,352 (89.7%) | 3,336/3,820 (87.3%) |
| VIOLATION: Illness/Injury - Endocrine disorder | | |
| No | 337/3,352 (10.1%) | 477/3,820 (12.5%) |
| Yes | 7/3,352 (0.2%) | 7/3,820 (0.2%) |
| Missing | 3,008/3,352 (89.7%) | 3,336/3,820 (87.3%) |
| VIOLATION: Illness/Injury - Hematological disease | | |
| No | 341/3,352 (10.2%) | 479/3,820 (12.5%) |
| Yes | 3/3,352 (0.1%) | 5/3,820 (0.1%) |
| Missing | 3,008/3,352 (89.7%) | 3,336/3,820 (87.3%) |
| VIOLATION: Illness/Injury - Mental/psychiatric disorder | | |
| No | 342/3,352 (10.2%) | 482/3,820 (12.6%) |
| Yes | 2/3,352 (0.1%) | 2/3,820 (0.1%) |
| Missing | 3,008/3,352 (89.7%) | 3,336/3,820 (87.3%) |
| VIOLATION: Illness/Injury - Neurological disorder | | |
| No | 330/3,352 (9.8%) | 466/3,820 (12.2%) |
| Yes | 14/3,352 (0.4%) | 18/3,820 (0.5%) |
| Missing | 3,008/3,352 (89.7%) | 3,336/3,820 (87.3%) |

| | Overall sample | |
|---|---|---|
| | Male<br>N=3,352 | Female<br>N=3,820 |
| VIOLATION: Illness/Injury - Eye/ear/nose/throat/teeth disease | | |
| No | 337/3,352 (10.1%) | 469/3,820 (12.3%) |
| Yes | 7/3,352 (0.2%) | 15/3,820 (0.4%) |
| Missing | 3,008/3,352 (89.7%) | 3,336/3,820 (87.3%) |
| VIOLATION: Illness/Injury - Digestive disease | | |
| No | 296/3,352 (8.8%) | 440/3,820 (11.5%) |
| Yes | 48/3,352 (1.4%) | 44/3,820 (1.2%) |
| Missing | 3,008/3,352 (89.7%) | 3,336/3,820 (87.3%) |
| VIOLATION: Illness/Injury - Urinary disease | | |
| No | 335/3,352 (10.0%) | 476/3,820 (12.5%) |
| Yes | 9/3,352 (0.3%) | 8/3,820 (0.2%) |
| Missing | 3,008/3,352 (89.7%) | 3,336/3,820 (87.3%) |
| VIOLATION: Illness/Injury - Sexual dysfunction | | |
| No | 343/3,352 (10.2%) | 484/3,820 (12.7%) |
| Yes | 1/3,352 (0.0%) | 0/3,820 (0.0%) |
| Missing | 3,008/3,352 (89.7%) | 3,336/3,820 (87.3%) |
| VIOLATION: Illness/Injury - Obstetrical/gynecological disease | | |
| No | 344/3,352 (10.3%) | 466/3,820 (12.2%) |
| Yes | 0/3,352 (0.0%) | 18/3,820 (0.5%) |
| Missing | 3,008/3,352 (89.7%) | 3,336/3,820 (87.3%) |
| VIOLATION: Illness/Injury - Dermatological disease | | |
| No | 336/3,352 (10.0%) | 477/3,820 (12.5%) |
| Yes | 8/3,352 (0.2%) | 7/3,820 (0.2%) |
| Missing | 3,008/3,352 (89.7%) | 3,336/3,820 (87.3%) |
| VIOLATION: Illness/Injury - Muscular/rheumatological disease | | |
| No | 328/3,352 (9.8%) | 461/3,820 (12.1%) |
| Yes | 16/3,352 (0.5%) | 23/3,820 (0.6%) |
| Missing | 3,008/3,352 (89.7%) | 3,336/3,820 (87.3%) |
| VIOLATION: Diagnosed with high blood pressure | | |
| No | 3,013/3,352 (89.9%) | 3,422/3,820 (89.6%) |
| Yes | 317/3,352 (9.5%) | 381/3,820 (10.0%) |
| No data | 9/3,352 (0.3%) | 7/3,820 (0.2%) |
| Missing | 13/3,352 (0.4%) | 10/3,820 (0.3%) |
| VIOLATION: Diagnosed with diabetes | | |
| No | 3,244/3,352 (96.8%) | 3,735/3,820 (97.8%) |
| Yes | 90/3,352 (2.7%) | 67/3,820 (1.8%) |
| No data | 5/3,352 (0.1%) | 8/3,820 (0.2%) |
| Missing | 13/3,352 (0.4%) | 10/3,820 (0.3%) |
| VIOLATION: Diagnosed with myocardial infarction | | |
| No | 3,319/3,352 (99.0%) | 3,777/3,820 (98.9%) |
| Yes | 19/3,352 (0.6%) | 26/3,820 (0.7%) |
| No data | 1/3,352 (0.0%) | 5/3,820 (0.1%) |
| Missing | 13/3,352 (0.4%) | 12/3,820 (0.3%) |
| VIOLATION: Diagnosed with apoplexy | | |
| No | 3,301/3,352 (98.5%) | 3,786/3,820 (99.1%) |
| Yes | 37/3,352 (1.1%) | 18/3,820 (0.5%) |
| No data | 1/3,352 (0.0%) | 4/3,820 (0.1%) |
| Missing | 13/3,352 (0.4%) | 12/3,820 (0.3%) |

| | Overall sample | |
|---|---|---|
| | Male N=3,352 | Female N=3,820 |
| VIOLATION: Dr. told you that you suffer from asthma | | |
| No | 3,306/3,352 (98.6%) | 3,780/3,820 (99.0%) |
| Yes | 30/3,352 (0.9%) | 26/3,820 (0.7%) |
| No data | 1/3,352 (0.0%) | 2/3,820 (0.1%) |
| Missing | 15/3,352 (0.4%) | 12/3,820 (0.3%) |
| VIOLATION: Past 12 months - had whistling/wheezing in chest | | |
| No | 9/3,352 (0.3%) | 10/3,820 (0.3%) |
| Yes | 20/3,352 (0.6%) | 15/3,820 (0.4%) |
| No data | 0/3,352 (0.0%) | 1/3,820 (0.0%) |
| Missing | 3,323/3,352 (99.1%) | 3,794/3,820 (99.3%) |
| VIOLATION: Last 4 weeks - been sick or injured | | |
| No | 2,932/3,352 (87.5%) | 3,260/3,820 (85.3%) |
| Yes | 400/3,352 (11.9%) | 543/3,820 (14.2%) |
| No data | 1/3,352 (0.0%) | 1/3,820 (0.0%) |
| Missing | 19/3,352 (0.6%) | 16/3,820 (0.4%) |
| VIOLATION: Last 4 weeks - fever, sore throat, cough | | |
| No | 3,102/3,352 (92.5%) | 3,534/3,820 (92.5%) |
| Yes | 234/3,352 (7.0%) | 270/3,820 (7.1%) |
| No data | 0/3,352 (0.0%) | 1/3,820 (0.0%) |
| Missing | 16/3,352 (0.5%) | 15/3,820 (0.4%) |
| VIOLATION: Last 4 weeks - diarrhea, stomachache | | |
| No | 3,285/3,352 (98.0%) | 3,744/3,820 (98.0%) |
| Yes | 50/3,352 (1.5%) | 58/3,820 (1.5%) |
| No data | 0/3,352 (0.0%) | 1/3,820 (0.0%) |
| Missing | 17/3,352 (0.5%) | 17/3,820 (0.4%) |
| VIOLATION: Last 4 weeks - stomachache | | |
| No | 3,259/3,352 (97.2%) | 3,712/3,820 (97.2%) |
| Yes | 79/3,352 (2.4%) | 95/3,820 (2.5%) |
| No data | 1/3,352 (0.0%) | 3/3,820 (0.1%) |
| Missing | 13/3,352 (0.4%) | 10/3,820 (0.3%) |
| VIOLATION: Last 4 weeks - asthma | | |
| No | 3,324/3,352 (99.2%) | 3,788/3,820 (99.2%) |
| Yes | 14/3,352 (0.4%) | 20/3,820 (0.5%) |
| No data | 1/3,352 (0.0%) | 2/3,820 (0.1%) |
| Missing | 13/3,352 (0.4%) | 10/3,820 (0.3%) |
| VIOLATION: Last 4 weeks - headache, dizziness | | |
| No | 3,245/3,352 (96.8%) | 3,588/3,820 (93.9%) |
| Yes | 91/3,352 (2.7%) | 217/3,820 (5.7%) |
| No data | 1/3,352 (0.0%) | 3/3,820 (0.1%) |
| Missing | 15/3,352 (0.4%) | 12/3,820 (0.3%) |
| VIOLATION: Last 4 weeks - joint, muscle pain | | |
| No | 3,239/3,352 (96.6%) | 3,603/3,820 (94.3%) |
| Yes | 97/3,352 (2.9%) | 203/3,820 (5.3%) |
| No data | 1/3,352 (0.0%) | 3/3,820 (0.1%) |
| Missing | 15/3,352 (0.4%) | 11/3,820 (0.3%) |
| VIOLATION: Last 4 weeks - rash, dermatitis | | |
| No | 3,319/3,352 (99.0%) | 3,789/3,820 (99.2%) |
| Yes | 16/3,352 (0.5%) | 19/3,820 (0.5%) |
| No data | 1/3,352 (0.0%) | 1/3,820 (0.0%) |

| | Overall sample | |
|---|---|---|
| | Male<br>N=3,352 | Female<br>N=3,820 |
| Missing | 16/3,352 (0.5%) | 11/3,820 (0.3%) |
| VIOLATION: Last 4 weeks - eye/ear disease | | |
| No | 3,322/3,352 (99.1%) | 3,785/3,820 (99.1%) |
| Yes | 13/3,352 (0.4%) | 23/3,820 (0.6%) |
| No data | 1/3,352 (0.0%) | 1/3,820 (0.0%) |
| Missing | 16/3,352 (0.5%) | 11/3,820 (0.3%) |
| VIOLATION: Last 4 weeks - heart disease/chest pain | | |
| No | 3,309/3,352 (98.7%) | 3,757/3,820 (98.4%) |
| Yes | 26/3,352 (0.8%) | 51/3,820 (1.3%) |
| No data | 1/3,352 (0.0%) | 1/3,820 (0.0%) |
| Missing | 16/3,352 (0.5%) | 11/3,820 (0.3%) |
| VIOLATION: Last 4 weeks - other infectious disease | | |
| No | 3,284/3,352 (98.0%) | 3,752/3,820 (98.2%) |
| Yes | 41/3,352 (1.2%) | 41/3,820 (1.1%) |
| No data | 2/3,352 (0.1%) | 6/3,820 (0.2%) |
| Missing | 25/3,352 (0.7%) | 21/3,820 (0.5%) |
| VIOLATION: Last 4 weeks - noncommunicable disease | | |
| No | 3,196/3,352 (95.3%) | 3,637/3,820 (95.2%) |
| Yes | 129/3,352 (3.8%) | 155/3,820 (4.1%) |
| No data | 2/3,352 (0.1%) | 7/3,820 (0.2%) |
| Missing | 25/3,352 (0.7%) | 21/3,820 (0.5%) |
| VIOLATION: Last 4 weeks - days hospitalised | | |
| Yes | 31/3,352 (0.9%) | 27/3,820 (0.7%) |
| Missing | 3,321/3,352 (99.1%) | 3,793/3,820 (99.3%) |
| VIOLATION: Currently pregnant or don't know | | |
| No | 0/3,352 (0.0%) | 2,124/3,820 (55.6%) |
| Yes | 0/3,352 (0.0%) | 64/3,820 (1.7%) |
| Missing | 3,352/3,352 (100.0%) | 1,632/3,820 (42.7%) |
| VIOLATION: # of months pregnant | | |
| Yes | 0/3,352 (0.0%) | 62/3,820 (1.6%) |
| Missing | 3,352/3,352 (100.0%) | 3,758/3,820 (98.4%) |
| VIOLATION: Taking anti-hypertension drugs | | |
| No | 88/3,352 (2.6%) | 80/3,820 (2.1%) |
| Yes | 228/3,352 (6.8%) | 300/3,820 (7.9%) |
| No data | 0/3,352 (0.0%) | 1/3,820 (0.0%) |
| Missing | 3,036/3,352 (90.6%) | 3,439/3,820 (90.0%) |
| VIOLATION: Province – Guizhou – Altitude | | |
| No | 3,073/3,352 (91.7%) | 3,506/3,820 (91.8%) |
| Yes | 279/3,352 (8.3%) | 314/3,820 (8.2%) |
| VIOLATION: still smokes cigarettes | | |
| No | 172/3,352 (5.1%) | 12/3,820 (0.3%) |
| Yes | 1,909/3,352 (57.0%) | 109/3,820 (2.9%) |
| No data | 1,271/3,352 (37.9%) | 3,699/3,820 (96.8%) |
| VIOLATION: Alcohol consumption almost every day | | |
| No | 1,521/3,352 (45.4%) | 317/3,820 (8.3%) |
| Yes | 605/3,352 (18.0%) | 34/3,820 (0.9%) |
| Missing | 1,226/3,352 (36.6%) | 3,469/3,820 (90.8%) |
| VIOLATION (# beer bottles per week): >6 for women and >8 for men | | |

| | Overall sample | |
|---|---|---|
| | Male N=3,352 | Female N=3,820 |
| No | 1,362/3,352 (40.6%) | 210/3,820 (5.5%) |
| Yes | 107/3,352 (3.2%) | 9/3,820 (0.2%) |
| Missing | 1,883/3,352 (56.2%) | 3,601/3,820 (94.3%) |
| VIOLATION (# wine drinks per week): >6 for women and >8 for men | | |
| No | 224/3,352 (6.7%) | 105/3,820 (2.7%) |
| Yes | 56/3,352 (1.7%) | 11/3,820 (0.3%) |
| Missing | 3,072/3,352 (91.6%) | 3,704/3,820 (97.0%) |
| VIOLATION (# liquor drinks per week): >6 for women and >8 for men | | |
| No | 1,075/3,352 (32.1%) | 134/3,820 (3.5%) |
| Yes | 561/3,352 (16.7%) | 36/3,820 (0.9%) |
| Missing | 1,716/3,352 (51.2%) | 3,650/3,820 (95.5%) |
| VIOLATION (# beer, wine, liquor): >6 for women and >8 for men | | |
| No | 1,357/3,352 (40.5%) | 285/3,820 (7.5%) |
| Yes | 759/3,352 (22.6%) | 64/3,820 (1.7%) |
| Missing | 1,236/3,352 (36.9%) | 3,471/3,820 (90.9%) |
| VIOLATION: Ferritin<15ug/L or Ferritin>150/200ug/L (F/M) | | |
| No | 2,343/3,352 (69.9%) | 2,808/3,820 (73.5%) |
| Yes | 1,009/3,352 (30.1%) | 1,012/3,820 (26.5%) |
| VIOLATION: C-Reactive Protein>5mg/L | | |
| No | 3,058/3,352 (91.2%) | 3,538/3,820 (92.6%) |
| Yes | 294/3,352 (8.8%) | 282/3,820 (7.4%) |
| VIOLATION: eGFR<90 ml/min/1.73m$^2$ | | |
| No | 1,570/3,352 (46.8%) | 1,281/3,820 (33.5%) |
| Yes | 1,782/3,352 (53.2%) | 2,538/3,820 (66.4%) |
| Missing | 0/3,352 (0.0%) | 1/3,820 (0.0%) |

CHN = China, CHNS = China Health and Nutrition Survey, BMI = Body Mass Index, F = Female, M = Male.
Data are presented as unweighted n/total (%) based on participants with non-missing haemoglobin, ferritin, and C-reactive protein.
No data: No answer/refused, Don't know; Missing: Not applicable, Not asked.
[1] Males: ferritin≥15 and ≤200ug/L and C-reactive protein≤5mg/L; females: ferritin≥15 and ≤150ug/L and C-reactive protein≤5mg/L.

#### Table 4.1.13 AUS NHS 2011-2012: Participant characteristics of violations (18-65 years)

| | Overall sample | |
|---|---|---|
| | Male<br>N=1,912 | Female<br>N=2,480 |
| Reference sample[1] | | |
| No | 1,699/1,912 (88.9%) | 2,198/2,480 (88.6%) |
| Yes | 213/1,912 (11.1%) | 282/2,480 (11.4%) |
| VIOLATION: BMI<18.5 or >30 kg/m$^2$ | | |
| No | 1,282/1,912 (67.1%) | 1,626/2,480 (65.6%) |
| Yes | 577/1,912 (30.2%) | 678/2,480 (27.3%) |
| Missing | 53/1,912 (2.8%) | 176/2,480 (7.1%) |
| VIOLATION: Co-morbidities | | |
| No | 1,143/1,912 (59.8%) | 1,338/2,480 (54.0%) |
| Yes | 769/1,912 (40.2%) | 1,142/2,480 (46.0%) |
| VIOLATION: Dialysis | | |
| No | Not displayed* | Not displayed* |
| Yes | <10/1,912 (<0.5%) | <10/2,480(<0.4%) |
| VIOLATION: Hospital last 12 months | | |
| No | Not displayed* | 2,460/2,480 (99.2%) |
| Yes | <10/1,912 (<0.5%) | 20/2,480 (0.8%) |
| VIOLATION: Hospital last 2 weeks | | |
| No | 1,902/1,912 (99.5%) | 2,457/2,480 (99.1%) |
| Yes | 10/1,912 (0.5%) | 23/2,480 (0.9%) |
| VIOLATION: Time off work or study due to illness | | |
| No | 1,445/1,912 (75.6%) | 1,592/2,480 (64.2%) |
| Yes | 162/1,912 (8.5%) | 257/2,480 (10.4%) |
| Missing | 305/1,912 (16.0%) | 631/2,480 (25.4%) |
| VIOLATION: Time off work due to illness | | |
| No | 1,411/1,912 (73.8%) | 1,540/2,480 (62.1%) |
| Yes | 151/1,912 (7.9%) | 233/2,480 (9.4%) |
| Missing | 350/1,912 (18.3%) | 707/2,480 (28.5%) |
| VIOLATION: Time off study due to illness | | |
| No | 181/1,912 (9.5%) | 306/2,480 (12.3%) |
| Yes | 15/1,912 (0.8%) | 30/2,480 (1.2%) |
| Missing | 1,716/1,912 (89.7%) | 2,144/2,480 (86.5%) |
| VIOLATION: Medication | | |
| No | 969/1,912 (50.7%) | 1,101/2,480 (44.4%) |
| Yes | 943/1,912 (49.3%) | 1,379/2,480 (55.6%) |
| VIOLATION: Breastfeeding | | |
| No | Not applicable | 2,388/2,480 (97.5%) |
| Yes | Not applicable | 62/2,480 (2.5%) |
| VIOLATION: Pregnancy | | |
| No | Not applicable | 2,388/2,480 (98.7%) |
| Yes | Not applicable | 32/2,480 (1.3%) |
| VIOLATION: Smoking | | |
| No | 1,615/1,912 (84.5%) | 2,103/2,480 (84.8%) |
| Yes | 297/1,912 (15.5%) | 377/2,480 (15.2%) |
| VIOLATION: Alcohol | | |
| No | 1,365/1,912 (71.4%) | 1,388/2,480 (56.0%) |
| Yes | 12/1,912 (0.6%) | 44/2,480 (1.8%) |
| Missing | 535/1,912 (28.0%) | 1,048/2,480 (42.3%) |

| | Overall sample | |
|---|---|---|
| | Male N=1,912 | Female N=2,480 |
| VIOLATION: Ferritin<15ug/L or Ferritin>150/200ug/L (F/M) | | |
| No | 1,072/1,912 (56.1%) | 1,933/2,480 (77.9%) |
| Yes | 840/1,912 (43.9%) | 547/2,480 (22.1%) |
| VIOLATION: C-Reactive Protein>5mg/L | | |
| No | 1,713/1,912 (89.6%) | 2,010/2,480 (81.0%) |
| Yes | 199/1,912 (10.4%) | 470/2,480 (19.0%) |
| VIOLATION: eGFR<90 ml/min/1.73m$^2$ | | |
| No | 1,416/1,910 (74.1%) | 1,919/2,478 (77.4%) |
| Yes | 494/1,910 (25.8%) | 559/2,478 (22.5%) |

AUS = Australia, NHS = National Health Survey, BMI = Body Mass Index, F = Female, M = Male.
Data are presented as unweighted n/total (%) based on participants with non-missing haemoglobin, ferritin, and C-reactive protein.

[*] Not displayed due to Australian Bureau of Statistics requirements for output clearance.

[1] Males: ferritin≥15 and ≤200ug/L and C-reactive protein≤5mg/L; females: ferritin≥15 and ≤150ug/L and C-reactive protein≤5mg/L.

#### Table 4.1.14 AUS NNPAS 2011-2012: Participant characteristics of violations (18-65 years)

| | Overall sample | |
|---|---|---|
| | **Male N=1,277** | **Female N=1,593** |
| Reference sample[1] | | |
| No | 983/1,277 (77.0%) | 1,114/1,593 (69.9%) |
| Yes | 294/1,277 (23.0%) | 479/1,593 (30.1%) |
| VIOLATION: BMI<18.5 or >30 kg/m$^2$ | | |
| No | 889/1,277 (69.6%) | 1,053/1,593 (66.1%) |
| Yes | 345/1,277 (27.0%) | 445/1,593 (27.9%) |
| Missing | 43/1,277 (3.4%) | 95/1,593 (6.0%) |
| VIOLATION: Co-morbidities | | |
| No | 1,084/1,277 (84.9%) | 1,367/1,593 (85.8%) |
| Yes | 193/1,277 (15.1%) | 226/1,593 (14.2%) |
| VIOLATION: Breastfeeding | | |
| No | Not applicable | 1,552/1,593 (97.4%) |
| Yes | Not applicable | 41/1,593 (2.6%) |
| VIOLATION: Pregnancy | | |
| No | Not applicable | 1,571/1,593 (98.6%) |
| Yes | Not applicable | 22/1,593 (1.4%) |
| VIOLATION: Smoking | | |
| No | 1,059/1,277 (82.9%) | 1,371/1,593 (86.1%) |
| Yes | 218/1,277 (17.1%) | 222/1,593 (13.9%) |
| VIOLATION: Ferritin<15ug/L or Ferritin>150/200ug/L (F/M) | | |
| No | 739/1,277 (57.9%) | 1,253/1,593 (78.7%) |
| Yes | 538/1,277 (42.1%) | 340/1,593 (21.3%) |
| VIOLATION: C-Reactive Protein>5mg/L | | |
| No | 1,145/1,277 (89.7%) | 1,288/1,593 (80.9%) |
| Yes | 132/1,277 (10.3%) | 305/1,593 (19.1%) |
| VIOLATION: eGFR<90 ml/min/1.73m$^2$ | | |
| No | 877/1,276 (68.7%) | 1,216/1,593 (76.3%) |
| Yes | 399/1,276 (31.2%) | 377/1,593 (23.7%) |

AUS = Australia, NNPAS = National Nutrition and Physical Activity Survey, BMI = Body Mass Index, F = Female, M = Male.

Data are presented as unweighted n/total (%) based on participants with non-missing haemoglobin, ferritin, and C-reactive protein.

[1] Males: ferritin≥15 and ≤200ug/L and C-reactive protein≤5mg/L; females: ferritin≥15 and ≤150ug/L and C-reactive protein≤5mg/L.

#### 4.2 Detailed Individual Exclusions Children (6-23 months)

##### Table 4.2.1 CAN TARGet Kids: Participant characteristics of violations (6-23 months)

| | **Overall sample Male and Female N=1,671** |
|---|---|
| Reference sample[1] | |
| No | 1,248/1,671 (74.7%) |
| Yes | 423/1,671 (25.3%) |
| VIOLATION: Low birth weight (<2.5 kg) if age ≤24 months | |
| No | 1,377/1,671 (82.4%) |
| Yes | 209/1,671 (12.5%) |
| Missing | 85/1,671 (5.1%) |
| VIOLATION: Weight z-score <-2 or >+2 | |
| No | 1,558/1,671 (93.2%) |
| Yes | 85/1,671 (5.1%) |
| Missing | 28/1,671 (1.7%) |
| VIOLATION: Length/height z-score <-2 or >+2 | |
| No | 1,421/1,671 (85.0%) |
| Yes | 212/1,671 (12.7%) |
| Missing | 38/1,671 (2.3%) |
| VIOLATION: Weight for length z-score <-2 or >+2 | |
| No | 1,521/1,671 (91.0%) |
| Yes | 109/1,671 (6.5%) |
| Missing | 41/1,671 (2.5%) |
| VIOLATION: BMI z-score <-2 or >+2 | |
| No | 1,507/1,671 (90.2%) |
| Yes | 123/1,671 (7.4%) |
| Missing | 41/1,671 (2.5%) |
| VIOLATION: Aged ≤12 months and >6 months exclusively breastfed | |
| No | 446/1,671 (26.7%) |
| Yes | 64/1,671 (3.8%) |
| Missing | 1,161/1,671 (69.5%) |
| VIOLATION: Anemia (mother) | |
| No | 1,449/1,671 (86.7%) |
| Yes | 110/1,671 (6.6%) |
| Missing | 112/1,671 (6.7%) |
| VIOLATION: Birth (<37 weeks or > 42 weeks) if age ≤24 months | |
| No | 1,263/1,671 (75.6%) |
| Yes | 219/1,671 (13.1%) |
| Missing | 189/1,671 (11.3%) |
| VIOLATION: Nursery if < 24 months | |
| No | 433/1,671 (25.9%) |
| Yes | 107/1,671 (6.4%) |
| Missing | 1,131/1,671 (67.7%) |
| VIOLATION: Allergies | |
| No | 1,076/1,671 (64.4%) |
| Yes | 53/1,671 (3.2%) |
| Missing | 542/1,671 (32.4%) |
| VIOLATION: Asthma | |

| | Overall sample<br>Male and Female<br>N=1,671 |
|---|---|
| No | 1,414/1,671 (84.6%) |
| Yes | 27/1,671 (1.6%) |
| Missing | 230/1,671 (13.8%) |
| VIOLATION: Austism/PDD | |
| No | 1,434/1,671 (85.8%) |
| Yes | 3/1,671 (0.2%) |
| Missing | 234/1,671 (14.0%) |
| VIOLATION: Development delay | |
| No | 1,106/1,671 (66.2%) |
| Yes | 13/1,671 (0.8%) |
| Missing | 552/1,671 (33.0%) |
| VIOLATION: Diabetes | |
| No | 1,434/1,671 (85.8%) |
| Yes | 3/1,671 (0.2%) |
| Missing | 234/1,671 (14.0%) |
| VIOLATION: At least one diagnosis | |
| No | 1,172/1,671 (70.1%) |
| Yes | 234/1,671 (14.0%) |
| Missing | 265/1,671 (15.9%) |
| VIOLATION: Overweight | |
| No | 1,114/1,671 (66.7%) |
| Yes | 3/1,671 (0.2%) |
| Missing | 554/1,671 (33.2%) |
| VIOLATION: IBD | |
| No | 333/1,671 (19.9%) |
| Missing | 1,338/1,671 (80.1%) |
| VIOLATION: Cancer | |
| No | 543/1,671 (32.5%) |
| Yes | 1/1,671 (0.1%) |
| Missing | 1,127/1,671 (67.4%) |
| VIOLATION: Other diagnosis | |
| No | 1,138/1,671 (68.1%) |
| Yes | 62/1,671 (3.7%) |
| Missing | 471/1,671 (28.2%) |
| VIOLATION: Ill past month | |
| No | 125/1,671 (7.5%) |
| Yes | 134/1,671 (8.0%) |
| Missing | 1,412/1,671 (84.5%) |
| VIOLATION: Regular prescribed medicine | |
| No | 1,511/1,671 (90.4%) |
| Yes | 68/1,671 (4.1%) |
| Missing | 92/1,671 (5.5%) |
| VIOLATION: Prescribed medicine past 2 weeks | |
| No | 301/1,671 (18.0%) |
| Yes | 25/1,671 (1.5%) |
| Missing | 1,345/1,671 (80.5%) |
| VIOLATION: Cold or flu medicine past month | |
| No | 119/1,671 (7.1%) |
| Yes | 46/1,671 (2.8%) |

| | Overall sample Male and Female N=1,671 |
|---|---|
| Missing | 1,506/1,671 (90.1%) |
| VIOLATION: Other medicine past month | |
| No | 97/1,671 (5.8%) |
| Yes | 99/1,671 (5.9%) |
| Missing | 1,475/1,671 (88.3%) |
| VIOLATION: Cold or flu medicine past 2 weeks | |
| No | 277/1,671 (16.6%) |
| Yes | 44/1,671 (2.6%) |
| Missing | 1,350/1,671 (80.8%) |
| VIOLATION: Other medicine past 2 weeks | |
| No | 273/1,671 (16.3%) |
| Yes | 48/1,671 (2.9%) |
| Missing | 1,350/1,671 (80.8%) |
| VIOLATION: Iron (e.g., Ferinsol, Palafer) intake | |
| No | 973/1,671 (58.2%) |
| Yes | 28/1,671 (1.7%) |
| Missing | 670/1,671 (40.1%) |
| VIOLATION: Other iron intake | |
| No | 328/1,671 (19.6%) |
| Yes | 1/1,671 (0.1%) |
| Missing | 1,342/1,671 (80.3%) |
| VIOLATION: Traditional remedies or cosmetics last 3 months | |
| No | 246/1,671 (14.7%) |
| Yes | 12/1,671 (0.7%) |
| Missing | 1,413/1,671 (84.6%) |
| VIOLATION: Lead (household) | |
| No | 232/1,671 (13.9%) |
| Yes | 25/1,671 (1.5%) |
| Missing | 1,414/1,671 (84.6%) |
| VIOLATION: Smoking (household) | |
| No | 1,402/1,671 (83.9%) |
| Yes | 178/1,671 (10.7%) |
| Missing | 91/1,671 (5.4%) |
| VIOLATION: Ferritin<12ug/L | |
| No | 1,459/1,671 (87.3%) |
| Yes | 212/1,671 (12.7%) |
| VIOLATION: C-Reactive Protein>5mg/L | |
| No | 1,568/1,671 (93.8%) |
| Yes | 103/1,671 (6.2%) |
| VIOLATION: Albumin<32g/L | |
| No | 1,630/1,671 (97.5%) |
| Missing | 41/1,671 (2.5%) |
| VIOLATION: Alkaline Phosphatase (age and sex dependent) | |
| No | 1,533/1,671 (91.7%) |
| Yes | 96/1,671 (5.7%) |
| Missing | 42/1,671 (2.5%) |
| VIOLATION: ALT serum≥35U/L | |
| No | 1,582/1,671 (94.7%) |
| Yes | 88/1,671 (5.3%) |

| | Overall sample Male and Female N=1,671 |
|---|---|
| Missing | 1/1,671 (0.1%) |

CAN = Canada, TARGet Kids! = The Applied Research Group for Kids, BMI = Body Mass Index.
Data are presented as n/total (%) based on participants with non-missing haemoglobin, ferritin, and C-reactive protein.
Missing: No answer.
[1] Ferritin≥12ug/L and C-reactive protein≤5mg/L.

#### Table 4.2.2 BAN BRISC 2017-2020: Participant characteristics of violations (6-23 months)

| | Overall sample Male and Female N=1,365 |
|---|---|
| Reference sample[1] | |
| No | 1,110/1,365 (81.3%) |
| Yes | 255/1,365 (18.7%) |
| VIOLATION: Weight z-score <-2 or >+2 | |
| No | 1,235/1,365 (90.5%) |
| Yes | 127/1,365 (9.3%) |
| Missing | 3/1,365 (0.2%) |
| VIOLATION: Length/height z-score <-2 or >+2 | |
| No | 1,053/1,365 (77.1%) |
| Yes | 309/1,365 (22.6%) |
| Missing | 3/1,365 (0.2%) |
| VIOLATION: Weight for length z-score <-2 or >+2 | |
| No | 1,300/1,365 (95.2%) |
| Yes | 62/1,365 (4.5%) |
| Missing | 3/1,365 (0.2%) |
| VIOLATION: Any meds in the 4 weeks before midline visit | |
| No | 806/1,365 (59.0%) |
| Yes | 545/1,365 (39.9%) |
| Missing | 14/1,365 (1.0%) |
| VIOLATION: Compliance<70% [Iron and MNP specific] | |
| No | 1,120/1,365 (82.1%) |
| Yes | 245/1,365 (17.9%) |
| VIOLATION: Any medical visit in the 4 weeks before midline visit | |
| No | 783/1,365 (57.4%) |
| Yes | 567/1,365 (41.5%) |
| Missing | 15/1,365 (1.1%) |
| VIOLATION: Any infection in the 4 weeks before midline visit | |
| No | 594/1,365 (43.5%) |
| Yes | 759/1,365 (55.6%) |
| Missing | 12/1,365 (0.9%) |
| VIOLATION: Ferritin<12ug/L | |
| No | 1,203/1,365 (88.1%) |
| Yes | 162/1,365 (11.9%) |
| VIOLATION: C-Reactive Protein>5mg/L | |
| No | 1,220/1,365 (89.4%) |
| Yes | 145/1,365 (10.6%) |

BAN = Bangladesh, BRISC = Benefits and Risks of Iron intervention in Children.
Data are presented as n/total (%) based on participants with non-missing haemoglobin, ferritin, and C-reactive protein.
[1] Ferritin≥12ug/L and C-reactive protein≤5mg/L.

#### Table 4.2.3 ECU ENSANUT 2011-2013: Participant characteristics of violations (6-23 months)

| | **Overall sample Male and Female N=787** |
|---|---|
| Reference sample[1] | 95/787 (12.1%) |
| VIOLATION: Altitude > 750 meters | |
| No | 493/787 (62.6%) |
| Yes | 294/787 (37.4%) |
| VIOLATION: Weighed less than 5.5 pounds or 2.5 kilograms | |
| No | 55/787 (7.0%) |
| Yes | 13/787 (1.7%) |
| No data | 124/787 (15.8%) |
| Missing | 595/787 (75.6%) |
| VIOLATION: Birthweight less than 2500 grams | |
| No | 445/787 (56.5%) |
| Yes | 25/787 (3.2%) |
| Missing | 317/787 (40.3%) |
| VIOLATION: Born ≥5 weeks early or late | |
| No | 90/787 (11.4%) |
| Yes | 12/787 (1.5%) |
| No data | 5/787 (0.6%) |
| Missing | 680/787 (86.4%) |
| VIOLATION: Health problems in the last 30 days | |
| No | 265/787 (33.7%) |
| Yes | 522/787 (66.3%) |
| VIOLATION: Hospitalisation in the last 30 days | |
| No | 732/787 (93.0%) |
| Yes | 55/787 (7.0%) |
| VIOLATION: Diarrhea in the last 2 weeks | |
| No | 614/787 (78.0%) |
| Yes | 163/787 (20.7%) |
| No data | 1/787 (0.1%) |
| Missing | 9/787 (1.1%) |
| VIOLATION: Cough, runny nose in the last 2 weeks | |
| No | 424/787 (53.9%) |
| Yes | 354/787 (45.0%) |
| Missing | 9/787 (1.1%) |
| VIOLATION: Ferritin<12ug/L | |
| No | 648/787 (82.3%) |
| Yes | 139/787 (17.7%) |
| VIOLATION: C-Reactive Protein>5mg/L | |
| No | 682/787 (86.7%) |
| Yes | 105/787 (13.3%) |

ECU = Ecuador, ENSANUT = Encuesta National de Salud y Nutricion.
Data are presented as unweighted n/total (%) based on participants with non-missing haemoglobin, ferritin, and C-reactive protein.
No data: No answer/refused, Don't know; Missing: Not applicable, Not asked.
[1] Ferritin≥12ug/L and C-reactive protein≤5mg/L.

#### Table 4.2.4 US NHANES 2003-2018: Participant characteristics of violations (12-23 months)

| | Overall sample<br>Male and Female<br>N=530 |
|---|---|
| Reference sample[1] | 151/530 (28.5%) |
| VIOLATION: Doctor told you have diabetes | |
| No | 529/530 (99.8%) |
| Yes | 1/530 (0.2%) |
| VIOLATION: Still have asthma | |
| No | 3/530 (0.6%) |
| Yes | 38/530 (7.2%) |
| No data | 1/530 (0.2%) |
| Missing | 488/530 (92.1%) |
| VIOLATION: Taking treatment for anemia/past 3 months | |
| No | 515/530 (97.2%) |
| Yes | 15/530 (2.8%) |
| VIOLATION: Overnight hospital patient in last year | |
| No | 470/530 (88.7%) |
| Yes | 60/530 (11.3%) |
| VIOLATION: SP have head cold or chest cold? | |
| No | 290/530 (54.7%) |
| Yes | 215/530 (40.6%) |
| No data | 25/530 (4.7%) |
| VIOLATION: SP have stomach or intestinal illness? | |
| No | 416/530 (78.5%) |
| Yes | 87/530 (16.4%) |
| No data | 27/530 (5.1%) |
| VIOLATION: SP have flu, pneumonia, ear infection? | |
| No | 447/530 (84.3%) |
| Yes | 54/530 (10.2%) |
| No data | 29/530 (5.5%) |
| VIOLATION: Taken prescription medicine, past month | |
| No | 414/530 (78.1%) |
| Yes | 116/530 (21.9%) |
| VIOLATION: For anemia, such as low iron | |
| No | 97/530 (18.3%) |
| Yes | 4/530 (0.8%) |
| No data | 117/530 (22.1%) |
| Missing | 312/530 (58.9%) |
| VIOLATION: Ferritin<12ug/L | |
| No | 461/530 (87.0%) |
| Yes | 69/530 (13.0%) |
| VIOLATION: C-Reactive Protein>5mg/L | |
| No | 487/530 (91.9%) |
| Yes | 43/530 (8.1%) |

US = United States, NHANES = National Health and Nutrition Examination Survey, SP = Survey Participant.
Data are presented as unweighted n/total (%) based on participants with non-missing haemoglobin, ferritin, and C-reactive protein. Data was combined from 1999-2018. No ferritin was measured in participants in survey years 1999-2000, 2001-2002, 2007-2008, 2009-2010, 2011-2012, 2013-2014. VIOLATION: For anemia, such as low iron - only asked in survey years 2015-2016 and 2017-2018.
No data: No answer/refused, Don't know; Missing: Not applicable, Not asked.

[1] Ferritin≥12ug/L and C-reactive protein≤5mg/L.

#### 4.3 Detailed Individual Exclusions Children (24-59 months)

##### Table 4.3.1 CAN TARGet Kids: Participant characteristics of violations (24-59 months)

| | Overall sample Male and Female N=2,050 |
|---|---|
| Reference sample[1] | |
| No | 1,293/2,050 (63.1%) |
| Yes | 757/2,050 (36.9%) |
| VIOLATION: Low birth weight (<2.5 kg) if age ≤24 months | |
| No | 1,861/2,050 (90.8%) |
| Yes | 43/2,050 (2.1%) |
| Missing | 146/2,050 (7.1%) |
| VIOLATION: Weight z-score <-2 or >+2 | |
| No | 1,903/2,050 (92.8%) |
| Yes | 101/2,050 (4.9%) |
| Missing | 46/2,050 (2.2%) |
| VIOLATION: Length/height z-score <-2 or >+2 | |
| No | 1,880/2,050 (91.7%) |
| Yes | 120/2,050 (5.9%) |
| Missing | 50/2,050 (2.4%) |
| VIOLATION: Weight for length z-score <-2 or >+2 | |
| No | 1,872/2,050 (91.3%) |
| Yes | 125/2,050 (6.1%) |
| Missing | 53/2,050 (2.6%) |
| VIOLATION: BMI z-score <-2 or >+2 | |
| No | 1,863/2,050 (90.9%) |
| Yes | 137/2,050 (6.7%) |
| Missing | 50/2,050 (2.4%) |
| VIOLATION: Anemia (mother) | |
| No | 1,757/2,050 (85.7%) |
| Yes | 120/2,050 (5.9%) |
| Missing | 173/2,050 (8.4%) |
| VIOLATION: Birth (<37 weeks or > 42 weeks) if age ≤24 months | |
| No | 1,721/2,050 (84.0%) |
| Yes | 37/2,050 (1.8%) |
| Missing | 292/2,050 (14.2%) |
| VIOLATION: Allergies | |
| No | 1,006/2,050 (49.1%) |
| Yes | 81/2,050 (4.0%) |
| Missing | 963/2,050 (47.0%) |
| VIOLATION: Asthma | |
| No | 1,650/2,050 (80.5%) |
| Yes | 103/2,050 (5.0%) |
| Missing | 297/2,050 (14.5%) |
| VIOLATION: Austism/PDD | |
| No | 1,737/2,050 (84.7%) |
| Yes | 12/2,050 (0.6%) |
| Missing | 301/2,050 (14.7%) |
| VIOLATION: Development delay | |
| No | 1,046/2,050 (51.0%) |

| | Overall sample<br>Male and Female<br>N=2,050 |
|---|---|
| Yes | 37/2,050 (1.8%) |
| Missing | 967/2,050 (47.2%) |
| VIOLATION: Diabetes | |
| No | 1,745/2,050 (85.1%) |
| Yes | 3/2,050 (0.1%) |
| Missing | 302/2,050 (14.7%) |
| VIOLATION: At least one diagnosis | |
| No | 1,323/2,050 (64.5%) |
| Yes | 372/2,050 (18.1%) |
| Missing | 355/2,050 (17.3%) |
| VIOLATION: Overweight | |
| No | 1,072/2,050 (52.3%) |
| Yes | 3/2,050 (0.1%) |
| Missing | 975/2,050 (47.6%) |
| VIOLATION: IBD | |
| No | 469/2,050 (22.9%) |
| Missing | 1,581/2,050 (77.1%) |
| VIOLATION: Cancer | |
| No | 598/2,050 (29.2%) |
| Missing | 1,452/2,050 (70.8%) |
| VIOLATION: Other diagnosis | |
| No | 1,052/2,050 (51.3%) |
| Yes | 124/2,050 (6.0%) |
| Missing | 874/2,050 (42.6%) |
| VIOLATION: Ill past month | |
| No | 354/2,050 (17.3%) |
| Yes | 259/2,050 (12.6%) |
| Missing | 1,437/2,050 (70.1%) |
| VIOLATION: Regular prescribed medicine | |
| No | 1,741/2,050 (84.9%) |
| Yes | 95/2,050 (4.6%) |
| Missing | 214/2,050 (10.4%) |
| VIOLATION: Prescribed medicine past 2 weeks | |
| No | 621/2,050 (30.3%) |
| Yes | 64/2,050 (3.1%) |
| Missing | 1,365/2,050 (66.6%) |
| VIOLATION: Cold or flu medicine past month | |
| No | 294/2,050 (14.3%) |
| Yes | 139/2,050 (6.8%) |
| Missing | 1,617/2,050 (78.9%) |
| VIOLATION: Other medicine past month | |
| No | 282/2,050 (13.8%) |
| Yes | 167/2,050 (8.1%) |
| Missing | 1,601/2,050 (78.1%) |
| VIOLATION: Cold or flu medicine past 2 weeks | |
| No | 622/2,050 (30.3%) |
| Yes | 55/2,050 (2.7%) |
| Missing | 1,373/2,050 (67.0%) |
| VIOLATION: Other medicine past 2 weeks | |

| | Overall sample Male and Female N=2,050 |
|---|---|
| No | 623/2,050 (30.4%) |
| Yes | 54/2,050 (2.6%) |
| Missing | 1,373/2,050 (67.0%) |
| VIOLATION: Iron (e.g., Ferinsol, Palafer) intake | |
| No | 1,347/2,050 (65.7%) |
| Yes | 21/2,050 (1.0%) |
| Missing | 682/2,050 (33.3%) |
| VIOLATION: Other iron intake | |
| No | 680/2,050 (33.2%) |
| Yes | 6/2,050 (0.3%) |
| Missing | 1,364/2,050 (66.5%) |
| VIOLATION: Traditional remedies or cosmetics last 3 months | |
| No | 571/2,050 (27.9%) |
| Yes | 35/2,050 (1.7%) |
| Missing | 1,444/2,050 (70.4%) |
| VIOLATION: Lead (household) | |
| No | 546/2,050 (26.6%) |
| Yes | 59/2,050 (2.9%) |
| Missing | 1,445/2,050 (70.5%) |
| VIOLATION: Smoking (household) | |
| No | 1,626/2,050 (79.3%) |
| Yes | 218/2,050 (10.6%) |
| Missing | 206/2,050 (10.0%) |
| VIOLATION: Ferritin<12ug/L | |
| No | 1,911/2,050 (93.2%) |
| Yes | 139/2,050 (6.8%) |
| VIOLATION: C-Reactive Protein>5mg/L | |
| No | 1,906/2,050 (93.0%) |
| Yes | 144/2,050 (7.0%) |
| VIOLATION: Albumin<32g/L | |
| No | 1,818/2,050 (88.7%) |
| Yes | 1/2,050 (0.0%) |
| Missing | 231/2,050 (11.3%) |
| VIOLATION: Alkaline Phosphatase (age and sex dependent) | |
| No | 1,777/2,050 (86.7%) |
| Yes | 41/2,050 (2.0%) |
| Missing | 232/2,050 (11.3%) |
| VIOLATION: ALT serum≥35U/L | |
| No | 2,023/2,050 (98.7%) |
| Yes | 27/2,050 (1.3%) |

CAN = Canada, TARGet Kids! = The Applied Research Group for Kids, BMI = Body Mass Index.
Data are presented as n/total (%) based on participants with non-missing haemoglobin, ferritin, and C-reactive protein.
Missing: No answer.
[1] Ferritin≥12ug/L and C-reactive protein≤5mg/L.

#### Table 4.3.2 ECU ENSANUT 2011-2013: Participant characteristics of violations (24-59 months)

| | **Overall sample Male and Female** |
|---|---|
| | **N=1,260** |
| Reference sample[1] | 180/1,260 (14.3%) |
| VIOLATION: Altitude > 750 meters | |
| No | 692/1,260 (54.9%) |
| Yes | 568/1,260 (45.1%) |
| VIOLATION: Health problems in the last 30 days | |
| No | 474/1,260 (37.6%) |
| Yes | 786/1,260 (62.4%) |
| VIOLATION: Hospitalisation in the last 30 days | |
| No | 1,201/1,260 (95.3%) |
| Yes | 59/1,260 (4.7%) |
| VIOLATION: Diarrhea in the last 2 weeks | |
| No | 1,112/1,260 (88.3%) |
| Yes | 134/1,260 (10.6%) |
| No data | 1/1,260 (0.1%) |
| Missing | 13/1,260 (1.0%) |
| VIOLATION: Cough, runny nose in the last 2 weeks | |
| No | 676/1,260 (53.7%) |
| Yes | 571/1,260 (45.3%) |
| Missing | 13/1,260 (1.0%) |
| VIOLATION: Ferritin<12ug/L | |
| No | 1,210/1,260 (96.0%) |
| Yes | 50/1,260 (4.0%) |
| VIOLATION: C-Reactive Protein>5mg/L | |
| No | 1,112/1,260 (88.3%) |
| Yes | 148/1,260 (11.7%) |

ECU = Ecuador, ENSANUT = Encuesta National de Salud y Nutricion.
Data are presented as unweighted n/total (%) based on participants with non-missing haemoglobin, ferritin, and C-reactive protein.
No data: No answer/refused, Don't know; Missing: Not applicable, Not asked
[1] Ferritin≥12ug/L and C-reactive protein≤5mg/L.

#### Table 4.3.3 US NHANES 1999-2018: Participant characteristics of violations (24-59 months)

| | Overall sample<br>Male and Female<br>N=2,559 |
|---|---|
| Reference sample[1] | 937/2,559 (36.6%) |
| VIOLATION: BMI age and sex dependent | |
| No | 2,179/2,559 (85.2%) |
| Yes | 266/2,559 (10.4%) |
| No data | 81/2,559 (3.2%) |
| Missing | 33/2,559 (1.3%) |
| VIOLATION: Doctor told you have diabetes | |
| No | 2,555/2,559 (99.8%) |
| Yes | 2/2,559 (0.1%) |
| No data | 2/2,559 (0.1%) |
| VIOLATION: Still have asthma | |
| No | 88/2,559 (3.4%) |
| Yes | 209/2,559 (8.2%) |
| No data | 6/2,559 (0.2%) |
| Missing | 2,256/2,559 (88.2%) |
| VIOLATION: Taking treatment for anemia/past 3 months | |
| No | 2,504/2,559 (97.9%) |
| Yes | 53/2,559 (2.1%) |
| No data | 2/2,559 (0.1%) |
| VIOLATION: Overnight hospital patient in last year | |
| No | 2,430/2,559 (95.0%) |
| Yes | 127/2,559 (5.0%) |
| No data | 2/2,559 (0.1%) |
| VIOLATION: SP have head cold or chest cold? | |
| No | 1,518/2,559 (59.3%) |
| Yes | 879/2,559 (34.3%) |
| No data | 162/2,559 (6.3%) |
| VIOLATION: SP have stomach or intestinal illness? | |
| No | 2,124/2,559 (83.0%) |
| Yes | 279/2,559 (10.9%) |
| No data | 156/2,559 (6.1%) |
| VIOLATION: SP have flu, pneumonia, ear infection? | |
| No | 2,176/2,559 (85.0%) |
| Yes | 228/2,559 (8.9%) |
| No data | 155/2,559 (6.1%) |
| VIOLATION: Taken prescription medicine, past month | |
| No | 2,102/2,559 (82.1%) |
| Yes | 455/2,559 (17.8%) |
| No data | 2/2,559 (0.1%) |
| VIOLATION: For anemia, such as low iron | |
| No | 319/2,559 (12.5%) |
| Yes | 22/2,559 (0.9%) |
| No data | 882/2,559 (34.5%) |
| Missing | 1,336/2,559 (52.2%) |
| VIOLATION: Ferritin<12ug/L | |
| No | 2,365/2,559 (92.4%) |
| Yes | 194/2,559 (7.6%) |
| VIOLATION: C-Reactive Protein>5mg/L | |

| | |
|---|---|
| No | 2,390/2,559 (93.4%) |
| Yes | 169/2,559 (6.6%) |

US = United States, NHANES = National Health and Nutrition Examination Survey, BMI = Body Mass Index, SP = Survey Participant.

Data are presented as unweighted n/total (%) based on participants with non-missing haemoglobin, ferritin, and C-reactive protein. Data was combined from 1999-2018. No ferritin was measured in participants less than 3 years in survey years 1999-2000, 2001-2002, 2007-2008 and 2009-2010. No ferritin was measured in participants in survey years 2011-2012 and 2013-2014. VIOLATION: For anemia, such as low iron - only asked in survey years 2007-2008, 2009-2010, 2015-2016, and 2017-2018.

No data: No answer/refused, Don't know; Missing: Not applicable, Not asked.

[1] Ferritin≥12ug/L and C-reactive protein≤5mg/L.

#### 4.4 Detailed Individual Exclusions Children (5-11 years)

##### Table 4.4.1 CAN TARGet Kids: Participant characteristics of violations (5-11 years)

| | Overall sample<br>Male and Female<br>N=979 |
|---|---|
| Reference sample[1] | |
| No | 587/979 (60.0%) |
| Yes | 392/979 (40.0%) |
| VIOLATION: Weight z-score <-2 or >+2 | |
| No | 866/979 (88.5%) |
| Yes | 71/979 (7.3%) |
| Missing | 42/979 (4.3%) |
| VIOLATION: Length/height z-score <-2 or >+2 | |
| No | 894/979 (91.3%) |
| Yes | 65/979 (6.6%) |
| Missing | 20/979 (2.0%) |
| VIOLATION: Weight for length z-score <-2 or >+2 | |
| No | 133/979 (13.6%) |
| Yes | 7/979 (0.7%) |
| Missing | 839/979 (85.7%) |
| VIOLATION: BMI z-score <-2 or >+2 | |
| No | 891/979 (91.0%) |
| Yes | 66/979 (6.7%) |
| Missing | 22/979 (2.2%) |
| VIOLATION: Anemia (mother) | |
| No | 873/979 (89.2%) |
| Yes | 49/979 (5.0%) |
| Missing | 57/979 (5.8%) |
| VIOLATION: Allergies | |
| No | 495/979 (50.6%) |
| Yes | 63/979 (6.4%) |
| Missing | 421/979 (43.0%) |
| VIOLATION: Asthma | |
| No | 777/979 (79.4%) |
| Yes | 86/979 (8.8%) |
| Missing | 116/979 (11.8%) |
| VIOLATION: Austism/PDD | |
| No | 837/979 (85.5%) |
| Yes | 15/979 (1.5%) |
| Missing | 127/979 (13.0%) |
| VIOLATION: Development delay | |
| No | 528/979 (53.9%) |
| Yes | 14/979 (1.4%) |
| Missing | 437/979 (44.6%) |
| VIOLATION: Diabetes | |
| No | 845/979 (86.3%) |
| Yes | 3/979 (0.3%) |
| Missing | 131/979 (13.4%) |
| VIOLATION: At least one diagnosis | |
| No | 631/979 (64.5%) |

| | Overall sample<br>Male and Female<br>N=979 |
|---|---|
| Yes | 186/979 (19.0%) |
| Missing | 162/979 (16.5%) |
| VIOLATION: Overweight | |
| No | 537/979 (54.9%) |
| Yes | 5/979 (0.5%) |
| Missing | 437/979 (44.6%) |
| VIOLATION: IBD | |
| No | 186/979 (19.0%) |
| Yes | 2/979 (0.2%) |
| Missing | 791/979 (80.8%) |
| VIOLATION: Cancer | |
| No | 277/979 (28.3%) |
| Yes | 2/979 (0.2%) |
| Missing | 700/979 (71.5%) |
| VIOLATION: Other diagnosis | |
| No | 558/979 (57.0%) |
| Yes | 62/979 (6.3%) |
| Missing | 359/979 (36.7%) |
| VIOLATION: Ill past month | |
| No | 194/979 (19.8%) |
| Yes | 79/979 (8.1%) |
| Missing | 706/979 (72.1%) |
| VIOLATION: Regular prescribed medicine | |
| No | 877/979 (89.6%) |
| Yes | 54/979 (5.5%) |
| Missing | 48/979 (4.9%) |
| VIOLATION: Prescribed medicine past 2 weeks | |
| No | 292/979 (29.8%) |
| Yes | 19/979 (1.9%) |
| Missing | 668/979 (68.2%) |
| VIOLATION: Cold or flu medicine past month | |
| No | 95/979 (9.7%) |
| Yes | 51/979 (5.2%) |
| Missing | 833/979 (85.1%) |
| VIOLATION: Other medicine past month | |
| No | 81/979 (8.3%) |
| Yes | 67/979 (6.8%) |
| Missing | 831/979 (84.9%) |
| VIOLATION: Cold or flu medicine past 2 weeks | |
| No | 279/979 (28.5%) |
| Yes | 24/979 (2.5%) |
| Missing | 676/979 (69.1%) |
| VIOLATION: Other medicine past 2 weeks | |
| No | 283/979 (28.9%) |
| Yes | 20/979 (2.0%) |
| Missing | 676/979 (69.1%) |
| VIOLATION: Iron (e.g. Ferinsol, Palafer) intake | |
| No | 602/979 (61.5%) |
| Yes | 7/979 (0.7%) |

| | Overall sample<br>Male and Female<br>N=979 |
|---|---|
| Missing | 370/979 (37.8%) |
| VIOLATION: Other iron intake | |
| No | 306/979 (31.3%) |
| Yes | 1/979 (0.1%) |
| Missing | 672/979 (68.6%) |
| VIOLATION: Traditional remedies or cosmetics last 3 months | |
| No | 259/979 (26.5%) |
| Yes | 12/979 (1.2%) |
| Missing | 708/979 (72.3%) |
| VIOLATION: Lead (household) | |
| No | 247/979 (25.2%) |
| Yes | 23/979 (2.3%) |
| Missing | 709/979 (72.4%) |
| VIOLATION: Smoking (household) | |
| No | 822/979 (84.0%) |
| Yes | 99/979 (10.1%) |
| Missing | 58/979 (5.9%) |
| VIOLATION: Ferritin<15ug/L | |
| No | 926/979 (94.6%) |
| Yes | 53/979 (5.4%) |
| VIOLATION: C-Reactive Protein>5mg/L | |
| No | 934/979 (95.4%) |
| Yes | 45/979 (4.6%) |
| VIOLATION: Albumin<32g/L | |
| No | 955/979 (97.5%) |
| Missing | 24/979 (2.5%) |
| VIOLATION: Alkaline Phosphatase (age and sex dependent) | |
| No | 942/979 (96.2%) |
| Yes | 13/979 (1.3%) |
| Missing | 24/979 (2.5%) |
| VIOLATION: ALT serum ≥35U/L | |
| No | 962/979 (98.3%) |
| Yes | 16/979 (1.6%) |
| Missing | 1/979 (0.1%) |

CAN = Canada, TARGet Kids! = The Applied Research Group for Kids, BMI = Body Mass Index.
Data are presented as n/total (%) based on participants with non-missing haemoglobin, ferritin, and C-reactive protein.
Missing: No answer.
[1] Ferritin≥15ug/L and C-reactive protein≤5mg/L.

#### Table 4.4.2 US NHANES 1999-2018: Participant characteristics of violations (5-11 years)

| | Overall sample Male and Female N=3,030 |
|---|---|
| Reference sample[1] | 1,201/3,030 (39.6%) |
| VIOLATION: BMI age and sex dependent | |
| No | 2,514/3,030 (83.0%) |
| Yes | 487/3,030 (16.1%) |
| No data | 29/3,030 (1.0%) |
| VIOLATION: Doctor told you have diabetes | |
| No | 3,023/3,030 (99.8%) |
| Yes | 4/3,030 (0.1%) |
| No data | 3/3,030 (0.1%) |
| VIOLATION: Still have asthma | |
| No | 146/3,030 (4.8%) |
| Yes | 292/3,030 (9.6%) |
| No data | 12/3,030 (0.4%) |
| Missing | 2,580/3,030 (85.1%) |
| VIOLATION: Taking treatment for anemia/past 3 months | |
| No | 2,992/3,030 (98.7%) |
| Yes | 36/3,030 (1.2%) |
| No data | 2/3,030 (0.1%) |
| VIOLATION: Overnight hospital patient in last year | |
| No | 2,926/3,030 (96.6%) |
| Yes | 100/3,030 (3.3%) |
| No data | 4/3,030 (0.1%) |
| VIOLATION: SP have head cold or chest cold? | |
| No | 2,121/3,030 (70.0%) |
| Yes | 742/3,030 (24.5%) |
| No data | 167/3,030 (5.5%) |
| VIOLATION: SP have stomach or intestinal illness? | |
| No | 2,568/3,030 (84.8%) |
| Yes | 300/3,030 (9.9%) |
| No data | 162/3,030 (5.3%) |
| VIOLATION: SP have flu, pneumonia, ear infection? | |
| No | 2,677/3,030 (88.3%) |
| Yes | 188/3,030 (6.2%) |
| No data | 165/3,030 (5.4%) |
| VIOLATION: Pregnancy status at exam | |
| No | 137/3,030 (4.5%) |
| No data | 189/3,030 (6.2%) |
| Missing | 2,704/3,030 (89.2%) |
| VIOLATION: Urine Pregnancy Result | |
| No | 34/3,030 (1.1%) |
| No data | 292/3,030 (9.6%) |
| Missing | 2,704/3,030 (89.2%) |
| VIOLATION: Taken prescription medicine, past month | |
| No | 2,471/3,030 (81.6%) |
| Yes | 558/3,030 (18.4%) |
| No data | 1/3,030 (0.0%) |
| VIOLATION: For anemia, such as low iron | |
| No | 120/3,030 (4.0%) |

| | Overall sample<br>Male and Female |
|---|---|
| | **N=3,030** |
| Yes | 7/3,030 (0.2%) |
| No data | 426/3,030 (14.1%) |
| Missing | 2,477/3,030 (81.7%) |
| VIOLATION: Ferritin<15ug/L | |
| No | 2,798/3,030 (92.3%) |
| Yes | 232/3,030 (7.7%) |
| VIOLATION: C-Reactive Protein>5mg/L | |
| No | 2,824/3,030 (93.2%) |
| Yes | 206/3,030 (6.8%) |

US = United States, NHANES = National Health and Nutrition Examination Survey, BMI = Body Mass Index, SP = Survey Participant.
Data are presented as unweighted n/total (%) based on participants with non-missing haemoglobin, ferritin, and C-reactive protein.  Data was combined from 1999-2018. No ferritin was measured in participants 6 to 11 years in survey years 2003-2004, 2005-2006, 2007-2008, 2009-2010, 2015-2016, 2017-2018. No ferritin was measured in participants in survey years 2011-2012 and 2013-2014. VIOLATION: For anemia, such as low iron - only asked in NHANES 2007-2008, 2009-2010, 2015-2016, 2017-2018.
No data: No answer/refused, Don't know; Missing: Not applicable, Not asked.
[1] Ferritin≥15ug/L and C-reactive protein≤5mg/L.

#### Table 4.4.3 CHN CHNS 2009: Participant characteristics of violations (5-11 years)

| | Overall sample Male and Female |
|---|---|
| | N=404 |
| Reference sample[1] | 249/404 (61.6%) |
| VIOLATION: BMI age and sex dependent | |
| No | 340/404 (84.2%) |
| Yes | 50/404 (12.4%) |
| Missing | 14/404 (3.5%) |
| VIOLATION: Illness/Injury - Infectious/parasitic disease | |
| No | 31/404 (7.7%) |
| Missing | 373/404 (92.3%) |
| VIOLATION: Illness/Injury - Heart disease | |
| No | 31/404 (7.7%) |
| Missing | 373/404 (92.3%) |
| VIOLATION: Illness/Injury - Tumor | |
| No | 31/404 (7.7%) |
| Missing | 373/404 (92.3%) |
| VIOLATION: Illness/Injury - Respiratory disease | |
| No | 15/404 (3.7%) |
| Yes | 16/404 (4.0%) |
| Missing | 373/404 (92.3%) |
| VIOLATION: Illness/Injury - Injury | |
| No | 30/404 (7.4%) |
| Yes | 1/404 (0.2%) |
| Missing | 373/404 (92.3%) |
| VIOLATION: Illness/Injury - Alcohol poisoning | |
| No | 31/404 (7.7%) |
| Missing | 373/404 (92.3%) |
| VIOLATION: Illness/Injury - Endocrine disorder | |
| No | 31/404 (7.7%) |
| Missing | 373/404 (92.3%) |
| VIOLATION: Illness/Injury - Hematological disease | |
| No | 31/404 (7.7%) |
| Missing | 373/404 (92.3%) |
| VIOLATION: Illness/Injury - Mental/psychiatric disorder | |
| No | 31/404 (7.7%) |
| Missing | 373/404 (92.3%) |
| VIOLATION: Illness/Injury - Neurological disorder | |
| No | 31/404 (7.7%) |
| Missing | 373/404 (92.3%) |
| VIOLATION: Illness/Injury - Eye/ear/nose/throat/teeth disease | |
| No | 31/404 (7.7%) |
| Missing | 373/404 (92.3%) |
| VIOLATION: Illness/Injury - Digestive disease | |
| No | 29/404 (7.2%) |
| Yes | 2/404 (0.5%) |
| Missing | 373/404 (92.3%) |
| VIOLATION: Illness/Injury - Urinary disease | |
| No | 31/404 (7.7%) |
| Missing | 373/404 (92.3%) |
| VIOLATION: Illness/Injury - Sexual dysfunction | |

| | Overall sample Male and Female |
|---|---|
| | N=404 |
| No | 31/404 (7.7%) |
| Missing | 373/404 (92.3%) |
| VIOLATION: Illness/Injury - Obstetrical/gynecological disease | |
| No | 31/404 (7.7%) |
| Missing | 373/404 (92.3%) |
| VIOLATION: Illness/Injury - Dermatological disease | |
| No | 31/404 (7.7%) |
| Missing | 373/404 (92.3%) |
| VIOLATION: Illness/Injury - Muscular/rheumatological disease | |
| No | 31/404 (7.7%) |
| Missing | 373/404 (92.3%) |
| VIOLATION: Diagnosed with high blood pressure | |
| No | 5/404 (1.2%) |
| Missing | 399/404 (98.8%) |
| VIOLATION: Diagnosed with diabetes | |
| No | 5/404 (1.2%) |
| Missing | 399/404 (98.8%) |
| VIOLATION: Diagnosed with myocardial infarction | |
| Missing | 404/404 (100.0%) |
| VIOLATION: Diagnosed with apoplexy | |
| Missing | 404/404 (100.0%) |
| VIOLATION: Dr. told you that you suffer from asthma | |
| Missing | 404/404 (100.0%) |
| VIOLATION: Past 12 months - had whistling/wheezing in chest | |
| Missing | 404/404 (100.0%) |
| VIOLATION: Last 4 weeks - been sick or injured | |
| No | 367/404 (90.8%) |
| Yes | 28/404 (6.9%) |
| Missing | 9/404 (2.2%) |
| VIOLATION: Last 4 weeks - fever, sore throat, cough | |
| No | 370/404 (91.6%) |
| Yes | 32/404 (7.9%) |
| Missing | 2/404 (0.5%) |
| VIOLATION: Last 4 weeks - diarrhea, stomachache | |
| No | 400/404 (99.0%) |
| Yes | 1/404 (0.2%) |
| Missing | 3/404 (0.7%) |
| VIOLATION: Last 4 weeks - stomachache | |
| No | 401/404 (99.3%) |
| Yes | 1/404 (0.2%) |
| Missing | 2/404 (0.5%) |
| VIOLATION: Last 4 weeks - asthma | |
| No | 402/404 (99.5%) |
| Missing | 2/404 (0.5%) |
| VIOLATION: Last 4 weeks - headache, dizziness | |
| No | 400/404 (99.0%) |
| Yes | 2/404 (0.5%) |
| Missing | 2/404 (0.5%) |
| VIOLATION: Last 4 weeks - joint, muscle pain | |

| | Overall sample Male and Female N=404 |
|---|---|
| No | 402/404 (99.5%) |
| Missing | 2/404 (0.5%) |
| VIOLATION: Last 4 weeks - rash, dermatitis | |
| No | 401/404 (99.3%) |
| Yes | 1/404 (0.2%) |
| Missing | 2/404 (0.5%) |
| VIOLATION: Last 4 weeks - eye/ear disease | |
| No | 402/404 (99.5%) |
| Missing | 2/404 (0.5%) |
| VIOLATION: Last 4 weeks - heart disease/chest pain | |
| No | 402/404 (99.5%) |
| Missing | 2/404 (0.5%) |
| VIOLATION: Last 4 weeks - other infectious disease | |
| No | 399/404 (98.8%) |
| Yes | 2/404 (0.5%) |
| Missing | 3/404 (0.7%) |
| VIOLATION: Last 4 weeks - noncommunicable disease | |
| No | 399/404 (98.8%) |
| Yes | 1/404 (0.2%) |
| No data | 1/404 (0.2%) |
| Missing | 3/404 (0.7%) |
| VIOLATION: Last 4 weeks - days hospitalised | |
| Yes | 1/404 (0.2%) |
| Missing | 403/404 (99.8%) |
| VIOLATION: Taking anti-hypertension drugs | |
| Missing | 404/404 (100.0%) |
| VIOLATION: Province – Guizhou – Altitude | |
| No | 345/404 (85.4%) |
| Yes | 59/404 (14.6%) |
| VIOLATION: Ferritin<15ug/L | |
| No | 392/404 (97.0%) |
| Yes | 12/404 (3.0%) |
| VIOLATION: C-Reactive Protein>5mg/L | |
| No | 383/404 (94.8%) |
| Yes | 21/404 (5.2%) |

CHN = China, CHNS = China Health and Nutrition Survey, BMI = Body Mass Index.
Data are presented as unweighted n/total (%) based on participants with non-missing haemoglobin, ferritin, and C-reactive protein.
No data: No answer/refused, Don't know; Missing: Not applicable, Not asked.
[1] Ferritin≥15ug/L and C-reactive protein≤5mg/L.

#### 4.5 Detailed Individual Exclusions Children (12-17 years)

##### Table 4.5.1 US NHANES 1999-2000: Participant characteristics of violations (12-17 years)

| | Overall sample | |
|---|---|---|
| | **Male N=836** | **Female N=793** |
| Reference sample[1] | 334/836 (40.0%) | 233/793 (29.4%) |
| VIOLATION: BMI age and sex dependent | | |
| No | 641/836 (76.7%) | 631/793 (79.6%) |
| Yes | 191/836 (22.8%) | 157/793 (19.8%) |
| No data | 4/836 (0.5%) | 5/793 (0.6%) |
| VIOLATION: Doctor told you have diabetes | | |
| No | 835/836 (99.9%) | 791/793 (99.7%) |
| Yes | 1/836 (0.1%) | 2/793 (0.3%) |
| VIOLATION: Still have asthma | | |
| No | 48/836 (5.7%) | 39/793 (4.9%) |
| Yes | 79/836 (9.4%) | 74/793 (9.3%) |
| No data | 4/836 (0.5%) | 1/793 (0.1%) |
| Missing | 705/836 (84.3%) | 679/793 (85.6%) |
| VIOLATION: Taking treatment for anemia/past 3 months | | |
| No | 832/836 (99.5%) | 780/793 (98.4%) |
| Yes | 4/836 (0.5%) | 12/793 (1.5%) |
| No data | 0/836 (0.0%) | 1/793 (0.1%) |
| VIOLATION: Overnight hospital patient in last year | | |
| No | 815/836 (97.5%) | 759/793 (95.7%) |
| Yes | 20/836 (2.4%) | 34/793 (4.3%) |
| No data | 1/836 (0.1%) | 0/793 (0.0%) |
| VIOLATION: SP donated blood in past 12 months | | |
| No | 253/836 (30.3%) | 225/793 (28.4%) |
| Yes | 14/836 (1.7%) | 14/793 (1.8%) |
| No data | 15/836 (1.8%) | 9/793 (1.1%) |
| Missing | 554/836 (66.3%) | 545/793 (68.7%) |
| VIOLATION: SP have head cold or chest cold? | | |
| No | 628/836 (75.1%) | 587/793 (74.0%) |
| Yes | 172/836 (20.6%) | 184/793 (23.2%) |
| No data | 36/836 (4.3%) | 22/793 (2.8%) |
| VIOLATION: SP have stomach or intestinal illness? | | |
| No | 732/836 (87.6%) | 693/793 (87.4%) |
| Yes | 70/836 (8.4%) | 79/793 (10.0%) |
| No data | 34/836 (4.1%) | 21/793 (2.6%) |
| VIOLATION: SP have flu, pneumonia, ear infection? | | |
| No | 742/836 (88.8%) | 706/793 (89.0%) |
| Yes | 57/836 (6.8%) | 66/793 (8.3%) |
| No data | 37/836 (4.4%) | 21/793 (2.6%) |
| VIOLATION: Are you pregnant now? | | |
| Yes | 0/836 (0.0%) | 21/793 (2.6%) |
| No data | 0/836 (0.0%) | 343/793 (43.3%) |
| Missing | 836/836 (100.0%) | 429/793 (54.1%) |
| VIOLATION: Taken prescription medicine, past month | | |
| No | 698/836 (83.5%) | 620/793 (78.2%) |
| Yes | 138/836 (16.5%) | 173/793 (21.8%) |

| | Overall sample | |
|---|---|---|
| | **Male** | **Female** |
| | **N=836** | **N=793** |
| VIOLATION: Ferritin<15ug/L | | |
| No | 791/836 (94.6%) | 632/793 (79.7%) |
| Yes | 45/836 (5.4%) | 161/793 (20.3%) |
| VIOLATION: C-Reactive Protein>5mg/L | | |
| No | 785/836 (93.9%) | 717/793 (90.4%) |
| Yes | 51/836 (6.1%) | 76/793 (9.6%) |
| VIOLATION: eGFR<90 ml/min/1.73m$^2$ | | |
| No | 816/836 (97.6%) | 775/793 (97.7%) |
| Yes | 6/836 (0.7%) | 6/793 (0.8%) |
| No data | 5/836 (0.6%) | 5/793 (0.6%) |
| Missing | 9/836 (1.1%) | 7/793 (0.9%) |

US = United States, NHANES = National Health and Nutrition Examination Survey, BMI = Body Mass Index, SP = Survey Participant.
Data are presented as unweighted n/total (%) based on participants with non-missing haemoglobin, ferritin, and C-reactive protein.
No data: No answer/refused, Don't know; Missing: Not applicable, Not asked.
[1] Ferritin≥15ug/L and C-reactive protein≤5mg/L.

#### Table 4.5.2 US NHANES 2001-2002: Participant characteristics of violations (12-17 years)

| | Overall sample | |
|---|---|---|
| | Male<br>N=820 | Female<br>N=879 |
| Reference sample[1] | 291/820 (35.5%) | 245/879 (27.9%) |
| VIOLATION: BMI age and sex dependent | | |
| No | 648/820 (79.0%) | 711/879 (80.9%) |
| Yes | 165/820 (20.1%) | 155/879 (17.6%) |
| No data | 7/820 (0.9%) | 13/879 (1.5%) |
| VIOLATION: Doctor told you have diabetes | | |
| No | 814/820 (99.3%) | 876/879 (99.7%) |
| Yes | 6/820 (0.7%) | 3/879 (0.3%) |
| VIOLATION: Still have asthma | | |
| No | 70/820 (8.5%) | 51/879 (5.8%) |
| Yes | 85/820 (10.4%) | 82/879 (9.3%) |
| No data | 1/820 (0.1%) | 4/879 (0.5%) |
| Missing | 664/820 (81.0%) | 742/879 (84.4%) |
| VIOLATION: Taking treatment for anemia/past 3 months | | |
| No | 818/820 (99.8%) | 868/879 (98.7%) |
| Yes | 2/820 (0.2%) | 11/879 (1.3%) |
| VIOLATION: Overnight hospital patient in last year | | |
| No | 796/820 (97.1%) | 834/879 (94.9%) |
| Yes | 24/820 (2.9%) | 45/879 (5.1%) |
| VIOLATION: SP donated blood in past 12 months | | |
| No | 273/820 (33.3%) | 248/879 (28.2%) |
| Yes | 13/820 (1.6%) | 11/879 (1.3%) |
| No data | 14/820 (1.7%) | 18/879 (2.0%) |
| Missing | 520/820 (63.4%) | 602/879 (68.5%) |
| VIOLATION: SP have head cold or chest cold? | | |
| No | 616/820 (75.1%) | 637/879 (72.5%) |
| Yes | 167/820 (20.4%) | 192/879 (21.8%) |
| No data | 37/820 (4.5%) | 50/879 (5.7%) |
| VIOLATION: SP have stomach or intestinal illness? | | |
| No | 727/820 (88.7%) | 753/879 (85.7%) |
| Yes | 57/820 (7.0%) | 76/879 (8.6%) |
| No data | 36/820 (4.4%) | 50/879 (5.7%) |
| VIOLATION: SP have flu, pneumonia, ear infection? | | |
| No | 735/820 (89.6%) | 772/879 (87.8%) |
| Yes | 49/820 (6.0%) | 57/879 (6.5%) |
| No data | 36/820 (4.4%) | 50/879 (5.7%) |
| VIOLATION: Pregnancy status at exam | | |
| No | 0/820 (0.0%) | 832/879 (94.7%) |
| Yes | 0/820 (0.0%) | 17/879 (1.9%) |
| No data | 0/820 (0.0%) | 30/879 (3.4%) |
| Missing | 820/820 (100.0%) | 0/879 (0.0%) |
| VIOLATION: Urine Pregnancy Result | | |
| No | 0/820 (0.0%) | 849/879 (96.6%) |
| Yes | 0/820 (0.0%) | 17/879 (1.9%) |
| No data | 0/820 (0.0%) | 13/879 (1.5%) |
| Missing | 820/820 (100.0%) | 0/879 (0.0%) |
| VIOLATION: Are you pregnant now? | | |
| No | 0/820 (0.0%) | 17/879 (1.9%) |

| | Overall sample | |
|---|---|---|
| | **Male** | **Female** |
| | **N=820** | **N=879** |
| Yes | 0/820 (0.0%) | 15/879 (1.7%) |
| No data | 0/820 (0.0%) | 842/879 (95.8%) |
| Missing | 820/820 (100.0%) | 5/879 (0.6%) |
| VIOLATION: Taken prescription medicine, past month | | |
| No | 622/820 (75.9%) | 685/879 (77.9%) |
| Yes | 198/820 (24.1%) | 194/879 (22.1%) |
| VIOLATION: Ferritin<15ug/L | | |
| No | 765/820 (93.3%) | 664/879 (75.5%) |
| Yes | 55/820 (6.7%) | 215/879 (24.5%) |
| VIOLATION: C-Reactive Protein>5mg/L | | |
| No | 766/820 (93.4%) | 822/879 (93.5%) |
| Yes | 54/820 (6.6%) | 57/879 (6.5%) |
| VIOLATION: eGFR<90 ml/min/1.73m[2] | | |
| No | 807/820 (98.4%) | 862/879 (98.1%) |
| Yes | 0/820 (0.0%) | 7/879 (0.8%) |
| No data | 7/820 (0.9%) | 5/879 (0.6%) |
| Missing | 6/820 (0.7%) | 5/879 (0.6%) |

US = United States, NHANES = National Health and Nutrition Examination Survey, BMI = Body Mass Index, SP = Survey Participant.
Data are presented as unweighted n/total (%) based on participants with non-missing haemoglobin, ferritin, and C-reactive protein.
No data: No answer/refused, Don't know; Missing: Not applicable, Not asked.
[1] Ferritin≥15ug/L and C-reactive protein≤5mg/L.

#### Table 4.5.3 US NHANES 2003-2004: Participant characteristics of violations (Females) (12-17 years)

| | Overall sample Female N=730 |
|---|---|
| Reference sample[1] | 243/730 (33.3%) |
| VIOLATION: BMI age and sex dependent | |
| No | 590/730 (80.8%) |
| Yes | 131/730 (17.9%) |
| No data | 9/730 (1.2%) |
| VIOLATION: Doctor told you have diabetes | |
| No | 724/730 (99.2%) |
| Yes | 4/730 (0.5%) |
| No data | 2/730 (0.3%) |
| VIOLATION: Still have asthma | |
| No | 39/730 (5.3%) |
| Yes | 80/730 (11.0%) |
| No data | 3/730 (0.4%) |
| Missing | 608/730 (83.3%) |
| VIOLATION: Taking treatment for anemia/past 3 months | |
| No | 714/730 (97.8%) |
| Yes | 16/730 (2.2%) |
| VIOLATION: Overnight hospital patient in last year | |
| No | 698/730 (95.6%) |
| Yes | 32/730 (4.4%) |
| VIOLATION: SP donated blood in past 12 months | |
| No | 221/730 (30.3%) |
| Yes | 11/730 (1.5%) |
| No data | 10/730 (1.4%) |
| Missing | 488/730 (66.8%) |
| VIOLATION: SP have head cold or chest cold? | |
| No | 535/730 (73.3%) |
| Yes | 157/730 (21.5%) |
| No data | 38/730 (5.2%) |
| VIOLATION: SP have stomach or intestinal illness? | |
| No | 646/730 (88.5%) |
| Yes | 47/730 (6.4%) |
| No data | 37/730 (5.1%) |
| VIOLATION: SP have flu, pneumonia, ear infection? | |
| No | 647/730 (88.6%) |
| Yes | 46/730 (6.3%) |
| No data | 37/730 (5.1%) |
| VIOLATION: Pregnancy status at exam | |
| No | 697/730 (95.5%) |
| Yes | 20/730 (2.7%) |
| No data | 13/730 (1.8%) |
| VIOLATION: Urine Pregnancy Result | |
| No | 705/730 (96.6%) |
| Yes | 20/730 (2.7%) |
| No data | 5/730 (0.7%) |
| VIOLATION: Are you pregnant now? | |
| No | 24/730 (3.3%) |

| | Overall sample<br>Female<br>N=730 |
|---|---|
| Yes | 15/730 (2.1%) |
| No data | 691/730 (94.7%) |
| VIOLATION: Taken prescription medicine, past month | |
| No | 555/730 (76.0%) |
| Yes | 174/730 (23.8%) |
| No data | 1/730 (0.1%) |
| VIOLATION: Ferritin<15ug/L | |
| No | 602/730 (82.5%) |
| Yes | 128/730 (17.5%) |
| VIOLATION: C-Reactive Protein>5mg/L | |
| No | 660/730 (90.4%) |
| Yes | 70/730 (9.6%) |
| VIOLATION: eGFR<90 ml/min/1.73m$^2$ | |
| No | 715/730 (97.9%) |
| Yes | 5/730 (0.7%) |
| No data | 10/730 (1.4%) |

US = United States, NHANES = National Health and Nutrition Examination Survey, BMI = Body Mass Index, SP = Survey Participant.
Data are presented as unweighted n/total (%) based on participants with non-missing haemoglobin, ferritin, and C-reactive protein. No ferritin was collected in males.
No data: No answer/refused, Don't know; Missing: Not applicable, Not asked.
[1] Ferritin≥15ug/L and C-reactive protein≤5mg/L.

#### Table 4.5.4 US NHANES 2005-2006: Participant characteristics of violations (Females) (12-17 years)

| | Overall sample<br>Female<br>N=732 |
|---|---|
| Reference sample[1] | 220/732 (30.1%) |
| VIOLATION: BMI age and sex dependent | |
| No | 552/732 (75.4%) |
| Yes | 175/732 (23.9%) |
| No data | 5/732 (0.7%) |
| VIOLATION: Doctor told you have diabetes | |
| No | 728/732 (99.5%) |
| Yes | 3/732 (0.4%) |
| No data | 1/732 (0.1%) |
| VIOLATION: Still have asthma | |
| No | 37/732 (5.1%) |
| Yes | 80/732 (10.9%) |
| No data | 2/732 (0.3%) |
| Missing | 613/732 (83.7%) |
| VIOLATION: Taking treatment for anemia/past 3 months | |
| No | 725/732 (99.0%) |
| Yes | 7/732 (1.0%) |
| VIOLATION: Overnight hospital patient in last year | |
| No | 692/732 (94.5%) |
| Yes | 40/732 (5.5%) |
| VIOLATION: SP donated blood in past 12 months | |
| No | 223/732 (30.5%) |
| Yes | 11/732 (1.5%) |
| No data | 19/732 (2.6%) |
| Missing | 479/732 (65.4%) |
| VIOLATION: SP have head cold or chest cold? | |
| No | 529/732 (72.3%) |
| Yes | 160/732 (21.9%) |
| No data | 43/732 (5.9%) |
| VIOLATION: SP have stomach or intestinal illness? | |
| No | 618/732 (84.4%) |
| Yes | 71/732 (9.7%) |
| No data | 43/732 (5.9%) |
| VIOLATION: SP have flu, pneumonia, ear infection? | |
| No | 646/732 (88.3%) |
| Yes | 43/732 (5.9%) |
| No data | 43/732 (5.9%) |
| VIOLATION: Pregnancy status at exam | |
| No | 697/732 (95.2%) |
| Yes | 17/732 (2.3%) |
| No data | 18/732 (2.5%) |
| VIOLATION: Urine Pregnancy Result | |
| No | 704/732 (96.2%) |
| Yes | 16/732 (2.2%) |
| No data | 12/732 (1.6%) |
| VIOLATION: Are you pregnant now? | |
| No | 28/732 (3.8%) |

| | Overall sample<br>Female<br>N=732 |
|---|---|
| Yes | 14/732 (1.9%) |
| No data | 690/732 (94.3%) |
| VIOLATION: Taken prescription medicine, past month | |
| No | 561/732 (76.6%) |
| Yes | 171/732 (23.4%) |
| VIOLATION: Ferritin<15ug/L | |
| No | 620/732 (84.7%) |
| Yes | 112/732 (15.3%) |
| VIOLATION: C-Reactive Protein>5mg/L | |
| No | 663/732 (90.6%) |
| Yes | 69/732 (9.4%) |
| VIOLATION: eGFR<90 ml/min/1.73m$^2$ | |
| No | 715/732 (97.7%) |
| Yes | 13/732 (1.8%) |
| No data | 4/732 (0.5%) |

US = United States, NHANES = National Health and Nutrition Examination Survey, BMI = Body Mass Index, SP = Survey Participant.
Data are presented as unweighted n/total (%) based on participants with non-missing haemoglobin, ferritin, and C-reactive protein. No ferritin was collected in males.
No data: No answer/refused, Don't know; Missing: Not applicable, Not asked.
[1] Ferritin≥15ug/L and C-reactive protein≤5mg/L.

#### Table 4.5.5 US NHANES 2007-2008: Participant characteristics of violations (Females) (12-17 years)

| | Overall sample<br>Female<br>N=383 |
|---|---|
| Reference sample[1] | 119/383 (31.1%) |
| VIOLATION: BMI age and sex dependent | |
| No | 293/383 (76.5%) |
| Yes | 82/383 (21.4%) |
| No data | 8/383 (2.1%) |
| VIOLATION: Doctor told you have diabetes | |
| No | 380/383 (99.2%) |
| Yes | 3/383 (0.8%) |
| VIOLATION: Still have asthma | |
| No | 28/383 (7.3%) |
| Yes | 46/383 (12.0%) |
| Missing | 309/383 (80.7%) |
| VIOLATION: Taking treatment for anemia/past 3 months | |
| No | 381/383 (99.5%) |
| Yes | 2/383 (0.5%) |
| VIOLATION: Overnight hospital patient in last year | |
| No | 367/383 (95.8%) |
| Yes | 16/383 (4.2%) |
| VIOLATION: SP donated blood in past 12 months | |
| No | 111/383 (29.0%) |
| Yes | 11/383 (2.9%) |
| No data | 8/383 (2.1%) |
| Missing | 253/383 (66.1%) |
| VIOLATION: SP have head cold or chest cold? | |
| No | 269/383 (70.2%) |
| Yes | 87/383 (22.7%) |
| No data | 27/383 (7.0%) |
| VIOLATION: SP have stomach or intestinal illness? | |
| No | 317/383 (82.8%) |
| Yes | 39/383 (10.2%) |
| No data | 27/383 (7.0%) |
| VIOLATION: SP have flu, pneumonia, ear infection? | |
| No | 337/383 (88.0%) |
| Yes | 19/383 (5.0%) |
| No data | 27/383 (7.0%) |
| VIOLATION: Taken prescription medicine, past month | |
| No | 288/383 (75.2%) |
| Yes | 94/383 (24.5%) |
| No data | 1/383 (0.3%) |
| VIOLATION: For anemia, such as low iron | |
| Yes | 1/383 (0.3%) |
| No data | 382/383 (99.7%) |
| VIOLATION: Ferritin<15ug/L | |
| No | 323/383 (84.3%) |
| Yes | 60/383 (15.7%) |
| VIOLATION: C-Reactive Protein>5mg/L | |
| No | 350/383 (91.4%) |

| | Overall sample Female N=383 |
|---|---|
| Yes | 33/383 (8.6%) |
| VIOLATION: eGFR<90 ml/min/1.73m$^2$ | |
| No | 371/383 (96.9%) |
| Yes | 8/383 (2.1%) |
| No data | 4/383 (1.0%) |

US = United States, NHANES = National Health and Nutrition Examination Survey, BMI = Body Mass Index, SP = Survey Participant.
Data are presented as unweighted n/total (%) based on participants with non-missing haemoglobin, ferritin, and C-reactive protein. No ferritin was collected in males.
No data: No answer/refused, Don't know; Missing: Not applicable, Not asked.
[1] Ferritin≥15ug/L and C-reactive protein≤5mg/L.

#### Table 4.5.6 US NHANES 2009-2010: Participant characteristics of violations (Females) (12-17 years)

| | Overall sample Female N=422 |
|---|---|
| Reference sample[1] | 147/422 (34.8%) |
| VIOLATION: BMI age and sex dependent | |
| No | 334/422 (79.1%) |
| Yes | 83/422 (19.7%) |
| No data | 5/422 (1.2%) |
| VIOLATION: Doctor told you have diabetes | |
| No | 421/422 (99.8%) |
| Yes | 1/422 (0.2%) |
| VIOLATION: Still have asthma | |
| No | 21/422 (5.0%) |
| Yes | 45/422 (10.7%) |
| Missing | 356/422 (84.4%) |
| VIOLATION: Taking treatment for anemia/past 3 months | |
| No | 419/422 (99.3%) |
| Yes | 3/422 (0.7%) |
| VIOLATION: Overnight hospital patient in last year | |
| No | 405/422 (96.0%) |
| Yes | 17/422 (4.0%) |
| VIOLATION: SP donated blood in past 12 months | |
| No | 122/422 (28.9%) |
| Yes | 15/422 (3.6%) |
| No data | 12/422 (2.8%) |
| Missing | 273/422 (64.7%) |
| VIOLATION: SP have head cold or chest cold? | |
| No | 319/422 (75.6%) |
| Yes | 75/422 (17.8%) |
| No data | 28/422 (6.6%) |
| VIOLATION: SP have stomach or intestinal illness? | |
| No | 367/422 (87.0%) |
| Yes | 28/422 (6.6%) |
| No data | 27/422 (6.4%) |
| VIOLATION: SP have flu, pneumonia, ear infection? | |
| No | 372/422 (88.2%) |
| Yes | 23/422 (5.5%) |
| No data | 27/422 (6.4%) |
| VIOLATION: Taken prescription medicine, past month | |
| No | 339/422 (80.3%) |
| Yes | 83/422 (19.7%) |
| VIOLATION: For anemia, such as low iron | |
| Yes | 3/422 (0.7%) |
| No data | 419/422 (99.3%) |
| VIOLATION: Ferritin<15ug/L | |
| No | 337/422 (79.9%) |
| Yes | 85/422 (20.1%) |
| VIOLATION: C-Reactive Protein>5mg/L | |
| No | 403/422 (95.5%) |
| Yes | 19/422 (4.5%) |

| | Overall sample<br>Female<br>N=422 |
|---|---|
| VIOLATION: eGFR<90 ml/min/1.73m$^2$ | |
| No | 417/422 (98.8%) |
| Yes | 2/422 (0.5%) |
| No data | 3/422 (0.7%) |

US = United States, NHANES = National Health and Nutrition Examination Survey, BMI = Body Mass Index, SP = Survey Participant.
Data are presented as unweighted n/total (%) based on participants with non-missing haemoglobin, ferritin, and C-reactive protein. No ferritin was collected in males.
No data: No answer/refused, Don't know; Missing: Not applicable, Not asked.
[1] Ferritin≥15ug/L and C-reactive protein≤5mg/L.

#### Table 4.5.7 US NHANES 2015-2016: Participant characteristics of violations (Females) (12-17 years)

| | Overall sample Female N=420 |
|---|---|
| Reference sample[1] | 154/420 (36.7%) |
| VIOLATION: BMI age and sex dependent | |
| No | 321/420 (76.4%) |
| Yes | 92/420 (21.9%) |
| No data | 7/420 (1.7%) |
| VIOLATION: Doctor told you have diabetes | |
| No | 418/420 (99.5%) |
| Yes | 2/420 (0.5%) |
| VIOLATION: Still have asthma | |
| No | 31/420 (7.4%) |
| Yes | 51/420 (12.1%) |
| Missing | 338/420 (80.5%) |
| VIOLATION: Taking treatment for anemia/past 3 months | |
| No | 410/420 (97.6%) |
| Yes | 10/420 (2.4%) |
| VIOLATION: Ever been told you have jaundice? | |
| No | 417/420 (99.3%) |
| Yes | 2/420 (0.5%) |
| No data | 1/420 (0.2%) |
| VIOLATION: Overnight hospital patient in last year | |
| No | 408/420 (97.1%) |
| Yes | 12/420 (2.9%) |
| VIOLATION: SP donated blood in past 12 months | |
| No | 135/420 (32.1%) |
| Yes | 19/420 (4.5%) |
| No data | 6/420 (1.4%) |
| Missing | 260/420 (61.9%) |
| VIOLATION: SP have head cold or chest cold? | |
| No | 338/420 (80.5%) |
| Yes | 62/420 (14.8%) |
| No data | 20/420 (4.8%) |
| VIOLATION: SP have stomach or intestinal illness? | |
| No | 366/420 (87.1%) |
| Yes | 34/420 (8.1%) |
| No data | 20/420 (4.8%) |
| VIOLATION: SP have flu, pneumonia, ear infection? | |
| No | 384/420 (91.4%) |
| Yes | 16/420 (3.8%) |
| No data | 20/420 (4.8%) |
| VIOLATION: Taken prescription medicine, past month | |
| No | 328/420 (78.1%) |
| Yes | 91/420 (21.7%) |
| No data | 1/420 (0.2%) |
| VIOLATION: For anemia, such as low iron | |
| Yes | 8/420 (1.9%) |
| No data | 412/420 (98.1%) |
| VIOLATION: Ferritin<15ug/L | |

| | **Overall sample Female N=420** |
|---|---|
| No | 332/420 (79.0%) |
| Yes | 88/420 (21.0%) |
| VIOLATION: C-Reactive Protein>5mg/L | |
| No | 394/420 (93.8%) |
| Yes | 26/420 (6.2%) |
| VIOLATION: eGFR<90 ml/min/1.73m$^2$ | |
| No | 418/420 (99.5%) |
| Yes | 2/420 (0.5%) |

US = United States, NHANES = National Health and Nutrition Examination Survey, BMI = Body Mass Index, SP = Survey Participant.
Data are presented as unweighted n/total (%) based on participants with non-missing haemoglobin, ferritin, and C-reactive protein. No ferritin was collected in males.
No data: No answer/refused, Don't know; Missing: Not applicable, Not asked.
[1] Ferritin≥15ug/L and C-reactive protein≤5mg/L.

Table 4.5.8 US NHANES 2017-2018: Participant characteristics of violations (12-17 years)

| | Overall sample | |
|---|---|---|
| | Male N=383 | Female N=366 |
| Reference sample[1] | 141/383 (36.8%) | 109/366 (29.8%) |
| VIOLATION: BMI age and sex dependent | | |
| No | 283/383 (73.9%) | 274/366 (74.9%) |
| Yes | 96/383 (25.1%) | 88/366 (24.0%) |
| No data | 4/383 (1.0%) | 4/366 (1.1%) |
| VIOLATION: Doctor told you have diabetes | | |
| No | 380/383 (99.2%) | 362/366 (98.9%) |
| Yes | 3/383 (0.8%) | 4/366 (1.1%) |
| VIOLATION: Still have asthma | | |
| No | 31/383 (8.1%) | 26/366 (7.1%) |
| Yes | 46/383 (12.0%) | 32/366 (8.7%) |
| No data | 2/383 (0.5%) | 1/366 (0.3%) |
| Missing | 304/383 (79.4%) | 307/366 (83.9%) |
| VIOLATION: Taking treatment for anemia/past 3 months | | |
| No | 383/383 (100.0%) | 353/366 (96.4%) |
| Yes | 0/383 (0.0%) | 12/366 (3.3%) |
| No data | 0/383 (0.0%) | 1/366 (0.3%) |
| VIOLATION: Ever told you had any liver condition (age12-19) | | |
| No | 382/383 (99.7%) | 366/366 (100.0%) |
| Yes | 1/383 (0.3%) | 0/366 (0.0%) |
| VIOLATION: Ever been told you have jaundice? | | |
| No | 382/383 (99.7%) | 364/366 (99.5%) |
| Yes | 0/383 (0.0%) | 1/366 (0.3%) |
| No data | 1/383 (0.3%) | 1/366 (0.3%) |
| VIOLATION: SP donated blood in past 12 months | | |
| No | 105/383 (27.4%) | 120/366 (32.8%) |
| Yes | 17/383 (4.4%) | 15/366 (4.1%) |
| No data | 5/383 (1.3%) | 5/366 (1.4%) |
| Missing | 256/383 (66.8%) | 226/366 (61.7%) |
| VIOLATION: Overnight hospital patient in last year | | |
| No | 376/383 (98.2%) | 358/366 (97.8%) |
| Yes | 7/383 (1.8%) | 8/366 (2.2%) |
| VIOLATION: SP have head cold or chest cold? | | |
| No | 320/383 (83.6%) | 299/366 (81.7%) |
| Yes | 48/383 (12.5%) | 55/366 (15.0%) |
| No data | 15/383 (3.9%) | 12/366 (3.3%) |
| VIOLATION: SP have stomach or intestinal illness? | | |
| No | 343/383 (89.6%) | 336/366 (91.8%) |
| Yes | 25/383 (6.5%) | 21/366 (5.7%) |
| No data | 15/383 (3.9%) | 9/366 (2.5%) |
| VIOLATION: SP have flu, pneumonia, ear infection? | | |
| No | 351/383 (91.6%) | 341/366 (93.2%) |
| Yes | 17/383 (4.4%) | 15/366 (4.1%) |
| No data | 15/383 (3.9%) | 10/366 (2.7%) |
| VIOLATION: For anemia, such as low iron | | |
| No | 382/383 (99.7%) | 358/366 (97.8%) |
| Yes | 1/383 (0.3%) | 8/366 (2.2%) |
| VIOLATION: Ferritin<15ug/L | | |

| | Overall sample | |
|---|---|---|
| | Male<br>N=383 | Female<br>N=366 |
| No | 367/383 (95.8%) | 287/366 (78.4%) |
| Yes | 16/383 (4.2%) | 79/366 (21.6%) |
| VIOLATION: C-Reactive Protein>5mg/L | | |
| No | 351/383 (91.6%) | 339/366 (92.6%) |
| Yes | 32/383 (8.4%) | 27/366 (7.4%) |
| VIOLATION: eGFR<90 ml/min/1.73m$^2$ | | |
| No | 380/383 (99.2%) | 361/366 (98.6%) |
| Yes | 2/383 (0.5%) | 3/366 (0.8%) |
| No data | 1/383 (0.3%) | 2/366 (0.5%) |

US = United States, NHANES = National Health and Nutrition Examination Survey, BMI = Body Mass Index, SP = Survey Participant.
Data are presented as unweighted n/total (%) based on participants with non-missing haemoglobin, ferritin, and C-reactive protein.
No data: No answer/refused, Don't know; Missing: Not applicable, Not asked.
[1] Ferritin≥15ug/L and C-reactive protein≤5mg/L.

#### Table 4.5.9 AUS NHS 2011-2012: Participant characteristics of violations (12-17 years)

| | Overall sample | |
|---|---|---|
| | Male<br>N=209 | Female<br>N=243 |
| Reference sample[1] | | |
| No | 119/209 (56.9%) | 164/243 (67.5%) |
| Yes | 90/209 (43.1%) | 79/243 (32.5%) |
| VIOLATION: BMI violation* | | |
| No | 172/209 (82.3%) | 189/243 (77.8%) |
| Yes | 23/209 (11.0%) | 37/243 (15.2%) |
| Missing | 14/209 (6.7%) | 17/243 (7.0%) |
| VIOLATION: Co-morbidities | | |
| No | 176/209 (84.2%) | 202/243 (83.1%) |
| Yes | 33/209 (15.8%) | 41/243 (16.9%) |
| VIOLATION: Dialysis | | |
| No | 209/209 (100.0%) | 243/243 (100.0%) |
| VIOLATION: Hospital last 12 months | | |
| No | Not displayed† | Not displayed† |
| Yes | <10/209 (<4.8%) | <10/243 (<4.1%) |
| VIOLATION: Hospital last 2 weeks | | |
| No | Not displayed† | Not displayed† |
| Yes | <10/209 (<4.8%) | <10/243 (<4.1%) |
| VIOLATION: Time off work or study due to illness | | |
| No | 162/206 (78.6%) | 184/235 (78.3%) |
| Yes | 44/206 (21.4%) | 51/235 (21.7%) |
| VIOLATION: Time off work due to illness | | |
| No | Not displayed† | Not displayed† |
| Yes | <10/209 (<4.8%) | <10/243 (<4.1%) |
| Missing | 169/209 (80.9%) | 168/243 (69.1%) |
| VIOLATION: Time off study due to illness | | |
| No | 161/204 (78.9%) | 181/229 (79.0%) |
| Yes | 43/204 (21.1%) | 48/229 (21.0%) |
| VIOLATION: Medication | | |
| No | 153/209 (73.2%) | 176/243 (72.4%) |
| Yes | 56/209 (26.8%) | 67/243 (27.6%) |
| VIOLATION: Smoking | | |
| No | Not displayed† | 133/243 (54.7%) |
| Yes | <10/209 (<4.8%) | 12/243 (4.9%) |
| Missing | 105/209 (50.2%) | 98/243 (40.3%) |
| VIOLATION: Alcohol | | |
| No | Not displayed† | Not displayed† |
| Yes | <10/209 (<4.8%) | <10/243 (<4.1%) |
| Missing | 189/209 (90.4%) | 218/243 (89.7%) |
| VIOLATION: Ferritin<15ug/L | | |
| No | Not displayed† | Not displayed† |
| Yes | <10/209 (<4.8%) | 34/243 (14.0%) |
| VIOLATION: C-Reactive Protein>5mg/L | | |
| No | 197/209 (94.3%) | 230/243 (94.7%) |
| Yes | 12/209 (5.7%) | 13/243 (5.3%) |

AUS = Australia, NHS = National Health Survey, BMI = Body Mass Index.

Data are presented as unweighted n/total (%) based on participants with non-missing haemoglobin, ferritin, and C-reactive protein.
*BMI – underweight class 1/2/3 and obese class 1.
† Not displayed due to Australian Bureau of Statistics requirements for output clearance.
[1] Ferritin≥15ug/L and C-reactive protein≤5mg/L.

#### Table 4.5.10 AUS NNPAS 2011-2012: Participant characteristics of violations (12-17 years)

| | Overall sample | |
|---|---|---|
| | Male N=149 | Female N=133 |
| Reference sample[1] | | |
| No | 32/149 (21.5%) | 39/133 (29.3%) |
| Yes | 117/149 (78.5%) | 94/133 (70.7%) |
| VIOLATION: BMI violation* | | |
| No | Not displayed† | Not displayed† |
| Yes | 11/149 (7.4%) | 20/133 (15.0%) |
| Missing | <10 | <10 |
| VIOLATION: Co-morbidities | | |
| No | Not displayed† | Not displayed† |
| Yes | <10/149 (<6.7%) | <10/133 (<7.5%) |
| VIOLATION: Smoking | | |
| No | Not displayed† | Not displayed† |
| Yes | <10/149 (<6.7%) | <10/133 (<7.5%) |
| Missing | 69/149 (46.3%) | 64/133 (48.1%) |
| VIOLATION: Ferritin<15ug/L | | |
| No | Not displayed† | 120/133 (90.2%) |
| Yes | <10/149 (<6.7%) | 13/133 (9.8%) |
| VIOLATION: C-Reactive Protein>5mg/L | | |
| No | 136/149 (91.3%) | Not displayed† |
| Yes | 13/149 (8.7%) | <10/133 (<7.5%) |

AUS = Australia, NNPAS = National Nutrition and Physical Activity Survey, BMI = Body Mass Index.
Data are presented as unweighted n/total (%) based on participants with non-missing haemoglobin, ferritin, and C-reactive protein.
*BMI – underweight class 1/2/3 and obese class 1.
† Not displayed due to Australian Bureau of Statistics requirements for output clearance.
[1] Ferritin≥15ug/L and C-reactive protein≤5mg/L.

#### Table 4.5.11 CHN CHNS 2009: Participant characteristics of violations (12-17 years)

| | Overall sample | |
|---|---|---|
| | Male<br>N=223 | Female<br>N=187 |
| Reference sample[1] | 130/223 (58.3%) | 102/187 (54.5%) |
| VIOLATION: BMI age and sex dependent | | |
| No | 186/223 (83.4%) | 171/187 (91.4%) |
| Yes | 23/223 (10.3%) | 13/187 (7.0%) |
| Missing | 14/223 (6.3%) | 3/187 (1.6%) |
| VIOLATION: Illness/Injury - Infectious/parasitic disease | | |
| No | 15/223 (6.7%) | 12/187 (6.4%) |
| Missing | 208/223 (93.3%) | 175/187 (93.6%) |
| VIOLATION: Illness/Injury - Heart disease | | |
| No | 14/223 (6.3%) | 12/187 (6.4%) |
| Yes | 1/223 (0.4%) | 0/187 (0.0%) |
| Missing | 208/223 (93.3%) | 175/187 (93.6%) |
| VIOLATION: Illness/Injury - Tumor | | |
| No | 15/223 (6.7%) | 12/187 (6.4%) |
| Missing | 208/223 (93.3%) | 175/187 (93.6%) |
| VIOLATION: Illness/Injury - Respiratory disease | | |
| No | 10/223 (4.5%) | 6/187 (3.2%) |
| Yes | 5/223 (2.2%) | 6/187 (3.2%) |
| Missing | 208/223 (93.3%) | 175/187 (93.6%) |
| VIOLATION: Illness/Injury - Injury | | |
| No | 14/223 (6.3%) | 12/187 (6.4%) |
| Yes | 1/223 (0.4%) | 0/187 (0.0%) |
| Missing | 208/223 (93.3%) | 175/187 (93.6%) |
| VIOLATION: Illness/Injury - Alcohol poisoning | | |
| No | 15/223 (6.7%) | 12/187 (6.4%) |
| Missing | 208/223 (93.3%) | 175/187 (93.6%) |
| VIOLATION: Illness/Injury - Endocrine disorder | | |
| No | 15/223 (6.7%) | 12/187 (6.4%) |
| Missing | 208/223 (93.3%) | 175/187 (93.6%) |
| VIOLATION: Illness/Injury - Hematological disease | | |
| No | 15/223 (6.7%) | 12/187 (6.4%) |
| Missing | 208/223 (93.3%) | 175/187 (93.6%) |
| VIOLATION: Illness/Injury - Mental/psychiatric disorder | | |
| No | 15/223 (6.7%) | 12/187 (6.4%) |
| Missing | 208/223 (93.3%) | 175/187 (93.6%) |
| VIOLATION: Illness/Injury - Neurological disorder | | |
| No | 15/223 (6.7%) | 12/187 (6.4%) |
| Missing | 208/223 (93.3%) | 175/187 (93.6%) |
| VIOLATION: Illness/Injury - Eye/ear/nose/throat/teeth disease | | |
| No | 14/223 (6.3%) | 12/187 (6.4%) |
| Yes | 1/223 (0.4%) | 0/187 (0.0%) |
| Missing | 208/223 (93.3%) | 175/187 (93.6%) |
| VIOLATION: Illness/Injury - Digestive disease | | |
| No | 14/223 (6.3%) | 10/187 (5.3%) |
| Yes | 1/223 (0.4%) | 2/187 (1.1%) |
| Missing | 208/223 (93.3%) | 175/187 (93.6%) |
| VIOLATION: Illness/Injury - Urinary disease | | |
| No | 15/223 (6.7%) | 12/187 (6.4%) |

| | Overall sample | |
|---|---|---|
| | Male<br>N=223 | Female<br>N=187 |
| Missing | 208/223 (93.3%) | 175/187 (93.6%) |
| VIOLATION: Illness/Injury - Sexual dysfunction | | |
| No | 15/223 (6.7%) | 12/187 (6.4%) |
| Missing | 208/223 (93.3%) | 175/187 (93.6%) |
| VIOLATION: Illness/Injury - Obstetrical/gynecological disease | | |
| No | 15/223 (6.7%) | 12/187 (6.4%) |
| Missing | 208/223 (93.3%) | 175/187 (93.6%) |
| VIOLATION: Illness/Injury - Dermatological disease | | |
| No | 14/223 (6.3%) | 12/187 (6.4%) |
| Yes | 1/223 (0.4%) | 0/187 (0.0%) |
| Missing | 208/223 (93.3%) | 175/187 (93.6%) |
| VIOLATION: Illness/Injury - Muscular/rheumatological disease | | |
| No | 15/223 (6.7%) | 12/187 (6.4%) |
| Missing | 208/223 (93.3%) | 175/187 (93.6%) |
| VIOLATION: Diagnosed with high blood pressure | | |
| No | 217/223 (97.3%) | 186/187 (99.5%) |
| No data | 0/223 (0.0%) | 1/187 (0.5%) |
| Missing | 6/223 (2.7%) | 0/187 (0.0%) |
| VIOLATION: Diagnosed with diabetes | | |
| No | 217/223 (97.3%) | 187/187 (100.0%) |
| Missing | 6/223 (2.7%) | 0/187 (0.0%) |
| VIOLATION: Diagnosed with myocardial infarction | | |
| No | 1/223 (0.4%) | 2/187 (1.1%) |
| Missing | 222/223 (99.6%) | 185/187 (98.9%) |
| VIOLATION: Diagnosed with apoplexy | | |
| No | 1/223 (0.4%) | 2/187 (1.1%) |
| Missing | 222/223 (99.6%) | 185/187 (98.9%) |
| VIOLATION: Dr. told you that you suffer from asthma | | |
| No | 1/223 (0.4%) | 2/187 (1.1%) |
| Missing | 222/223 (99.6%) | 185/187 (98.9%) |
| VIOLATION: Past 12 months - had whistling/wheezing in chest | | |
| Missing | 223/223 (100.0%) | 187/187 (100.0%) |
| VIOLATION: Last 4 weeks - been sick or injured | | |
| No | 201/223 (90.1%) | 169/187 (90.4%) |
| Yes | 17/223 (7.6%) | 14/187 (7.5%) |
| No data | 0/223 (0.0%) | 2/187 (1.1%) |
| Missing | 5/223 (2.2%) | 2/187 (1.1%) |
| VIOLATION: Last 4 weeks - fever, sore throat, cough | | |
| No | 202/223 (90.6%) | 164/187 (87.7%) |
| Yes | 18/223 (8.1%) | 20/187 (10.7%) |
| No data | 0/223 (0.0%) | 2/187 (1.1%) |
| Missing | 3/223 (1.3%) | 1/187 (0.5%) |
| VIOLATION: Last 4 weeks - diarrhea, stomachache | | |
| No | 216/223 (96.9%) | 178/187 (95.2%) |
| Yes | 4/223 (1.8%) | 5/187 (2.7%) |
| No data | 0/223 (0.0%) | 3/187 (1.6%) |
| Missing | 3/223 (1.3%) | 1/187 (0.5%) |
| VIOLATION: Last 4 weeks - stomachache | | |
| No | 219/223 (98.2%) | 181/187 (96.8%) |

| | Overall sample | |
|---|---|---|
| | Male<br>N=223 | Female<br>N=187 |
| Yes | 1/223 (0.4%) | 4/187 (2.1%) |
| No data | 0/223 (0.0%) | 2/187 (1.1%) |
| Missing | 3/223 (1.3%) | 0/187 (0.0%) |
| VIOLATION: Last 4 weeks - asthma | | |
| No | 220/223 (98.7%) | 184/187 (98.4%) |
| Yes | 0/223 (0.0%) | 1/187 (0.5%) |
| No data | 0/223 (0.0%) | 2/187 (1.1%) |
| Missing | 3/223 (1.3%) | 0/187 (0.0%) |
| VIOLATION: Last 4 weeks - headache, dizziness | | |
| No | 216/223 (96.9%) | 177/187 (94.7%) |
| Yes | 4/223 (1.8%) | 7/187 (3.7%) |
| No data | 0/223 (0.0%) | 2/187 (1.1%) |
| Missing | 3/223 (1.3%) | 1/187 (0.5%) |
| VIOLATION: Last 4 weeks - joint, muscle pain | | |
| No | 217/223 (97.3%) | 183/187 (97.9%) |
| Yes | 3/223 (1.3%) | 1/187 (0.5%) |
| No data | 0/223 (0.0%) | 2/187 (1.1%) |
| Missing | 3/223 (1.3%) | 1/187 (0.5%) |
| VIOLATION: Last 4 weeks - rash, dermatitis | | |
| No | 219/223 (98.2%) | 183/187 (97.9%) |
| Yes | 1/223 (0.4%) | 1/187 (0.5%) |
| No data | 0/223 (0.0%) | 2/187 (1.1%) |
| Missing | 3/223 (1.3%) | 1/187 (0.5%) |
| VIOLATION: Last 4 weeks - eye/ear disease | | |
| No | 220/223 (98.7%) | 183/187 (97.9%) |
| Yes | 0/223 (0.0%) | 1/187 (0.5%) |
| No data | 0/223 (0.0%) | 2/187 (1.1%) |
| Missing | 3/223 (1.3%) | 1/187 (0.5%) |
| VIOLATION: Last 4 weeks - heart disease/chest pain | | |
| No | 220/223 (98.7%) | 184/187 (98.4%) |
| No data | 0/223 (0.0%) | 2/187 (1.1%) |
| Missing | 3/223 (1.3%) | 1/187 (0.5%) |
| VIOLATION: Last 4 weeks - other infectious disease | | |
| No | 220/223 (98.7%) | 183/187 (97.9%) |
| No data | 0/223 (0.0%) | 2/187 (1.1%) |
| Missing | 3/223 (1.3%) | 2/187 (1.1%) |
| VIOLATION: Last 4 weeks - noncommunicable disease | | |
| No | 219/223 (98.2%) | 182/187 (97.3%) |
| Yes | 1/223 (0.4%) | 1/187 (0.5%) |
| No data | 0/223 (0.0%) | 2/187 (1.1%) |
| Missing | 3/223 (1.3%) | 2/187 (1.1%) |
| VIOLATION: Last 4 weeks - days hospitalised | | |
| Missing | 223/223 (100.0%) | 187/187 (100.0%) |
| VIOLATION: Taking anti-hypertension drugs | | |
| Missing | 223/223 (100.0%) | 187/187 (100.0%) |
| VIOLATION: Province – Guizhou – Altitude | | |
| No | 198/223 (88.8%) | 166/187 (88.8%) |
| Yes | 25/223 (11.2%) | 21/187 (11.2%) |
| VIOLATION: still smokes cigarettes | | |

| | Overall sample | |
|---|---|---|
| | Male N=223 | Female N=187 |
| No | 4/223 (1.8%) | 0/187 (0.0%) |
| Yes | 9/223 (4.0%) | 0/187 (0.0%) |
| No data | 210/223 (94.2%) | 187/187 (100.0%) |
| VIOLATION: Alcohol consumption almost every day | | |
| No | 24/223 (10.8%) | 7/187 (3.7%) |
| Yes | 0/223 (0.0%) | 1/187 (0.5%) |
| Missing | 199/223 (89.2%) | 179/187 (95.7%) |
| VIOLATION (# beer bottles per week): >6 for women and >8 for men | | |
| No | 23/223 (10.3%) | 3/187 (1.6%) |
| Missing | 200/223 (89.7%) | 184/187 (98.4%) |
| VIOLATION (# wine drinks per week): >6 for women and >8 for men | | |
| No | 8/223 (3.6%) | 5/187 (2.7%) |
| Yes | 0/223 (0.0%) | 1/187 (0.5%) |
| Missing | 215/223 (96.4%) | 181/187 (96.8%) |
| VIOLATION (# liquor drinks per week): >6 for women and >8 for men | | |
| No | 9/223 (4.0%) | 2/187 (1.1%) |
| Missing | 214/223 (96.0%) | 185/187 (98.9%) |
| VIOLATION (# beer, wine, liquor): >6 for women and >8 for men | | |
| No | 23/223 (10.3%) | 7/187 (3.7%) |
| Yes | 1/223 (0.4%) | 1/187 (0.5%) |
| Missing | 199/223 (89.2%) | 179/187 (95.7%) |
| VIOLATION: Ferritin<15ug/L | | |
| No | 213/223 (95.5%) | 157/187 (84.0%) |
| Yes | 10/223 (4.5%) | 30/187 (16.0%) |
| VIOLATION: C-Reactive Protein>5mg/L | | |
| No | 208/223 (93.3%) | 179/187 (95.7%) |
| Yes | 15/223 (6.7%) | 8/187 (4.3%) |

CHN = China, CHNS = China Health and Nutrition Survey, BMI = Body Mass Index
Data are presented as unweighted n/total (%) based on participants with non-missing haemoglobin, ferritin, and C-reactive protein.
No data: No answer/refused, Don't know; Missing: Not applicable, Not asked.
[1] Ferritin≥15ug/L and C-reactive protein≤5mg/L.

#### 4.6 Detailed Individual Exclusions Pregnant women

##### Table 4.6.1 NL Generation R: Participant characteristics of violations (Trimester 1) (18-45 years)

| | **Overall sample**<br>**Female**<br>**N = 5,054** |
|---|---|
| Reference sample[1] | |
| No | 4,279 / 5,054 (84.7%) |
| Yes | 775 / 5,054 (15.3%) |
| VIOLATION: Outcome pregnancy not live birth singleton or missing | |
| No | 4,904 / 5,054 (97.0%) |
| Yes | 150 / 5,054 (3.0%) |
| VIOLATION: PIH, PE, HELLP, Eclampsia, superimposed PE/HELLP, preexisting RR (from medical records) or missing | |
| No | 4,562 / 5,054 (90.3%) |
| Yes | 492 / 5,054 (9.7%) |
| VIOLATION: Preeclampsia or missing | |
| No | 4,562 / 5,054 (90.3%) |
| Yes | 492 / 5,054 (9.7%) |
| VIOLATION: Preexisting hypertension (based on questionnaire and medical records) or missing | |
| No | 4,307 / 5,054 (85.2%) |
| Yes | 747 / 5,054 (14.8%) |
| VIOLATION: Pregnancy Induced Hypertension (positive PE_total excluded) or missing | |
| No | 4,562 / 5,054 (90.3%) |
| Yes | 492 / 5,054 (9.7%) |
| VIOLATION: Ferritin (<15ug/L or >150ug/L) or missing | |
| No | 4,098 / 5,054 (81.1%) |
| Yes | 956 / 5,054 (18.9%) |
| VIOLATION: Folic acid plasma mother <18 weeks (≤6.0nmol/l) or missing | |
| No | 4,505 / 5,054 (89.1%) |
| Yes | 549 / 5,054 (10.9%) |
| VIOLATION: Active vitamin B12 serum mother <18 weeks (≤23pmol/L) or missing | |
| No | 3,088 / 5,054 (61.1%) |
| Yes | 1,966 / 5,054 (38.9%) |
| VIOLATION: C-reactive protein plasma mother <18 weeks (>5mg/L) or missing | |
| No | 2,601 / 5,054 (51.5%) |
| Yes | 2,453 / 5,054 (48.5%) |
| VIOLATION: Body Mass Index first trimester <18.5kg/m$^2$ or >30kg/m$^2$ or missing | |
| No | 4,307 / 5,054 (85.2%) |
| Yes | 747 / 5,054 (14.8%) |
| VIOLATION: Smoked till pregnancy was known or continued smoking in pregnancy or missing | |
| No | 3,244 / 5,054 (64.2%) |
| Yes | 1,810 / 5,054 (35.8%) |

NL = The Netherlands.

Data are presented as n/total (%) based on participants with participants with non-missing haemoglobin in first trimester of pregnancy.
PIH = Pregnancy Induced Hypertension, PE = Pulmonary Embolus, HELLP = Hemolysis, Elevated Liver enzymes and Low Platelets, RR = …
[1] Ferritin≥15 and ≤150 ug/L and C-reactive protein≤5mg/L (both first trimester for pregnancy). If a measurement of one or more of the healthy criteria is missing, then the participant is excluded from the reference sample.

Table 4.6.2 NL Generation R: Participant characteristics of violations (Trimester 2) (18-45 years)

| | Overall sample<br>Female<br>N = 2,194 |
|---|---|
| Reference sample[1] | |
| No | 2,083 / 2,194 (94.9%) |
| Yes | 111 / 2,194 (5.1%) |
| VIOLATION: Outcome pregnancy not live birth singleton or missing | |
| No | 2,144 / 2,194 (97.7%) |
| Yes | 50 / 2,194 (2.3%) |
| VIOLATION: PIH, PE, HELLP, Eclampsia, superimposed PE/HELLP, preexisting RR (from medical records) or missing | |
| No | 1,968 / 2,194 (89.7%) |
| Yes | 226 / 2,194 (10.3%) |
| VIOLATION: Preeclampsia or missing | |
| No | 1,968 / 2,194 (89.7%) |
| Yes | 226 / 2,194 (10.3%) |
| VIOLATION: Preexisting hypertension (based on questionnaire and medical records) or missing | |
| No | 1,776 / 2,194 (80.9%) |
| Yes | 418 / 2,194 (19.1%) |
| VIOLATION: Pregnancy Induced Hypertension (positive PE_total excluded) or missing | |
| No | 1,968 / 2,194 (89.7%) |
| Yes | 226 / 2,194 (10.3%) |
| VIOLATION: Ferritin (<15ug/L or >150ug/L) or missing | |
| No | 567 / 2,194 (25.8%) |
| Yes | 1,627 / 2,194 (74.2%) |
| VIOLATION: Folic acid plasma mother <18 weeks (≤6.0nmol/l) or missing | |
| No | 607 / 2,194 (27.7%) |
| Yes | 1,587 / 2,194 (72.3%) |
| VIOLATION: Active vitamin B12 serum mother <18 weeks (≤23pmol/L) or missing | |
| No | 423 / 2,194 (19.3%) |
| Yes | 1,771 / 2,194 (80.7%) |
| VIOLATION: C-reactive protein plasma mother <18 weeks (>5mg/L) or missing | |
| No | 373 / 2,194 (17.0%) |
| Yes | 1,821 / 2,194 (83.0%) |
| VIOLATION: Body Mass Index first trimester <18.5kg/m$^2$ or >30kg/m$^2$ or missing | |
| No | 684 / 2,194 (31.2%) |
| Yes | 1,510 / 2,194 (68.8%) |
| VIOLATION: Smoked till pregnancy was known or continued smoking in pregnancy or missing | |
| No | 1,368 / 2,194 (62.4%) |
| Yes | 826 / 2,194 (37.6%) |

NL = The Netherlands.
Data are presented as n/total (%) based on participants with non-missing haemoglobin in second trimester of pregnancy.
PIH = Pregnancy Induced Hypertension, PE = Pulmonary Embolus, HELLP = Hemolysis, Elevated Liver enzymes and Low Platelets, RR = Pre-existing Hypertension

[1] Ferritin≥15 and ≤150 ug/L and C-reactive protein≤5mg/L (both first trimester for pregnancy). If a measurement of one or more of the healthy criteria is missing, then the participant is excluded from the reference sample.

#### Table 4.6.3 NL Generation R: Participant characteristics of violations (Trimester 3) (18-45 years)

| | **Overall sample**<br>**Female**<br>**N = 239** |
|---|---|
| Reference sample[1] | |
| No | 238 / 239 (99.6%) |
| Yes | 1 / 239 (0.4%) |
| VIOLATION: Outcome pregnancy not live birth singleton or missing | |
| No | 232.0 / 239.0 (97.1%) |
| Yes | 7.0 / 239.0 (2.9%) |
| VIOLATION: PIH, PE, HELLP, Eclampsia, superimposed PE/HELLP, preexisting RR (from medical records) or missing | |
| No | 211.0 / 239.0 (88.3%) |
| Yes | 28.0 / 239.0 (11.7%) |
| VIOLATION: Preeclampsia or missing | |
| No | 211.0 / 239.0 (88.3%) |
| Yes | 28.0 / 239.0 (11.7%) |
| VIOLATION: Preexisting hypertension (based on questionnaire and medical records) or missing | |
| No | 159.0 / 239.0 (66.5%) |
| Yes | 80.0 / 239.0 (33.5%) |
| VIOLATION: Pregnancy Induced Hypertension (positive PE_total excluded) or missing | |
| No | 211.0 / 239.0 (88.3%) |
| Yes | 28.0 / 239.0 (11.7%) |
| VIOLATION: Ferritin (<15ug/L or >150ug/L) or missing | |
| No | 6.0 / 239.0 (2.5%) |
| Yes | 233.0 / 239.0 (97.5%) |
| VIOLATION: Folic acid plasma mother <18 weeks (≤6.0nmol/l) or missing | |
| No | 6.0 / 239.0 (2.5%) |
| Yes | 233.0 / 239.0 (97.5%) |
| VIOLATION: Active vitamin B12 serum mother <18 weeks (≤23pmol/L) or missing | |
| No | 4.0 / 239.0 (1.7%) |
| Yes | 235.0 / 239.0 (98.3%) |
| VIOLATION: C-reactive protein plasma mother <18 weeks (>5mg/L) or missing | |
| No | 4.0 / 239.0 (1.7%) |
| Yes | 235.0 / 239.0 (98.3%) |
| VIOLATION: Body Mass Index first trimester <18.5kg/m$^2$ or >30kg/m$^2$ or missing | |
| No | 7.0 / 239.0 (2.9%) |
| Yes | 232.0 / 239.0 (97.1%) |
| VIOLATION: Smoked till pregnancy was known or continued smoking in pregnancy or missing | |
| No | 130.0 / 239.0 (54.4%) |
| Yes | 109.0 / 239.0 (45.6%) |

NL = The Netherlands.
Data are presented as n/total (%) based on participants with participants with non-missing haemoglobin in third trimester of pregnancy.
PIH = Pregnancy Induced Hypertension, PE = Pulmonary Embolus, HELLP = Hemolysis, Elevated Liver enzymes and Low Platelets, RR = …
[1] Ferritin≥15 and ≤150 ug/L and C-reactive protein≤5mg/L (both first trimester for pregnancy). If a measurement of one or more of the healthy criteria is missing, then the participant is excluded from the reference sample.

### 5. Discrete thresholds

#### Figure 5.1: Adults (18-65 years): 2.5th centile (90% CI)

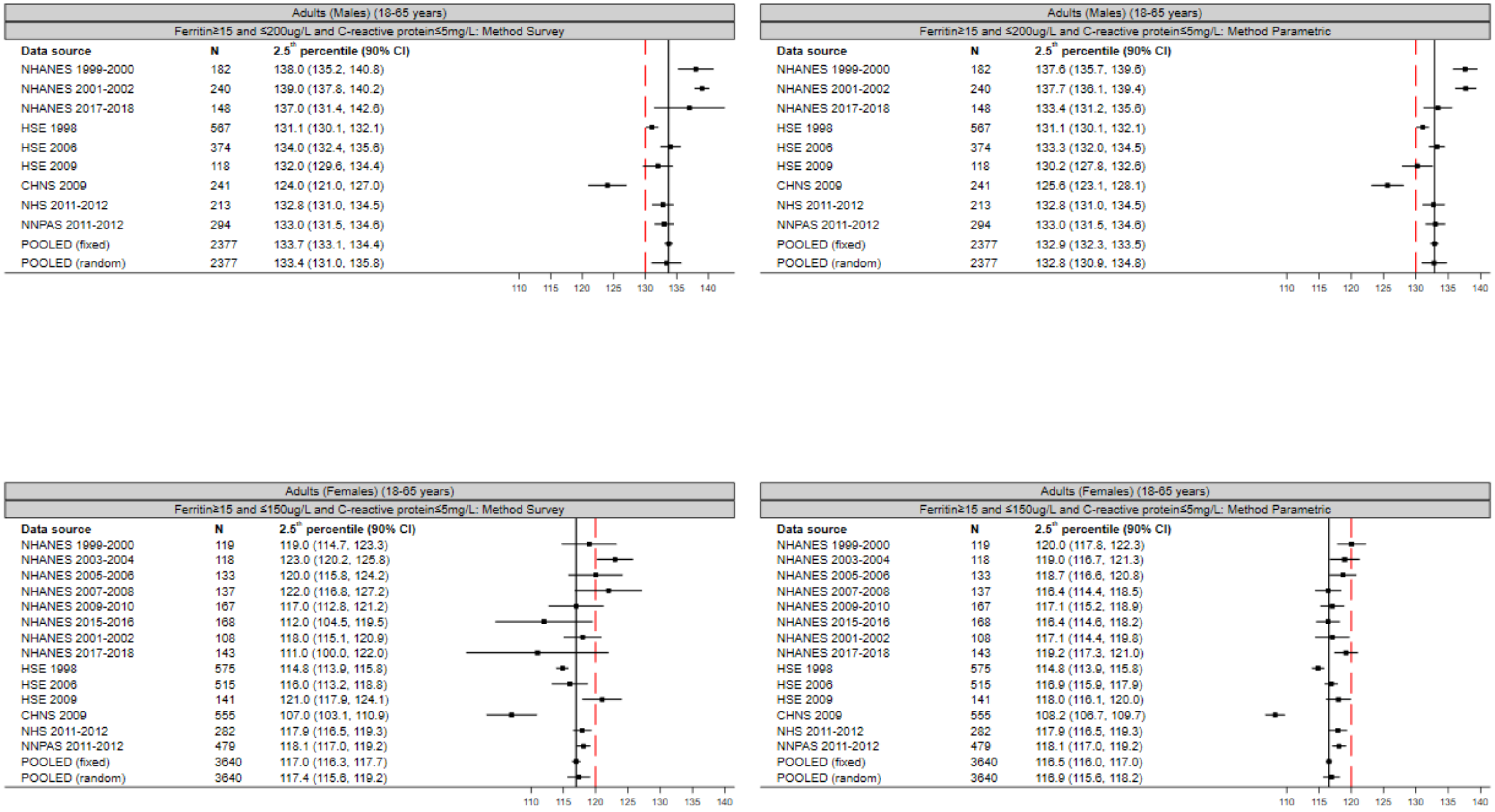

CI = Confidence Interval. Red line = cut-off anaemia according to WHO guideline (2011). Black line = pooled estimate of fixed-effect meta-analysis.

#### Figure 5.2: Children (6-23 months, 24-59 months, 5-11 years): 2.5th centile (90% CI)

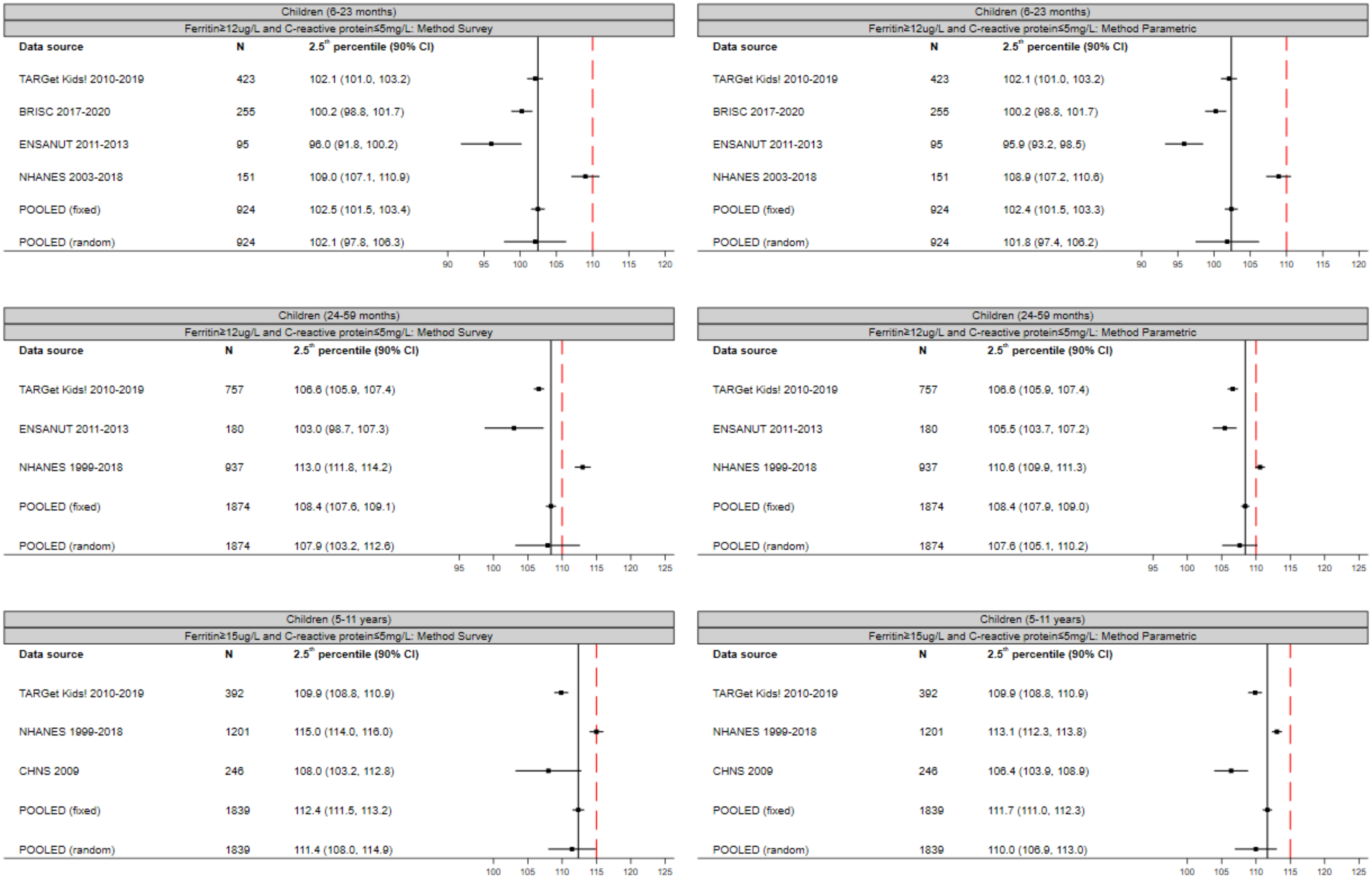

CI = Confidence Interval. Red line = cut-off anaemia according to WHO guideline (2011). Black line = pooled estimate of fixed-effect meta-analysis.

#### Figure 5.3: Children (12-17 years): 2.5th centile (90% CI)

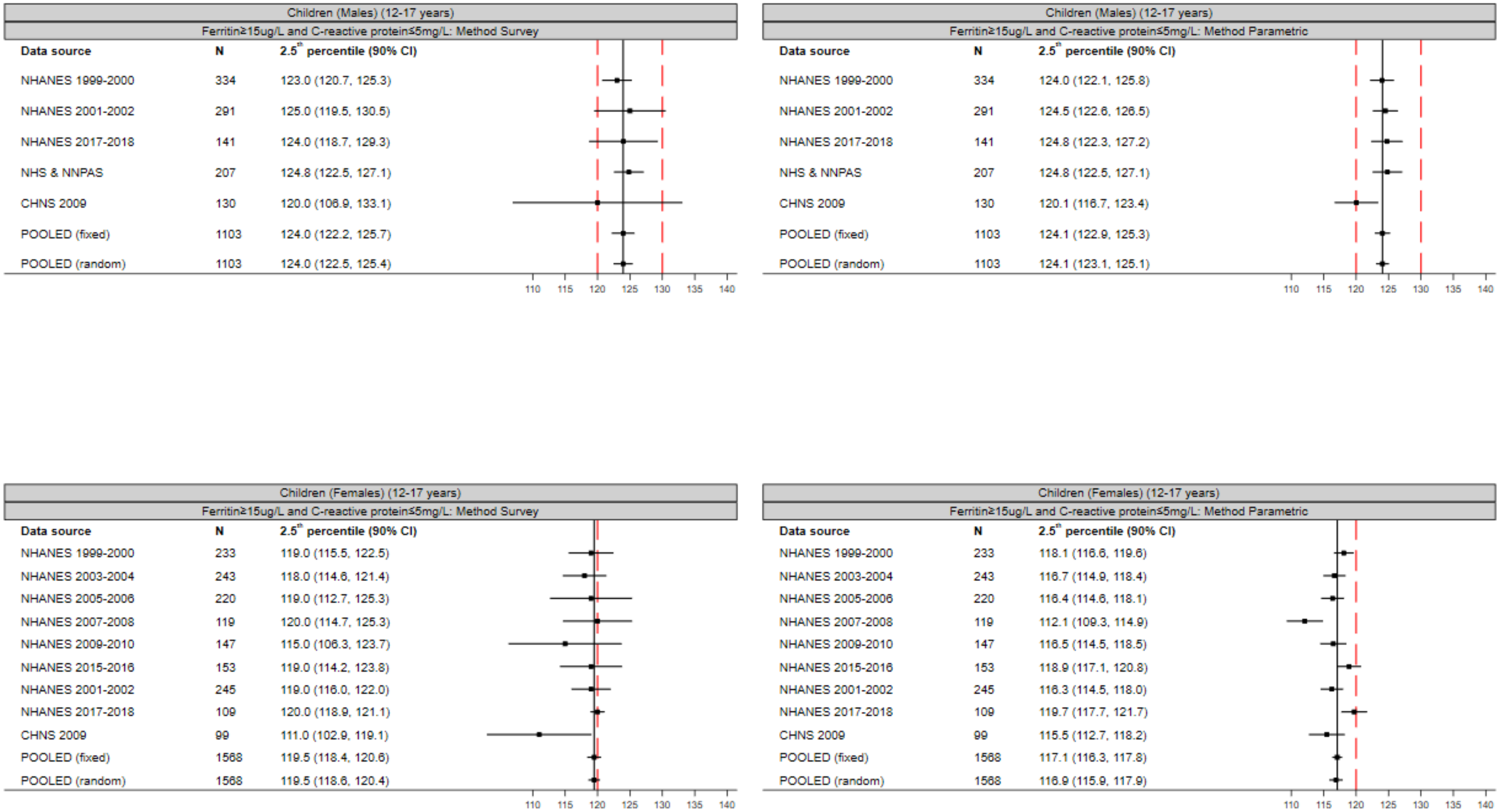

CI = Confidence Interval. Red line = cut-off anaemia according to WHO guideline (2011). Black line = pooled estimate of fixed-effect meta-analysis.

#### Figure 5.4: Sensitivity Analyses Adults (Males) (18-65 years): 5th centile (90% CI)

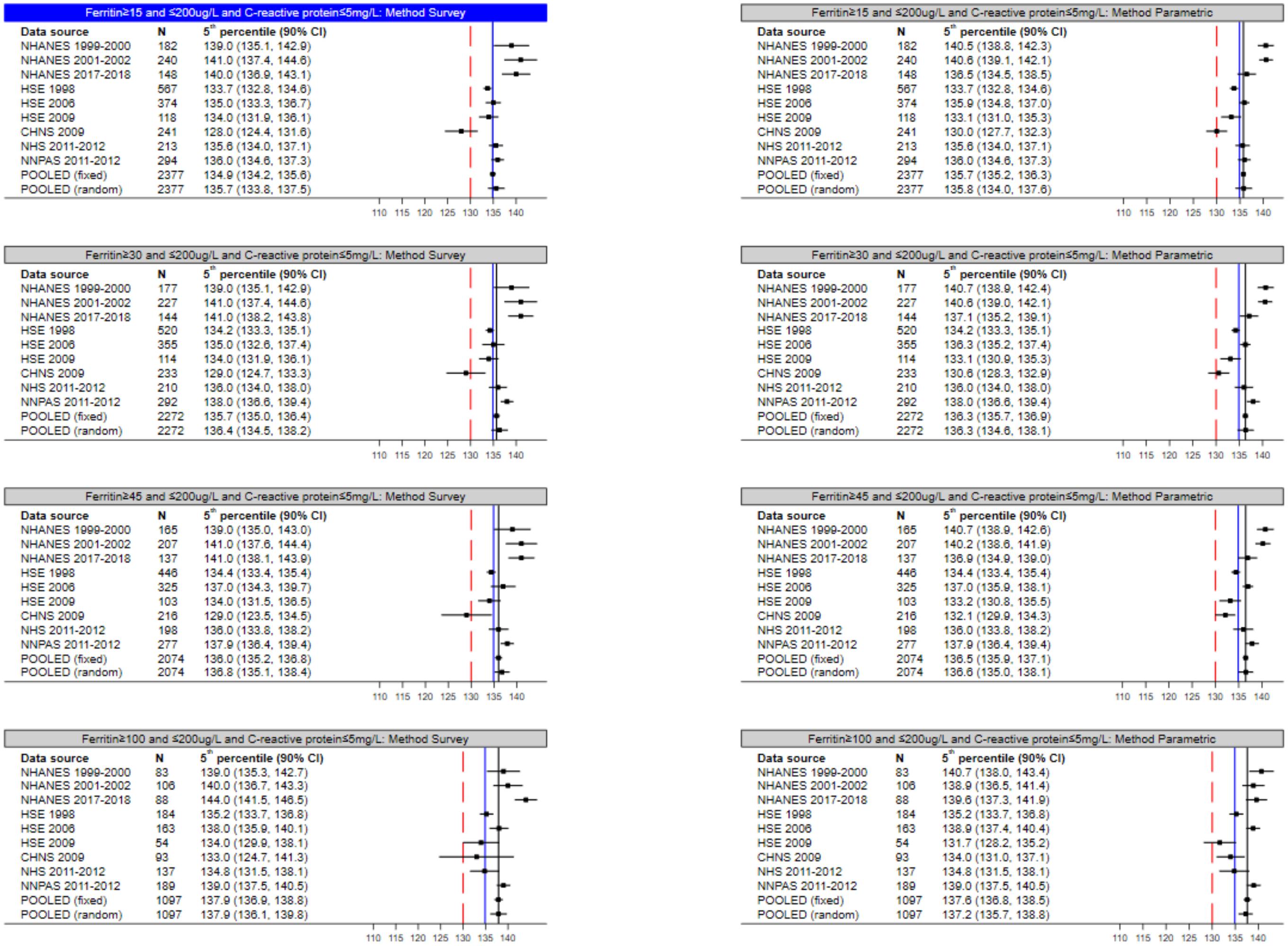

CI = Confidence Interval. Red line = cut-off anaemia according to WHO guideline (2011). Blue line = pooled estimate of fixed-effect meta-analysis of primary definition and analysis method (top left figure). Black line = pooled estimate of fixed-effect meta-analysis. If N<20, no centile is derived. NHS or NNPAS centile not displayed if sample size less than 100, as per Australian Bureau of Statistics requirements for output clearance.

#### Figure 5.5: Sensitivity Analyses Adults (Females) (18-65 years): 5th centile (90% CI)

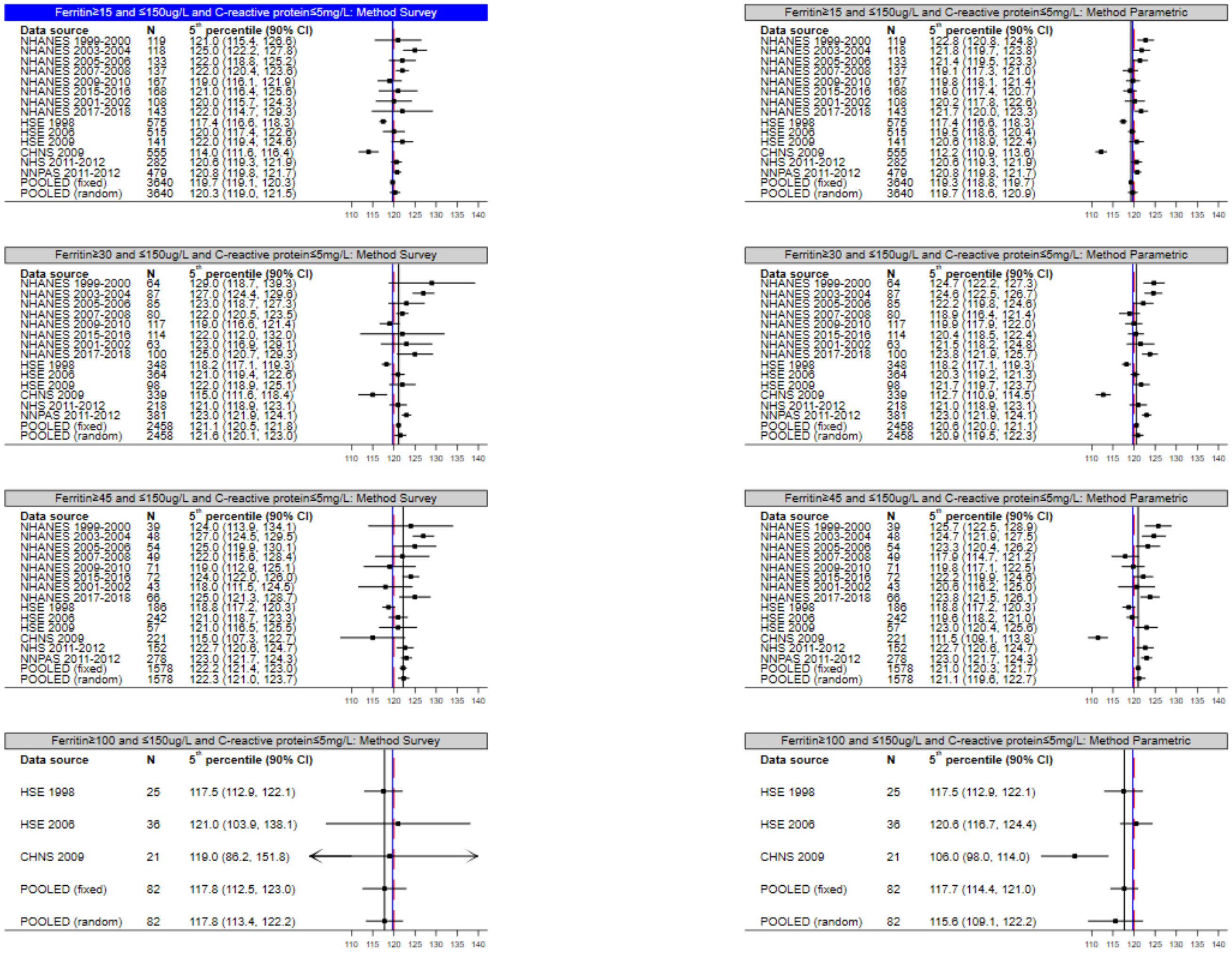

CI = Confidence Interval. Red line = cut-off anaemia according to WHO guideline (2011). Blue line = pooled estimate of fixed-effect meta-analysis of primary definition and analysis method (top left figure). Black line = pooled estimate of fixed-effect meta-analysis. If N<20, no centile is derived. NHS or NNPAS centile not displayed if sample size less than 100, as per Australian Bureau of Statistics requirements for output clearance.

#### Figure 5.6: Sensitivity Analyses Children (6-23 months): 5th centile (90% CI) (Part 1)

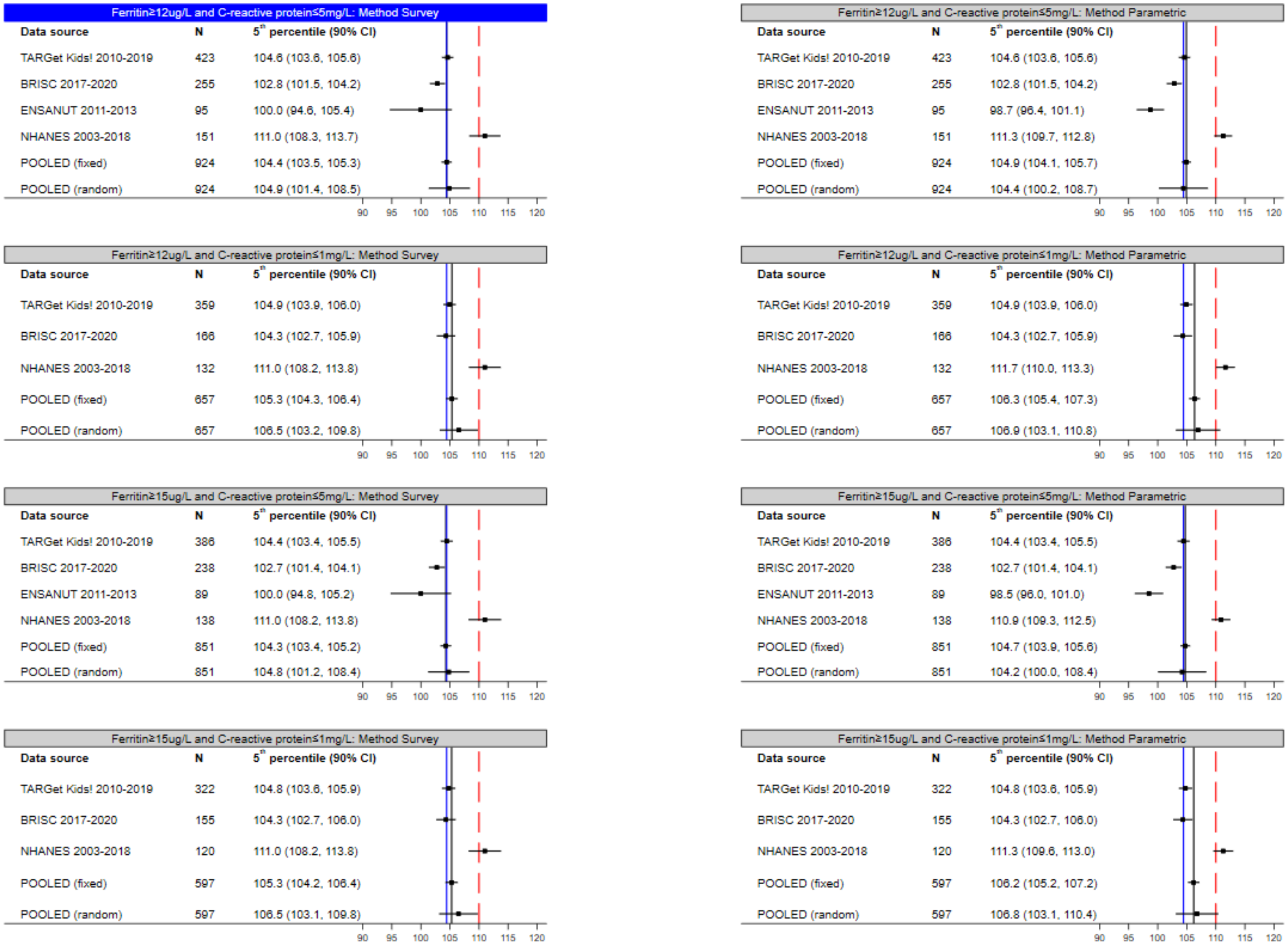

CI = Confidence Interval. Red line = cut-off anaemia according to WHO guideline (2011). Blue line = pooled estimate of fixed-effect meta-analysis of primary definition and analysis method (top left figure). Black line = pooled estimate of fixed-effect meta-analysis. If N<20, no centile is derived.

#### Figure 5.7: Sensitivity Analyses Children (6-23 months): 5th centile (90% CI) (Part 2)

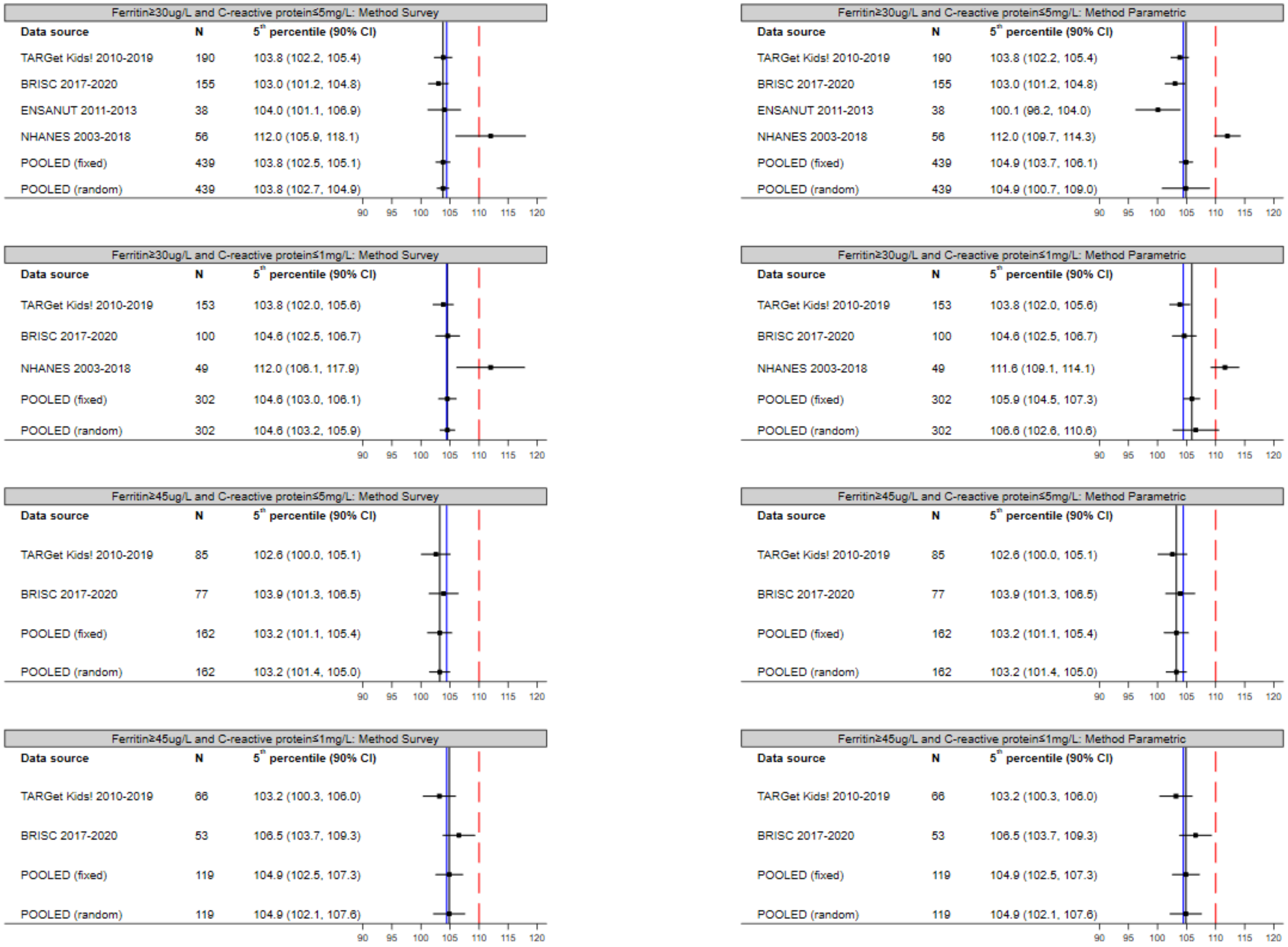

CI = Confidence Interval. Red line = cut-off anaemia according to WHO guideline (2011). Blue line = pooled estimate of fixed-effect meta-analysis of primary definition and analysis method. Black line = pooled estimate of fixed-effect meta-analysis. If N<20, no centile is derived.

#### Figure 5.8: Sensitivity Analyses Children (6-23 months): 5th centile (90% CI) (Part 3)

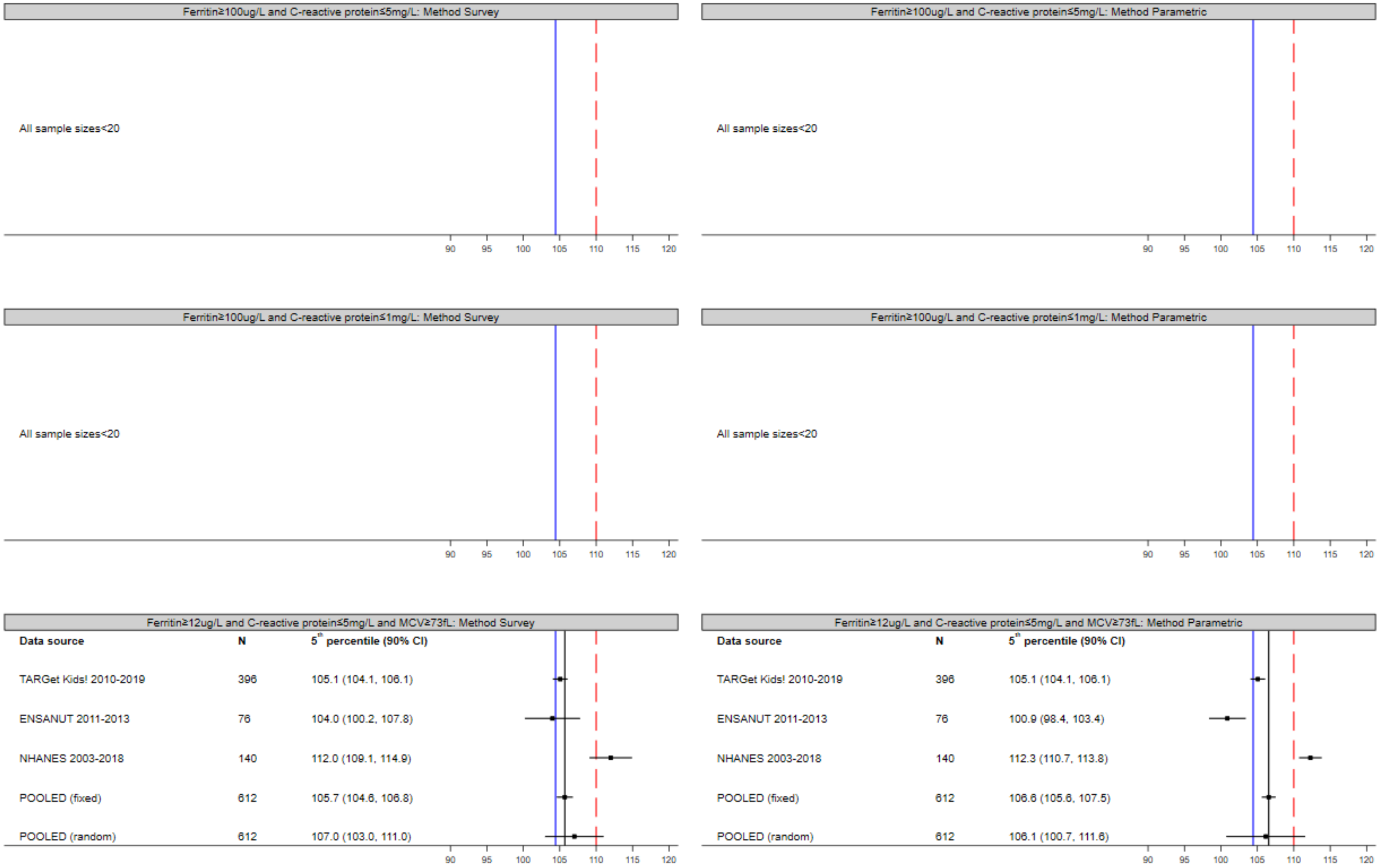

CI = Confidence Interval. Red line = cut-off anaemia according to WHO guideline (2011). Blue line = pooled estimate of fixed-effect meta-analysis of primary definition and analysis method. Black line = pooled estimate of fixed-effect meta-analysis. If N<20, no centile is derived. No MCV collected as part of BRISC.

#### Figure 5.9: Sensitivity Analyses Children (24-59 months): 5th centile (90% CI) (Part 1)

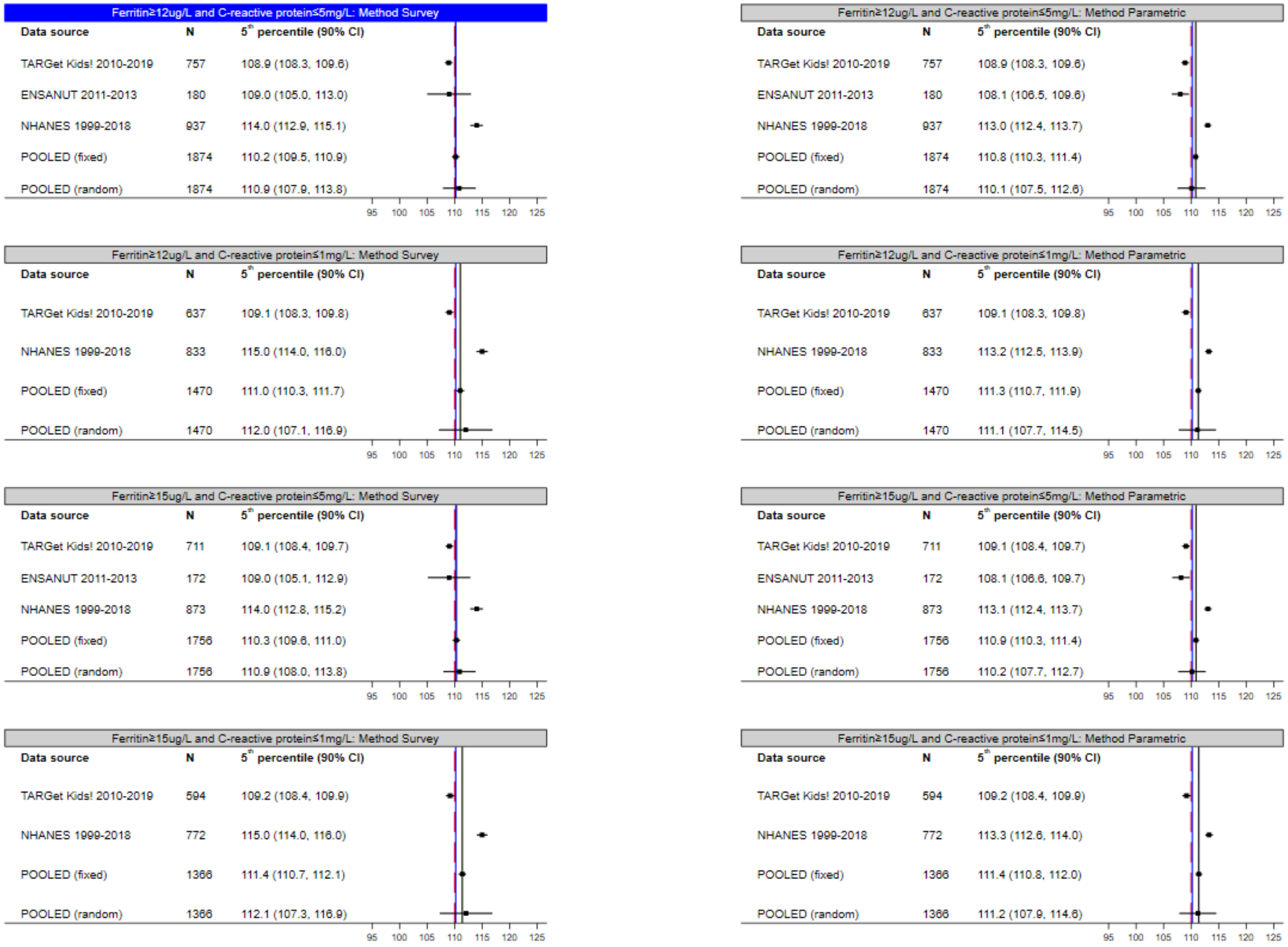

CI = Confidence Interval. Red line = cut-off anaemia according to WHO guideline (2011). Blue line = pooled estimate of fixed-effect meta-analysis of primary definition and analysis method (top left figure). Black line = pooled estimate of fixed-effect meta-analysis. If N<20, no centile is derived.

#### Figure 5.10: Sensitivity Analyses Children (24-59 months): 5th centile (90% CI) (Part 2)

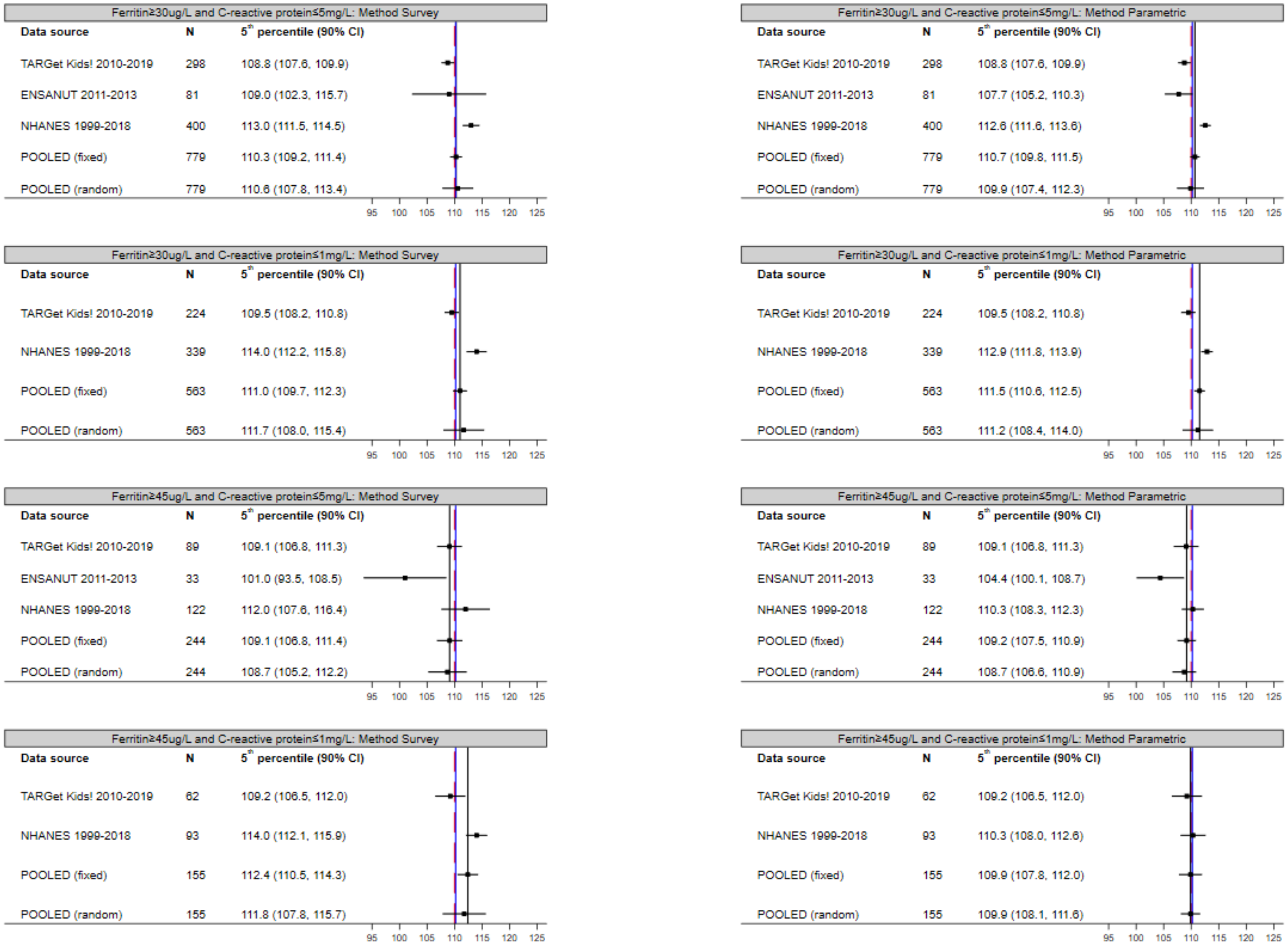

CI = Confidence Interval. Red line = cut-off anaemia according to WHO guideline (2011). Blue line = pooled estimate of fixed-effect meta-analysis of primary definition and analysis method. Black line = pooled estimate of fixed-effect meta-analysis. If N<20, no centile is derived.

Figure 5.11: Sensitivity Analyses Children (24-59 months): 5th centile (90% CI) (Part 3)

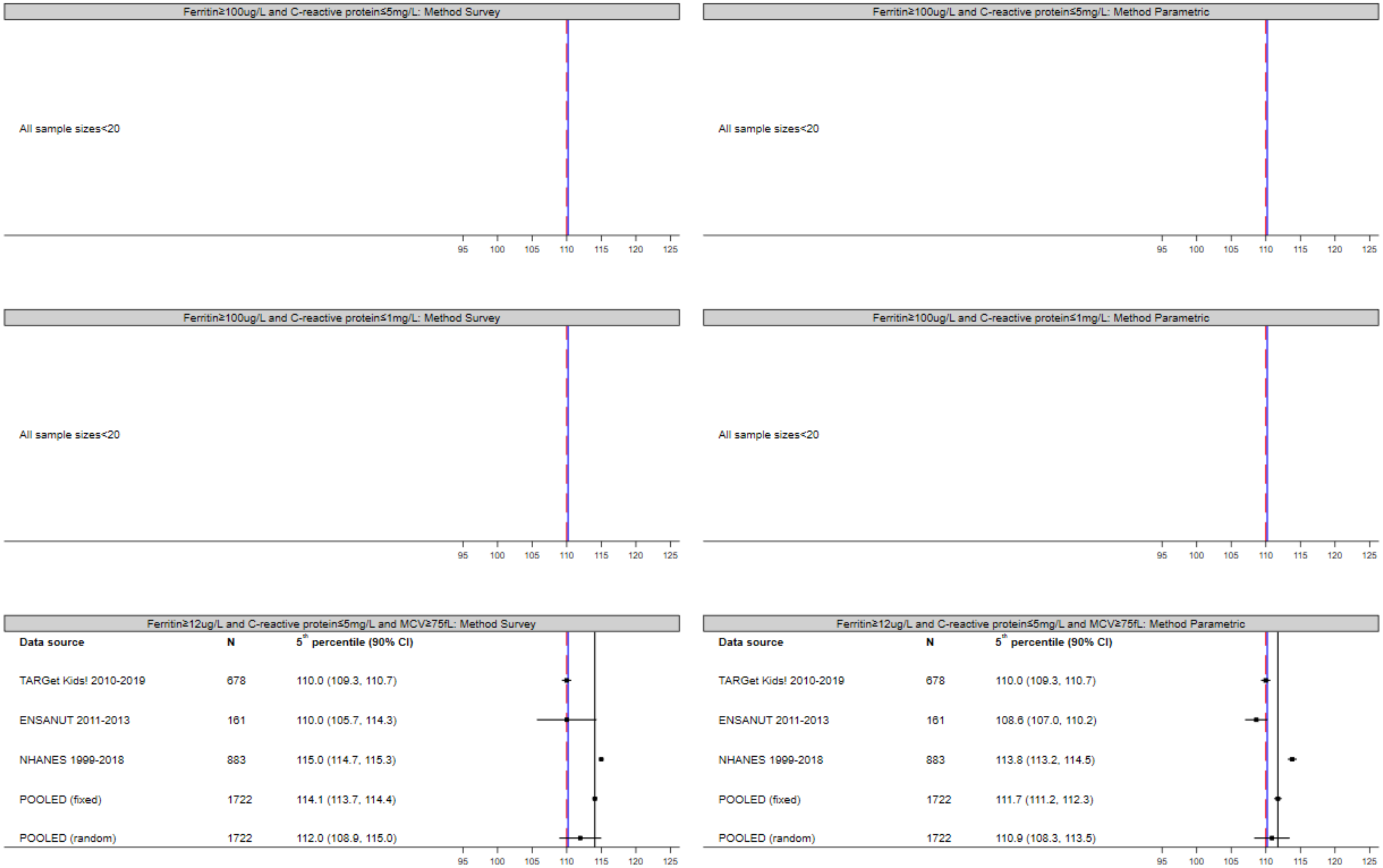

CI = Confidence Interval. Red line = cut-off anaemia according to WHO guideline (2011). Blue line = pooled estimate of fixed-effect meta-analysis of primary definition and analysis method. Black line = pooled estimate of fixed-effect meta-analysis. If N<20, no centile is derived.

#### Figure 5.12: Sensitivity Analyses Children (5-11 years): 5th centile (90% CI) (Part 1)

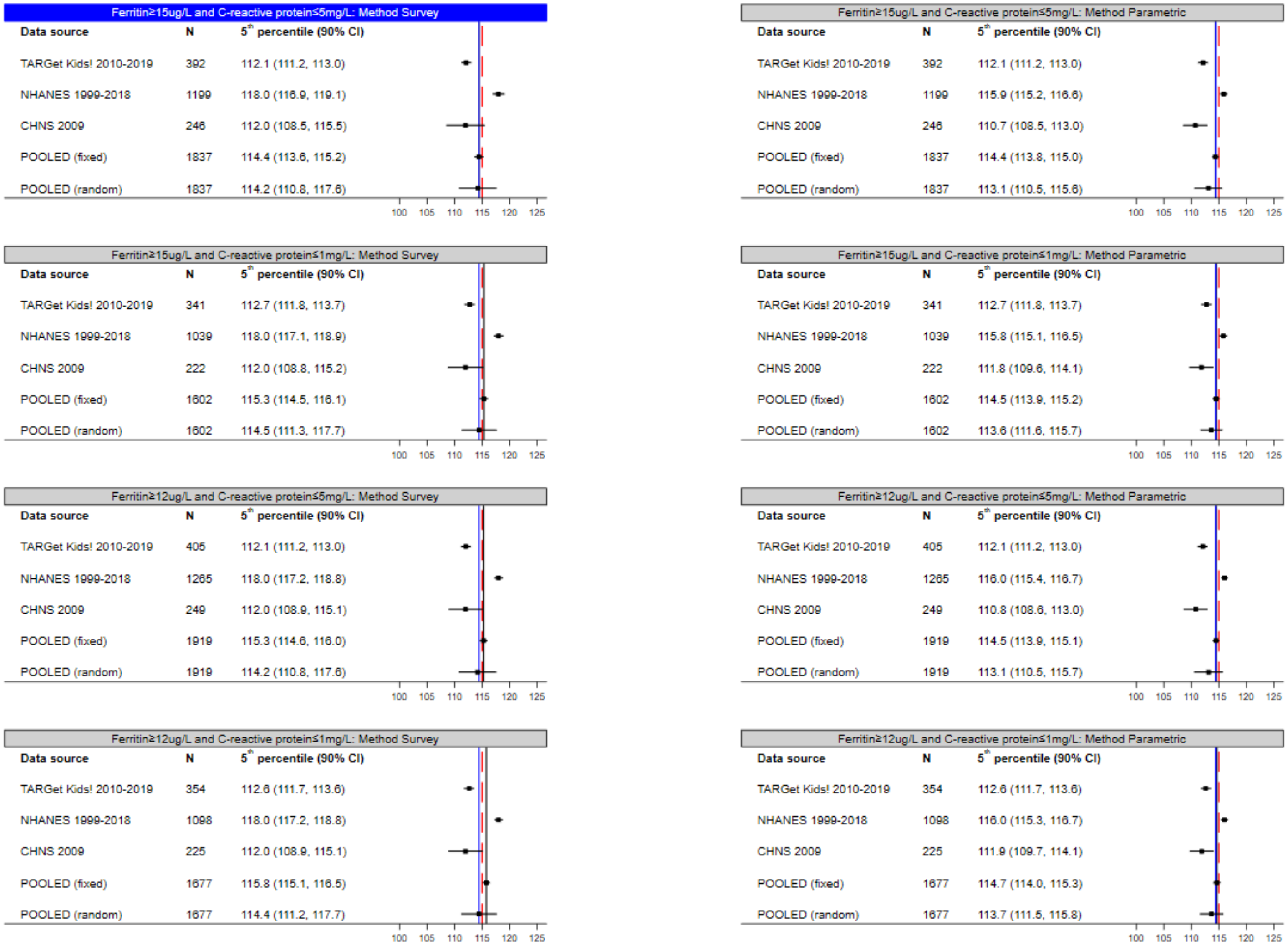

CI = Confidence Interval. Red line = cut-off anaemia according to WHO guideline (2011). Blue line = pooled estimate of fixed-effect meta-analysis of primary definition and analysis method (top left figure). Black line = pooled estimate of fixed-effect meta-analysis. If N<20, no centile is derived.

#### Figure 5.13: Sensitivity Analyses Children (5-11 years): 5th centile (90% CI) (Part 2)

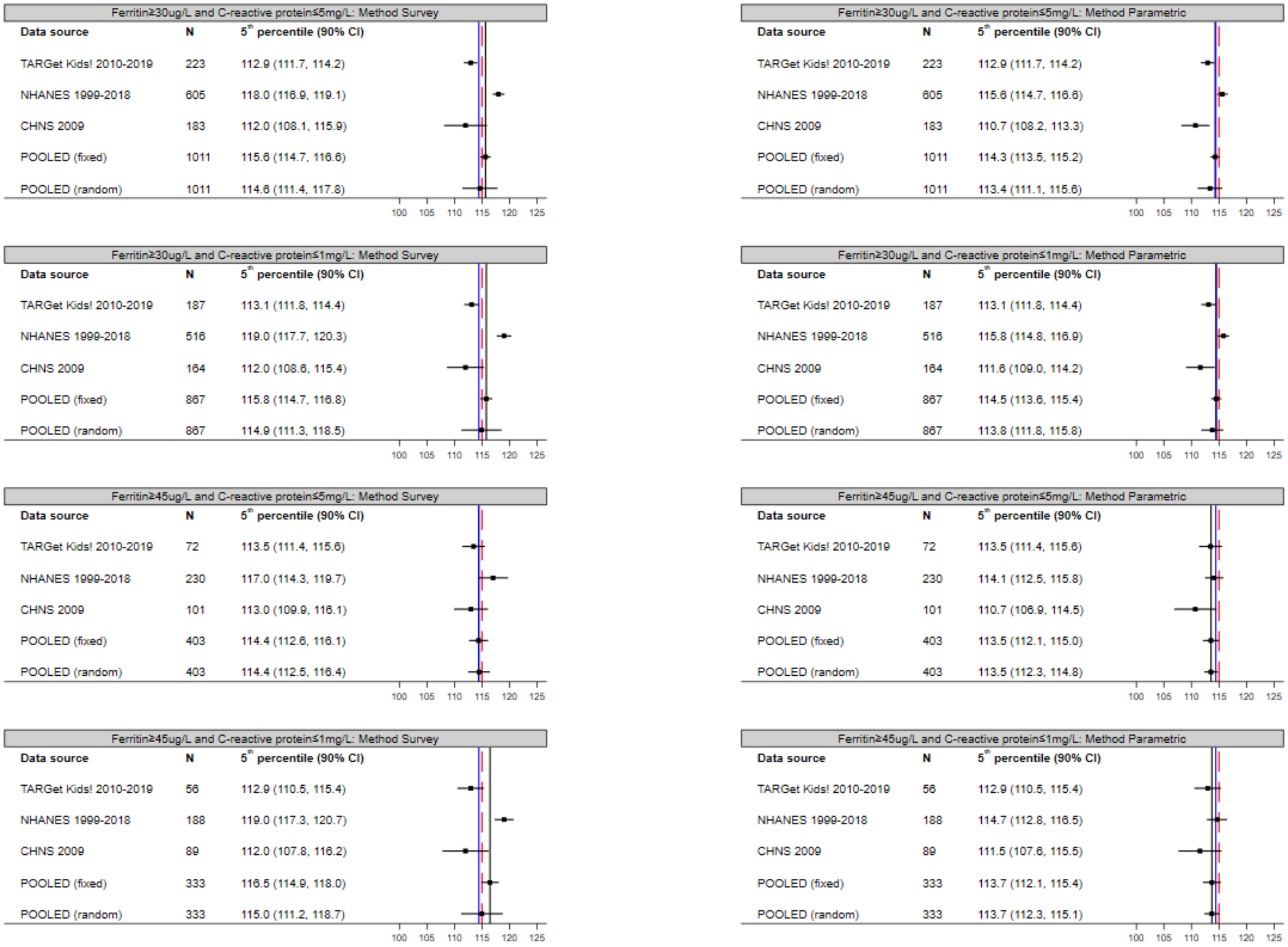

CI = Confidence Interval. Red line = cut-off anaemia according to WHO guideline (2011). Blue line = pooled estimate of fixed-effect meta-analysis of primary definition and analysis method. Black line = pooled estimate of fixed-effect meta-analysis. If N<20, no centile is derived.

#### Figure 5.14: Sensitivity Analyses Children (5-11 years): 5th centile (90% CI) (Part 3)

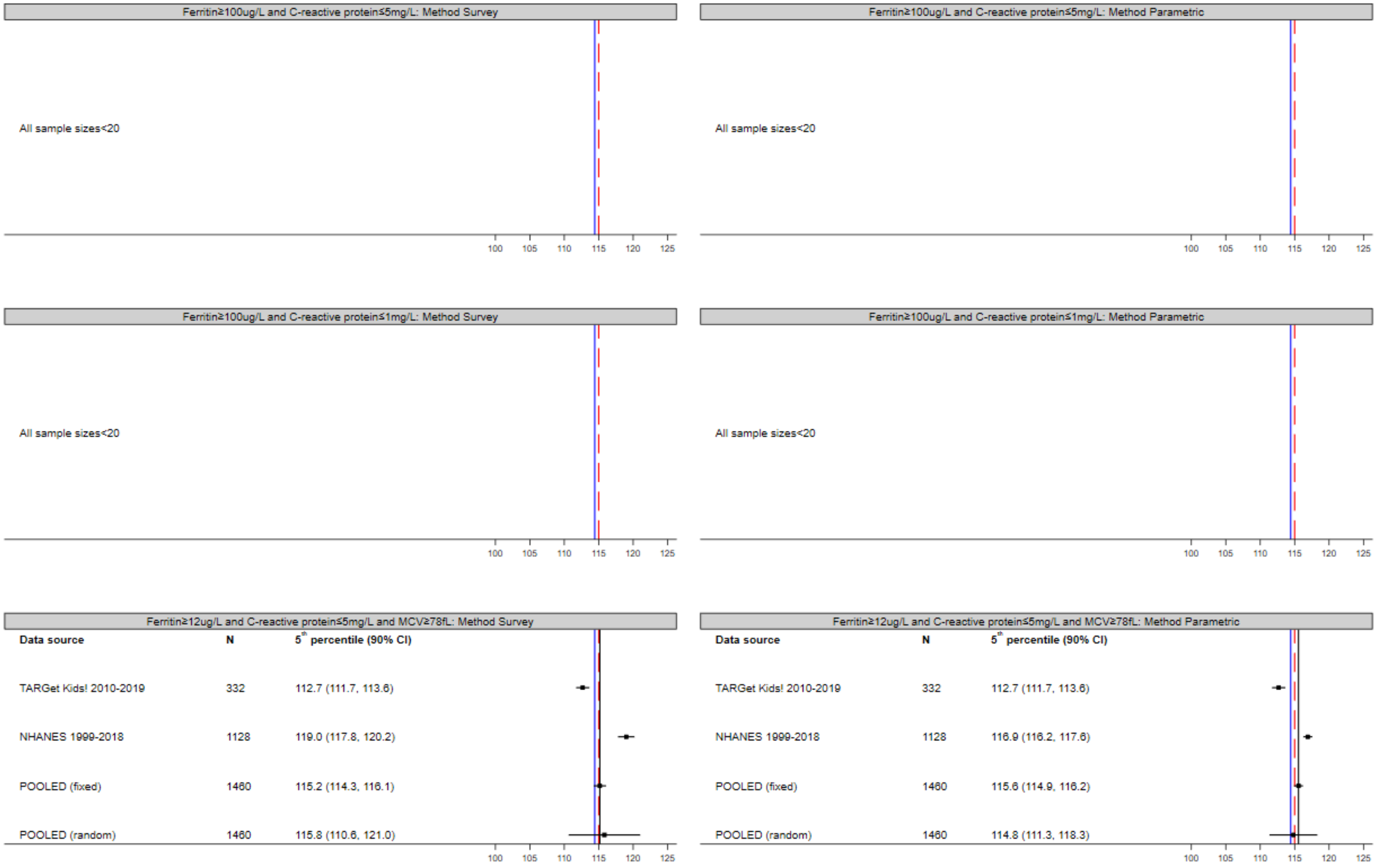

CI = Confidence Interval. Red line = cut-off anaemia according to WHO guideline (2011). Blue line = pooled estimate of fixed-effect meta-analysis of primary definition and analysis method. Black line = pooled estimate of fixed-effect meta-analysis. If N<20, no centile is derived. No MCV collected as part of CHNS.

#### Figure 5.15: Sensitivity Analyses Children (Males) (12-17 years): 5th centile (90% CI) (Part 1)

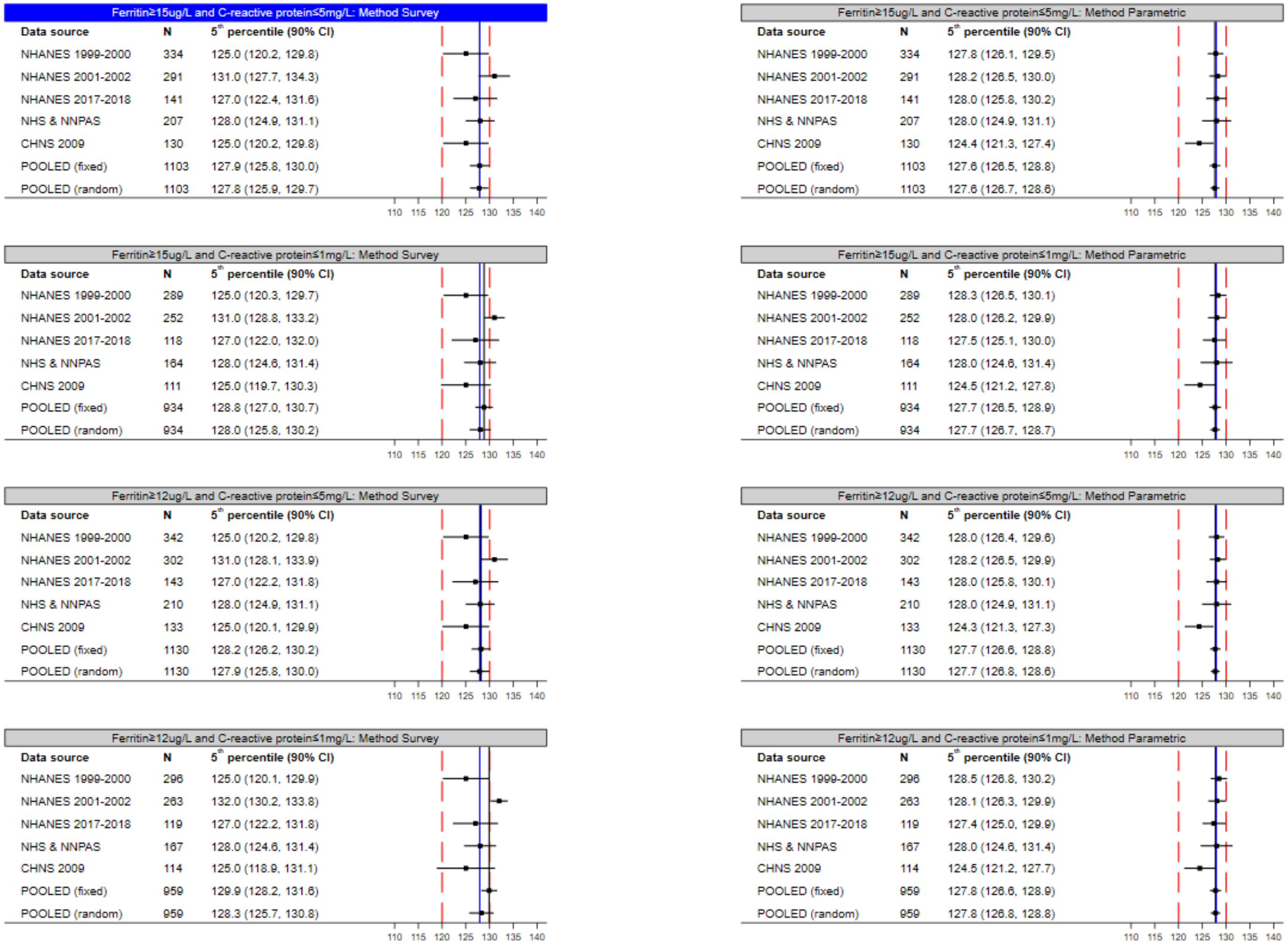

CI = Confidence Interval. Red line = cut-off anaemia according to WHO guideline (2011). Blue line = pooled estimate of fixed-effect meta-analysis of primary definition and analysis method (top left figure). Black line = pooled estimate of fixed-effect meta-analysis. If N<20, no centile is derived. NHS & NNPAS centile not displayed if sample size  less than 100, as per Australian Bureau of Statistics requirements for output clearance.

#### Figure 5.16: Sensitivity Analyses Children (Males) (12-17 years): 5th centile (90% CI) (Part 2)

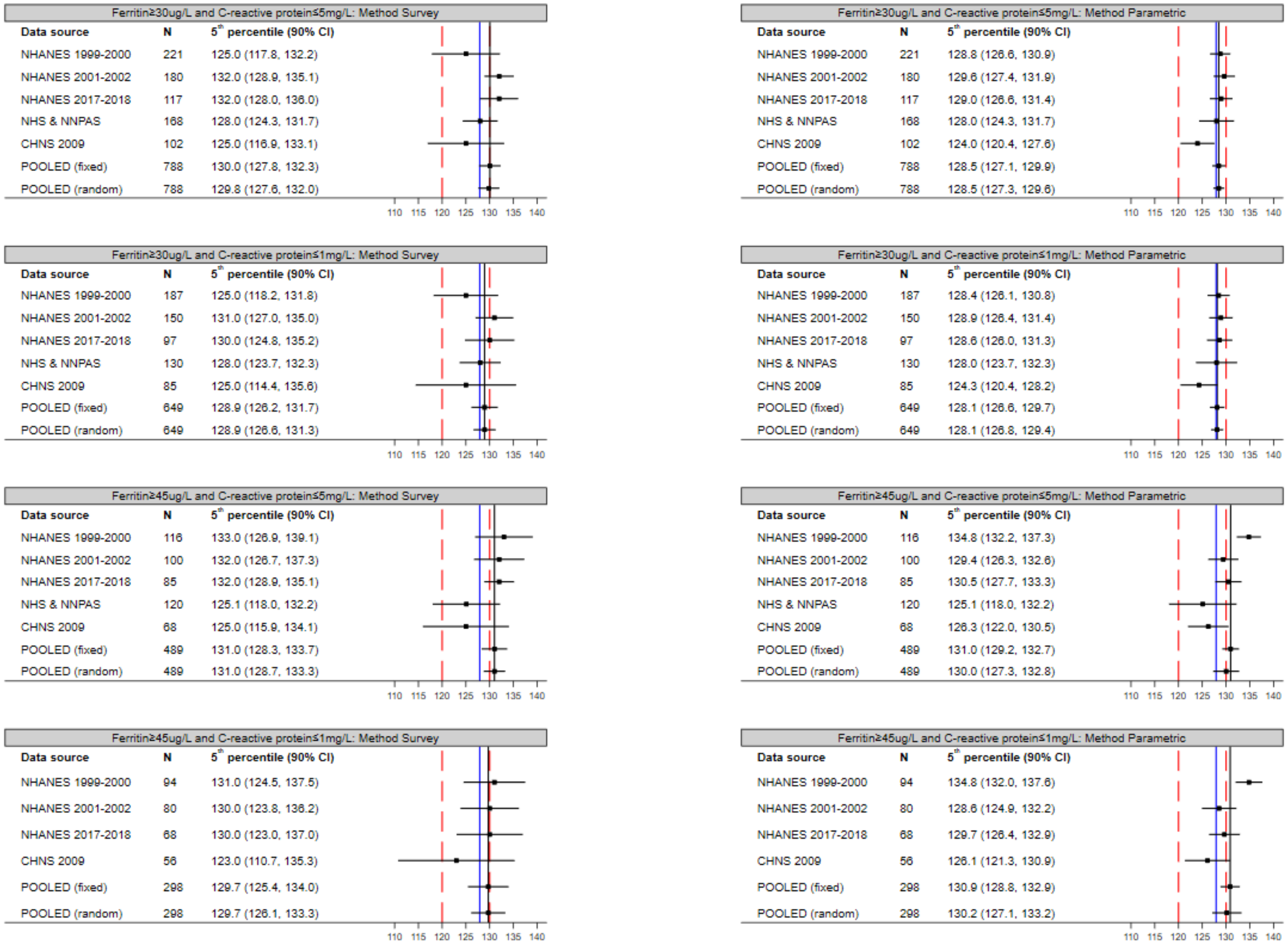

CI = Confidence Interval. Red line = cut-off anaemia according to WHO guideline (2011). Blue line = pooled estimate of fixed-effect meta-analysis of primary definition and analysis method. Black line = pooled estimate of fixed-effect meta-analysis. If N<20, no centile is derived. NHS & NNPAS centile not displayed if sample size  less than 100, as per Australian Bureau of Statistics requirements for output clearance.

Figure 5.17: Sensitivity Analyses Children (Males) (12-17 years): 5th centile (90% CI) (Part 3)

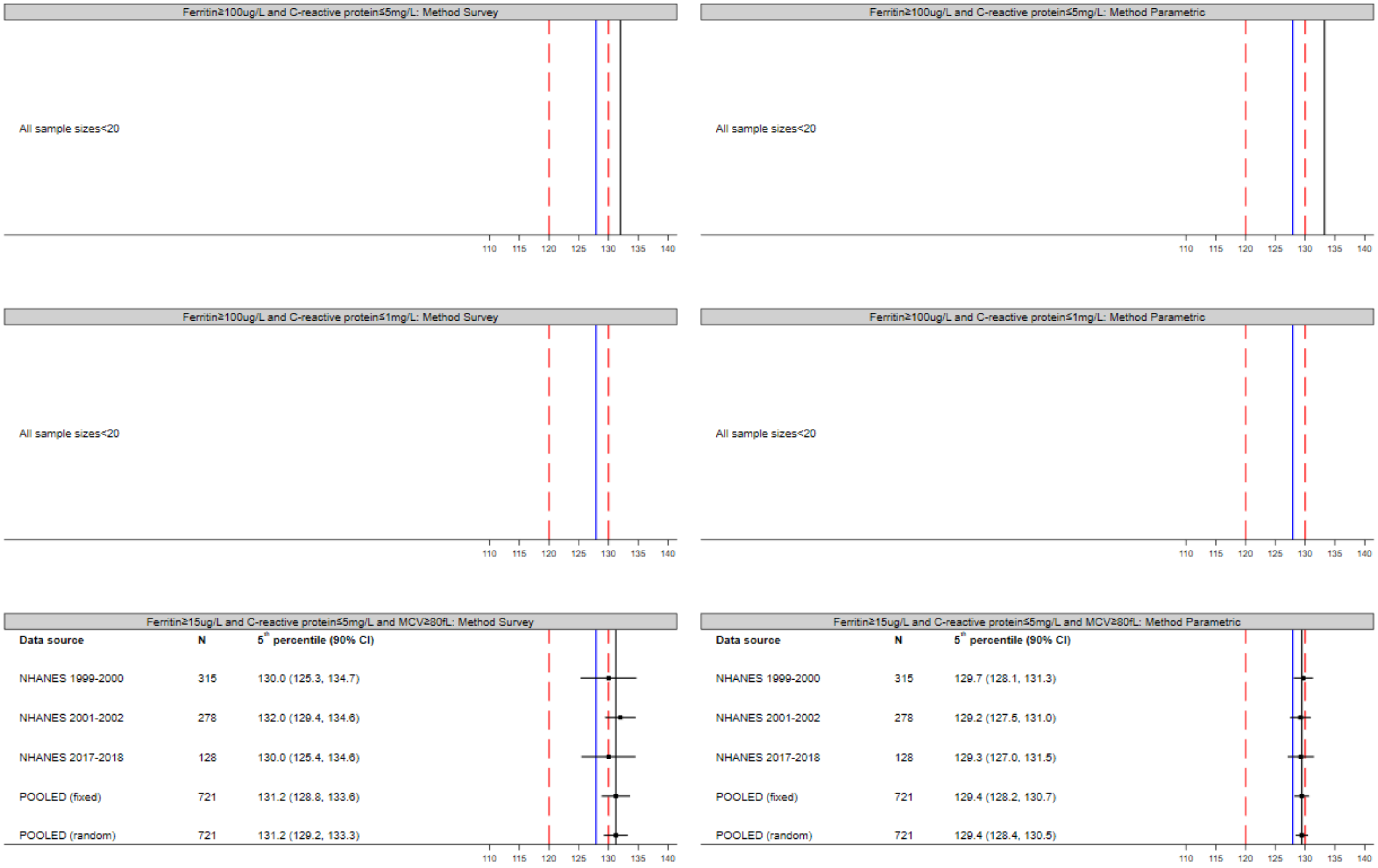

CI = Confidence Interval. Red line = cut-off anaemia according to WHO guideline (2011). Blue line = pooled estimate of fixed-effect meta-analysis of primary definition and analysis method. Black line = pooled estimate of fixed-effect meta-analysis. If N<20, no centile is derived. NHS & NNPAS centile not displayed if sample size less than 100, as per Australian Bureau of Statistics requirements for output clearance. No MCV collected as part of NHS, NNPAS, and CHNS.

#### Figure 5.18: Sensitivity Analyses Children (Females) (12-17 years): 5th centile (90% CI) (Part 1)

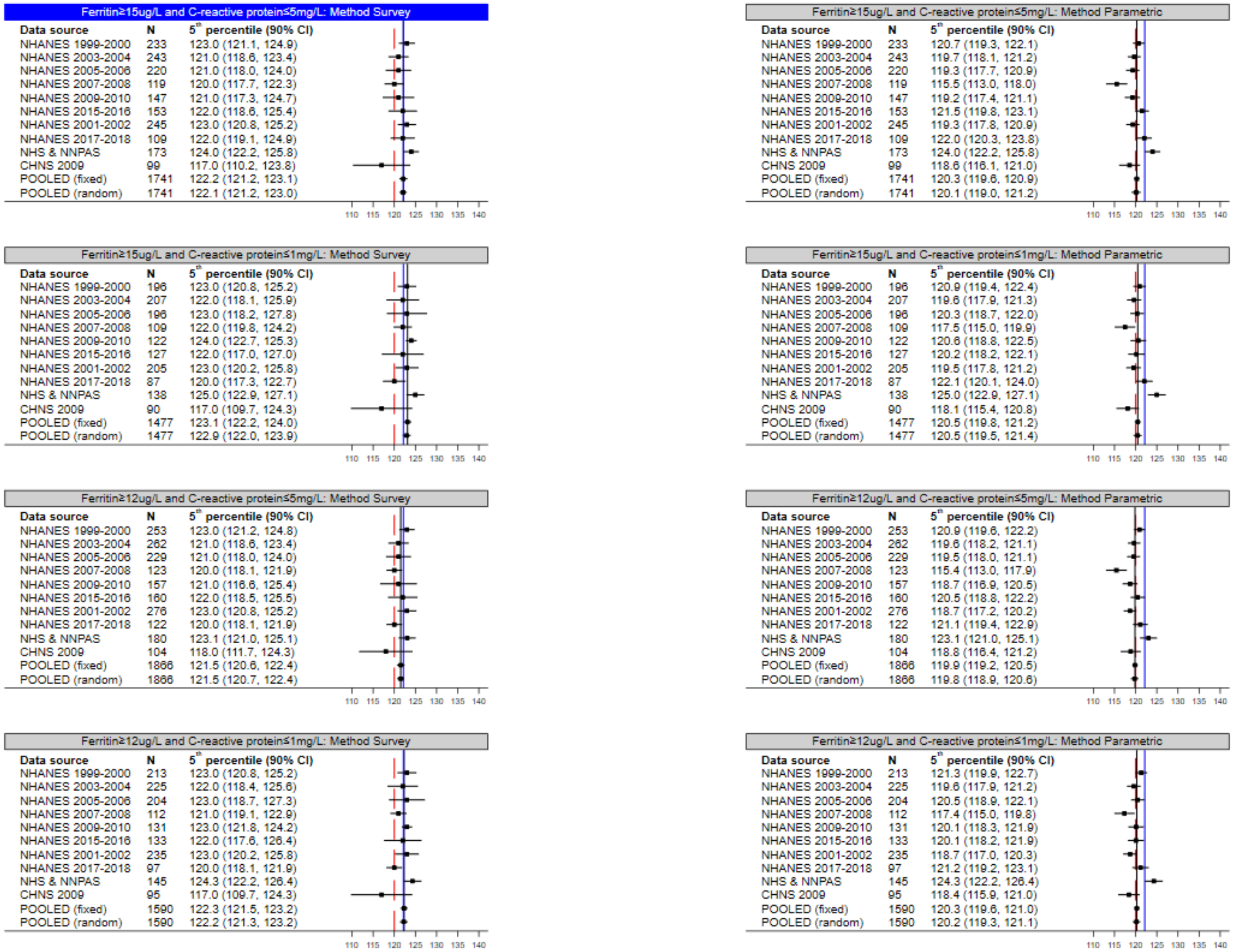

CI = Confidence Interval. Red line = cut-off anaemia according to WHO guideline (2011). Blue line = pooled estimate of fixed-effect meta-analysis of primary definition and analysis method (top left figure). Black line = pooled estimate of fixed-effect meta-analysis. If N<20, no centile is derived. NHS & NNPAS centile not displayed if sample size less than 100, as per Australian Bureau of Statistics requirements for output clearance.

#### Figure 5.19: Sensitivity Analyses Children (Females) (12-17 years): 5th centile (90% CI) (Part 2)

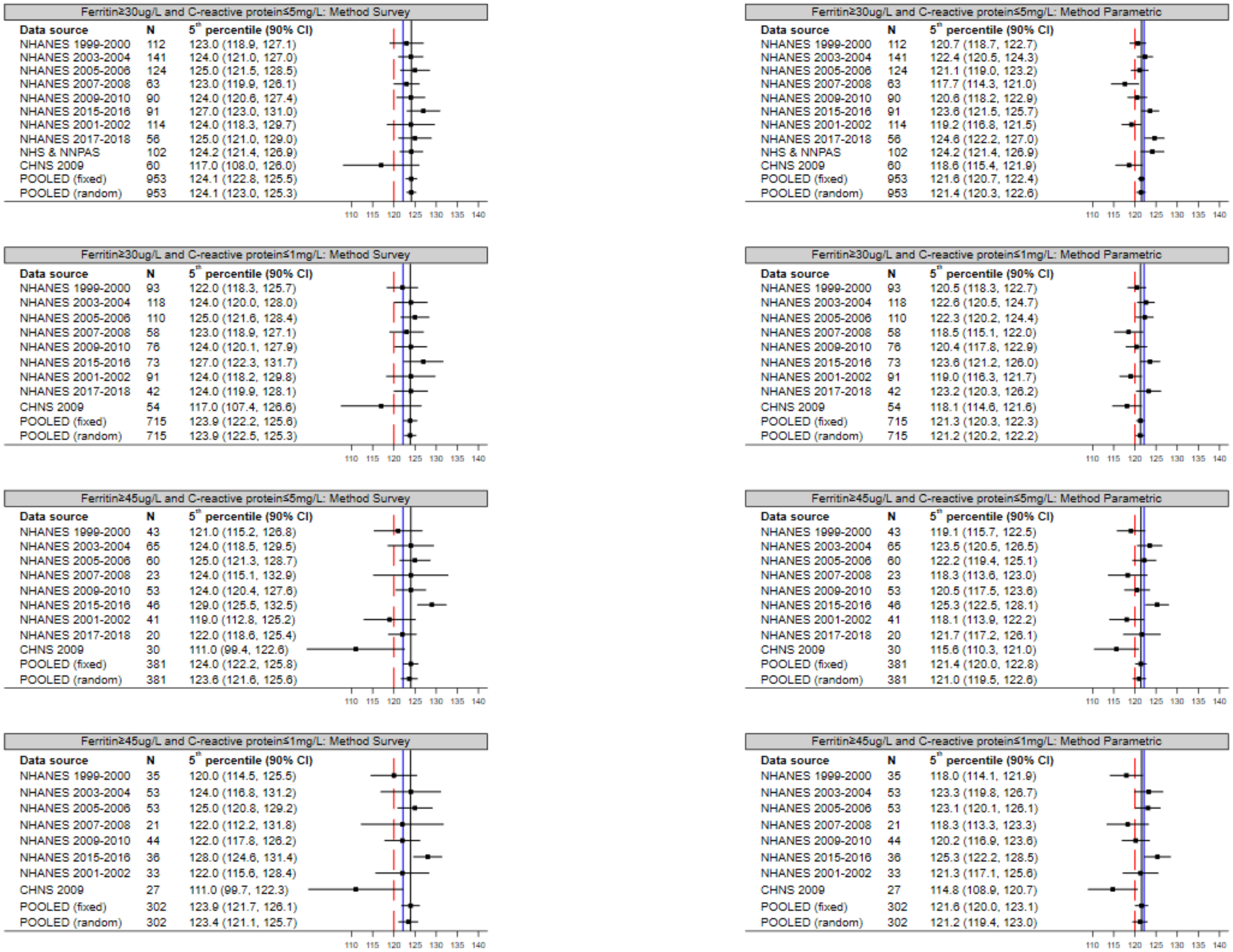

CI = Confidence Interval. Red line = cut-off anaemia according to WHO guideline (2011). Blue line = pooled estimate of fixed-effect meta-analysis of primary definition and analysis method. Black line = pooled estimate of fixed-effect meta-analysis. If N<20, no centile is derived. NHS & NNPAS centile not displayed if sample size less than 100, as per Australian Bureau of Statistics requirements for output clearance.

#### Figure 5.20: Sensitivity Analyses Children (Females) (12-17 years): 5th centile (90% CI) (Part 3)

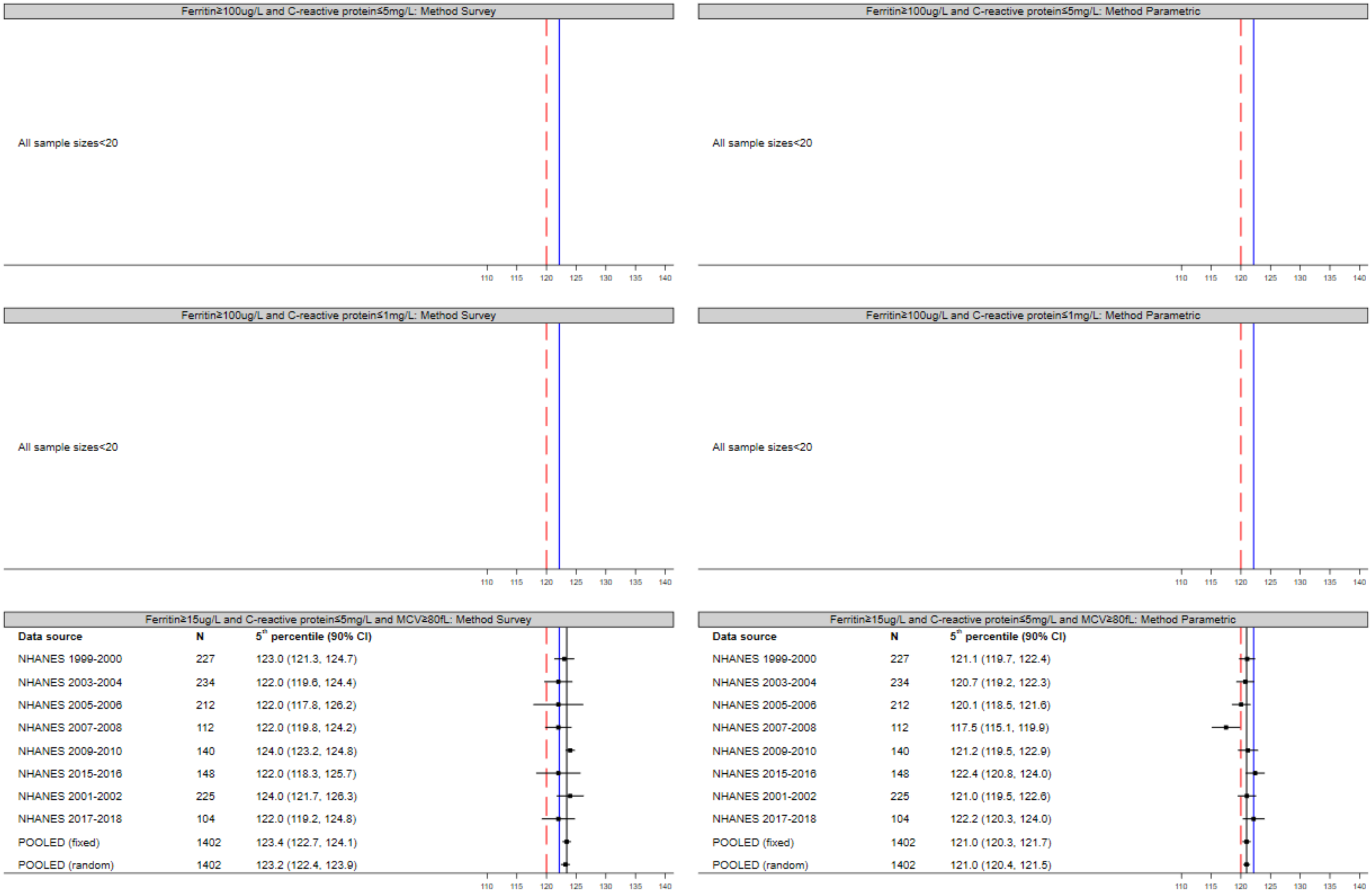

CI = Confidence Interval. Red line = cut-off anaemia according to WHO guideline (2011). Blue line = pooled estimate of fixed-effect meta-analysis of primary definition and analysis method. Black line = pooled estimate of fixed-effect meta-analysis. If N<20, no centile is derived. NHS & NNPAS centile not displayed if sample size less than 100, as per Australian Bureau of Statistics requirements for output clearance. No MCV collected as part of NHS, NNPAS, and CHNS.

Figure 5.21: Sensitivity Analyses Pregnant Women (18-45 years): 5th centile (90% CI)

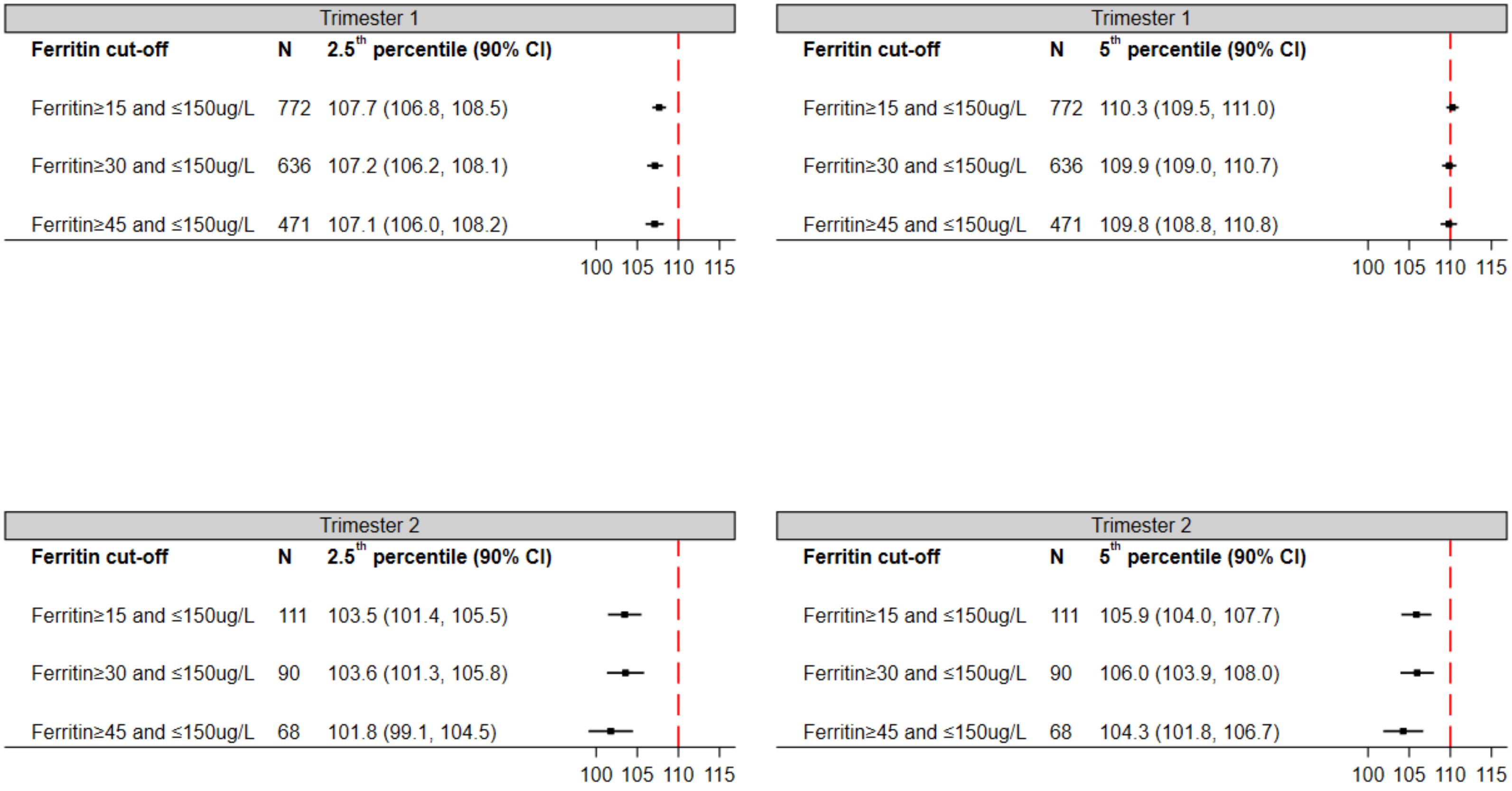

CI = Confidence Interval. Red line = cut-off anaemia according to WHO guideline (2011). All include only women with C-reactive protein≤5mg/L. Ferritin and C-reactive protein as collected in the first trimester of pregnancy.

### 6. Ancestry/genetics

Table 6.1: Genetic association results (summary statistics) of haemoglobin concentrations used

| Variant | Sample size | Number of variants | Ancestry | Reference |
|---|---|---|---|---|
| SNP | 746,667 | ~ 45 million autosomal variants | 76% European<br>20% East Asian<br>2% African<br>1% Hispanic/Latino<br>1% South Asian | Chen et al., 2020[19] |
| SNP | 35,418 | 24,092,221 autosomal variants | South Asian | Genes & Health (G&H) Study[20] |
| SV | 50,675 | 96,049 structural variants | 59% European<br>24% African<br>16% Hispanic/Latino<br>1% Asian | Wheeler et al., 2022[21] |

SNP = Single Nucleotide Polymorphisms, SV = Structural Variants.

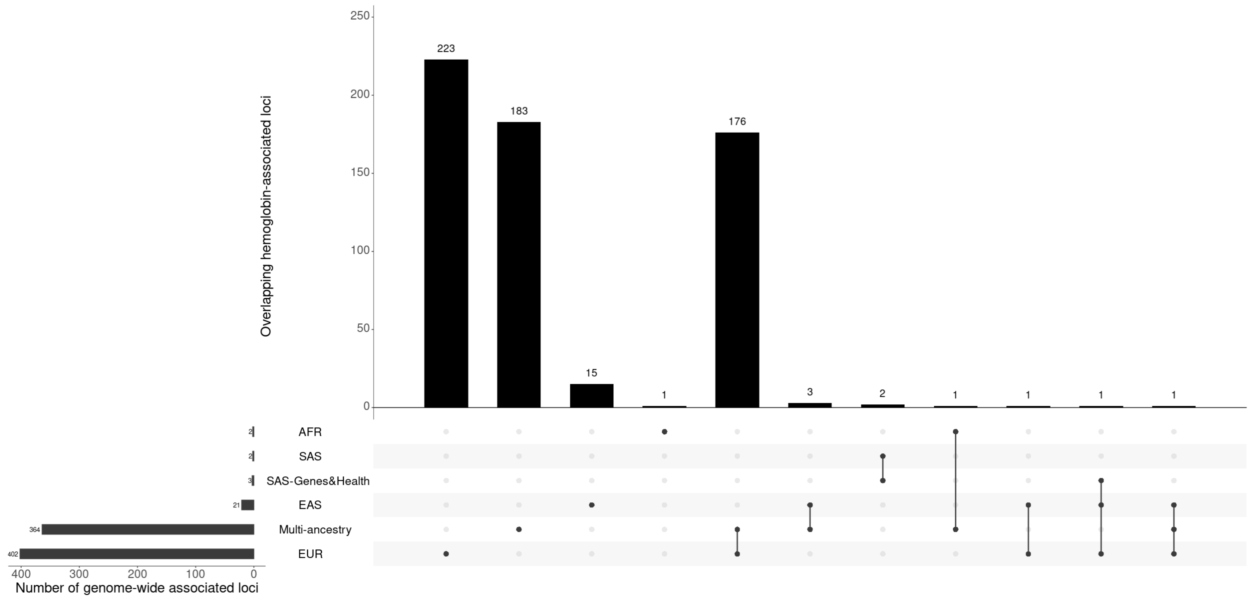

Figure 6.1: Multi-ancestry and ancestry-specific haemoglobin-associated genome-wide significant loci.

EUR, European; EAS, East Asian; AFR, African; SAS, South Asian; SAS-Genes&Health, South Asian from Genes & Health study.

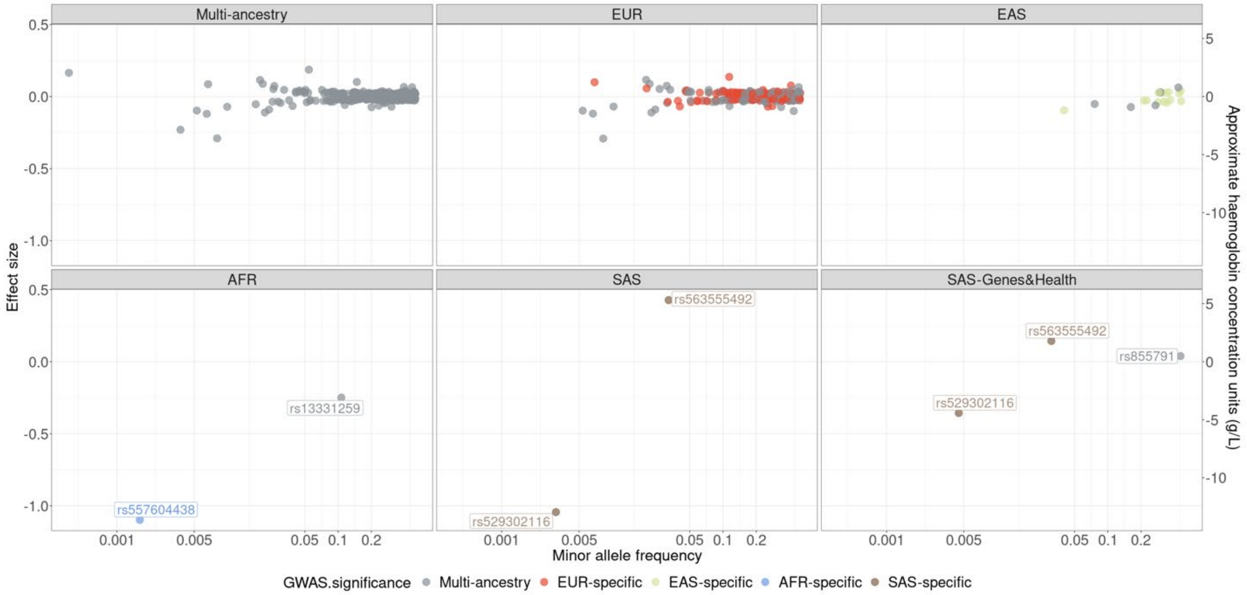

Figure 6.2: Scatter plot of minor allele frequency (MAF) and standardised effect size

We plotted the MAF (x-axis on a log10 scale) and effect size (y-axis) of the minor alleles of the 607 variants (ancestry-specific and cross-ancestry loci) that reached genome-wide significance ($P < 5 \times 10^{-9}$) in the GWAS summary statistics. Coloured by Ancestry-specific or cross-ancestry variants. To convert the standardized effect size to an approximate haemoglobin unit (g/L), we regarded standard deviation per minor allele as 12.41 units (g/L).

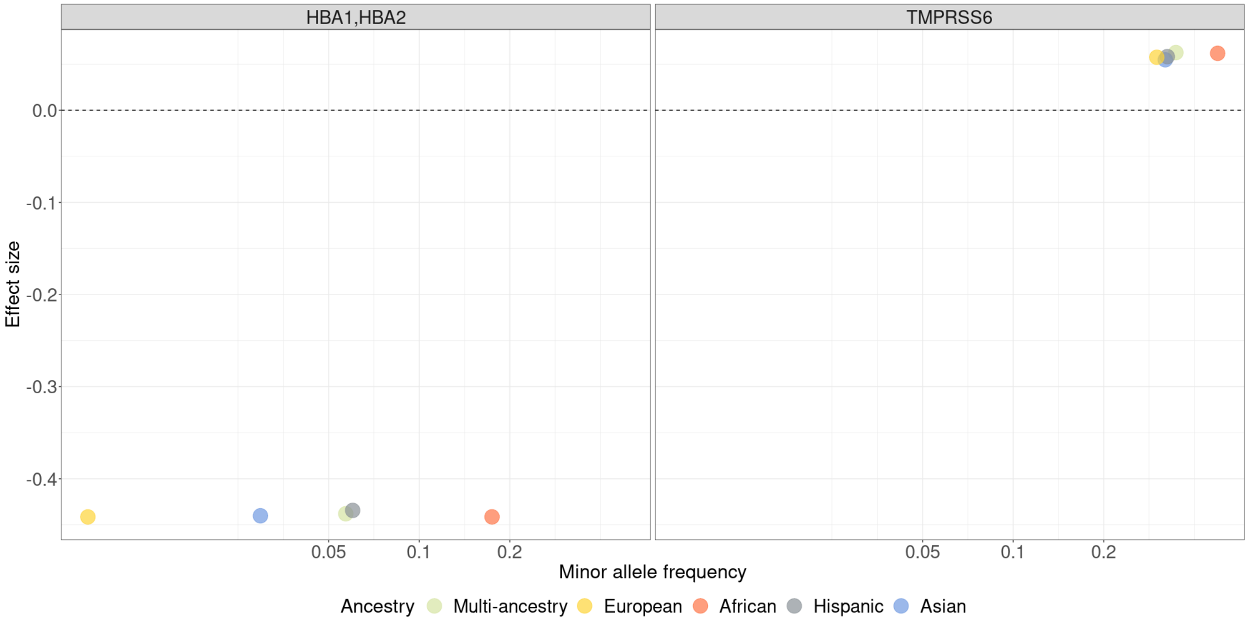

Figure 6.3. Scatter plot of minor allele frequency (MAF) and standardised effect size

We plotted the MAF (x-axis on a log10 scale) and effect size (y-axis) of the minor alleles of the two structural variants that reached genome-wide significance ($P < 5 \times 10^{-8}$) in the GWAS summary statistics. Colored by ancestry.

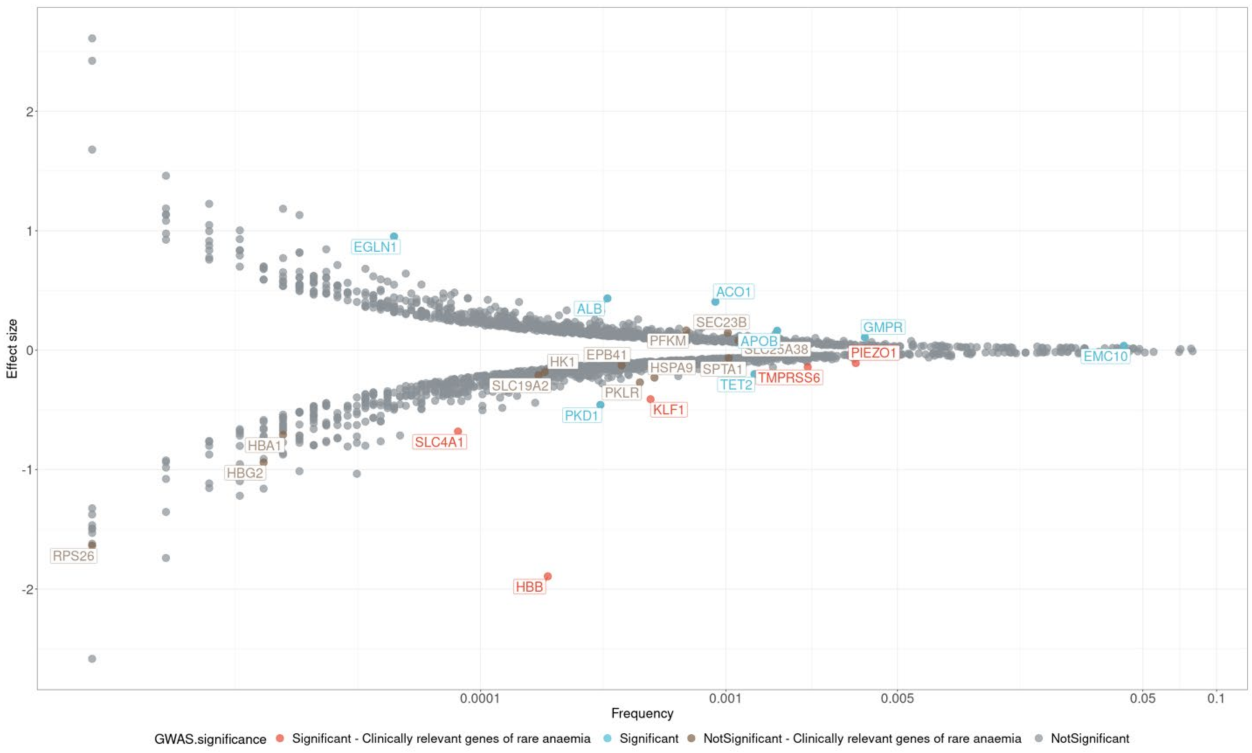

Figure 6.4: Scatter plot of frequency and effect size in the UKBB cohort

Frequency represents the proportion of individuals with a qualifying variant for gene-level results. The effect size is computed from the gene-based collapsing model. Significant associations are those with $P < 2.665 \times 10^{-6}$. The color is represented by whether it is significant and whether it is a clinically relevant cause of rare anaemia from the Genomics England PanelApp 'Green' gene list.

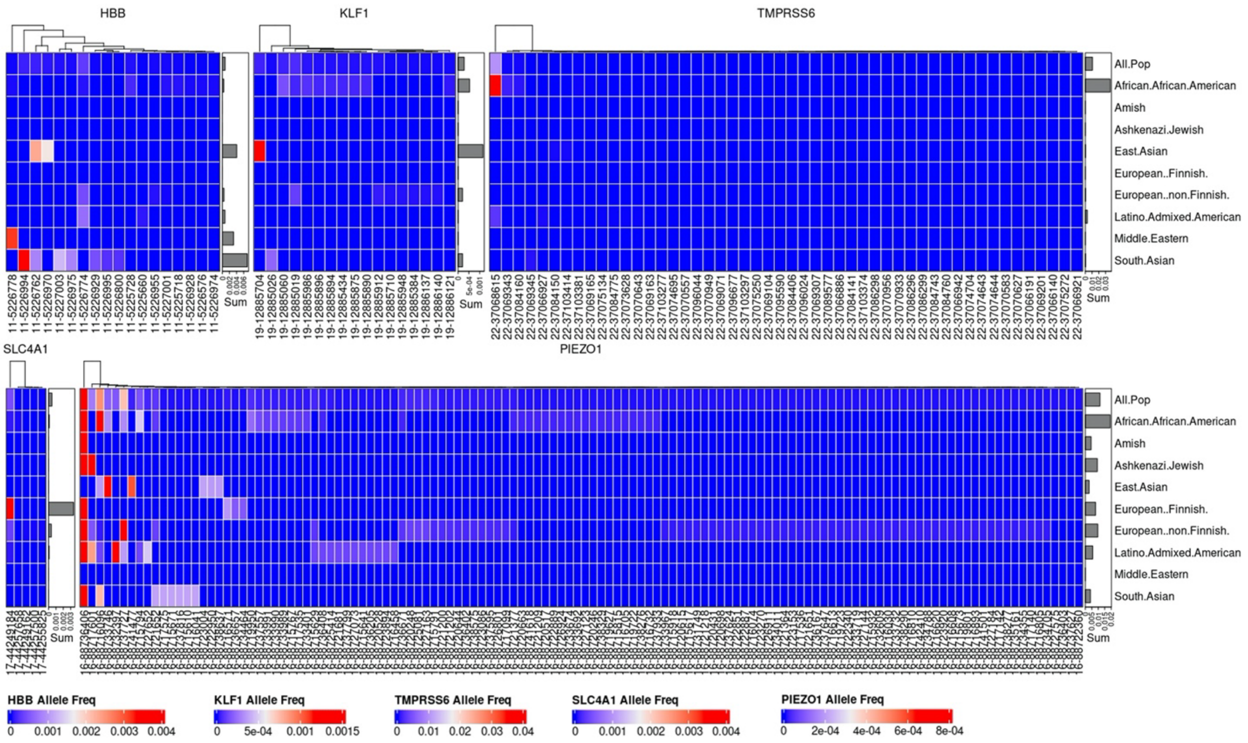

Figure 6.5: Allele frequency plot of predicted loss-of-function (pLOF) variants.

Frequency distribution in different gnomAD populations of five significant genes ($P < 2.665 \times 10^{-6}$) that have been identified as clinically relevant causes of rare anaemia from the Genomics England PanelApp 'Green' gene list. The colour scale for each gene ranges from blue for the lowest allele frequency to red for the highest allele frequency.

### 7. References

1. World Health Organization. WHO guideline on use of ferritin concentrations to assess iron status in individuals and populations. *Geneva: World Health Organization* (2020).
2. Inker, L.A.*, et al.* New Creatinine- and Cystatin C-Based Equations to Estimate GFR without Race. *N Engl J Med* **385**, 1737-1749 (2021).
3. Delgado, C.*, et al.* A Unifying Approach for GFR Estimation: Recommendations of the NKF-ASN Task Force on Reassessing the Inclusion of Race in Diagnosing Kidney Disease. *Am J Kidney Dis* **79**, 268-288.e261 (2022).
4. Institute C and LS. Defining, establishing, and verifying reference intervals in the clinical laboratory; approved guideline. *CLSI document C28–A3* (2008).
5. Hickman, P.E.*, et al.* Choice of Statistical Tools for Outlier Removal Causes Substantial Changes in Analyte Reference Intervals in Healthy Populations. *Clinical Chemistry* **66**, 1558-1561 (2020).
6. Tukey, J.W. Exploratory Data Analysis. *Addison-Wesley.* (1977).
7. Daly, C.H., Higgins, V., Adeli, K., Grey, V.L. & Hamid, J.S. Reference interval estimation: Methodological comparison using extensive simulations and empirical data. *Clin Biochem* **50**, 1145-1158 (2017).
8. Box, G.E.P. & Cox, D.R. An Analysis of Transformations. *Journal of the Royal Statistical Society. Series B (Methodological)* **26**, 211-252 (1964).
9. Canty AJ & AC, D. Resampling-based Variance Estimation for Labour Force Surveys. Journal of the Royal Statistical Society: Series D. *The Statistician* **48**, 379-391 (1999).
10. Woodruff, R.S. Confidence Intervals for Medians and Other Position Measures. *Journal of the American Statistical Association* **47**, 635-646 (1952).
11. Viechtbauer, W. Conducting Meta-Analyses in R with the metafor Package. *Journal of Statistical Software* **36**, 1 - 48 (2010).
12. Hoq, M.*, et al.* Reference Values for 30 Common Biochemistry Analytes Across 5 Different Analyzers in Neonates and Children 30 Days to 18 Years of Age. 1317 (Oxford University Press, 2019).
13. Royston, P. & Wright, E.M. A Method for Estimating Age-Specific Reference Intervals (`Normal Ranges') Based on Fractional Polynomials and Exponential Transformation. *Journal of the Royal Statistical Society. Series A (Statistics in Society)* **161**, 79-101 (1998).
14. Chernick MR. *An introduction to bootstrap methods with applications to R.*, (Wiley, 2011).
15. Davison, A.C. & Hinkley, D.V. *Bootstrap Methods and their Application*, (Cambridge University Press, Cambridge, 1997).
16. StataCorp. Stata Statistical Software: Release 16. (College Station, TX: StataCorp LLC., 2019).
17. R Core Team. R: A language and environment for statistical computing. (R Foundation for Statistical Computing, Vienna, Austria, 2021).
18. StataCorp. Stata Statistical Software: Release 17. (College Station, TX: StataCorp LLC., 2021).

19. Chen, M.-H., *et al.* Trans-ethnic and Ancestry-Specific Blood-Cell Genetics in 746,667 Individuals from 5 Global Populations. *Cell* **182**, 1198-1213.e1114 (2020).
20. Finer, S., *et al.* Cohort Profile: East London Genes & Health (ELGH), a community-based population genomics and health study in British Bangladeshi and British Pakistani people. *International Journal of Epidemiology* **49**, 20-21i (2020).
21. Wheeler, M.M., *et al.* Whole genome sequencing identifies structural variants contributing to hematologic traits in the NHLBI TOPMed program. *Nat Commun* **13**, 7592 (2022).
